# Does the Disease-free Survival Benefit of Adding Belzutifan to Adjuvant Pembrolizumab Persist Beyond the Planned Treatment Period? A Reconstructed Individual Patient Data Analysis of Phase 3 Renal Cell Carcinoma Trials

**DOI:** 10.64898/2026.10.14.26365591

**Authors:** Hao-Han Chang, Chun-Kai Chang, Tsz-Yi Tang, Jhen-Hao Jhan, Shao-Wei Dong, Yung-Shun Juan, Jiun-Hung Geng, Hsien-Che Ou, Hsiang-Ying Lee, Chia-Yang Li, Hsin-Chih Yeh, Mei-Kar Leong, Chia-Hsin Liu, Yung-Chin Lee, Shu-Pin Huang

**Affiliations:** Graduate Institute of Clinical Medicine, College of Medicine, Kaohsiung Medical University, Kaohsiung 80708, Taiwan; Department of Urology, Kaohsiung Medical University Hospital, Kaohsiung 80756, Taiwan; Department of Urology, Kaohsiung Municipal Siaogang Hospital, Kaohsiung 81267, Taiwan; Department of Urology, School of Medicine, College of Medicine, Kaohsiung Medical University, Kaohsiung 80708, Taiwan; Department of Urology, Shuang Ho Hospital, Taipei Medical University, New Taipei City 235, Taiwan; Taipei Medical University (TMU) Research Center of Urology and Kidney, Taipei Medical University, Taipei City 110, Taiwan; Research Center for Environmental Medicine, Kaohsiung Medical University, Kaohsiung 80708, Taiwan; Graduate Institute of Medicine, College of Medicine, Kaohsiung Medical University, Kaohsiung 80708, Taiwan; Center of Applied Genomics, Kaohsiung Medical University, Kaohsiung 80708, Taiwan; Department of Medical Research, Kaohsiung Medical University Hospital, Kaohsiung 80756, Taiwan; Institute of Medical Science and Technology, College of Medicine, National Sun Yat-Sen University, Kaohsiung 804201, Taiwan

**Keywords:** Renal cell carcinoma, Adjuvant therapy, Belzutifan, Pembrolizumab, Disease-free survival, Restricted mean survival time

## Abstract

**Background and Objective:** Adding belzutifan to adjuvant pembrolizumab improved disease-free survival (DFS) in LITESPARK-022, but its durability beyond the 1-year treatment is unknown.

**Methods:** Following a registered plan, we reconstructed individual patient data from published Kaplan-Meier curves of ten phase 3 adjuvant renal cell carcinoma trials. Within each trial, we estimated restricted mean survival time differences (ΔRMST), hazard ratios (HRs) before and after planned treatment ended, and DFS differences.

**Key Findings and Limitations:** In LITESPARK-022 (central review), the 24-month ΔRMST was 1.34 months (95% confidence interval 0.73–1.95), and the belzutifan arm remained ahead after month 12 (ΔRMST increment 0.74 months/yr, 0.32–1.19). The HR was 0.55 (0.43–0.71) before month 12 and 1.16 (0.87–1.54) afterwards (post/pre ratio 2.10); investigator assessment showed similar attenuation. Similar ratios occurred in KEYNOTE-564 at its first interim analysis (2.02), whose benefit persisted, and in SORCE and PROTECT (2.09, 2.59), whose benefit faded. From months 12 to 30 (post hoc), the DFS difference changed by −3.7 percentage points (−8.1 to 0.8) with central review and 0.6 (−4.0 to 5.0) with investigator assessment. Data were reconstructed, and follow-up was short.

**Conclusions and Clinical Implications:** Relative hazard reductions with belzutifan were greater before month 12; durability of the absolute DFS advantage is uncertain and assessment-dependent, so single-HR projections should be avoided.

**Patient Summary:** In a trial after kidney cancer surgery, adding belzutifan to pembrolizumab increased the average time without cancer returning by about 6 weeks over 2 years, but caused more serious side effects. Whether this advantage lasts after treatment ends is not yet clear.

## 1. Introduction

Among the adjuvant trials of vascular endothelial growth factor receptor tyrosine kinase inhibitors (VEGFR-TKIs) after nephrectomy for renal cell carcinoma (RCC), only S-TRAC met its disease-free survival (DFS) endpoint, and none improved overall survival (OS)^1–7^. In KEYNOTE-564, pembrolizumab improved both DFS and OS, and the DFS benefit was maintained at 5 years (hazard ratio [HR] 0.71, 95% confidence interval [CI] 0.59–0.86; 5-year DFS 60.9% vs 52.2%)^8–10^. Other immune checkpoint inhibitor (ICI) regimens did not improve DFS^11–13^.

LITESPARK-022 compared pembrolizumab plus the hypoxia-inducible factor 2α inhibitor belzutifan with pembrolizumab plus placebo, given for up to 1 year^14^. Investigator-assessed DFS improved (HR 0.72, 95% CI 0.59–0.87; 24-month DFS 80.7% vs 73.7%), grade ≥3 adverse events were more frequent (52.1% vs 30.2%), and OS data were immature^14^.

A single summary HR does not show whether an advantage gained during a fixed treatment period is maintained or diminishes after treatment ends. Earlier adjuvant trials show both patterns: in S-TRAC, sunitinib reduced the hazard of recurrence mainly in the first 1.5 years^15^, whereas the DFS benefit of pembrolizumab was sustained through 5 years^10^. Analyses that extended LITESPARK-022 to a comparison with surveillance summarised the effect with a single HR, from Cox models of reconstructed arms pooled across trials^16^ or from an anchored indirect comparison under proportional hazards^17^. The restricted mean survival time (RMST), the average event-free time up to a fixed time point, and time-varying effect measures do not require this assumption^18,19^.

We reconstructed individual patient data from the published Kaplan-Meier curves of phase 3 adjuvant RCC trials and analysed each trial separately. We asked how the DFS benefit of adding belzutifan to pembrolizumab evolves beyond the planned treatment period on relative and absolute scales, and compared this time course with that in other adjuvant trials.

## 2. Patients and methods

### 2.1 Study design and registration

Individual patient data (IPD) were reconstructed from published Kaplan-Meier curves, and every treatment effect was estimated within its own trial; patients or arms from different trials were never combined, and only within-trial estimates were pooled across trials. The statistical analysis plan was registered on the Open Science Framework before the LITESPARK-022 and KEYNOTE-564 curves were digitised (https://osf.io/fk8hz); data access before registration and all deviations are described in Supplementary Table S1. The study used only published aggregate data, and ethics committee approval was not required.

### 2.2 Trial selection

Eligible trials were phase 3 randomised trials of adjuvant or perioperative systemic therapy versus placebo, observation, or placebo added to a common background therapy in resected RCC, including no evidence of disease after metastasectomy, with a published Kaplan-Meier curve of the primary endpoint and numbers at risk. Trials reported only as abstracts, trials of agents no longer in use, and trials of everolimus, which is neither a VEGFR-TKI nor an ICI, were excluded. Trials were identified in PubMed by one reviewer; one trial, ECOG-ACRIN E2810^6^, was added after registration (search history in Supplementary Methods and Supplementary Table S1). Eleven trials were included, ten of which contributed to the main analyses (Table 1). LITESPARK-022 was the trial of primary interest. ICI trials were KEYNOTE-564, IMmotion010, and CheckMate 914 parts A and B (separately randomised cohorts); VEGFR-TKI trials were S-TRAC, ASSURE, PROTECT, ATLAS, SORCE, and E2810. PROSPER (open-label, perioperative, observation-controlled)^20^ was used only in sensitivity analyses.

**Table 1.**
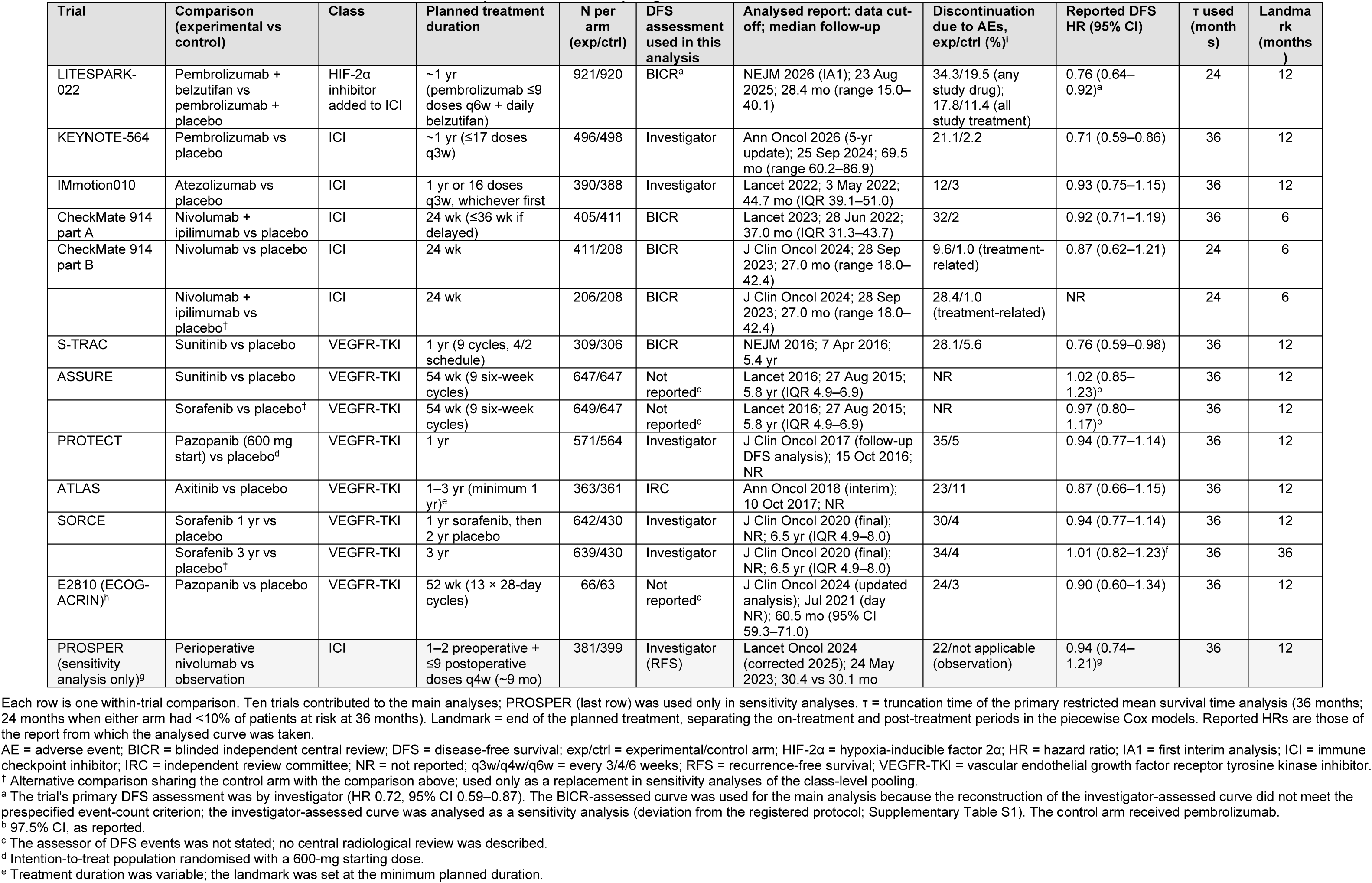
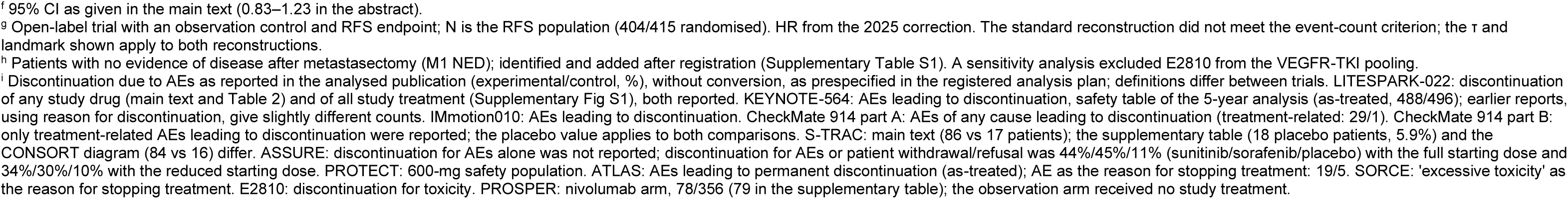
Characteristics of the included trials, the curves analysed, and the analysis parameters.

| Trial | Comparison (experimental vs control) | Class | Planned treatment duration | N per arm (exp/ctrl) | DFS assessment used in this analysis | Analysed report: data cut-off; median follow-up | Discontinuation due to AEs, exp/ctrl (%)<sup>i</sup> | Reported DFS HR (95% CI) | τ used (months) | Landmark (months) |
| --- | --- | --- | --- | --- | --- | --- | --- | --- | --- | --- |
| LITESPARK-022 | Pembrolizumab + belzutifan vs pembrolizumab + placebo | HIF-2α inhibitor added to ICI | ~1 yr (pembrolizumab ≤9 doses q6w + daily belzutifan) | 921/920 | BICR<sup>a</sup> | NEJM 2026 (IA1); 23 Aug 2025; 28.4 mo (range 15.0–40.1) | 34.3/19.5 (any study drug); 17.8/11.4 (all study treatment) | 0.76 (0.64–0.92)<sup>a</sup> | 24 | 12 |
| KEYNOTE-564 | Pembrolizumab vs placebo | ICI | ~1 yr (≤17 doses q3w) | 496/498 | Investigator | Ann Oncol 2026 (5-yr update); 25 Sep 2024; 69.5 mo (range 60.2–86.9) | 21.1/2.2 | 0.71 (0.59–0.86) | 36 | 12 |
| IMmotion010 | Atezolizumab vs placebo | ICI | 1 yr or 16 doses q3w, whichever first | 390/388 | Investigator | Lancet 2022; 3 May 2022; 44.7 mo (IQR 39.1–51.0) | 12/3 | 0.93 (0.75–1.15) | 36 | 12 |
| CheckMate 914 part A | Nivolumab + ipilimumab vs placebo | ICI | 24 wk (≤36 wk if delayed) | 405/411 | BICR | Lancet 2023; 28 Jun 2022; 37.0 mo (IQR 31.3–43.7) | 32/2 | 0.92 (0.71–1.19) | 36 | 6 |
| CheckMate 914 part B | Nivolumab vs placebo | ICI | 24 wk | 411/208 | BICR | J Clin Oncol 2024; 28 Sep 2023; 27.0 mo (range 18.0–42.4) | 9.6/1.0 (treatment-related) | 0.87 (0.62–1.21) | 24 | 6 |
|  | Nivolumab + ipilimumab vs placebo<sup>†</sup> | ICI | 24 wk | 206/208 | BICR | J Clin Oncol 2024; 28 Sep 2023; 27.0 mo (range 18.0–42.4) | 28.4/1.0 (treatment-related) | NR | 24 | 6 |
| S-TRAC | Sunitinib vs placebo | VEGFR-TKI | 1 yr (9 cycles, 4/2 schedule) | 309/306 | BICR | NEJM 2016; 7 Apr 2016; 5.4 yr | 28.1/5.6 | 0.76 (0.59–0.98) | 36 | 12 |
| ASSURE | Sunitinib vs placebo | VEGFR-TKI | 54 wk (9 six-week cycles) | 647/647 | Not reported<sup>c</sup> | Lancet 2016; 27 Aug 2015; 5.8 yr (IQR 4.9–6.9) | NR | 1.02 (0.85–1.23)<sup>b</sup> | 36 | 12 |
|  | Sorafenib vs placebo<sup>†</sup> | VEGFR-TKI | 54 wk (9 six-week cycles) | 649/647 | Not reported<sup>c</sup> | Lancet 2016; 27 Aug 2015; 5.8 yr (IQR 4.9–6.9) | NR | 0.97 (0.80–1.17)<sup>b</sup> | 36 | 12 |
| PROTECT | Pazopanib (600 mg start) vs placebo<sup>d</sup> | VEGFR-TKI | 1 yr | 571/564 | Investigator | J Clin Oncol 2017 (follow-up DFS analysis); 15 Oct 2016; NR | 35/5 | 0.94 (0.77–1.14) | 36 | 12 |
| ATLAS | Axitinib vs placebo | VEGFR-TKI | 1–3 yr (minimum 1 yr)<sup>e</sup> | 363/361 | IRC | Ann Oncol 2018 (interim); 10 Oct 2017; NR | 23/11 | 0.87 (0.66–1.15) | 36 | 12 |
| SORCE | Sorafenib 1 yr vs placebo | VEGFR-TKI | 1 yr sorafenib, then 2 yr placebo | 642/430 | Investigator | J Clin Oncol 2020 (final); NR; 6.5 yr (IQR 4.9–8.0) | 30/4 | 0.94 (0.77–1.14) | 36 | 12 |
|  | Sorafenib 3 yr vs placebo<sup>†</sup> | VEGFR-TKI | 3 yr | 639/430 | Investigator | J Clin Oncol 2020 (final); NR; 6.5 yr (IQR 4.9–8.0) | 34/4 | 1.01 (0.82–1.23)<sup>f</sup> | 36 | 36 |
| E2810 (ECOG-ACRIN)<sup>h</sup> | Pazopanib vs placebo | VEGFR-TKI | 52 wk (13 × 28-day cycles) | 66/63 | Not reported<sup>c</sup> | J Clin Oncol 2024 (updated analysis); Jul 2021 (day NR); 60.5 mo (95% CI 59.3–71.0) | 24/3 | 0.90 (0.60–1.34) | 36 | 12 |
| PROSPER (sensitivity analysis only)<sup>g</sup> | Perioperative nivolumab vs observation | ICI | 1–2 preoperative + ≤9 postoperative doses q4w (~9 mo) | 381/399 | Investigator (RFS) | Lancet Oncol 2024 (corrected 2025); 24 May 2023; 30.4 vs 30.1 mo | 22/not applicable (observation) | 0.94 (0.74–1.21)<sup>g</sup> | 36 | 12 |

Each row is one within-trial comparison. Ten trials contributed to the main analyses; PROSPER (last row) was used only in sensitivity analyses. τ = truncation time of the primary restricted mean survival time analysis (36 months; 24 months when either arm had <10% of patients at risk at 36 months). Landmark = end of the planned treatment, separating the on-treatment and post-treatment periods in the piecewise Cox models. Reported HRs are those of the report from which the analysed curve was taken.
AE = adverse event; BICR = blinded independent central review; DFS = disease-free survival; exp/ctrl = experimental/control arm; HIF-2α = hypoxia-inducible factor 2α; HR = hazard ratio; IA1 = first interim analysis; ICI = immune checkpoint inhibitor; IRC = independent review committee; NR = not reported; q3w/q4w/q6w = every 3/4/6 weeks; RFS = recurrence-free survival; VEGFR-TKI = vascular endothelial growth factor receptor tyrosine kinase inhibitor.
<sup>†</sup> Alternative comparison sharing the control arm with the comparison above; used only as a replacement in sensitivity analyses of the class-level pooling.
<sup>a</sup> The trial's primary DFS assessment was by investigator (HR 0.72, 95% CI 0.59–0.87). The BICR-assessed curve was used for the main analysis because the reconstruction of the investigator-assessed curve did not meet the prespecified event-count criterion; the investigator-assessed curve was analysed as a sensitivity analysis (deviation from the registered protocol; Supplementary Table S1). The control arm received pembrolizumab.
<sup>b</sup> 97.5% CI, as reported.
<sup>c</sup> The assessor of DFS events was not stated; no central radiological review was described.
<sup>d</sup> Intention-to-treat population randomised with a 600-mg starting dose.
<sup>e</sup> Treatment duration was variable; the landmark was set at the minimum planned duration.
<sup>f</sup> 95% CI as given in the main text (0.83–1.23 in the abstract).

### 2.3 Curve selection and reconstruction

For each comparison, one curve was selected by prespecified rules: the intention-to-treat population, the trial’s primary assessment method, and the latest analysis with full text, preferring the publisher’s PDF (Supplementary Table S2). Vector curves were extracted from the PDF drawing paths with PyMuPDF 1.28.2; raster curves, used only in sensitivity or secondary analyses, were digitised by colour tracing (Supplementary Methods). IPD were reconstructed with the Guyot algorithm in the IPDfromKM package^21,22^. Prespecified accuracy criteria were a reconstructed HR within 0.05 of, and inside the CI of, the reported value; event counts within 5% in each arm; survival at reported time points within 2 percentage points; and close visual overlay. When a publication’s figure and text disagreed, the figure and its numbers at risk were taken as the reference, as prespecified, and each such adjudication was documented (Supplementary Table S1). A second investigator checked every digitisation and reconstruction overlay, and curves that failed were moved to sensitivity analyses. For LITESPARK-022, the standard reconstruction of the investigator-assessed DFS curve (the trial’s primary endpoint) undercounted events by 7.0% in the belzutifan arm, so the main analysis used the blinded independent central review (BICR)-assessed curve from the same report. This decision was made before any time-course analysis. In a post hoc analysis, curves that failed because of non-uniform censoring were reconstructed using the censoring marks or counts printed on the figure (censoring-informed reconstruction); the investigator-assessed LITESPARK-022 curve met all criteria with this method and is reported alongside the BICR results.

### 2.4 Endpoints and time parameters

The primary endpoint was DFS as defined in each trial; OS was secondary. The RMST truncation time (τ) was 36 months, reduced to 24 months if any arm had fewer than 10% of patients at risk at 36 months; this set τ at 24 months for LITESPARK-022 and CheckMate 914 part B. Follow-up was divided at a landmark set at the end of planned treatment: 12 months for most trials, including ATLAS, whose planned treatment lasted 1–3 years; 6 months for CheckMate 914; and 36 months for the 3-year sorafenib arm of SORCE. These periods were defined by calendar time, not by individual treatment discontinuation.

### 2.5 Statistical analysis

The primary analysis was the within-trial difference in RMST (ΔRMST) at τ^18^. To describe how the benefit evolved, ΔRMST was also computed at truncation times from 6 months in 3-month steps up to the longest follow-up at which each arm retained at least 10% of patients at risk. The post-landmark increment, ΔRMST at this longest truncation time minus ΔRMST at the landmark, was expressed per year. Secondary analyses were piecewise Cox models with a treatment-by-period interaction, fitted to all follow-up, giving HRs before and after the landmark and their ratio (post-landmark divided by pre-landmark HR; values above 1 indicate a smaller relative reduction after the landmark). We also estimated Kaplan-Meier DFS differences at 12–60 months and tested proportional hazards with Schoenfeld residuals; time-varying HRs from flexible parametric models^23^ and smoothed hazards are reported in Supplementary Table S12 and the Supplementary Figures. The log ratio of post-landmark to pre-landmark HRs was pooled separately for VEGFR-TKI and ICI trials by random-effects meta-analysis (restricted maximum likelihood with Hartung-Knapp adjustment), with one comparison per trial or independently randomised cohort (CheckMate 914 parts A and B entered separately). LITESPARK-022, whose control arm received pembrolizumab, was not pooled, and no formal cross-trial test was made. The registered objective of describing which class LITESPARK-022 more closely resembled was addressed descriptively only. The registered statement that a fading benefit yields an increment near zero was corrected, because the increment equals the average DFS difference after the landmark (Supplementary Table S1). Registered sensitivity analyses used alternative landmarks, earlier KEYNOTE-564 data cut-offs, alternative populations and assessment methods, and PROSPER (Supplementary Tables S3, S6, and S16–S18). Post hoc analyses were bootstrap CIs (2000 resamples within arms) for the increment; a window-matched analysis estimating the change in DFS difference and the increment over the 18 months after the landmark, the post-landmark follow-up available in LITESPARK-022; a modified Hartung-Knapp adjustment, which does not allow the adjusted standard error to fall below the unadjusted one; and pooling without E2810. Secondary and post hoc results are descriptive, with nominal CIs. Analyses used R 4.6.0 (details in Supplementary Methods).

## 3. Results

### 3.1 Reconstruction

Fourteen within-trial DFS comparisons from ten trials were analysed (Table 1). All curves used in the main analyses met the accuracy criteria, after a documented adjudication for PROTECT, whose tabulated 12-month DFS corresponded to the curve after a drop at the 12-month imaging visit (Supplementary Table S1). Reconstructed HRs were within 0.04 of the reported values, and event counts were within 3.1% in every arm; the second investigator confirmed every overlay (Supplementary Table S5). For LITESPARK-022, the BICR-assessed curve reproduced the reported HR (0.756 vs 0.76); the investigator-assessed curve met the criteria with the censoring-informed reconstruction (0.719 vs 0.72).

### 3.2 Restricted mean DFS time

In LITESPARK-022, adding belzutifan increased restricted mean DFS time over 24 months by 1.34 months (BICR; 95% CI 0.73–1.95), about 6 weeks of additional disease-free time on average, and by 1.29 months (0.75–1.83) with investigator assessment. Over 24 months, pembrolizumab in KEYNOTE-564 increased restricted mean DFS time by 1.98 months (1.08–2.88), an effect against placebo rather than on top of pembrolizumab. Results for the other comparisons are shown in Figs. 1 and 2 and Supplementary Table S8.

**Fig. 1.**
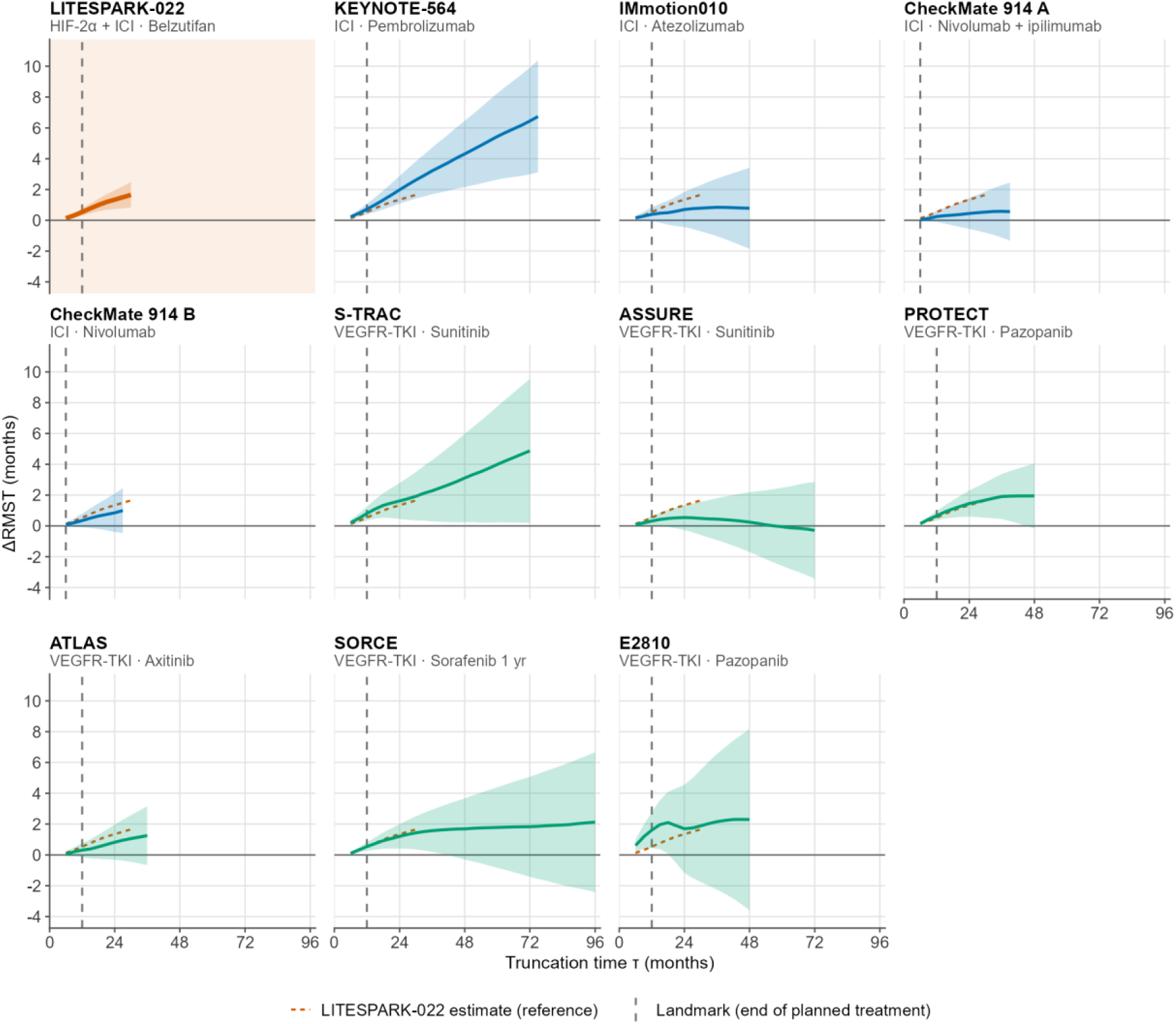
Difference in restricted mean disease-free survival time (ΔRMST) as a function of the truncation time τ for the 11 main-analysis comparisons. Each panel shows one within-trial comparison (experimental minus control arm): the solid line is the ΔRMST estimate and the shaded band its pointwise 95% confidence interval, computed from 6 months in 3-month steps up to the longest follow-up at which each arm retained ≥10% of patients at risk. The vertical dashed line marks the landmark at the end of planned treatment (12 months; 6 months for CheckMate 914). The orange dashed line repeats the LITESPARK-022 estimate in every panel for reference. Colours denote the drug class: orange, HIF-2α inhibitor added to an ICI; blue, ICI; green, VEGFR-TKI. LITESPARK-022 is shown with BICR-assessed DFS and a pembrolizumab-plus-placebo control arm; KEYNOTE-564 is shown with the 5-year analysis, and SORCE with 1 year of sorafenib. Alternative comparisons sharing a control arm and the sensitivity curves are shown in Supplementary Figure S1. BICR = blinded independent central review; DFS = disease-free survival; HIF-2α = hypoxia-inducible factor 2α; ICI = immune checkpoint inhibitor; RMST = restricted mean survival time; VEGFR-TKI = vascular endothelial growth factor receptor tyrosine kinase inhibitor.

**Fig. 2.**
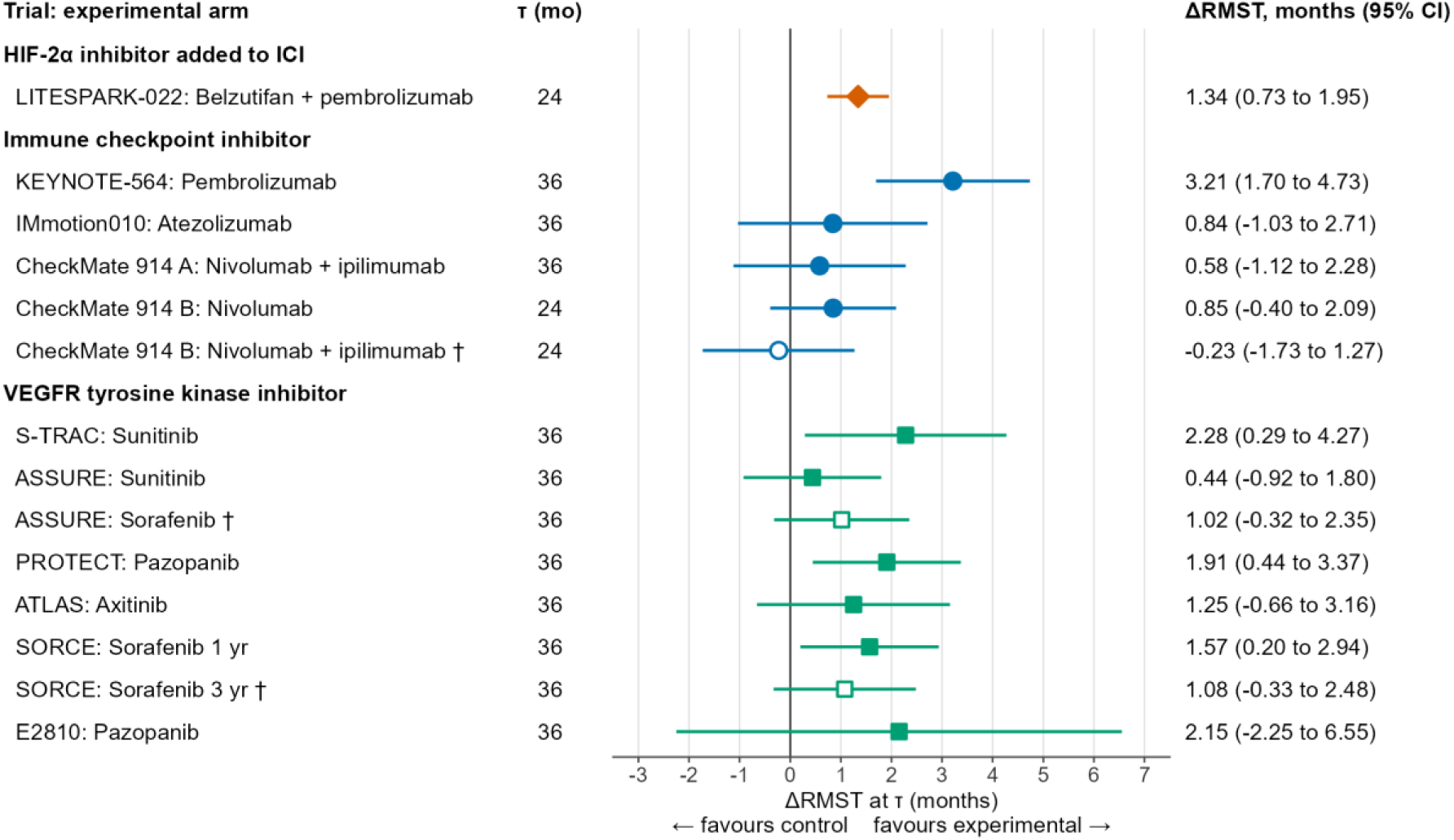
Difference in restricted mean disease-free survival time (ΔRMST) at the primary truncation time τ, by drug class. Symbols show the within-trial estimate (experimental minus control arm) and horizontal lines the 95% confidence interval; values above zero favour the experimental arm. τ was 36 months, or 24 months when either arm had <10% of patients at risk at 36 months (LITESPARK-022 and CheckMate 914 part B). Open symbols marked † are alternative comparisons sharing the control arm with the comparison above, used only in sensitivity analyses. LITESPARK-022 is shown with BICR-assessed DFS; its control arm received pembrolizumab, so its ΔRMST is incremental to pembrolizumab. Diamond, HIF-2α inhibitor added to an ICI; circles, ICI; squares, VEGFR-TKI. BICR = blinded independent central review; DFS = disease-free survival; HIF-2α = hypoxia-inducible factor 2α; ICI = immune checkpoint inhibitor; RMST = restricted mean survival time; VEGFR = vascular endothelial growth factor receptor; VEGFR-TKI = VEGFR tyrosine kinase inhibitor.

### 3.3 Time course of the DFS benefit

In LITESPARK-022 (BICR), the HR was 0.55 (95% CI 0.43–0.71) before the 12-month landmark, a 45% lower hazard of recurrence or death, and 1.16 (0.87–1.54) afterwards; the post-to-pre-landmark HR ratio was 2.10 (1.44–3.08) (Fig. 3). With investigator assessment, the HRs were 0.52 (0.39–0.69) and 0.95 (0.73–1.24), a ratio of 1.83 (1.24–2.70). Thus, the relative hazard reduction with belzutifan was greater before month 12 under both assessments. On the absolute scale, the BICR-assessed DFS difference was 7.7 percentage points (4.5–11.0) at 12 months and 4.9 points (0.7–9.1) at 24 months. The registered post-landmark increment was positive (0.74 months per year, bootstrap 95% CI 0.32–1.19), corresponding to an average DFS difference of about 6 points over months 12–30 (Fig. 1). This shows that the belzutifan arm remained ahead, but not whether the difference was stable.

**Fig. 3.**
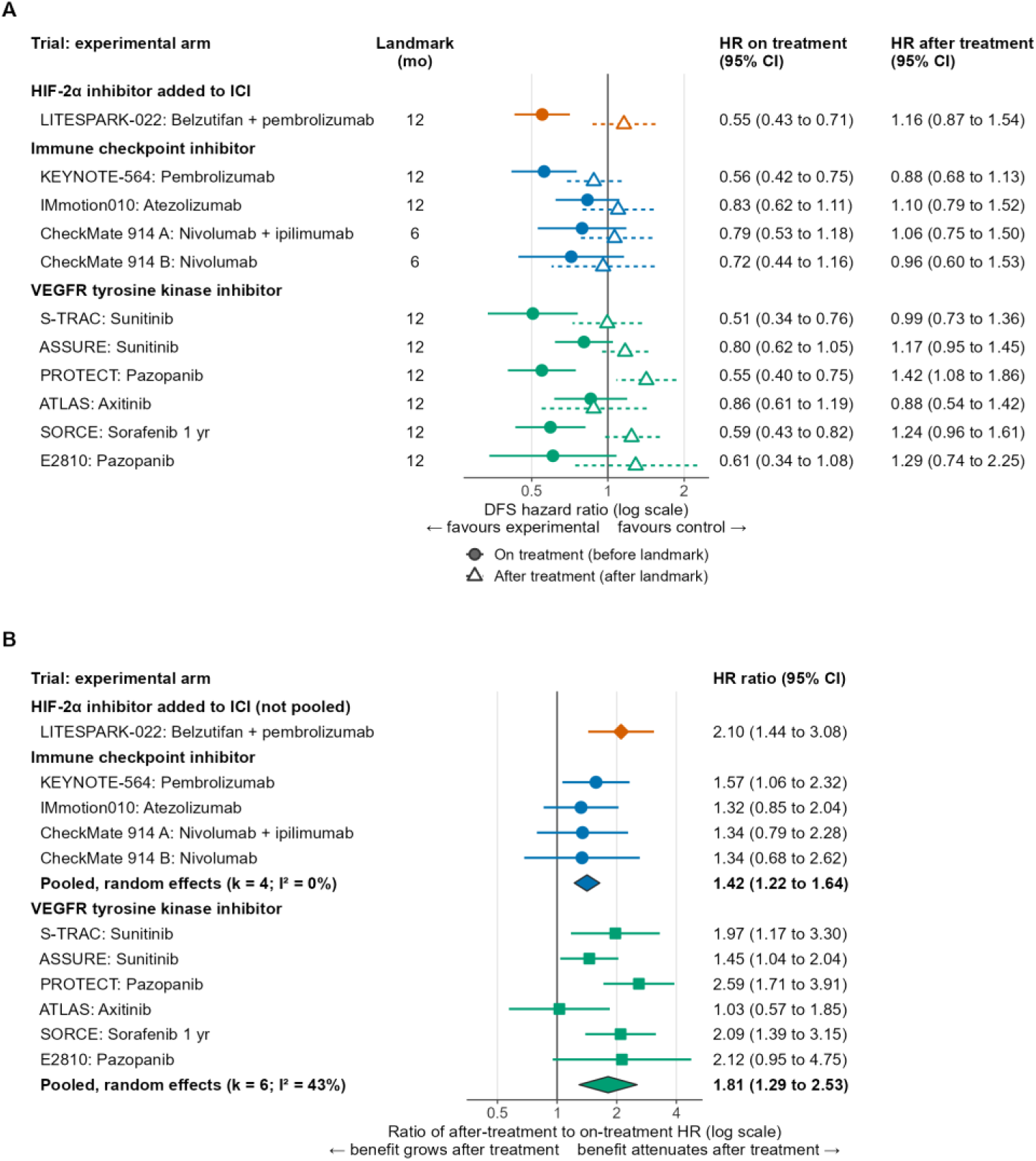
Disease-free survival hazard ratios before and after the landmark (A) and their ratio (B). (A) HRs (log scale) from piecewise Cox models with a treatment-by-period interaction, fitted to all follow-up: filled circles with solid lines, on treatment (before the landmark); open triangles with dashed lines, after treatment (after the landmark). The landmark (end of planned treatment) is listed for each comparison; HRs <1 favour the experimental arm. (B) Ratio of the after-treatment to the on-treatment HR (log scale); ratios >1 indicate a smaller relative hazard reduction after the landmark. Diamonds at the bottom of each class show the random-effects pooled ratio for ICI and VEGFR-TKI trials (restricted maximum likelihood with Hartung-Knapp adjustment; k = number of comparisons). LITESPARK-022 (BICR-assessed DFS; orange diamond) is shown alongside but was not pooled, and no formal cross-trial test was made. Horizontal lines are 95% confidence intervals; these secondary analyses are descriptive, with nominal confidence intervals. BICR = blinded independent central review; DFS = disease-free survival; HIF-2α = hypoxia-inducible factor 2α; HR = hazard ratio; ICI = immune checkpoint inhibitor; VEGFR = vascular endothelial growth factor receptor; VEGFR-TKI = VEGFR tyrosine kinase inhibitor.

In KEYNOTE-564, the ratio at the 5-year analysis was 1.57 (1.06–2.32), while the DFS difference stayed between 8.6 and 11.1 points from 12 to 60 months. At its first interim analysis (median follow-up 24.1 months; investigator assessment; 73 events after month 12), the ratio was 2.02 (1.16–3.49), and the DFS difference narrowed from 9.6 to 6.0 points between 12 and 30 months (change −3.6 points, 95% CI −9.4 to 2.1; post hoc). At the 30-month update, closer in follow-up to LITESPARK-022 (28.4 months), the ratio was 1.35 (0.82–2.21), and the difference did not narrow (9.3 to 9.9 points) (Supplementary Table S16). Among VEGFR-TKI trials, the ratio ranged from 1.03 for ATLAS to 2.59 for PROTECT and was 2.09 for SORCE (1 year) (Fig. 3). In PROTECT and SORCE, the DFS difference fell from 8.7 and 6.8 points at 12 months to 0.1 and 0.7 points at 48 months. Thus, ratios near 2 occurred both in KEYNOTE-564 at its first interim analysis, whose absolute benefit persisted, and in PROTECT and SORCE, whose benefit faded.

Proportional hazards were rejected for LITESPARK-022 (P < 0.001) and four other trials (Supplementary Table S12).

### 3.4 Class patterns

The pooled ratio of post-landmark to pre-landmark HRs was 1.81 (95% CI 1.29–2.53; I² 43%) for VEGFR-TKI trials and 1.42 (1.22–1.64; I² 0%) for ICI trials (Fig. 3B). Without E2810 (metastasectomy only), the VEGFR-TKI ratio was 1.78 (1.17–2.72). Both pooled ratios exceeded 1. With the modified Hartung-Knapp adjustment, the ICI interval widened to 0.96–2.09 and included 1, as did six of eight ICI sensitivity analyses. VEGFR-TKI intervals excluded 1 with either adjustment (Supplementary Table S6).

### 3.5 Window-matched analysis (post hoc)

We next compared the change in the absolute DFS difference over the same 18 months after the landmark in each main comparison (Fig. 4). In LITESPARK-022, the BICR-assessed difference changed from 7.7 to 4.0 points (−3.7, 95% CI −8.1 to 0.8), and the investigator-assessed difference from 6.7 to 7.2 points (0.6, −4.0 to 5.0). Among the other ten main comparisons, the difference narrowed in eight, by 0.2 to 7.9 points, including S-TRAC, PROTECT, SORCE, and E2810, and did not narrow in KEYNOTE-564 (5-year analysis) or ATLAS; the CI excluded zero only for PROTECT (Supplementary Table S13). Thus, whether the absolute DFS advantage of belzutifan narrowed after the landmark depended on the assessment method.

**Fig. 4.**
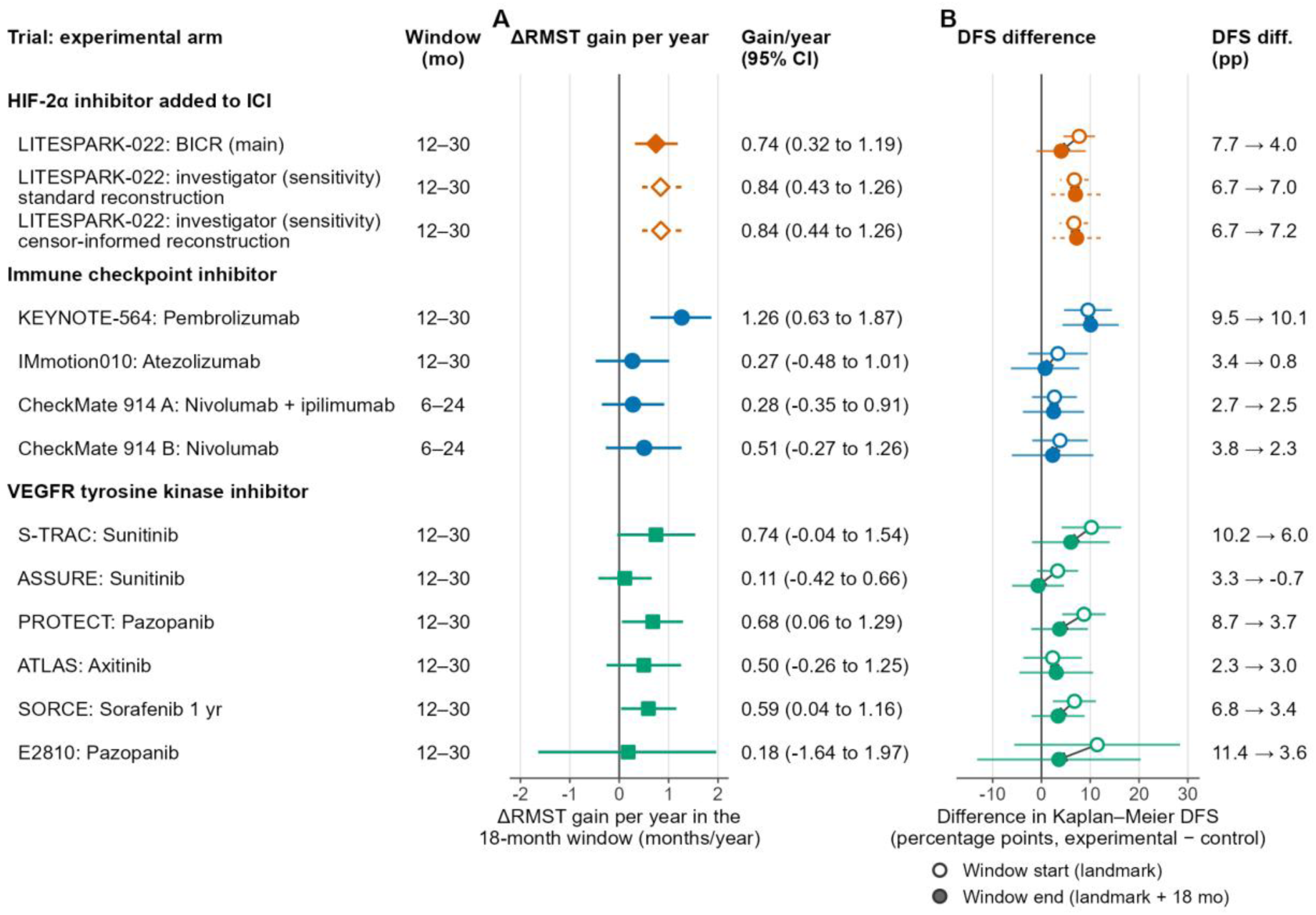
Window-matched analysis over the 18 months after the landmark (post hoc). The window was months 12–30 (months 6–24 for CheckMate 914). (A) Gain in ΔRMST per year within the window (months per year), with bootstrap 95% confidence intervals (2000 resamples within arms). (B) Kaplan-Meier DFS difference (experimental minus control arm, percentage points) at the start (open circles) and end (filled circles) of the window, with Greenwood 95% confidence intervals; arrows show the direction of change, and the start and end values are listed on the right. LITESPARK-022 is shown with BICR-assessed DFS (main analysis) and with investigator-assessed DFS (sensitivity analyses; standard and censoring-informed reconstructions; open diamonds in A and dashed interval lines). KEYNOTE-564 is shown with the 5-year analysis. Colours denote the drug class: orange, HIF-2α inhibitor added to an ICI; blue, ICI; green, VEGFR-TKI. This analysis was not prespecified; alternative comparisons and PROSPER are shown in Supplementary Figure S9. BICR = blinded independent central review; DFS = disease-free survival; HIF-2α = hypoxia-inducible factor 2α; ICI = immune checkpoint inhibitor; RMST = restricted mean survival time; VEGFR-TKI = vascular endothelial growth factor receptor tyrosine kinase inhibitor. Changes were computed before rounding.

### 3.6 Overall survival

The OS ΔRMST was 0.04 months (95% CI −0.18 to 0.25) at 24 months in LITESPARK-022 (descriptive) and 0.60 months (−0.01 to 1.20) at 36 months in KEYNOTE-564. OS results for the other trials, including E2810, are given in Supplementary Table S7.

## 4. Discussion

Adding belzutifan to pembrolizumab increased restricted mean DFS time, and its relative hazard reduction was greater before month 12. Whether the absolute advantage narrowed afterwards depended on the assessment method.

Attenuation of the hazard reduction after month 12 did not by itself predict loss of benefit. KEYNOTE-564 showed a similar attenuation, and at its first interim analysis a similar narrowing of the DFS difference, yet its benefit persisted with longer follow-up. VEGFR-TKI trials with a similar attenuation included some whose benefit faded. Period-specific HRs after month 12 compare only patients still disease-free, who may differ between arms, and cannot isolate the effect of stopping treatment^24^, so we placed more weight on absolute DFS differences and ΔRMST, which compare the arms as randomised.

The VEGFR-TKI trials did not share one pattern, and E2810, which enrolled only patients after metastasectomy, changed the pooled ratio little.

The disagreement between assessment methods is the main source of uncertainty. It is not an artefact of reconstruction, because the published 12- and 30-month DFS estimates show the same divergence (BICR: 7.9 points at 12 months and 3.8 at 30 months; investigator: 6.7 and 7.2 points)^14^. As both assessments come from the same patients, the disagreement may reflect the small number of events after month 12 and differences in which events, and which dates, each assessment recorded.

Earlier analyses of adjuvant RCC trials^25,26^ and of the LITESPARK-022 combination^16,17^ summarised effects with a single HR. Because hazards were not proportional in LITESPARK-022, such estimates should not be read as a constant effect or extrapolated beyond the observed follow-up.

For patients, adding belzutifan meant about 6 weeks of additional disease-free time over 2 years, against an increase in grade ≥3 adverse events from 30.2% to 52.1%^14^. Our data cannot show whether recurrences were prevented or delayed, and counselling should convey this uncertainty. If the benefit is durable, later LITESPARK-022 analyses should show a stable DFS difference beyond month 30 and an undiminished post-landmark increment; a narrowing difference with an increment approaching zero would indicate fading benefit.

Our study has limitations. Reconstructed data approximate the original data and cannot recover covariates or exact event times, and bootstrap intervals did not include uncertainty from digitisation. Follow-up after month 12 in LITESPARK-022 was short, OS data were immature, and periods were defined by planned treatment duration, although about 30% of patients in each arm stopped trial treatment early^14^. The trials differed in population, era, imaging schedule, and DFS definition, and the LITESPARK-022 effect is incremental to pembrolizumab. Finally, several analyses were post hoc, trials were screened by one reviewer, and one trial was added after registration.

## 5. Conclusions

In LITESPARK-022, adding belzutifan to pembrolizumab improved restricted mean DFS over 24 months, and the relative reduction in hazard was greater before month 12, a pattern seen with both durable and non-durable adjuvant therapies. Whether the absolute DFS advantage persists is uncertain and depends on the assessment method; longer follow-up, analysed with time-varying and absolute measures, is needed.

## Supporting information

Registered Statistical Analysis Plan (OSF, 12 October 2026)

## Ethics statement

This study used only published aggregate data and did not require ethics committee approval.

## Data availability

The statistical analysis plan was registered on the Open Science Framework on 12 October 2026 (https://osf.io/fk8hz; under embargo until publication) and is provided as a supplementary file. The reconstructed individual patient data and analysis code are available from the corresponding author on request and will be deposited on the Open Science Framework at publication.

## Declaration of generative AI and AI-assisted technologies in the writing process

During the preparation of this work, the authors used Claude (Anthropic), an AI assistant, to assist with literature searching, data extraction, writing of the analysis code, and drafting and editing of the manuscript. An additional AI model, Codex (OpenAI), was used for an internal pre-submission review. All outputs were checked by the authors, who reviewed and edited the content as needed and take full responsibility for the content of the publication.

## Author contributions

Conceptualization: Hao-Han Chang, Shu-Pin Huang.

Methodology: Hao-Han Chang.

Software: Hao-Han Chang.

Formal analysis: Hao-Han Chang.

Investigation: Hao-Han Chang.

Data curation: Hao-Han Chang.

Validation: Chun-Kai Chang.

Visualization: Hao-Han Chang.

Supervision: Chia-Yang Li, Shu-Pin Huang.

Project administration: Hao-Han Chang.

Resources: Shu-Pin Huang.

Funding acquisition: Shu-Pin Huang.

Writing – original draft: Hao-Han Chang.

Writing – review & editing: all authors.

Shu-Pin Huang had full access to all the data in the study and takes responsibility for the integrity of the data and the accuracy of the data analysis.

## References

1. Ravaud A, Motzer RJ, Pandha HS, George DJ, Pantuck AJ, Patel A, et al. Adjuvant sunitinib in high-risk renal-cell carcinoma after nephrectomy. N Engl J Med. 2016;375(23):2246–2254. doi:10.1056/NEJMoa1611406

2. Haas NB, Manola J, Uzzo RG, Flaherty KT, Wood CG, Kane C, et al. Adjuvant sunitinib or sorafenib for high-risk, non-metastatic renal-cell carcinoma (ECOG-ACRIN E2805): a double-blind, placebo-controlled, randomised, phase 3 trial. Lancet. 2016;387(10032):2008–2016. doi:10.1016/S0140-6736(16)00559-6

3. Motzer RJ, Haas NB, Donskov F, Gross-Goupil M, Varlamov S, Kopyltsov E, et al. Randomized phase III trial of adjuvant pazopanib versus placebo after nephrectomy in patients with localized or locally advanced renal cell carcinoma. J Clin Oncol. 2017;35(35):3916–3923. doi:10.1200/JCO.2017.73.5324

4. Gross-Goupil M, Kwon TG, Eto M, Ye D, Miyake H, Seo SI, et al. Axitinib versus placebo as an adjuvant treatment of renal cell carcinoma: results from the phase III, randomized ATLAS trial. Ann Oncol. 2018;29(12):2371–2378. doi:10.1093/annonc/mdy454

5. Eisen T, Frangou E, Oza B, Ritchie AWS, Smith B, Kaplan R, et al. Adjuvant sorafenib for renal cell carcinoma at intermediate or high risk of relapse: results from the SORCE randomized phase III intergroup trial. J Clin Oncol. 2020;38(34):4064–4075. doi:10.1200/JCO.20.01800

6. Appleman LJ, Kim SE, Harris WB, Pal SK, Pins MR, Kolesar J, et al. Randomized, double-blind phase III study of pazopanib versus placebo in patients with metastatic renal cell carcinoma who have no evidence of disease after metastasectomy: ECOG-ACRIN E2810. J Clin Oncol. 2024;42(17):2061–2070. doi:10.1200/JCO.23.01544

7. González-Padilla DA, Subiela JD, Develtere D, Villacampa-Aubá F. Adjuvant therapy in renal cell carcinoma. Does the end justify the means? A reflection on KEYNOTE-564 and LITESPARK-022 benefits and harms. J Urol. 2026 (in press). doi:10.1097/JU.0000000000005283

8. Choueiri TK, Tomczak P, Park SH, Venugopal B, Ferguson T, Chang YH, et al. Adjuvant pembrolizumab after nephrectomy in renal-cell carcinoma. N Engl J Med. 2021;385(8):683–694. doi:10.1056/NEJMoa2106391

9. Choueiri TK, Tomczak P, Park SH, Venugopal B, Ferguson T, Symeonides SN, et al. Overall survival with adjuvant pembrolizumab in renal-cell carcinoma. N Engl J Med. 2024;390(15):1359–1371. doi:10.1056/NEJMoa2312695

10. Haas NB, Powles TB, Tomczak P, Park SH, Venugopal B, Symeonides SN, et al. Adjuvant pembrolizumab for the treatment of clear cell renal cell carcinoma: five-year results from the phase III KEYNOTE-564 study. Ann Oncol. 2026 (in press). doi:10.1016/j.annonc.2026.08.006

11. Pal SK, Uzzo R, Karam JA, Master VA, Donskov F, Suarez C, et al. Adjuvant atezolizumab versus placebo for patients with renal cell carcinoma at increased risk of recurrence following resection (IMmotion010): a multicentre, randomised, double-blind, phase 3 trial. Lancet. 2022;400(10358):1103–1116. doi:10.1016/S0140-6736(22)01658-0

12. Motzer RJ, Russo P, Grünwald V, Tomita Y, Zurawski B, Parikh O, et al. Adjuvant nivolumab plus ipilimumab versus placebo for localised renal cell carcinoma after nephrectomy (CheckMate 914): a double-blind, randomised, phase 3 trial. Lancet. 2023;401(10379):821–832. doi:10.1016/S0140-6736(22)02574-0

13. Motzer RJ, Bex A, Russo P, Tomita Y, Cutuli HJ, Rojas C, et al. Adjuvant nivolumab for localized renal cell carcinoma at high risk of recurrence after nephrectomy: Part B of the randomized, placebo-controlled, phase III CheckMate 914 trial. J Clin Oncol. 2025;43(2):189–200. doi:10.1200/JCO.24.00773

14. Choueiri TK, Motzer RJ, Karam JA, Yip W, Suárez C, Ye D, et al. Adjuvant pembrolizumab plus belzutifan for renal-cell carcinoma. N Engl J Med. 2026;395(1):32–43. doi:10.1056/NEJMoa2518245

15. Shao N, Su H, Ye D. Conditional disease-free survival in high-risk renal cell carcinoma treated with sunitinib. Aging (Albany NY). 2019;11(23):11490–11503. doi:10.18632/aging.102549

16. Cigliola A, Larcher A, Maiorano BA, Robesti D, Fallara G, Mercinelli C, et al. Adjuvant pembrolizumab plus belzutifan versus placebo in clear cell renal cell carcinoma: a reconstructed survival data analysis of the KEYNOTE-564 and the LITESPARK-022 trials. ESMO Open. 2026;11(10):108543. doi:10.1016/j.esmoop.2026.108543

17. Das A, Cotta BH, Msaouel P, Matin SF, Karam JA, Shapiro DD. A risk-adapted framework for adjuvant pembrolizumab plus belzutifan in resected clear cell renal cell carcinoma. Clin Genitourin Cancer. 2026:102679. doi:10.1016/j.clgc.2026.102679

18. Royston P, Parmar MK. Restricted mean survival time: an alternative to the hazard ratio for the design and analysis of randomized trials with a time-to-event outcome. BMC Med Res Methodol. 2013;13:152. doi:10.1186/1471-2288-13-152

19. Uno H, Claggett B, Tian L, Inoue E, Gallo P, Miyata T, et al. Moving beyond the hazard ratio in quantifying the between-group difference in survival analysis. J Clin Oncol. 2014;32(22):2380–2385. doi:10.1200/JCO.2014.55.2208

20. Allaf ME, Kim SE, Master V, McDermott DF, Harshman LC, Cole SM, et al. Perioperative nivolumab versus observation in patients with renal cell carcinoma undergoing nephrectomy (PROSPER ECOG-ACRIN EA8143): an open-label, randomised, phase 3 study. Lancet Oncol. 2024;25(8):1038–1052. doi:10.1016/S1470-2045(24)00211-0

21. Guyot P, Ades AE, Ouwens MJ, Welton NJ. Enhanced secondary analysis of survival data: reconstructing the data from published Kaplan-Meier survival curves. BMC Med Res Methodol. 2012;12:9. doi:10.1186/1471-2288-12-9

22. Liu N, Zhou Y, Lee JJ. IPDfromKM: reconstruct individual patient data from published Kaplan-Meier survival curves. BMC Med Res Methodol. 2021;21(1):111. doi:10.1186/s12874-021-01308-8

23. Royston P, Parmar MK. Flexible parametric proportional-hazards and proportional-odds models for censored survival data, with application to prognostic modelling and estimation of treatment effects. Stat Med. 2002;21(15):2175–2197. doi:10.1002/sim.1203

24. Hernán MA. The hazards of hazard ratios. Epidemiology. 2010;21(1):13–15. doi:10.1097/EDE.0b013e3181c1ea43

25. Ossato A, Gasperoni L, Del Bono L, Messori A, Damuzzo V. Efficacy of immune checkpoint inhibitors vs. tyrosine kinase inhibitors/everolimus in adjuvant renal cell carcinoma: indirect comparison of disease-free survival. Cancers (Basel). 2024;16(3):557. doi:10.3390/cancers16030557

26. Zhou Q, Liu J, Xie S. Adjuvant therapy in renal cell carcinoma: tyrosine kinase inhibitor versus immune checkpoint inhibitor. Medicine (Baltimore). 2024;103(22):e38329. doi:10.1097/MD.0000000000038329

