## Supplementary Material for "Does the Disease-free Survival Benefit of Adding Belzutifan to Adjuvant Pembrolizumab Persist Beyond the Planned Treatment Period? A Reconstructed Individual Patient Data Analysis of Phase 3 Renal Cell Carcinoma Trials"

Version 7, 15 October 2026. Changes from version 6: every supplementary figure (Figures S1–S16, with each panel of Figure S10 shown separately) is now embedded as an image directly under its legend; internal file paths, file names and script names were removed from the legends, table notes and text and replaced by plain descriptions. The pending items were completed: the initials of the investigator who performed the digitisations and reconstructions and of the second investigator (S-M4; Table S1, E6), the data-availability statement (S-M5, Software), the execution note on points added at censoring marks for the KEYNOTE-564 OS curve (S-M2) and the reason for excluding the 1971 van der Werf-Messing report (S-M1). No analysis was re-run and no number changed. Changes from version 5: the title now matches the manuscript. Table S1 opens with a statement on registration timing and data access before registration; row D10 now follows the deviations log; row E6 reports the second-investigator review; a new row E7 records that no ethics approval was applied for; row D8-g states that PROSPER OS was not performed. S-M1 states that the trial search was done by one reviewer, that the initial search (9–10 October 2026) was not recorded, and that the repeat search on 13 October 2026, after registration, is a post hoc confirmatory search that identified E2810; it also lists the sources searched for overlapping work (moved from the manuscript). S-M4, Table S5 and Figure S10 report the completed second-investigator review (14 October 2026): all 46 overlays were checked and no discrepancy was found; the final PROTECT OS curve keeps the authors' stricter judgement, now worded "did not pass the authors' visual check". S-M5 explains why the modified Hartung-Knapp adjustment was added for the ICI pooling and points to the tables and figures that report the flexible parametric and smoothed-hazard results. Table S6 adds the count of ICI and VEGFR-TKI sensitivity analyses whose modified Hartung-Knapp CI included 1 across both landmarks; Table S8 notes which comparisons have no 36-month value; Table S16 adds the number of events before and after each landmark. No analysis was re-run and no number changed. Changes from version 4: S-M5 now gives the correct number of DFS comparisons whose selected baseline df was at the upper limit after E2810 was added; Table S1 has a new row D10; Table S6 has a sentence describing where the LITESPARK-022 ratio CIs lie relative to the pooled point estimates; the window-matched analysis (D7 method) was extended post hoc to the KEYNOTE-564 NEJM 2021 and Lancet Oncol 2022 cut-offs (S-M6; Tables S3, S13 and S16); Table 1 of the manuscript is cited as version 3. No other number changed. Changes from version 3: status labels only, following decisions of 14 October 2026 (Table S1, D8-g and D9); no number changed and no analysis was re-run. E2810 OS is now labelled "accepted after adjudication" (reported HR 2.55 vs HR 2.11 derived exactly from the curve; reported model not stated), replacing "OS adjudication pending". The final PROTECT OS curve (Eur Urol 2021 Fig 1A) was not digitised again; it is labelled "did not pass visual check; shown for reference only, not used in any pooled analysis", replacing "pending re-digitisation". PROSPER OS is recorded as not reconstructed (raster resolution insufficient; reported values described in the Table S7 notes). The planned re-check of ESMO Congress 2026 abstracts was dropped; the supplementary search is reported as performed on 13 October 2026 (S-M1). Changes from version 2: ECOG-ACRIN E2810 (pazopanib vs placebo after metastasectomy), found by the re-run PubMed search, was added as a VEGFR-TKI main-analysis trial (deviation D9), and all analyses were re-run; this changes the VEGFR-TKI pooled results and the figures, and adds E2810 rows to the tables; a post hoc sensitivity analysis excluding E2810 from the VEGFR-TKI pooling was added (Table S6). All results not involving E2810 were reproduced exactly (Table S1, D9). E2810 OS is reported with the label "OS adjudication pending". Changes from version 1: analyses that were registered but had not been performed are now reported (deviation D8): the KEYNOTE-564 curves from earlier data cut-offs, the PROSPER curve with Day-1 events censored, the ASSURE, PROTECT, ATLAS and SORCE subset, population and assessment curves (SAP section 9, items 2–7; Tables S16–S18), substitution of these curves in the class-level pooling (Table S6), and OS for IMmotion010, S-TRAC, PROTECT, SORCE and CheckMate 914 part A (SAP section 8.5; Table S7); a post hoc modified Hartung-Knapp sensitivity analysis was added to every pooled analysis (Table S6); the PubMed trial-identification search is documented (S-M1); Tables S1–S5 are updated accordingly; figure panels now carry readable labels. No result reported in version 1 changed in version 2. All numbers in this document were copied from the project's analysis output tables, the reconstruction quality-control reports, the digitisation reports, the registered statistical analysis plan (SAP; OSF, https://osf.io/fk8hz, registered 12 October 2026), the deviations log and the execution-decision log. Hazard ratios (HRs), restricted mean survival time (RMST) differences and HR ratios are rounded to two decimals, RMST ratios to three decimals and percentage points (pp) to one decimal; full precision is available in the CSV files.

#### Contents


- Supplementary Methods S-M1 to S-M6
- Supplementary Table S1. Deviations from the registered protocol and execution notes
- Supplementary Table S2. Curve used for each comparison
- Supplementary Table S3. Feasibility of the truncation time and landmarks
- Supplementary Table S4. Execution decisions J1–J25
- Supplementary Table S5. Reconstruction accuracy for each curve

- Supplementary Table S6. Sensitivity analyses of the class-level pooling
- Supplementary Table S7. Overall survival (A, RMST and HR(t); B, descriptive piecewise HRs)
- Supplementary Table S8. Restricted mean disease-free survival time at 24 months and at the primary truncation time
- Supplementary Table S9. ΔRMST over the truncation time and post-treatment increment
- Supplementary Table S10. Piecewise Cox models
- Supplementary Table S11. Kaplan-Meier disease-free survival differences at 12–60 months
- Supplementary Table S12. Proportional hazards tests and flexible parametric time-varying HRs
- Supplementary Table S13. Window-matched post-treatment analysis (post hoc)
- Supplementary Table S14. LITESPARK-022: central review versus investigator assessment
- Supplementary Table S15. SORCE, 3 years versus 1 year of sorafenib (exploratory)
- Supplementary Table S16. KEYNOTE-564: five-year analysis and earlier data cut-offs
- Supplementary Table S17. PROSPER: main-text curve and curve with Day-1 events censored
- Supplementary Table S18. ASSURE, PROTECT, ATLAS and SORCE: subsets, populations and assessment methods
- Supplementary Figures S1–S16

Abbreviations: AIC = Akaike's information criterion; BICR = blinded independent central review; CI = confidence interval; ctrl = control arm; DFS = disease-free survival; df = degrees of freedom; exp = experimental arm; HIF-2α = hypoxia-inducible factor 2α; HR = hazard ratio; ICI = immune checkpoint inhibitor; IPD = individual patient data; IRC = independent review committee; ITT = intention to treat; KM = Kaplan-Meier; NR = not reached; OS = overall survival; PH = proportional hazards; pp = percentage points; QC = quality control; REML = restricted maximum likelihood; RFS = recurrence-free survival; RMST = restricted mean survival time; ΔRMST = between-arm difference in RMST (experimental minus control); SAP = statistical analysis plan; τ = truncation time; tvc = time-varying coefficient; VEGFR-TKI = vascular endothelial growth factor receptor tyrosine kinase inhibitor.

---

### Supplementary Methods

#### S-M1. Literature and registry search

#### Trial identification

Trials were identified in PubMed by one reviewer, without an independent second screener. The initial search (9–10 October 2026) was not recorded. The search was therefore repeated and documented on 13 October 2026, after registration of the analysis plan; this repeat search is a post hoc confirmatory search, and it identified one eligible trial that had not been considered, ECOG-ACRIN E2810 (Supplementary Table S1, D8 and D9). Details follow.

##### *Search and screening*

The eligible trials were first identified on 9–10 October 2026, but the search strings, date limits and screening counts of that initial search were not recorded. To provide a reproducible record, we therefore re-ran and documented a PubMed search on 13 October 2026, after registration of the statistical analysis plan; this search is a post hoc confirmatory search and is reported as such. No start date, language restriction or PubMed filter was applied, and the end date was 13 October 2026 (publication date). The primary search combined renal cell carcinoma terms (MeSH "Carcinoma, Renal Cell" and "Kidney Neoplasms", plus free-text synonyms) with adjuvant-setting terms (adjuvant, perioperative, postoperative, "after nephrectomy", MeSH "Chemotherapy, Adjuvant") and trial-design terms (Publication Types "Randomized Controlled Trial" and "Clinical Trial, Phase III", plus randomized/randomised and phase 3/phase III in title or abstract). Three supplementary searches were run: a search not relying on adjuvant-setting terms (nephrectomy, resection, disease-free or recurrence-free survival combined with the same disease and design terms, excluding records retrieved by the primary search); a known-item search for trial acronyms; and a search for randomised trials after metastasectomy or in patients with no evidence of disease. The exact search strings are given in Table S-M1.1. Searches were run through the PubMed application programming interface (NCBI E-utilities and a

PubMed connector); the primary search returned the same number of records through both interfaces. One reviewer screened all titles and abstracts and assessed the remaining reports against the prespecified eligibility criteria using the abstract, the PubMed record and, where available, the PubMed Central full text; there was no independent second screener. For the included trials, the presence of a Kaplan–Meier curve of the primary endpoint with a number-at-risk table was confirmed from the published full texts during data extraction.

##### *Results of the search*

The four searches yielded 1,227 records (1,097 after removal of duplicates). After title and abstract screening, 55 reports of 40 randomised or phase 3 trials of adjuvant or perioperative therapy were assessed for eligibility (Fig. S-M1.1). All 10 trials prespecified in the statistical analysis plan were retrieved (16 reports; Table S-M1.2). Twenty-nine trials (38 reports) were excluded: 18 trials of cytokines, vaccines, cell therapy, immunomodulators or a monoclonal antibody (girentuximab); 3 trials of other agents no longer in use (medroxyprogesterone acetate, tegafur–uracil and thalidomide); 1 trial of an mTOR inhibitor (EVEREST), which belongs to neither the immune checkpoint inhibitor nor the VEGFR tyrosine kinase inhibitor class defined in the analysis plan; 1 trial of postoperative radiotherapy; 1 trial without a peer-reviewed report of the primary results (RAMPART); 3 trials that were not phase 3 or not randomised (including the phase 2 RESORT trial); and 2 reports that described a trial without reporting results or without an abstract allowing assessment (Table S-M1.3). The search identified one additional phase 3 trial that met the population, intervention, comparator and publication criteria but had not been considered in the initial identification: ECOG-ACRIN E2810, which compared 1 year of pazopanib with placebo after complete metastasectomy (PMID 38531002). E2810 met all eligibility criteria; it was added to the main analyses as a VEGFR-TKI trial after protocol registration (deviation D9; Supplementary Table S1). No other eligible trial was identified. With E2810, 11 trials were included (Table S-M1.2).

*Table S-M1.1. PubMed search strings (run 13 October 2026; publication date 1800/01/01 to 2026/10/13)*

| Search | Purpose | Records |
|---|---|---|
| S1 | Primary search | 685 |
| S2 | Supplementary search without adjuvant-setting terms, excluding S1 | 226 |
| S3 | Known-item search for trial acronyms | 271 |
| S4 | Randomised trials after metastasectomy or with no evidence of disease | 45 |

S1:

```
("Carcinoma, Renal Cell"[MeSH Terms] OR "Kidney Neoplasms"[MeSH Terms] OR "renal cell carcinoma"[Title/Abstract] OR "renal cell cancer"[Title/Abstract] OR "kidney cancer"[Title/Abstract] OR "renal cancer"[Title/Abstract]) AND (adjuvant[Title/Abstract] OR perioperative[Title/Abstract] OR postoperative[Title/Abstract] OR "after nephrectomy"[Title/Abstract] OR "Chemotherapy, Adjuvant"[MeSH Terms]) AND ("Randomized Controlled Trial"[Publication Type] OR "Clinical Trial, Phase III"[Publication Type] OR randomized[Title/Abstract] OR randomised[Title/Abstract] OR "phase 3"[Title/Abstract] OR "phase III"[Title/Abstract])
```

S2:

```
(("Carcinoma, Renal Cell"[MeSH Terms] OR "Kidney Neoplasms"[MeSH Terms] OR "renal cell carcinoma"[Title/Abstract] OR "renal cell cancer"[Title/Abstract] OR "kidney cancer"[Title/Abstract] OR "renal cancer"[Title/Abstract]) AND ("Nephrectomy"[MeSH Terms] OR nephrectomy[Title/Abstract] OR resected[Title/Abstract] OR resection[Title/Abstract] OR "after surgery"[Title/Abstract] OR "risk of recurrence"[Title/Abstract] OR "disease-free survival"[Title/Abstract] OR "recurrence-free survival"[Title/Abstract]) AND ("Randomized Controlled Trial"[Publication Type] OR "Clinical Trial, Phase III"[Publication Type] OR "phase 3"[Title/Abstract] OR "phase III"[Title/Abstract])) NOT (("Carcinoma, Renal Cell"[MeSH Terms] OR "Kidney Neoplasms"[MeSH Terms] OR "renal cell carcinoma"[Title/Abstract] OR "renal cell cancer"[Title/Abstract] OR "kidney cancer"[Title/Abstract] OR "renal cancer"[Title/Abstract]) AND (adjuvant[Title/Abstract] OR perioperative[Title/Abstract] OR postoperative[Title/Abstract] OR "after nephrectomy"[Title/Abstract] OR "Chemotherapy, Adjuvant"[MeSH Terms]) AND ("Randomized Controlled Trial"[Publication Type] OR "Clinical Trial, Phase III"[Publication Type] OR randomized[Title/Abstract] OR randomised[Title/Abstract] OR "phase 3"[Title/Abstract] OR "phase III"[Title/Abstract]))
```

S3:

```
("renal cell carcinoma"[Title/Abstract] OR "renal cell cancer"[Title/Abstract] OR "kidney cancer"[Title/Abstract]) AND (S-TRAC[Title/Abstract] OR ASSURE[Title/Abstract] OR E2805[Title/Abstract] OR PROTECT[Title/Abstract] OR ATLAS[Title/Abstract] OR SORCE[Title/Abstract] OR "KEYNOTE-564"[Title/Abstract] OR IMmotion010[Title/Abstract] OR "CheckMate 914"[Title/Abstract] OR PROSPER[Title/Abstract] OR EA8143[Title/Abstract] OR "LITESPARK-022"[Title/Abstract] OR EVEREST[Title/Abstract] OR S0931[Title/Abstract] OR ARISER[Title/Abstract] OR RAMPART[Title/Abstract] OR SWOG[Title/Abstract]) AND (adjuvant[Title/Abstract] OR perioperative[Title/Abstract] OR nephrectomy[Title/Abstract])
```

S4:

```
(metastasectomy[Title/Abstract] OR "no evidence of disease"[Title/Abstract]) AND ("renal cell carcinoma"[Title/Abstract] OR "Carcinoma, Renal Cell"[MeSH Terms]) AND ("Randomized Controlled Trial"[Publication Type] OR randomized[Title/Abstract] OR randomised[Title/Abstract])
```

To reproduce a search in the PubMed web interface, append AND `("1800/01/01"[Date - Publication] : "2026/10/13"[Date - Publication])`.

*Fig. S-M1.1. Flow of records (PRISMA-style)*

```
Records identified through PubMed (n = 1,227)
  S1 n = 685; S2 n = 226; S3 n = 271; S4 n = 45
        |
        v
Duplicates removed (n = 130)
        |
        v
Records screened on title and abstract (n = 1,097)  --->  Records excluded (n = 1,042)
        |                                                   paediatric renal tumours, upper tract urothelial
        |                                                   carcinoma, surgical or anaesthetic trials, metastatic
        |                                                   disease, reviews and guidelines, secondary analyses
        |                                                   and commentaries on included trials
        v
Reports assessed for eligibility (n = 55 reports; 40 trials)  --->  Excluded (n = 38 reports; 29 trials)
        |                                                             cytokine, vaccine, cell therapy, immunomodulator
        |                                                             or monoclonal antibody: 18 trials
        |                                                             other agents no longer in use: 3
        |                                                             mTOR inhibitor (outside prespecified classes): 1
        |                                                             radiotherapy (not systemic therapy): 1
        |                                                             no peer-reviewed results report: 1
        |                                                             not phase 3 or not randomised: 3
        |                                                             protocol only or not assessable: 2
        |
        +--->  Eligible, not in prespecified set (n = 1 report; 1 trial: ECOG-ACRIN E2810) → added to the included trials (D9)
```

```
        |
        v
Trials included (n = 11 trials; 17 reports, including E2810 added under D9)
  primary and reference analyses: 10 trials (CheckMate 914 Parts A and B counted as one trial)
  sensitivity analysis only: 1 trial (PROSPER)
```

*Table S-M1.2. Included trials and identified reports*

| Trial | Comparison | PubMed PMIDs |
|---|---|---|
| LITESPARK-022 | Pembrolizumab plus belzutifan vs pembrolizumab plus placebo | 42384869 |
| KEYNOTE-564 | Pembrolizumab vs placebo | 34407342; 36055304; 38631003; 42648402 |
| IMmotion010 | Atezolizumab vs placebo | 36099926 |
| CheckMate 914 Part A | Nivolumab plus ipilimumab vs placebo | 36774933 |
| CheckMate 914 Part B | Nivolumab vs placebo | 39303200 |
| S-TRAC | Sunitinib vs placebo | 27718781 |
| ASSURE (ECOG-ACRIN E2805) | Sunitinib or sorafenib vs placebo | 26969090; 28278333 |
| PROTECT | Pazopanib vs placebo | 28902533; 33461782 |
| ATLAS | Axitinib vs placebo | 30346481 |
| SORCE | Sorafenib for 1 or 3 years vs placebo | 33052759 |
| PROSPER (ECOG-ACRIN EA8143) | Perioperative nivolumab vs observation | 38942046 |
| ECOG-ACRIN E2810 (added after registration; D9) | Pazopanib vs placebo after complete metastasectomy | 38531002 |

*Table S-M1.3. Trials assessed and excluded*

| Trial or report | Intervention vs control | PMIDs | Reason for exclusion |
|---|---|---|---|
| EVEREST (SWOG S0931) | Everolimus vs placebo | 37524096; 38811313 | mTOR inhibitor; outside the drug classes defined in the analysis plan (§3.4) |
| ARISER | Girentuximab vs placebo | 27787547 | Monoclonal antibody no longer in use (§3.2, item 3) |
| RAMPART | Durvalumab with or without tremelimumab vs active monitoring | 34538401; 34538402; 42766600 | No peer-reviewed report of primary results at the search date (§3.1, item 4) |
| ECOG/Intergroup | Interferon alfa-NL vs observation | 12663707 | Cytokine (§3.2, item 3) |
| Reniale vaccine trial | Autologous tumour cell vaccine vs no treatment | 14987883; 12779015 | Vaccine (§3.2, item 3) |
| Vitespen trial | HSPPC-96 vaccine vs observation | 18602688 | Vaccine (§3.2, item 3) |
| EORTC/NCRI | Fluorouracil, interferon alfa and interleukin-2 vs observation | 24074763 | Cytokine (§3.2, item 3) |
| GOIRC | Low-dose interleukin-2 plus interferon alfa vs observation | 25304727 | Cytokine (§3.2, item 3) |
| Cytokine Working Group | High-dose interleukin-2 vs observation | 12810695 | Cytokine (§3.2, item 3) |
| Pizzocaro et al. | Interferon alfa-2b vs observation | 11208835 | Cytokine (§3.2, item 3) |
| DGCIN | Interleukin-2, interferon alfa and fluorouracil vs observation | 15756254 | Cytokine (§3.2, item 3) |
| Hinotsu et al. | Natural interferon alfa vs observation | 22068465 | Cytokine (§3.2, item 3) |
| Delta-P | Interferon alfa-2a vs no treatment | 8448744 | Cytokine (§3.2, item 3) |
| French multicentre trial | Interleukin-2 or interferon alfa | 10851508 | Cytokine (§3.2, item 3) |
| Galligioni et al. | Autologous tumour cells plus BCG vs control | 8640706; 8368758 | Vaccine (§3.2, item 3) |
| Adler et al. | Hormono-immunotherapy vs hormonotherapy | 3330126 | Vaccine (§3.2, item 3) |
| Jurincic-Winkler et al. | Keyhole limpet haemocyanin vs control | 7941592 | Immunomodulator no longer in use (§3.2, item 3) |
| Zhan et al. | Dendritic cell–cytokine-induced killer cells or interferon alfa vs control | 23106871 | Cell therapy; not phase 3 (§3.1, item 1; §3.2, item 3) |
| Zhang et al. | Cytokine-induced killer cells vs control | 24382970 | Cell therapy; not phase 3 (§3.1, item 1) |
| Yang et al. | Enhanced cytokine-induced killer cells | 34540187 | Cell therapy; not phase 3 (§3.1, item 1) |
| Pizzocaro et al. | Medroxyprogesterone acetate vs no treatment | 2824861; 2934557; 6223819 | Agent no longer in use (§3.2, item 3) |
| Naito et al. | Tegafur–uracil vs no adjuvant therapy | 9179659 | Agent no longer in use (§3.2, item 3) |
| Margulis et al. | Thalidomide vs observation | 18950837 | Agent no longer in use; not phase 3 (§3.1, item 1; §3.2, item 3) |
| Zhang et al. | Herbal decoction plus sunitinib vs control | 37867340 | Not phase 3 (§3.1, item 1) |
| Copenhagen Renal Cancer Study Group | Postoperative radiotherapy vs observation | 3445125; 3553111 | Not systemic therapy (§3.1, item 3) |
| RESORT | Sorafenib vs observation after metastasectomy | 31542243; 33472450 | Phase 2 (§3.1, item 1) |
| Wang et al. | Penpulimab vs surveillance | 42431710 | Non-randomised phase 2 (§3.1, item 1) |

| Trial or report | Intervention vs control | PMIDs | Reason for exclusion |
|---|---|---|---|
| Italian multicentre interferon trial | Interferon alfa after nephrectomy | 2141713 | Protocol description only, no results (§3.1, item 4) |
| van der Werf-Messing et al. | Not assessable (no abstract) | 5141536 | Excluded: 1971 Dutch-language early report without abstract; the intervention could not be confirmed and no eligible report with a Kaplan-Meier curve and numbers at risk was identified. |

##### *Eligibility criteria*

Eligibility criteria were prespecified (SAP sections 3.1 and 3.2). Included were phase 3 randomised trials in patients with RCC after radical or partial nephrectomy (including patients with no evidence of disease after metastasectomy), in which the experimental arm received adjuvant or perioperative systemic therapy and the control arm received placebo, observation, or placebo added to a common background therapy. The trial had to be published in full in a peer-reviewed journal and to show at least one KM curve of the primary efficacy endpoint with a number-at-risk table. Excluded were trials available only as conference abstracts or posters, KM curves without a number-at-risk table or with insufficient resolution to pass the quality checks, and trials of treatments no longer in use (cytokines, vaccines and monoclonal antibodies such as girentuximab). No data were requested from authors or sponsors. EVEREST (everolimus, a mammalian target of rapamycin inhibitor) was not included because it belongs to neither drug class compared in this study (SAP section 3.4).

The prespecified trials and their roles were LITESPARK-022 (trial of primary interest; pembrolizumab plus belzutifan vs pembrolizumab plus placebo), KEYNOTE-564 (ICI, positive reference), IMmotion010 and CheckMate 914 parts A and B (ICI, negative references), S-TRAC, ASSURE, PROTECT, ATLAS and SORCE (VEGFR-TKI references), and PROSPER (sensitivity analyses only).

#### Supplementary search for overlapping work (13 October 2026)

To identify ongoing or unpublished work addressing the same question, PROSPERO, OSF Registries, preprint servers (medRxiv and the Europe PMC preprint index), 2026 meeting abstracts (ASCO Genitourinary Cancers Symposium, ASCO Annual Meeting and EAU Congress) and PubMed were searched on 13 October 2026. The search terms and numbers of records are listed below.

| Source | Interface | Query | Records |
|---|---|---|---|
| PROSPERO | PROSPERO 2.0 web search, all fields | `belzutifan` | 21 |
| PROSPERO | Same | `belzutifan AND adjuvant` | 6 |
| PROSPERO | Same | `renal AND restricted mean survival` | 27 |
| PROSPERO | Same | `adjuvant AND renal AND pembrolizumab` | 63 |
| PROSPERO | Same | `adjuvant AND renal AND reconstructed` | 27 |
| PROSPERO | Same | `adjuvant AND renal AND sunitinib` | 29 |
| PROSPERO | Same | `adjuvant AND renal AND hazards` | 12 |
| PROSPERO | Same | `adjuvant AND renal AND piecewise` | 1 (metastatic RCC; not relevant) |
| PROSPERO | Same | `adjuvant AND renal AND "time varying"` | 0 |
| PROSPERO | Same | `renal AND reconstructed` | 252 (too broad; first page screened) |
| OSF Registries | SHARE search API (registrations and registration components); details read with OSF API v2 | `belzutifan` | 1 (not relevant) |
| OSF Registries | Same | `adjuvant renal cell carcinoma` | 4 |
| OSF Registries | Same | `renal cell carcinoma restricted mean survival` | 1 |
| OSF Registries | Same | `reconstructed individual patient data renal` | 1 |
| OSF Registries | Same | `renal cell carcinoma reconstructed` | 1 |
| OSF Registries | Same | `restricted mean survival time adjuvant` | 2 |
| OSF Registries | Same | `renal cell carcinoma Kaplan-Meier` | 1 |
| OSF Registries | Same | `KEYNOTE-564` | 1 |
| OSF Registries | Same | `adjuvant pembrolizumab` | 2 (not relevant) |
| OSF Registries | Same | `kidney cancer restricted mean` | 1 (not relevant) |
| OSF Registries | Same | `HIF-2` | 4 (not relevant) |
| OSF Registries | Same | `LITESPARK`; `adjuvant kidney cancer`; `renal cell carcinoma time-varying hazard` | 0 each |
| medRxiv | medrxiv.org search | `belzutifan` (no date limit) | 7 (not relevant) |
| medRxiv | Same | `adjuvant renal cell carcinoma` (1 Jan 2025 to 13 Oct 2026) | 57 (titles screened; not relevant) |
| medRxiv | Same | `restricted mean survival time renal` (same dates) | 1,817 (first 75 titles screened; not relevant) |
| Europe PMC preprints (SRC:PPR; medRxiv, Research | Europe PMC | `belzutifan AND (adjuvant OR LITESPARK)` | 0 |

| Source | Interface | Query | Records |
|---|---|---|---|
| Square, Preprints.org; first posted 1 Jan 2025 to 13 Oct 2026) | | | |
| Europe PMC preprints | Same | `adjuvant AND ("renal cell carcinoma" OR "kidney cancer") AND ("restricted mean" OR RMST OR reconstructed OR "time-varying" OR piecewise OR "non-proportional" OR durab*)` | 4 (not relevant) |
| Europe PMC preprints | Same | `"KEYNOTE-564" OR "S-TRAC" OR "LITESPARK-022"` | 1 (case report) |
| Europe PMC preprints | Same | Broader adjuvant RCC query | 10 (not relevant) |
| ASCO (ASCO GU 2026; ASCO Annual Meeting 2026) | asco.org site search | `belzutifan adjuvant` | 7 |
| ASCO | Same | `LITESPARK-022` | 7 |
| ASCO | Same | `KEYNOTE-564` | 35 (first page screened) |
| ASCO | Same | `restricted mean survival renal`; `restricted mean survival adjuvant renal`; `reconstructed individual patient data adjuvant renal`; `adjuvant renal reconstructed` | 0 each |
| ASCO | Same | `adjuvant renal cell carcinoma disease-free survival meta-analysis` | 2 (2017) |
| EAU 2026 | Web search; meeting report read in full | `EAU 2026 abstract adjuvant renal cell carcinoma belzutifan reconstructed survival` | 1 meeting report (LITESPARK-022 exploratory outcomes; no overlap) |
| ESMO Congress 2026 (23–27 Oct 2026) | OncologyPRO; web search | Not searchable (abstracts not public on 13 Oct 2026) | — |
| PubMed | PubMed | `belzutifan adjuvant` | 23 |
| PubMed | Same | `LITESPARK-022` | 6 |
| PubMed | Same | `adjuvant renal cell carcinoma restricted mean survival time` | 6 |
| PubMed | Same | `adjuvant renal cell carcinoma reconstructed individual patient data` | 7 |
| PubMed | Same | `(pembrolizumab AND belzutifan) AND (adjuvant OR LITESPARK)` | 18 |
| PubMed | Same | `(adjuvant AND (renal OR kidney)) AND ("restricted mean" OR RMST OR "time-varying" OR "piecewise" OR "flexible parametric" OR "non-proportional" OR "reconstructed" OR "IPDfromKM" OR "Guyot")`, 2024 onward | 16 |
| PubMed | Same | `(adjuvant AND (renal cell carcinoma) AND (sunitinib OR pembrolizumab OR KEYNOTE-564 OR S-TRAC OR atezolizumab OR nivolumab)) AND (durability OR "treatment discontinuation" OR "off-treatment" OR "carryover" OR "hazard ratio over time" OR "landmark")`, 2024 onward | 37 |
| PubMed | Same | `(belzutifan OR LITESPARK) OR (adjuvant AND (renal cell carcinoma OR kidney cancer) AND (pembrolizumab OR sunitinib OR "disease-free survival" OR restricted OR reconstructed))`, entry date 1–13 Oct 2026 | 4 |

In PROSPERO, 15 registrations on adjuvant or perioperative RCC were opened and searched for the terms restricted mean, RMST, reconstruct, time-varying, piecewise, flexible parametric, proportional hazard, belzutifan, LITESPARK, discontinuation, durability and landmark. The direct medRxiv search was stopped when the site presented a human-verification page; the remaining terms were covered through the Europe PMC preprint index. All PubMed records not already known to the investigators were read at the level of title and abstract.

No identical published article, preprint, abstract or registration was found. Four registrations overlapped partially. PROSPERO CRD420261473448 (also OSF f3v5k; registered 9 August 2026) plans a reconstructed-IPD network meta-analysis of adjuvant and perioperative ICIs with 3- and 5-year RMST and tests of proportional hazards; it does not include belzutifan and does not examine post-treatment effects within trials. PROSPERO CRD420261454205 (registered 17 July 2026) plans a Bayesian network meta-analysis with reconstructed IPD, RMST and a multi-criteria decision analysis. PROSPERO CRD420261533360 (registered 8 October 2026) plans an RCT-only network meta-analysis including pembrolizumab plus belzutifan, without reconstructed IPD or RMST. PROSPERO CRD42024494438 (registered 4 January 2024) appears to correspond to a published reconstruction of adjuvant ICI and TKI trials (Zhou et al., Medicine 2024); this link was not confirmed on the registration page.

### S-M2. Curve extraction

#### Curve selection

For each comparison, one curve was prespecified for the main analysis by four rules applied in order (SAP section 5.1): (1) the ITT population (for PROTECT, the population randomised with a 600-mg starting dose); (2) the assessment method of the trial's primary analysis; (3) the latest published DFS analysis with full text available, a later analysis replacing an earlier one only if obtained before digitisation of that trial began; and (4) the publisher's final PDF in preference to the PubMed Central version. OS curves were selected by the same rules, with population taking precedence over analysis time. The curves used, and the reason for each departure from the curves listed in the registered plan, are given in Supplementary Tables S1 and S2.

#### Vector figures

Curves stored as vector drawing paths in the PDF were extracted with a purpose-written Python script (version 1.3; Python 3.14, PyMuPDF 1.28.2), without manual point selection. For each figure, a configuration file specified the PDF page, the bounding boxes of the plotting panel and of the number-at-risk table, the colour (and, where needed, line width) of each arm, the time unit and the survival scale. The script read all drawing segments in the panel and grouped them by colour. Axes were calibrated by matching tick labels (read as text from the PDF) to tick marks and fitting a least-squares linear transformation from page coordinates to time and survival; the residual of every calibration point was reported, and the maximum residuals are listed in Supplementary Table S2. Legend samples were identified and excluded as short isolated horizontal segments followed by text. Censoring marks were separated from the curve as short segments not joined to other segments (or, for figures in which marks and curves share colour and width, by line width, by duplicate-segment removal, or by recognising "+" marks), and were retained only if they lay on the curve. The curve was built from the remaining segment end points sorted by time, taking the running minimum of survival so that the curve was monotone non-increasing, keeping only the step corners and fixing the start at time 0 and survival 1. Numbers at risk were read from the text layer of the figure and compared cell by cell with the values transcribed in the extraction files (Supplementary Table S2). For visual checking, each extracted curve was drawn over a 300-dpi rendering of the original page.

#### Points added at censoring marks (`curve_points_at_censor`)

Vector extraction returns only the step corners, so points can be sparse in the tail of a curve. In that situation, the Guyot algorithm places the censored patients of an interval at the few available points, the estimated number at risk at a later drop becomes very small, and the event at that drop can be rounded to zero. The extraction script therefore has an option that adds a point on the horizontal segment at the time of each censoring mark, with the survival value of the curve at that time; the step shape is unchanged. The option was applied when the reconstruction from the standard input deviated from the digitised curve by more than 2 pp in the tail (execution note in the deviations log). It was applied to ATLAS Fig 1 (maximum deviation in the axitinib arm reduced from 4.56 to 0.64 pp) and to the KEYNOTE-564 DFS curve (placebo arm from 80.9 months, reduced from 5.45 to 1.6 pp), and later to the curves listed in Table S1 (D8-a, D8-e and D9; for E2810 the filled "+" censoring marks were read with a purpose-written script). It was also enabled for the KEYNOTE-564 OS curve, by analogy with ATLAS; without it, the OS reconstruction also met all numerical criteria. Points at censoring marks were added to the KEYNOTE-564 OS curve although the trigger (tail deviation >2 percentage points) was not met; the maximum deviation without them was 1.71 percentage points. This is recorded as an execution note.

#### Raster figure

The only curve available as a raster image was the CheckMate 914 part B comparison of nivolumab plus ipilimumab with placebo (JCO 2024 Data Supplement Fig S2), used only as an alternative comparison in sensitivity analyses. It was digitised with WebPlotDigitizer 4.8 using automatic colour extraction (averaging-window algorithm, $\Delta X = \Delta Y = 2$ pixels, colour distance 50), with the axis calibration points adjusted to the pixel positions of the tick marks to within 0.5 pixels. Because the censoring marks in this figure are vertical ticks of the same colour as the curve, colour extraction also captures the marks. A run-based anchor script (version 1.0) therefore converted pixels to time and survival using the calibration points, grouped points by pixel column, and treated a column containing a single point as lying on a horizontal part of the curve. Only runs of at least three consecutive single-point columns with a vertical spread of at most 0.75 pixels were accepted as horizontal anchors, so that partly hidden censoring ticks were not mistaken for the curve. The curve was built from the anchors alone, taking the running minimum of survival; each drop was placed at the multi-point column immediately before the first anchor of the new level. The number-at-risk table was transcribed manually.

Three further curves added under deviation D8 were available only as raster images: PROSPER Supplemental Fig 3 (768 × 480 pixels), the CheckMate 914 part A OS curve (Supplementary Fig S3) and the final PROTECT OS curve (Eur Urol 2021 Fig 1A; the PubMed Central author version was used because each panel of the publisher's composite image had a lower resolution, a departure from rule 4 of SAP section 5.1). WebPlotDigitizer was not available on the analysis computer for these figures, so the embedded images were extracted with PyMuPDF without resampling and the curves were traced by colour with purpose-written scripts (one for the PROSPER curves and one, version 1.2, for the raster overall survival curves); the traced points were then converted to curves with the same run-based anchor script as above. Axes were calibrated by least-squares fits to the tick marks, and the numbers at risk were transcribed manually from the images (Supplementary Table S2). The final PROTECT OS curve met the numerical criteria, but its shape after 60 months was judged coarse; under SAP section 5.3 it should have been digitised again once, but no manual re-digitisation was performed (decision of 14 October 2026; Table S1, D8-g); it did not pass the authors' visual check; shown for reference only, not used in any pooled analysis. The second investigator found no discrepancy in this overlay (S-M4), but the stricter judgement was retained.

### S-M3. Reconstruction and the censoring-informed modification

#### Standard reconstruction

IPD were reconstructed with the algorithm of Guyot et al. as implemented in the R package IPDfromKM 0.1.10, using the extracted curve, the number-at-risk table and the reported total number of events in each arm. Before reconstruction, the input curve was aligned with the number-at-risk table automatically, and every change was written to the QC report: a point was added on the curve at each number-at-risk time (otherwise `IPDfromKM::preprocess()` skips intervals without points); a horizontal end point slightly beyond the time at which the number at risk reached zero (a drawing artefact) was moved to just before that time; a sloping segment between two points (a step corner falling in a gap of a dashed line) was replaced by a horizontal segment followed by a drop; and a duplicate point was added when the curve ended with a drop. For a figure showing three arms with a shared control (ASSURE), all arms were reconstructed once and each comparison was checked separately. For comparison with the published HR, the HR was estimated with an unstratified Cox model, whereas most trials reported stratified HRs. The reported total number of events constrains only the last at-risk interval, so the event-count check is not fully independent of the input.

#### Censoring-informed reconstruction (post hoc)

The Guyot algorithm assumes that censoring is uniform within each at-risk interval. In trials with staggered accrual and short follow-up, censoring is concentrated late in each interval; the true number at risk at each drop is then larger than the algorithm assumes, and the number of events is systematically underestimated even when the curve and the number-at-risk table are reproduced exactly. This was the cause of the failed event-count criterion for the LITESPARK-022 investigator-assessed DFS curve (D1), the LITESPARK-022 OS curve (D2) and the PROSPER RFS curve (D4).

A modified reconstruction (purpose-written R code, version 1.1) changes only where censored observations are placed and otherwise follows the original Guyot algorithm; the curve pre-processing is identical to the standard method. Two sources of censoring information were used. When the figure prints cumulative numbers censored at each number-at-risk time (PROSPER Fig 2), the number censored in each interval was taken from the figure, the number of events in each interval was obtained by arithmetic (patients at risk at the start minus at risk at the next time point minus censored), events were allocated to the drops within the interval in proportion to the size of each drop and the number at risk at that drop (largest-remainder method), and censored times were placed at the censoring marks where present and uniformly otherwise. When the figure shows only censoring marks (LITESPARK-022 Fig 1A and Fig 2), the marks overlap and their number does not equal the number censored, so they were used only to shape the distribution of censored times within each interval (a window of 0.25 months on each side of each mark); the number censored per interval was found by the original iterative Guyot procedure so that the reconstructed number at risk at the next time point equalled the published value. Without either source, the method reduces to uniform censoring and should reproduce the standard result. As in the original algorithm, the reported total number of events constrained only the last interval.

The method was validated on three curves that had already passed with the standard method (SORCE Fig 2A without censoring information, and S-TRAC Fig 2 and IMmotion010 Fig 2A with censoring marks); all three again met the criteria (Supplementary Table S5, last three rows). These validation reconstructions were not used in any analysis (execution decision J23). For PROSPER, the number of events before the last interval is derived from the printed censored counts, so the event-count check is not independent and the HR and visual overlay are the main checks; in the observation arm, the reconstructed KM curve fell progressively below the digitised curve, with a maximum difference of about 5 pp at 45.8 months, where fewer than 30 patients remained at risk. The QC report for PROSPER shows that in most intervals the drop in the published curve was smaller than the minimum compatible with the published numbers of events and censored patients, so the two cannot be reproduced simultaneously.

### S-M4. Accuracy criteria and second-investigator review

The following criteria were prespecified (SAP section 5.3) and were not changed during the study:

| Item | Acceptance criterion |
|---|---|
| HR | Absolute difference between reconstructed and reported HR ≤ 0.05, and reconstructed HR within the reported CI |
| Number of events | Within 5% of the reported value in each arm |
| Median survival (if reported) | Difference ≤ 1 month, or within the reported CI |

| Item | Acceptance criterion |
|---|---|
| Landmark survival (if reported) | Difference ≤ 2 pp at each time point |
| Visual | No apparent deviation of the reconstructed KM curve overlaid on the original figure |

A curve that failed was to be digitised again once; if it still failed, it was used only in sensitivity analyses. When the publication was internally inconsistent, the number-at-risk and censoring information on the KM figure took precedence and the discrepancy was recorded (for example, SORCE arm B events are given as 245 in the text and 242 in the CONSORT diagram, and 242 was used; for CheckMate 914 part A, the total of 228 events in the final published version was used). For ASSURE, which reported 97.5% CIs, the published CI was used. For the comparison without a published HR (CheckMate 914 part B, nivolumab plus ipilimumab vs placebo), the HR item was not applicable and the check relied on the event counts printed on the figure and the number-at-risk table. For vector figures, re-digitisation reproduces the same coordinates, so a failure was attributed to the reconstruction step and handled as described in S-M3 and Supplementary Table S1.

The numerical items were computed automatically (the acceptance thresholds are fixed constants copied from the SAP). Each digitisation and reconstruction was performed by one investigator (H.-H.C.) and was checked visually and independently by a second investigator (C.-K.C.). The second investigator received two review packages: version 1 (13 October 2026; curves R01–R22) and version 2 (14 October 2026; curves R23–R46, the curves added under deviations D8 and D9). For each curve, the packages contained the extracted curve drawn over the original figure, the reconstructed KM curve drawn over the digitised curve, the QC values and the locations needing particular attention; the first investigator's visual judgements were withheld. The review was completed on 14 October 2026. The second investigator independently checked all 46 digitisation and reconstruction overlays and found no discrepancy (Supplementary Table S5, which gives the curve number of each overlay). For the final PROTECT OS curve (R42), the authors' stricter judgement was retained: it did not pass the authors' visual check and is shown for reference only (Supplementary Table S1, D8-g). The three validation reconstructions of the censoring-informed method (S-M3) were not included in the packages because they were not used in any analysis.

### S-M5. Statistical details

#### Coding and data handling

In every comparison, the experimental arm was coded 1 and the control arm 0; ΔRMST is experimental minus control, and HRs are experimental versus control. In LITESPARK-022, the experimental arm received pembrolizumab plus belzutifan and the control arm pembrolizumab plus placebo; in PROSPER, the control arm was observation. Records at time 0 (events or censored observations) were moved to 0.001 months in all analyses, because the counting-process format of the piecewise Cox model and the log-time scale of the flexible parametric model require positive times (J3; numbers per comparison in Supplementary Table S3).

#### Truncation time and its feasibility

The primary τ was 36 months, with 24 months as the secondary τ. τ was feasible for a comparison if, in each arm, the number at risk at τ was at least 10% of the number randomised. If 36 months was not feasible, the primary τ was 24 months. Feasibility was judged from the number-at-risk tables alone, without reading survival estimates from the curves. When τ fell between two time points of the table, the number at risk at the next table time point at or after τ was used (a lower bound), and τ was considered infeasible if this lower bound was below 10% (J1). The program determined the primary τ for every comparison and checked it against the determinations listed in the registered plan; all agreed (J2). For the ΔRMST(τ) curves, τ ran from 6 months in 3-month steps up to the maximum feasible τ (Supplementary Table S3). RMST and its pointwise asymptotic 95% CI were estimated at each τ with `survRM2::rmst2` (unadjusted); no simultaneous confidence bands were computed (J5).

#### Landmarks

The landmark separating the on-treatment and post-treatment periods was the end of planned treatment, set directly in months and not aligned with the number-at-risk time points (SAP section 6.2). The main landmark was 12 months for LITESPARK-022, KEYNOTE-564, IMmotion010, S-TRAC, ASSURE, PROTECT, ATLAS (shortest planned duration), SORCE 1 year, E2810 (52 weeks) and PROSPER, 6 months for CheckMate 914 parts A and B, and 36 months for SORCE 3 years. The sensitivity landmark was 15 months for most trials (later, because recurrence is detected only at the next imaging visit), 9 months for CheckMate 914, 36 months for ATLAS (longest planned duration), 12 months for SORCE 3 years (actual median exposure about 10.7 months) and 10 months for PROSPER (the authors' own landmark) (Supplementary Table S3).

#### Post-treatment increment

The post-treatment ΔRMST increment was ΔRMST at the maximum feasible $\tau$ minus ΔRMST at the main landmark, and the increment per year was the increment divided by the number of years between the two (SAP section 7.2; J6). Because ΔRMST($\tau$) is the area between the two survival curves up to $\tau$, its slope at any time equals the difference in survival at that time; the increment per year is therefore the average DFS difference (expressed in months per year) over the post-treatment period.

#### Piecewise Cox models

Follow-up was split at the landmark with `survival::survSplit`, and the model `Surv(tstart, tstop, event) ~ trt + trt:late` was fitted with Efron's method for ties, where `late` indicates the post-treatment period (J8). No main effect of period was included, because period is a function of time and is absorbed by the baseline hazard. The on-treatment HR is exp(b1), the post-treatment HR is exp(b1 + b2) with variance Var(b1) + Var(b2) + 2Cov(b1, b2), and the ratio of post-treatment to on-treatment HRs is exp(b2); model-based standard errors and Wald CIs were used. The number of events in each arm and period is reported to show the stability of each estimate (Supplementary Table S10).

#### Kaplan-Meier differences and proportional hazards

Between-arm differences in KM DFS were computed at 12, 24, 36, 48 and 60 months, within the time points feasible under the same 10% rule, with 95% CIs from Greenwood standard errors assuming independent arms and a normal approximation (J7). Proportional hazards were tested with the global Schoenfeld residual test (`survival::cox.zph`, default `transform = "km"`) in a Cox model with treatment as the only covariate; $P < 0.05$ was taken as evidence against proportional hazards (J9). Log(−log) plots are shown in Supplementary Figures S5 and S6.

#### Flexible parametric models

Royston-Parmar models with a time-varying treatment effect were fitted with `rstpm2::stpm2(Surv(time, event) ~ trt, df = baseline df, tvc = list(trt = time-varying df))` (J10–J12). All 15 combinations of baseline df 1–5 and time-varying df 1–3 were fitted, and the model with the smallest AIC was selected; ties (AIC difference < $10^{-6}$) were to be broken by the smaller number of parameters and then the smaller baseline df, but none occurred. A model was regarded as not converged, and excluded from selection, if fitting failed, the optimiser returned a non-zero convergence code, a coefficient had a non-finite standard error or refitting failed, or its maximised log-likelihood was more than 0.5 below that of the nested df 1/tvc 1 model. The prespecified sensitivity model with baseline df 3 and time-varying df 2 was fitted for every comparison. The selected baseline df was at the upper limit of 5 for 11 DFS comparisons; the prespecified range was not extended. HR(t) and its 95% CI were evaluated every 0.25 months from 1 month to the maximum feasible $\tau$; tables report HR(t) at the landmark and at the maximum feasible $\tau$ (Supplementary Table S12), and Supplementary Figures S2–S4, S12 and S13 show the full HR(t) curves.

#### Smoothed hazards

Hazard functions for each arm were smoothed with `muhaz::muhaz` using its defaults (local bandwidth, boundary correction at both ends, Epanechnikov kernel) from 0 to the maximum feasible $\tau$, for DFS only (J14); they are descriptive and are shown in Supplementary Figures S7, S8 and S15.

#### Class-level pooling

For each comparison, the effect size was the interaction coefficient b2 (the log of the ratio of post-treatment to on-treatment HRs) with its model-based standard error. These were pooled separately for VEGFR-TKI and ICI trials with `meta::metagen` using a random-effects model, REML estimation of between-study variance and the Hartung-Knapp adjustment of the CI, without the ad hoc variance correction (default in meta 8.5.0); only random-effects estimates are reported, with $\tau^2$ and $I^2$ (J13). E2810, identified after registration, entered the VEGFR-TKI main pooling under SAP section 3.1 (deviation D9); because all its patients had M1 disease resected to no evidence of disease, a post hoc sensitivity analysis excluded it. One comparison per trial entered the main pooling (ASSURE: sunitinib; SORCE: 1 year; CheckMate 914 part B: nivolumab), and the alternative comparisons were substituted one at a time and, for the two TKI trials, together, in sensitivity analyses (J17; Supplementary Table S6). LITESPARK-022 was not pooled with either class, and no formal test between LITESPARK-022 and either class was performed.

The curves added under deviation D8 (SAP section 9, items 2 and 4–7) were substituted one at a time for the main curve of the same trial, at the main landmarks: each earlier KEYNOTE-564 cut-off for the five-year curve; ASSURE clear cell (sunitinib) for ASSURE ITT (sunitinib); PROTECT ITT800mg and ITTAll for PROTECT ITT600mg; ATLAS highest-risk (IRC), highest-risk (investigator) and ITT (investigator) for ATLAS ITT (IRC); and SORCE Leibovich high-risk and clear cell (1 year) for SORCE ITT (1 year). The PROSPER curve with Day-1 events censored was added to the ICI pooling, as for the other PROSPER curves (J18).

Post hoc modified Hartung-Knapp analysis. With few studies and similar estimates, the Hartung-Knapp variance can be smaller than the variance of the conventional random-effects estimate, giving an implausibly narrow CI. The Hartung-Knapp variance equals $q/\Sigma w$, where $w = 1/(SE^2 + \tau^2)$ and $q = \Sigma w(y - \mu)^2/(k - 1)$. With only four ICI comparisons, the estimates were less dispersed than their standard errors imply ($q = 0.146$), which narrows the ordinary Hartung-Knapp interval; the modified adjustment, which does not allow the adjusted standard error to fall below the unadjusted one, was therefore added post hoc. As a post hoc sensitivity analysis (decision K5; suggested during internal review and agreed on 13 October 2026), every pooled analysis was repeated with q replaced by $q^* = \max(1, q)$, keeping the t distribution with $k - 1$ degrees of freedom (Knapp and Hartung 2003). In meta 8.5.0 this corresponds to `metagen(..., method.random.ci = "HK", adhoc.hakn.ci = "se")`, which uses the conventional random-effects standard error whenever the Hartung-Knapp standard error is smaller; the option `"IQWiG6"` compares widths against a DerSimonian-Laird interval and `"ci"` takes the wider of two intervals using normal quantiles, so neither is identical to $q^* = \max(1, q)$. Every modified interval was checked against a direct calculation of $q^*$ (agreement within $10^{-8}$).

#### Software

Analyses were run in R 4.6.0 (2026-04-24, ucrt) on Windows 11 x64 (build 26200) with survival 3.8-6, survRM2 1.0-4, rstpm2 1.7.1, muhaz 1.2.6.5 and meta 8.5-0; reconstruction used IPDfromKM 0.1.10; figures used ggplot2 4.0.3, patchwork 1.3.2 and ragg 1.5.2. The full analysis was run with the master analysis script on 13 October 2026, 23:49:53 to 00:01:10 (after E2810 was added; the first run on 13 October 2026, 11:55:12 to 12:04:28, is archived); the complete session information is stored with the analysis outputs. Vector extraction used Python 3.14 with PyMuPDF 1.28.2, numpy and matplotlib; raster digitisation used WebPlotDigitizer 4.8. The analyses added under deviation D8 and the modified Hartung-Knapp analysis were run with a separate SAP-completion script (14 October 2026, 00:01:23 to 00:09:21; session information stored with the analysis outputs), with the same software versions. That script reuses the function definitions of the main analysis scripts and writes only new files; before computing the new curves it recomputed every existing comparison and reproduced all existing result tables, including the bootstrap results, to within $10^{-8}$, and it confirmed by checksum that no existing output file had changed. `set.seed(20261013)` was set at the start of the analysis although the main analyses contain no random step (J24). The reconstructed individual patient data and analysis code are available from the corresponding author on request and will be deposited on the Open Science Framework at publication (SAP section 11).

### S-M6. Bootstrap and window-matched analyses (post hoc)

#### Bootstrap CI for the post-treatment increment (D5)

The registered plan defined the post-treatment increment without a CI. After registration, 95% CIs were added for ΔRMST at the landmark, ΔRMST at the maximum feasible τ, the increment and the increment per year. Within each trial, reconstructed patients were resampled with replacement separately within each arm, keeping the arm sizes fixed (B = 2000; `set.seed(20261013)` at the start of the script). The landmark and the maximum feasible τ were kept fixed and not re-determined in each resample. CIs are percentile intervals (2.5th and 97.5th percentiles; quantile type 7); point estimates are those from the original reconstructed data. The proportion of resamples with an increment above zero is also reported. No resample failed (a failure would have been a resample in which RMST could not be computed at τ because no patient in an arm remained at risk); all 2000 resamples were valid for every comparison.

#### Window-matched increment (D7)

Post-treatment follow-up differed widely between trials, so increments per year averaged over different windows could not be compared. In a second post hoc analysis, each comparison was restricted to the window from its main landmark to 18 months after the landmark, the post-treatment follow-up available in LITESPARK-022 (12 to 30 months). A window was to be declared infeasible, and not shortened, if landmark plus 18 months exceeded the comparison's maximum feasible τ; no window was infeasible. For each window, the increment was ΔRMST at the end minus ΔRMST at the start, and the increment per year was the increment divided by 1.5. The KM DFS differences at the start and end of the window were also computed, with Greenwood CIs, and the change in the difference with a bootstrap CI. Bootstrap CIs for ΔRMST at the start and end, the increment, the increment per year and the change in the KM difference used the same method, number of resamples and seed as D5. For LITESPARK-022, the window coincides with the whole post-treatment period, and the per-year increment and its CI are identical to those of D5, because both scripts produce the same resampling sequence. Drug class was used only for grouping in tables and figures; no pooling or test across trials was performed.

#### Curves added under deviation D8

The post-treatment increments of the curves added under D8 (Tables S16–S18) used the same bootstrap method and number of resamples, but the seed `set.seed(20261013)` was set before the resampling of each curve rather than once per script (K1), so that adding a trial later (for example ECOG-ACRIN E2810) cannot change the intervals of other curves; no resample failed. On 14 October 2026 the window-matched analysis (D7 method) was extended post hoc to the two earlier KEYNOTE-564 cut-offs (NEJM 2021 and Lancet Oncol 2022), with the same window (12 to 30 months), method and number of resamples and the per-curve seed (K1); it was not extended to the other curves added under D8. For the NEJM 2021 curve the window end equals the maximum feasible τ (30 months), so its increment and CI are identical to the post-treatment increment in Table S16 (verified by the script); no resample failed.

---

### Supplementary Tables

#### Supplementary Table S1. Deviations from the registered protocol and execution notes

**Registration timing.** The plan was registered at 21:10 on 12 October 2026. Before registration, the published Kaplan-Meier figures of the candidate trials had been viewed when the hypotheses were formulated, and curve coordinates from three VEGFR-TKI trials (S-TRAC, PROTECT, and SORCE) had been extracted to test the extraction software; reconstruction and effect estimation began after registration.

Registered plan: SAP version 4, registered on the Open Science Framework on 12 October 2026 (https://osf.io/fk8hz). Source: deviations log.

| ID | Date | SAP section | What happened | Decision | Potential impact | Status |
|---|---|---|---|---|---|---|
| D1 | 12 Oct 2026 | 5.1 (main curve for LITESPARK-022: Fig 1A, investigator); 5.3 (failed curves move to sensitivity analyses) | The standard reconstruction of Fig 1A undercounted events by 7.0% in the belzutifan arm and 3.3% in the placebo arm, exceeding 5%; the HR (0.700 vs reported 0.72), landmark survival (within 0.4 pp) and numbers at risk met the criteria. Re-extraction from the vector paths gave identical coordinates. The cause was judged to be non-uniform censoring within at-risk intervals (staggered accrual with censoring concentrated late), not digitisation or coding error. | Fig 1A moved to sensitivity analyses (SAP 5.3). The main analysis used Fig 1B (BICR), which met all numerical criteria (HR 0.756 vs 0.76; events −2.9%/−0.8%; landmarks within 0.3 pp). The SAP ranks a blinded assessment next when the primary assessment cannot be used but does not name the replacement curve, so this is recorded as a deviation. A post hoc sensitivity analysis reconstructed Fig 1A with the censoring-informed method. | The main-analysis assessment differs from the trial's primary (investigator) assessment; the reported HRs are similar (0.72 vs 0.76). | Deviation; censoring-informed method post hoc |
| D2 | 12 Oct 2026 | 5.3; 8.5 | The standard reconstruction of LITESPARK-022 OS (Fig 2) undercounted deaths by 7.9% and 6.1% (reported 38 vs 49 deaths, so a shortfall of about 3 deaths already exceeds 5%); HR and landmarks met the criteria. | OS curve designated for descriptive and sensitivity use (SAP 5.3); LITESPARK-022 OS was prespecified as descriptive only. | None on inference. | Deviation |
| D3 | 12 Oct 2026 | 5.3 (landmark within 2 pp; rule for internally inconsistent publications) | PROTECT Fig A1A: the 12-month DFS in the publication's Table A1 is 85%/76%, whereas the reconstruction at 12.0 months gives 87.8%/79.1% (difference 2.8/3.1 pp). The published curve drops sharply | Judged to be an inconsistency between the figure and the table of the publication; under the prespecified rule, the KM figure took precedence and the curve was accepted. | None expected. | Application of a prespecified rule, reported for transparency |

| ID | Date | SAP section | What happened | Decision | Potential impact | Status |
|---|---|---|---|---|---|---|
| | | | at 12.0–12.6 months (1-year imaging visit), and the published 1-year values correspond to the curve after this drop (about 12.1–12.6 months); there, the reconstructed and digitised curves differ by 0.1 pp. The 24-, 36- and 48-month landmarks, HR (0.930 vs 0.936) and events (about −2%) met the criteria. | | | |
| D4 | 12 Oct 2026 | 5.3; 9 (item 3: PROSPER used only in sensitivity analyses) | PROSPER Fig 2 (corrected version): events 118 vs 125 (−5.6%) and 124 vs 133 (−6.8%), exceeding 5%; HR 0.955 vs 0.94, numbers at risk 18/18 matched and close overlay. Same cause as D1. The publication is also internally inconsistent: the observation arm drops by about 34 patients at time 0, whereas 27 Day-1 events are reported; the drawn curve was retained. | (A) Standard reconstruction retained in sensitivity analyses, labelled as having failed the QC. (C) Reconstruction repeated with the censoring-informed method using the censored counts printed for each interval, to be used (labelled as a method outside the plan) if it met the criteria; it did (Supplementary Table S5). | None (PROSPER is not in the main analyses). | Deviation; method post hoc |
| D5 | 13 Oct 2026 | 7.2 (post-treatment increment defined without a CI) | The increment is the central measure for the primary question but had only a point estimate. | Within-trial nonparametric bootstrap stratified by arm (resampling reconstructed patients) for 95% percentile CIs of the increment and the increment per year (S-M6). | Adds uncertainty estimates; point estimates unchanged. | Post hoc addition |
| D6 | 13 Oct 2026 | 9 (item 1: sensitivity landmarks); 8.6 (class pooling) | The plan did not state whether the sensitivity landmarks also apply to the class-level pooling (J16). | Pooling with the sensitivity landmarks retained and labelled as a consequence of SAP 9 item 1; the main pooling remains the SAP 8.6 version with the main landmarks. | None on the main pooling. | Execution decision recorded as a deviation |
| D7 | 13 Oct 2026 | 7.2 (increment from the main landmark to the maximum feasible τ) | Post-treatment follow-up differs widely (1.5 years in LITESPARK-022; 2–7 years in TKI trials), so per-year increments over different windows cannot be compared. | Window-matched analysis: for every comparison, only the window from the main landmark to 18 months after it; ΔRMST increment, increment per year (bootstrap CI as in D5) and KM DFS differences at the start and end of the window (S-M6). | Adds a comparison of patterns over equal windows. | Post hoc addition |
| E1 | 12 Oct 2026 | 5.2 (raster figures; details not specified) | CheckMate 914 part B Fig S2 (raster): digitised with WebPlotDigitizer 4.8 (version 5 requires an account), automatic colour extraction (averaging window, ΔX = ΔY = 2 pixels, colour distance 50), axis calibration corrected to within 0.5 pixels; because censoring ticks share the curve colour, only runs of horizontal anchors were used (run-based anchor script; S-M2). | Methodological detail. | — | Execution note (not a deviation) |
| E2 | 12–13 Oct 2026 | 5.2 (reconstruction input; details not specified) | With vector extraction (corners only), sparse tail points can make the Guyot algorithm round events to zero. | Option `curve_points_at_censor`: when the standard input deviated from the digitised | — | Execution note (not a deviation) |

| ID | Date | SAP section | What happened | Decision | Potential impact | Status |
|---|---|---|---|---|---|---|
| | | | | curve by more than 2 pp in the tail, a point with the curve value was added at each censoring time (step shape unchanged). Applied to ATLAS Fig 1 (axitinib 4.56 → 0.64 pp) and the KEYNOTE-564 five-year DFS curve (placebo from 80.9 months, 5.45 → 1.6 pp; confirmed 13 Oct 2026). | | |
| E3 | 13 Oct 2026 | 5.1 (rule 4: publisher's final PDF preferred) | CheckMate 914 part B, nivolumab vs placebo: the registered plan listed the PubMed Central raster version (3951 px) of JCO 2024 Fig 2; the publisher's final PDF, in which the figure is a vector graphic, was obtained on 13 Oct 2026. | Same figure, better version, used under rule 4. | — | Execution note (not a deviation) |
| E4 | 13 Oct 2026 | 5.1 (rule 3: a later analysis obtained before digitisation replaces the earlier one) | The KEYNOTE-564 five-year analysis (Ann Oncol 2026; vector figures) was obtained on 13 Oct 2026, before digitisation of KEYNOTE-564 began. | Used as the main curve under rule 3; NEJM 2024 Fig 2A became a sensitivity curve. | — | Execution note (not a deviation) |
| E5 | 12 Oct 2026 | 5.1 (rule 4) | ASSURE: the registered plan listed the PubMed Central raster version (636 × 375 pixels) of Lancet 2016 Fig 2; the publisher's final PDF (vector) was obtained on 12 Oct 2026. | Same figure, better version, used under rule 4. | — | Execution note (not a deviation) |
| E6 | 12 Oct 2026; review completed 14 Oct 2026 | 5.2 (independent verification by a second investigator) | Second investigator (C.-K.C.) identified for visual verification of all digitisations and reconstructions. Review package version 1 (curves R01–R22) and version 2 (R23–R46, the curves added under D8 and D9) were checked independently, without the first investigator's visual judgements. | All 46 overlays checked; no discrepancy found (S-M4; Table S5). For the final PROTECT OS curve (R42), the authors' stricter judgement was retained: did not pass the authors' visual check; shown for reference only (D8-g). | None; no curve changed. | Execution note; completed |
| E7 | 15 Oct 2026 | 2 (ethics review not expected to be required; exemption to be obtained if required) | Only published aggregate data (Kaplan-Meier figures and published summary statistics) were used; no actual patient-level data were requested or accessed (the reconstructed IPD are derived from the published figures). | Ethics committee (institutional review board) approval was not applied for, because only published aggregate data were used; an application will be made only if a journal requires it. | None. | Execution note (not a deviation) |
| D8 | 13 Oct 2026 | 8.5 (OS of each trial); 9, items 2–7; Appendix A (search strings) | When the main analyses were completed, the sensitivity curves of SAP section 9 items 2–7 and the OS curves of five trials had not been reconstructed or analysed, and the PubMed trial-identification search strings had not been recorded; this was found while preparing the supplement and during internal review. | All items performed and reported in this supplement; items that proved impossible are listed with the pages searched and the reason (below). The PubMed search was re-run on 13 Oct 2026 and reported with that date (S-M1), without reconstructing the earlier record. | Adds sensitivity analyses; main analyses unchanged. | Deviation (late execution); completed |

| ID | Date | SAP section | What happened | Decision | Potential impact | Status |
|---|---|---|---|---|---|---|
| D8-a | 13 Oct 2026 | 9, item 2 | KEYNOTE-564 NEJM 2021 Fig 1A, Lancet Oncol 2022 Fig 2A and NEJM 2024 Fig 2A (vector) reconstructed; all met the criteria (Table S5). Points at censoring marks were added to the first two under the existing trigger (tail deviation > 2 pp), not to NEJM 2024. The Lancet Oncol 2022 24-month landmark appears only as quoted in NEJM 2024 (secondary source) and was not used for the QC judgement. | Analysed within trial (Table S16) and substituted for the five-year curve in the ICI pooling (Table S6). | — | Completed |
| D8-b | 13 Oct 2026 | 9, item 3; 5.2 | PROSPER Supplemental Fig 3 (Day-1 events censored; raster 768 × 480): the embedded image was extracted with PyMuPDF and traced with a purpose-written colour-tracing script instead of WebPlotDigitizer; numbers at risk and censored were transcribed from the image. It met the criteria (Table S5). The observation arm still drops at time 0 (the inconsistency noted in D4), and the image is truncated at 50.4 months. | Reported separately (Table S17) and added to the ICI pooling (Table S6), as prespecified for a curve passing QC. | — | Completed |
| D8-c | 13 Oct 2026 | 9, item 4; 5.3 (rule for internally inconsistent publications) | ASSURE clear cell (Lancet 2016 Supplemental Fig 1; first-ranked source, so the JAMA Oncol 2017 high-risk subset was not needed): HRs met the criteria, but reconstructed events exceeded the printed counts by 7.5–10.1% and a median did not match. The printed event counts were shown to be lower than the minimum compatible with the drawn curve and the number-at-risk table, and the text reports the placebo median inconsistently (event-bound calculation in the ASSURE reconstruction records). | Accepted after adjudication under the rule for internally inconsistent publications, as in D3 (decided 13 Oct 2026); labelled in all tables. | Sensitivity analysis only. | Application of a prespecified rule |
| D8-d | 13 Oct 2026 | 9, item 5; 5.3 | PROTECT Fig A1B (ITT800mg) and Fig A1C (ITTAll): HRs and events met the criteria; the 60-month placebo landmark (both) and the 12-month placebo landmark (A1C) did not, each at a sharp drop at an imaging visit, where the reconstruction and the digitised curve differed by ≤ 0.6 pp (the pattern of D3). | D3 extended to both curves (decided 13 Oct 2026); accepted and labelled. | Sensitivity analysis only. | Application of a prespecified rule |
| D8-e | 13 Oct 2026 | 9, item 6 | SAP item 6 (“highest-risk subset and investigator assessment”) was read as two sensitivity analyses; three | Each substituted for ATLAS in the TKI pooling. In the highest-risk IRC curve no placebo event occurred after 36 months, | — | Completed |

| ID | Date | SAP section | What happened | Decision | Potential impact | Status |
|---|---|---|---|---|---|---|
| | | | curves were reconstructed (Fig 3A highest risk, IRC; Fig 3B highest risk, investigator; Fig S2 ITT, investigator), all meeting the criteria; points at censoring marks were added to Fig 3A and 3B under the existing trigger. | so the piecewise model at the 36-month sensitivity landmark is not estimable. | | |
| D8-f | 13 Oct 2026 | 9, item 7 | SORCE Appendix Fig A1 (Leibovich high risk) and A2 (clear cell), 1 and 3 years vs placebo: all HRs met the criteria; subset event counts, landmarks and medians were not reported, so QC consisted of the HR and the visual check, supplemented post hoc by comparison with the 10-year RMST in the publication's Table A3. | 1-year curves substituted in the TKI pooling; 3-year curves analysed within trial only. | — | Completed |
| D8-g | 13 Oct 2026 | 8.5; 5.1 | OS reconstructed and analysed for IMmotion010 (Fig 3), S-TRAC (updated OS, Eur Urol Fig 3; latest full-text analysis), PROTECT (JCO 2017 Fig 4A, ITT600mg; the population rule precedes the time-point rule; deaths per arm not reported), SORCE (Fig 3A and 3B) and CheckMate 914 part A (Supplementary Fig S3; no HR reported, so the HR item was not applicable). The final PROTECT OS curve (Eur Urol 2021 Fig 1A, ITTAll; sensitivity) used the PubMed Central author version (departure from rule 4) and met the numerical criteria, but digitisation after 60 months was coarse. ATLAS and CheckMate 914 part B published no OS KM curve (all pages of the main texts and supplements checked). The PROSPER OS curve (raster) could not be extracted automatically. | OS analysed per SAP 8.5 (Table S7). Final PROTECT OS: no manual re-digitisation (decision of 14 Oct 2026); analysed but labelled "did not pass the authors' visual check; shown for reference only, not used in any pooled analysis" (K7). ATLAS and CheckMate 914 part B: reported values only (Table S7 notes). PROSPER OS: not performed (not reconstructed): raster resolution insufficient (768×480 px; overlapping curves); reported values described (Table S7 notes; decision of 14 Oct 2026). | None on the main analyses. | Partly completed (closed 14 Oct 2026) |
| D8-h | 13 Oct 2026 | 8.6 | Internal review noted that the four ICI estimates are very similar, so the ordinary Hartung-Knapp interval may be too narrow. | Post hoc modified Hartung-Knapp sensitivity analysis ($q^* = \max(1, q)$) for every pooled analysis (S-M5; Table S6). | Adds a more conservative interval; main estimates unchanged. | Post hoc addition |
| D8-i | 13 Oct 2026 | 3.1–3.3 | The re-run PubMed search identified one eligible phase 3 trial not considered initially: ECOG-ACRIN E2810 (pazopanib vs placebo after metastasectomy). | Added (D9). | See D9. | Resolved by D9 |
| D9 | 13 Oct 2026 | 3.1 (eligibility includes M1 NED after resection); Appendix A | E2810 (J Clin Oncol 2024; PMID 38531002): randomised, double-blind phase 3 trial of | Included as a VEGFR-TKI main-analysis trial under SAP 3.1 (decision of 13 Oct 2026, | Changes the VEGFR-TKI pooled results; all other results reproduced exactly (verified | Deviation (trial added after registration); OS accepted after adjudication (14 Oct 2026) |

| ID | Date | SAP section | What happened | Decision | Potential impact | Status |
|---|---|---|---|---|---|---|
| | | | pazopanib vs placebo for 52 weeks after complete metastasectomy, with a DFS KM curve and number-at-risk table. It was missed by the initial search (its abstract does not use “adjuvant” or “postoperative”) and had never been considered. Curves: publisher’s final PDF, Fig 2 (DFS) and Fig 3 (OS), vector graphics (rule 4); the PubMed Central raster version was used only for cross-checking. DFS assessor not stated; the only DFS curve was used (rule 2). τ = 36 months and landmarks 12/15 months were set by the SAP 6.1/6.2 rules (not listed in the registered tables). | option A); all analyses re-run; post hoc sensitivity analysis excluding E2810 from the VEGFR-TKI pooling because its population (100% M1 NED; much higher risk) differs from the other trials. DFS reconstruction (points at censoring marks) met all criteria (Table S5). OS: every drop is one death and death counts match, but the reconstructed unstratified HR differs from the published 2.55 (model not stated) by more than 0.05, so the HR item failed. The unstratified HR derived exactly from the curve is 2.11 (reconstructions 2.07–2.14), so the difference reflects the unstated published model rather than reconstruction error; by analogy with D3 (published numbers inconsistent with the figure; the KM curve takes precedence), OS was accepted after adjudication (decision of 14 Oct 2026; reported HR 2.55 vs HR 2.11 derived exactly from the curve; reported model not stated) and is labelled accordingly. | table by table against the archived pre-E2810 outputs). | |
| D10 | 14 Oct 2026 | 1.3 (secondary objective, item 2: describe which drug class LITESPARK-022 more closely resembles); 8.6 (item 4: position of the LITESPARK-022 ratio CI relative to the two pooled class estimates); 7.2 (interpretive statement that a converging benefit yields an increment approaching zero) | Internal review and an independent pre-submission review noted that the post/on-treatment HR ratio does not measure whether the absolute benefit persists (ratios near 2 occurred both in the first interim analysis of KEYNOTE-564, whose benefit persisted, and in SORCE and PROTECT, whose benefit faded), so it cannot be used to classify LITESPARK-022; and that the cumulative post-treatment increment equals the average DFS difference over the window, so it is the current slope, not the cumulative increment, that approaches zero when a benefit converges. | SAP 1.3 item 2 and SAP 8.6 item 4 are reported descriptively only (Table S6 states where the LITESPARK-022 ratio CIs lie relative to the pooled point estimates), without a classification. The SAP 7.2 interpretive statement is corrected here; the increment is still reported as registered. | The main message is the conservative version adopted on 13 Oct 2026; no number changes. | Deviation (14 Oct 2026) |

### Supplementary Table S2. Curve used for each comparison

Sources: reconstruction configuration files, Table 1 of the manuscript (version 3), the deviations log (execution notes), the number-at-risk files and the digitisation QC reports.

| Trial | Comparison | Endpoint | Source publication; figure | Assessment | Population (n exp/ctrl) | Data cut-off | Figure type; extraction | Number-at-risk interval | Axis calibration, max residual x/y (pt) | Number-at-risk cells matching extraction file | Use |
|---|---|---|---|---|---|---|---|---|---|---|---|
| LITESPARK-022 | Pembrolizumab + belzutifan vs pembrolizumab + placebo | DFS | Choueiri, N Engl J Med 2026; Fig 1B | BICR | ITT (921/920) | 23 Aug 2025 (IA1) | Vector (PyMuPDF) | every 6 mo (0–42) | 0.001/0.005 | 16/16 cells | Main analysis (D1) |
| LITESPARK-022 | Same | DFS | Choueiri, N Engl J Med 2026; Fig 1A | Investigator (trial primary) | ITT (921/920) | 23 Aug 2025 (IA1) | Vector (PyMuPDF) | every 6 mo (0–42) | 0.001/0.005 | 16/16 cells | Sensitivity (failed QC with standard method; censoring-informed reconstruction post hoc) (D1) |
| LITESPARK-022 | Same | OS | Choueiri, N Engl J Med 2026; Fig 2 | — | ITT (921/920) | 23 Aug 2025 (IA1) | Vector (PyMuPDF) | every 6 mo (0–42) | 0.001/0.005 | 16/16 cells | Descriptive only (D2) |
| KEYNOTE-564 | Pembrolizumab vs placebo | DFS | Haas, Ann Oncol 2026 (5-year analysis); Fig 1A | Investigator | ITT (496/498) | 25 Sep 2024 (IA4) | Vector (PyMuPDF); points added at censoring marks | every 5 mo (0–90) | 0.047/0.043 | 38/38 cells | Main analysis |
| KEYNOTE-564 | Same | OS | Haas, Ann Oncol 2026 (5-year analysis); Fig 1B | — | ITT (496/498) | 25 Sep 2024 (IA4) | Vector (PyMuPDF); points added at censoring marks | every 5 mo (0–90) | 0.041/0.037 | 38/38 cells | Secondary endpoint |
| IMmotion010 | Atezolizumab vs placebo | DFS | Pal, Lancet 2022; Fig 2A | Investigator | ITT (390/388) | 3 May 2022 | Vector (PyMuPDF) | every 3 mo (0–60) | 0.001/0.008 | 42/42 cells | Main analysis |
| CheckMate 914 part A | Nivolumab + ipilimumab vs placebo | DFS | Motzer, Lancet 2023; Fig 2 | BICR | ITT (405/411) | 28 Jun 2022 | Vector (PyMuPDF) | every 3 mo (0–54) | 0.000/0.278 | 38/38 cells | Main analysis |
| CheckMate 914 part B | Nivolumab vs placebo | DFS | Motzer, J Clin Oncol 2024; Fig 2 (publisher's final PDF) | BICR | ITT (411/208) | 28 Sep 2023 | Vector (PyMuPDF) | every 3 mo (0–42) | 0.080/0.080 | 30/30 cells | Main analysis |
| CheckMate 914 part B | Nivolumab + ipilimumab vs placebo | DFS | Motzer, J Clin Oncol 2024; Data Supplement Fig S2 | BICR | ITT (206/208) | 28 Sep 2023 | Raster (WebPlotDigitizer 4.8 + run-based anchors) | every 3 mo (0–42) | — (raster; calibration points corrected to within 0.5 px) | Transcribed manually; consistent with extraction file at 0, 24 and 36 mo | Alternative comparison (sensitivity pooling only) |
| S-TRAC | Sunitinib vs placebo | DFS | Ravaud, N Engl J Med 2016; Fig 2 | BICR | ITT (309/306) | 7 Apr 2016 | Vector (PyMuPDF) | every 12 mo (0–108) | 0.001/0.006 | 20/20 cells | Main analysis |
| ASSURE | Sunitinib vs placebo | DFS | Haas, Lancet 2016; Fig 2 (publisher's final PDF; three arms) | Not reported | ITT (647/647) | 27 Aug 2015 | Vector (PyMuPDF) | every 12 mo (0–96) | 0.001/0.000 | 27/27 cells | Main analysis |
| ASSURE | Sorafenib vs placebo | DFS | Same figure | Not reported | ITT (649/647) | 27 Aug 2015 | Vector (PyMuPDF) | every 12 mo (0–96) | 0.001/0.000 | 27/27 cells | Alternative comparison |
| ASSURE | Sunitinib vs placebo; sorafenib vs placebo | OS | Haas, Lancet 2016; Fig 3 | — | ITT (647/649/647; sunitinib/sorafenib/placebo) | 27 Aug 2015 | Vector (PyMuPDF) | every 12 mo (0–96) | 0.001/0.000 | 27/27 cells | Secondary endpoint |
| PROTECT | Pazopanib (600-mg start) vs placebo | DFS | Motzer, J Clin Oncol 2017; Appendix Fig A1A (follow-up DFS analysis) | Investigator | ITT600mg (571/564) | 15 Oct 2016 | Vector (PyMuPDF) | every 6 mo (0–60) | 0.312/0.184 | 22/22 cells | Main analysis (D3) |
| ATLAS | Axitinib vs placebo | DFS | Gross-Goupil, Ann Oncol 2018; | IRC | ITT (363/361) | 10 Oct 2017 (interim) | Vector (PyMuPDF); | every 12 mo (0–60) | 0.001/0.000 | 12/12 cells | Main analysis |

| Trial | Comparison | Endpoint | Source publication; figure | Assessment | Population (n exp/ctrl) | Data cut-off | Figure type; extraction | Number-at-risk interval | Axis calibration, max residual x/y (pt) | Number-at-risk cells matching extraction file | Use |
|---|---|---|---|---|---|---|---|---|---|---|---|
| | | | Fig 1 | | | | points added at censoring marks | | | | |
| SORCE | Sorafenib 1 yr vs placebo | DFS | Eisen, J Clin Oncol 2020; Fig 2B | Investigator | ITT (642/430) | Not reported | Vector (PyMuPDF) | every 24 mo (0–144) | 0.005/0.005 | 14/14 cells | Main analysis |
| SORCE | Sorafenib 3 yr vs placebo | DFS | Eisen, J Clin Oncol 2020; Fig 2A | Investigator | ITT (639/430) | Not reported | Vector (PyMuPDF) | every 24 mo (0–144) | 0.005/0.005 | 14/14 cells | Alternative comparison; exploratory 3 yr vs 1 yr |
| E2810 (ECOG-ACRIN) | Pazopanib vs placebo | DFS | Appleman, J Clin Oncol 2024; Fig 2 (publisher's final PDF) | Not reported | ITT, M1 NED after metastasectomy (66/63) | Jul 2021 | Vector (PyMuPDF); points added at censoring marks | every 12 mo (0–96) | 0.003/0.004 | 18/18 cells | Main analysis (added after registration; D9) |
| E2810 (ECOG-ACRIN) | Same | OS | Appleman, J Clin Oncol 2024; Fig 3 | — | ITT (66/63) | Jul 2021 | Vector (PyMuPDF); points added at censoring marks | every 12 mo (0–96) | 0.003/0.003 | 18/18 cells | Secondary endpoint; OS accepted after adjudication (reported HR 2.55 vs HR 2.11 derived exactly from the curve; reported model not stated — see Table S1, D9) |
| PROSPER | Perioperative nivolumab vs observation | RFS | Allaf, Lancet Oncol 2024; Fig 2 (corrected version, 24 Nov 2025) | Investigator | RFS population (381/399) | 24 May 2023 | Vector (PyMuPDF) | every 6 mo (0–48) | 0.000/0.000 | 18/18 cells | Sensitivity only (D4) |
| KEYNOTE-564 | Pembrolizumab vs placebo | DFS | Choueiri, N Engl J Med 2021; Fig 1A (first interim analysis) | Investigator | ITT (496/498) | 14 Dec 2020 | Vector (PyMuPDF); points added at censoring marks | every 5 mo (0–45) | 0.001/0.005 | 20/20 cells | Sensitivity (SAP 9, item 2) |
| KEYNOTE-564 | Same | DFS | Powles, Lancet Oncol 2022; Fig 2A (30-month update) | Investigator | ITT (496/498) | 14 Jun 2021 | Vector (PyMuPDF); points added at censoring marks | every 5 mo (0–50) | 0.001/0.159 | 22/22 cells | Sensitivity (SAP 9, item 2) |
| KEYNOTE-564 | Same | DFS | Choueiri, N Engl J Med 2024; Fig 2A (third interim analysis) | Investigator | ITT (496/498) | 15 Sep 2023 | Vector (PyMuPDF) | every 5 mo (0–75) | 0.002/0.005 | 32/32 cells | Sensitivity (SAP 9, item 2; registered main curve before E4) |
| PROSPER | Perioperative nivolumab vs observation | RFS | Allaf, Lancet Oncol 2024; Supplemental Fig 3 (Day-1 events censored) | Investigator | RFS population (381/399) | 24 May 2023 | Raster 768×480 (colour tracing script + run-based anchor script) | every 6 mo (0–48) | — (raster; colour tracing; tick-fit residuals x ≤ 0.035 mo, y ≤ 0.0022) | Transcribed manually from the image; identical to the extraction file cell by cell | Sensitivity (SAP 9, item 3); reported separately |
| ASSURE | Sunitinib vs placebo; sorafenib vs placebo | DFS | Haas, Lancet 2016; Supplemental Fig 1 (clear cell; three arms) | Not reported | Clear cell (542/540/540; sunitinib/sorafenib/placebo) | 27 Aug 2015 | Vector (PyMuPDF) | every 12 mo (0–96) | 0.005/0.004 | 27/27 cells | Sensitivity (SAP 9, item 4); accepted after adjudication (D8) |
| PROTECT | Pazopanib (800-mg start) vs placebo | DFS | Motzer, J Clin Oncol 2017; Appendix Fig A1B | Investigator | ITT800mg (198/205) | 15 Oct 2016 | Vector (PyMuPDF) | every 6 mo (0–66) | 0.341/0.185 | 24/24 cells | Sensitivity (SAP 9, item 5); accepted after adjudication (D8) |
| PROTECT | Pazopanib (all doses) vs | DFS | Motzer, J Clin Oncol 2017; | Investigator | ITTAll (769/769) | 15 Oct 2016 | Vector (PyMuPDF) | every 6 mo (0–66) | 0.347/0.188 | 24/24 cells | Sensitivity (SAP 9, item 5); |

| Trial | Comparison | Endpoint | Source publication; figure | Assessment | Population (n exp/ctrl) | Data cut-off | Figure type; extraction | Number-at-risk interval | Axis calibration, max residual x/y (pt) | Number-at-risk cells matching extraction file | Use |
|---|---|---|---|---|---|---|---|---|---|---|---|
| | placebo | | Appendix Fig A1C | | | | | | | | accepted after adjudication (D8) |
| ATLAS | Axitinib vs placebo | DFS | Gross-Goupil, Ann Oncol 2018; Fig 3A | IRC | Highest-risk subset (209/200) | 10 Oct 2017 (interim) | Vector (PyMuPDF); points added at censoring marks | every 12 mo (0–60) | 0.004/0.000 | 12/12 cells | Sensitivity (SAP 9, item 6) |
| ATLAS | Same | DFS | Gross-Goupil, Ann Oncol 2018; Fig 3B | Investigator | Highest-risk subset (209/200) | 10 Oct 2017 (interim) | Vector (PyMuPDF); points added at censoring marks | every 12 mo (0–60) | 0.004/0.000 | 12/12 cells | Sensitivity (SAP 9, item 6) |
| ATLAS | Same | DFS | Gross-Goupil, Ann Oncol 2018; Supplementary Fig S2 | Investigator | ITT (363/361) | 10 Oct 2017 (interim) | Vector (PyMuPDF) | every 12 mo (0–60) | 0.000/0.000 | 12/12 cells | Sensitivity (SAP 9, item 6) |
| SORCE | Sorafenib 1 yr vs placebo | DFS | Eisen, J Clin Oncol 2020; Appendix Fig A1 (right) | Investigator | Leibovich high risk (300/206) | Not reported | Vector (PyMuPDF) | every 24 mo (0–144) | 0.007/0.297 | 14/14 cells | Sensitivity (SAP 9, item 7) |
| SORCE | Sorafenib 3 yr vs placebo | DFS | Eisen, J Clin Oncol 2020; Appendix Fig A1 (left) | Investigator | Leibovich high risk (295/206) | Not reported | Vector (PyMuPDF) | every 24 mo (0–144) | 0.005/0.004 | 14/14 cells | Sensitivity (SAP 9, item 7); within-trial only |
| SORCE | Sorafenib 1 yr vs placebo | DFS | Eisen, J Clin Oncol 2020; Appendix Fig A2 (right) | Investigator | Clear cell (534/361) | Not reported | Vector (PyMuPDF) | every 24 mo (0–144) | 0.006/0.005 | 14/14 cells | Sensitivity (SAP 9, item 7) |
| SORCE | Sorafenib 3 yr vs placebo | DFS | Eisen, J Clin Oncol 2020; Appendix Fig A2 (left) | Investigator | Clear cell (550/361) | Not reported | Vector (PyMuPDF) | every 24 mo (0–144) | 0.006/0.005 | 14/14 cells | Sensitivity (SAP 9, item 7); within-trial only |
| IMmotion010 | Atezolizumab vs placebo | OS | Pal, Lancet 2022; Fig 3 | — | ITT (390/388) | 3 May 2022 | Vector (PyMuPDF) | every 3 mo (0–60) | 0.067/0.007 | 42/42 cells | Secondary endpoint (SAP 8.5) |
| S-TRAC | Sunitinib vs placebo | OS | Motzer, Eur Urol 2018; Fig 3 (updated OS) | — | ITT (309/306) | 31 Jan 2017 | Vector (PyMuPDF) | every 12 mo (0–120) | 0.002/0.001 | 22/22 cells | Secondary endpoint (SAP 8.5) |
| PROTECT | Pazopanib (600-mg start) vs placebo | OS | Motzer, J Clin Oncol 2017; Fig 4A (interim OS) | — | ITT600mg (571/564) | 15 Oct 2016 | Vector (PyMuPDF) | every 6 mo (0–60) | 0.064/0.057 | 22/22 cells | Secondary endpoint (SAP 8.5; population rule precedes time point) |
| PROTECT | Pazopanib (all doses) vs placebo | OS | Motzer, Eur Urol 2021; Fig 1A (final OS; PubMed Central author version) | — | ITTAll (769/769) | 15 Apr 2019 | Raster, PubMed Central JPEG (colour extraction script, version 1.2) | every 10 mo (0–100) | — (raster, PubMed Central author-manuscript JPEG; colour extraction; residuals not reported) | Transcribed manually from the image; identical to the extraction file cell by cell | Sensitivity; did not pass the authors' visual check; shown for reference only, not used in any pooled analysis |
| SORCE | Sorafenib 3 yr vs placebo | OS | Eisen, J Clin Oncol 2020; Fig 3A | — | ITT (638/430) | Not reported | Vector (PyMuPDF) | every 24 mo (0–144) | 0.005/0.004 | 14/14 cells | Secondary endpoint (SAP 8.5) |
| SORCE | Sorafenib 1 yr vs placebo | OS | Eisen, J Clin Oncol 2020; Fig 3B | — | ITT (641/430) | Not reported | Vector (PyMuPDF) | every 24 mo (0–144) | 0.004/0.004 | 14/14 cells | Secondary endpoint (SAP 8.5) |
| CheckMate 914 part A | Nivolumab + ipilimumab vs placebo | OS | Motzer, Lancet 2023; Supplementary | — | ITT (405/411) | 28 Jun 2022 | Raster, native PNG (colour extraction script, | every 3 mo (0–60) | — (raster; colour extraction; tick-fit residuals x 0.006 | Transcribed manually from the image; | Secondary endpoint (SAP 8.5) |

| Trial | Comparison | Endpoint | Source publication; figure | Assessment | Population (n exp/ctrl) | Data cut-off | Figure type; extraction | Number-at-risk interval | Axis calibration, max residual x/y (pt) | Number-at-risk cells matching extraction file | Use |
|---|---|---|---|---|---|---|---|---|---|---|---|
| | | | Fig S3 | | | | version 1.2) | | mo, y 0.20 pp) | identical to the extraction file cell by cell | |

Data cut-off and population as reported in each publication; "Not reported" means the publication does not state it. Axis calibration residuals are the largest distances (in PDF points; 1 pt = 1/72 inch) between a tick label's position and the fitted calibration line. The number-at-risk cells extracted from the figure were compared with the values transcribed independently in the extraction files. For ASSURE, the DFS assessor was not stated, and no central radiological review was described. CheckMate 914 part B appears online in 2024 and in print in J Clin Oncol 2025;43:189–200. Curves added under deviation D8 are listed after the main curves; curves that could not be analysed are listed in Supplementary Table S1 (D8-g).

#### Supplementary Table S3. Feasibility of the truncation time and landmarks

Source: analysis output tables (feasibility and window-matched increment).

| Comparison | Endpoint | Role | n (exp/ctrl) | % at risk at 24 mo (exp/ctrl) | % at risk at 36 mo (exp/ctrl) (grid point used, mo) | Primary τ (mo) | Primary τ in registered plan (mo) | Maximum feasible τ (mo) | Main landmark (mo) | Sensitivity landmark (mo) | Window-matched interval, D7 (mo) | Records at t = 0 set to 0.001 mo |
|---|---|---|---|---|---|---|---|---|---|---|---|---|
| LITESPARK-022, BICR (Fig 1B) | DFS | Main | 921/920 | 46.0/43.9 | 3.7/4.0 (36) | 24 | 24 | 30 | 12 | 15 | 12→30 | 0 |
| KEYNOTE-564 | DFS | Main | 496/498 | 71.8/64.7 | 62.3/56.8 (40) | 36 | 36 | 75 | 12 | 15 | 12→30 | 0 |
| IMmotion010 | DFS | Main | 390/388 | 62.6/59.8 | 49.7/48.2 (36) | 36 | 36 | 48 | 12 | 15 | 12→30 | 0 |
| CheckMate 914 part A | DFS | Main | 405/411 | 55.3/55.2 | 22.0/21.9 (36) | 36 | 36 | 39 | 6 | 9 | 6→24 | 0 |
| CheckMate 914 part B, nivolumab | DFS | Main | 411/208 | 26.8/27.4 | 1.7/1.0 (36) | 24 | 24 | 27 | 6 | 9 | 6→24 | 0 |
| CheckMate 914 part B, nivolumab + ipilimumab | DFS | Alternative | 206/208 | 26.2/27.4 | 0.5/1.0 (36) | 24 | 24 | 27 | 6 | 9 | 6→24 | 0 |
| S-TRAC | DFS | Main | 309/306 | 56.0/59.2 | 49.5/49.0 (36) | 36 | 36 | 72 | 12 | 15 | 12→30 | 0 |
| ASSURE, sunitinib | DFS | Main | 647/647 | 61.4/64.0 | 52.2/55.6 (36) | 36 | 36 | 72 | 12 | 15 | 12→30 | 0 |
| ASSURE, sorafenib | DFS | Alternative | 649/647 | 65.2/64.0 | 55.0/55.6 (36) | 36 | 36 | 72 | 12 | 15 | 12→30 | 0 |
| PROTECT | DFS | Main | 571/564 | 60.6/61.7 | 49.4/50.0 (36) | 36 | 36 | 48 | 12 | 15 | 12→30 | 2 |
| ATLAS | DFS | Main | 363/361 | 50.1/45.4 | 22.3/19.9 (36) | 36 | 36 | 36 | 12 | 36 | 12→30 | 0 |
| SORCE, sorafenib 1 yr | DFS | Main | 642/430 | 77.1/72.8 | 65.7/65.6 (48) | 36 | 36 | 96 | 12 | 15 | 12→30 | 4 |
| SORCE, sorafenib 3 yr | DFS | Alternative | 639/430 | 75.7/72.8 | 62.4/65.6 (48) | 36 | 36 | 96 | 36 | 12 | 36→54 | 0 |
| E2810 (M1 NED after metastasectomy) | DFS | Main | 66/63 | 28.8/31.7 | 21.2/15.9 (36) | 36 | 36 | 48 | 12 | 15 | 12→30 | 0 |
| LITESPARK-022, investigator (Fig 1A), standard reconstruction | DFS | Sensitivity | 921/920 | 49.0/45.3 | 3.8/4.1 (36) | 24 | 24 | 30 | 12 | 15 | 12→30 | 0 |
| LITESPARK-022, | DFS | Sensitivity | 921/920 | 49.0/45.3 | 3.8/4.1 (36) | 24 | 24 | 30 | 12 | 15 | 12→30 | 0 |

| Comparison | Endpoint | Role | n (exp/ctrl) | % at risk at 24 mo (exp/ctrl) | % at risk at 36 mo (exp/ctrl) (grid point used, mo) | Primary τ (mo) | Primary τ in registered plan (mo) | Maximum feasible τ (mo) | Main landmark (mo) | Sensitivity landmark (mo) | Window-matched interval, D7 (mo) | Records at t = 0 set to 0.001 mo |
|---|---|---|---|---|---|---|---|---|---|---|---|---|
| investigator (Fig 1A), censoring-informed reconstruction | | | | | | | | | | | | |
| PROSPER, standard reconstruction | RFS | Sensitivity | 381/399 | 39.1/34.1 | 19.9/18.5 (36) | 36 | 36 | 36 | 12 | 10 | 12→30 | 73 |
| PROSPER, censoring-informed reconstruction | RFS | Sensitivity | 381/399 | 39.1/34.1 | 19.9/18.5 (36) | 36 | 36 | 36 | 12 | 10 | 12→30 | 78 |
| SORCE, sorafenib 3 yr vs 1 yr | DFS | Exploratory | 639/642 | 75.7/77.1 | 62.4/65.7 (48) | 36 | 36 | 96 | 36 | 12 | — | 2 |
| KEYNOTE-564 | OS | Main | 496/498 | 94.8/93.0 | 91.1/86.9 (40) | 36 | 36 | 75 | — | — | — | 0 |
| ASSURE, sunitinib | OS | Main | 647/647 | 83.9/87.9 | 77.7/82.4 (36) | 36 | 36 | 84 | — | — | — | 0 |
| ASSURE, sorafenib | OS | Main | 649/647 | 86.6/87.9 | 79.2/82.4 (36) | 36 | 36 | 84 | — | — | — | 0 |
| LITESPARK-022, standard reconstruction | OS | Descriptive | 921/920 | 68.5/69.3 | 7.6/7.5 (36) | 24 | 24 | 30 | — | — | — | 0 |
| LITESPARK-022, censoring-informed reconstruction | OS | Descriptive | 921/920 | 68.5/69.3 | 7.6/7.5 (36) | 24 | 24 | 30 | — | — | — | 0 |
| E2810 [OS accepted after adjudication (reported HR 2.55 vs HR 2.11 derived exactly from the curve; reported model not stated — see Table S1, D9)] | OS | Main | 66/63 | 81.8/85.7 | 72.7/77.8 (36) | 36 | 36 | 72 | — | — | — | 0 |
| KEYNOTE-564, NEJM 2021 (cut-off Dec 2020) | DFS | Sensitivity | 496/498 | 30.4/29.1 | 0.2/0.2 (40) | 24 | — (rule applied) | 30 | 12 | 15 | 12→30 | 0 |
| KEYNOTE-564, Lancet Oncol 2022 (cut-off Jun 2021) | DFS | Sensitivity | 496/498 | 51.4/46.2 | 7.5/6.6 (40) | 24 | — (rule applied) | 33 | 12 | 15 | 12→30 | 0 |
| KEYNOTE-564, NEJM 2024 (cut-off Sep 2023) | DFS | Sensitivity | 496/498 | 71.6/64.3 | 61.9/56.6 (40) | 36 | 36 | 60 | 12 | 15 | — | 0 |
| PROSPER, Day-1 events censored (Suppl Fig 3) | RFS | Sensitivity | 381/399 | 39.1/34.1 | 19.9/18.5 (36) | 36 | — (rule applied) | 36 | 12 | 10 | — | 0 |
| ASSURE clear cell, sunitinib | DFS | Sensitivity | 542/540 | 61.8/63.0 | 51.1/54.3 (36) | 36 | — (rule applied) | 72 | 12 | 15 | — | 5 |

| Comparison | Endpoint | Role | n (exp/ctrl) | % at risk at 24 mo (exp/ctrl) | % at risk at 36 mo (exp/ctrl) (grid point used, mo) | Primary τ (mo) | Primary τ in registered plan (mo) | Maximum feasible τ (mo) | Main landmark (mo) | Sensitivity landmark (mo) | Window-matched interval, D7 (mo) | Records at t = 0 set to 0.001 mo |
|---|---|---|---|---|---|---|---|---|---|---|---|---|
| ASSURE clear cell, sorafenib | DFS | Alternative | 540/540 | 65.9/63.0 | 55.2/54.3 (36) | 36 | — (rule applied) | 72 | 12 | 15 | — | 3 |
| PROTECT, ITT 800 mg | DFS | Sensitivity | 198/205 | 65.2/58.0 | 57.6/48.3 (36) | 36 | — (rule applied) | 60 | 12 | 15 | — | 2 |
| PROTECT, ITT all randomised | DFS | Sensitivity | 769/769 | 61.8/60.7 | 51.5/49.5 (36) | 36 | — (rule applied) | 54 | 12 | 15 | — | 10 |
| ATLAS highest risk, IRC | DFS | Sensitivity | 209/200 | 45.9/37.0 | 19.1/11.0 (36) | 36 | — (rule applied) | 36 | 12 | 36 | — | 0 |
| ATLAS highest risk, investigator | DFS | Sensitivity | 209/200 | 43.1/35.0 | 16.3/10.0 (36) | 36 | — (rule applied) | 36 | 12 | 36 | — | 0 |
| ATLAS ITT, investigator | DFS | Sensitivity | 363/361 | 48.2/44.0 | 20.1/18.6 (36) | 36 | — (rule applied) | 36 | 12 | 36 | — | 2 |
| SORCE Leibovich high risk, sorafenib 1 yr | DFS | Sensitivity | 300/206 | 66.3/59.2 | 53.3/53.9 (48) | 36 | — (rule applied) | 96 | 12 | 15 | — | 5 |
| SORCE clear cell, sorafenib 1 yr | DFS | Sensitivity | 534/361 | 77.7/73.1 | 66.1/65.4 (48) | 36 | — (rule applied) | 96 | 12 | 15 | — | 3 |
| SORCE Leibovich high risk, sorafenib 3 yr | DFS | Alternative | 295/206 | 62.7/59.2 | 48.5/53.9 (48) | 36 | — (rule applied) | 96 | 36 | 12 | — | 2 |
| SORCE clear cell, sorafenib 3 yr | DFS | Alternative | 550/361 | 75.8/73.1 | 62.2/65.4 (48) | 36 | — (rule applied) | 96 | 36 | 12 | — | 1 |
| IMmotion010 | OS | Main | 390/388 | 91.0/85.3 | 83.6/78.4 (36) | 36 | — (rule applied) | 54 | 12 | 15 | — | 0 |
| S-TRAC (updated analysis) | OS | Main | 309/306 | 83.5/87.9 | 76.4/81.7 (36) | 36 | — (rule applied) | 84 | 12 | 15 | — | 0 |
| PROTECT, ITT 600 mg (interim) | OS | Main | 571/564 | 84.8/85.8 | 75.0/75.7 (36) | 36 | — (rule applied) | 54 | 12 | 15 | — | 0 |
| PROTECT, ITT all (final; Eur Urol 2021) [did not pass the authors' visual check; shown for reference only, not used in any pooled analysis] | OS | Sensitivity | 769/769 | 83.0/85.2 | 78.7/79.7 (40) | 36 | — (rule applied) | 90 | 12 | 15 | — | 0 |
| SORCE, sorafenib 3 yr | OS | Main | 638/430 | 91.2/91.6 | 80.6/81.6 (48) | 36 | — (rule applied) | 96 | 36 | 12 | — | 0 |
| SORCE, sorafenib 1 yr | OS | Main | 641/430 | 92.7/91.6 | 80.7/81.6 (48) | 36 | — (rule applied) | 96 | 12 | 15 | — | 0 |
| CheckMate 914 part A | OS | Main | 405/411 | 85.9/87.8 | 45.4/44.8 (36) | 36 | — (rule applied) | 45 | 6 | 9 | — | 0 |

Percentages are numbers at risk divided by numbers randomised, at the number-at-risk time point at or after the stated time (J1); a lower bound when the time falls between table time points. Primary τ = 36 months if both arms had ≥10% at risk at 36 months, otherwise 24 months; all program determinations agreed with those in the registered plan. Maximum feasible τ = the latest 3-month step at which both arms had ≥10% at risk (used for the ΔRMST(τ) curves, the post-treatment increment and the HR(t) plots). LITESPARK-022 OS: no RMST at a fixed τ of 30 months was reported (SAP 8.5). For the SORCE 3-year vs 1-year comparison, the 3-year arm is the experimental arm and the 1-year arm the reference (J15).

### Supplementary Table S4. Execution decisions J1–J25 (condensed)

Source: execution-decision log (version 2). Principle: where the plan was not explicit, the most conservative option closest to the wording of the plan was chosen, and no analysis absent from the plan was added (exceptions are D5–D7 in Supplementary Table S1).

| No. | Topic (SAP section) | Decision |
|---|---|---|
| J1 | τ between number-at-risk time points (6.1, 7.2) | The number at risk at τ is taken from the nearest time point at or after τ (a lower bound); τ is infeasible if this is below 10%. Example: for LITESPARK-022, τ = 33 months lies between the 30-month (≥15%) and 36-month (<5%) time points and was not included. |
| J2 | Primary τ; τ for OS (6.1, 8.5) | Determined by the program with the 6.1 rule and checked against the plan's table (all agreed). KEYNOTE-564 was re-determined with the Ann Oncol 2026 table (36 months) and CheckMate 914 part B with the vector Fig 2 (24 months). For OS, the same rule was applied to each OS curve (KEYNOTE-564 and ASSURE 36 months; LITESPARK-022 24 months). Comparisons with a primary τ of 36 months were also reported at 24 months. |
| J3 | Records at time 0 (not specified) | Moved to 0.001 months (about 0.03 days) once, at data loading, for all analyses. Affected: PROTECT 2, SORCE 1 year vs placebo 4, PROSPER standard 73, PROSPER censoring-informed 78, SORCE 3 years vs 1 year 2 records. |
| J4 | Direction of effects | Experimental arm = 1; ΔRMST = experimental − control; HRs and ratios experimental vs control. |
| J5 | CIs of the ΔRMST(τ) curve (7.2) | Pointwise asymptotic CIs from `rmst2` at each τ; no simultaneous bands. The `rmst2` limit (τ ≤ shorter maximum follow-up of the two arms) was never binding. |
| J6 | Post-treatment increment (7.2) | Computed from the main landmark only; years after the landmark = (maximum feasible τ − landmark)/12; for SORCE 3 years vs placebo, from 36 months. Bootstrap CIs added post hoc (D5). |
| J7 | CIs of KM differences (8.2) | Greenwood standard errors, arms independent, normal approximation; feasible time points by the 10% rule. |
| J8 | Piecewise Cox model (8.1) | `trt + trt:late` without a period main effect; Efron ties; model-based standard errors; Wald CIs; events per arm and period reported. |
| J9 | PH test (8.3) | `cox.zph` with `transform = "km"`; with one covariate the global test equals the treatment test. The whole-period Cox HR is listed for reference only. |
| J10 | AIC selection (8.4) | Smallest AIC of 15 models; tie rule prespecified (none occurred); AIC difference to the second-best model recorded. The baseline df of 10 selected DFS models was at the upper limit of 5; the range was not extended. Fixed df 3/tvc 2 shown alongside. |
| J11 | Non-converged models (not specified) | Excluded if fitting failed, the convergence code was non-zero, a standard error was not finite or refitting failed, or the log-likelihood was more than 0.5 below the nested df 1/tvc 1 model. Excluded: PROTECT df 4/tvc 1; SORCE 1 year df 4/tvc 1; PROSPER standard 8 models; PROSPER censoring-informed 10 models. For PROSPER, the fixed df 3/tvc 2 model could not be estimated. The excluded PROTECT and SORCE models were not the AIC-best. |
| J12 | Range of HR(t) (8.4) | From 1 month to the maximum feasible τ, every 0.25 months. |
| J13 | Pooling (8.6) | Effect size = interaction coefficient; model-based standard error; `metagen` with REML and Hartung-Knapp; no ad hoc variance correction; random effects only; $\tau^2$ and $I^2$ reported. |
| J14 | Smoothed hazards (8.7) | `muhaz` defaults (local bandwidth, boundary correction at both ends, Epanechnikov kernel), 0 to maximum feasible τ, DFS only. |
| J15 | SORCE 3 years vs 1 year (9A) | 3-year arm (Fig 2A) experimental, 1-year arm (Fig 2B) reference; landmarks 36 months (main) and 12 months (sensitivity); primary τ 36 months, also 24 months; RMST difference and piecewise HRs only; only the two sorafenib arms used. |
| J16 | Sensitivity landmarks and pooling (9.1, 8.6) | Pooling re-run with the sensitivity landmarks; retained as a consequence of SAP 9 item 1 (D6). |
| J17 | Replacing shared-control comparisons (8.6, 9.8) | One at a time (ASSURE sorafenib; SORCE 3 years; CheckMate 914 part B nivolumab plus ipilimumab), plus both TKI replacements together; SORCE 3 years uses its own main landmark of 36 months. |
| J18 | Adding PROSPER (9.3) | All within-trial analyses run (landmarks 12 and 10 months) and PROSPER |

| No. | Topic (SAP section) | Decision |
|---|---|---|
| | | added to the ICI pooling once for each reconstruction, labelled as failing QC (standard, D4-A) or as a method outside the plan (censoring-informed, D4-C). The Day-1-censored supplementary curve had no IPD and was not analysed. |
| J19 | Excluding raster-derived curves (post hoc) | The main analyses contain no raster-derived curve; the only raster curve (CheckMate 914 part B Fig S2) enters only one sensitivity pooling, so this analysis equals the main analysis. |
| J20 | Curves failing QC (9.9) | The main analyses contain no curve that failed QC. “Inclusion” versions: LITESPARK-022 Fig 1A standard (D1), LITESPARK-022 OS standard (D2) and PROSPER standard (D4; also added to the ICI pooling). |
| J21 | Scope of OS (8.5) | RMST difference (primary τ and 24 months) and flexible parametric HR(t) only; no piecewise HRs, PH tests or ΔRMST(τ) curves. LITESPARK-022 OS descriptive (D2), both reconstructions shown. |
| J22 | KEYNOTE-564 IPD version | IPD with points added at censoring marks used (confirmed 13 Oct 2026). |
| J23 | Unused censoring-informed IPD | IMmotion010 Fig 2A, SORCE Fig 2A and S-TRAC Fig 2 censoring-informed IPD were used only to validate the method. |
| J24 | Random seed | `set.seed(20261013)` recorded although the main analyses have no random step; the bootstrap scripts (D5, D7) set the same seed. |
| K1 | Bootstrap seed for curves added under D8 | `set.seed(20261013)` before the resampling of each curve (not once per script), so that adding a curve cannot change other intervals; B = 2000, within-arm resampling and percentile CIs as in D5. |
| K2 | τ for curves added under D8 (6.1) | Determined by the program with the 6.1 rule; only KEYNOTE-564 NEJM 2024 appears in the plan’s table (36 months), and the determination agreed. |
| K3 | Landmarks for curves added under D8 (6.2) | The trial’s main and sensitivity landmarks (subsets and assessment methods do not change planned treatment duration); SORCE 3-year subsets 36 and 12 months. |
| K4 | OS piecewise HRs (8.5) | Added for description only (not pooled, no inference), using the landmarks of the trial’s DFS analysis; also computed for the OS curves analysed in version 1. Supersedes the scope statement of J21 for OS. |
| K5 | Modified Hartung-Knapp (8.6; post hoc) | `adhoc.hakn.ci = "se"` in meta 8.5.0, equivalent to q* = max(1, q) with t(k − 1); verified by direct calculation (S-M5). |
| K6 | Substitution sensitivity analyses (9, items 2–7) | One curve at a time, main landmarks; PROSPER Day-1-censored curve added to the ICI pooling as in J18. Supersedes the J18 statement that this curve had no IPD. |
| K7 | Final PROTECT OS (Eur Urol 2021) | Analysed, but labelled “did not pass the authors’ visual check; shown for reference only, not used in any pooled analysis” wherever reported (no manual re-digitisation; decision of 14 Oct 2026). |
| K8 | ASSURE clear cell; PROTECT Fig A1B and A1C | Accepted after adjudication (D8-c, D8-d); labelled wherever reported. |
| J25 | Reporting | ΔRMST, HRs and ratios rounded to 2 decimals, RMST ratios to 3, KM differences to 1; full precision in the CSV files. RMST P values are those from `rmst2` and were not required by the plan. |

#### Supplementary Table S5. Reconstruction accuracy for each curve

Source: reconstruction quality-control reports (section “quality checks” and “overall judgement” of each report).

| Curve | Endpoint | Use | Reconstruction | Reported HR (CI) | Reconstructed HR (95% CI) | Difference in HR | Events reported → reconstructed, exp/ctrl (% difference) | Median, reported vs reconstructed (mo), exp; ctrl | Landmark survival, reconstructed minus reported (pp), exp/ctrl | Maximum deviation of reconstructed KM from digitised curve, exp/ctrl (pp, at time) | Numerical criteria | Visual check (second investigator) |
|---|---|---|---|---|---|---|---|---|---|---|---|---|
| LITESPARK-022 Fig 1B (BICR) | DFS | Main | Standard | 0.760 (95% CI 0.640–0.920) | 0.756 (95% CI 0.628–0.911) | −0.004; within reported CI | 205/252 → 199/250 (−2.9%/−0.8%) | NR in both (reconstruction also NR) | 12 mo: −0.2/−0.1; 24 mo: 0.0/−0.3; 30 mo: +0.2/−0.1 | 1.3 (36.6 mo) / 0.8 (33.1 mo) | Pass | Pass (second investigator; R01) |
| KEYNOTE-564 | DFS | Main | Standard ¶ | 0.710 (95% CI | 0.724 (95% CI | +0.014; within | 188/241 → | NR vs NR; | 24 mo: | 0.4 (75.6 mo) / | Pass | Pass (second |

| Curve | Endpoint | Use | Reconstruction | Reported HR (CI) | Reconstructed HR (95% CI) | Difference in HR | Events reported → reconstructed, exp/ctrl (% difference) | Median, reported vs reconstructed (mo), exp; ctrl | Landmark survival, reconstructed minus reported (pp), exp/ctrl | Maximum deviation of reconstructed KM from digitised curve, exp/ctrl (pp, at time) | Numerical criteria | Visual check (second investigator) |
|---|---|---|---|---|---|---|---|---|---|---|---|---|
| Fig 1A (Ann Oncol 2026) | | | | 0.590–0.860) | 0.598–0.877) | reported CI | 187/239 (−0.5%/−0.8%) | 68.3 vs 68.3 | +0.1/0.0; 36 mo: 0.0/−0.1; 48 mo: +0.1/0.0; 60 mo: −0.1/−0.1; 72 mo: −0.1/−0.1 | 1.6 (80.9 mo) | | investigator; R02) |
| IMmotion010 Fig 2A | DFS | Main | Standard | 0.930 (95% CI 0.750–1.150) | 0.940 (95% CI 0.758–1.167) | +0.010; within reported CI | 164/168 → 164/166 (0.0%/−1.2%) | 57.2 vs 57.2; 49.5 vs 49.5 | 12 mo: 0.0/−0.1; 24 mo: 0.0/−0.1; 36 mo: 0.0/+0.1 | 0.4 (48.4 mo) / 1.1 (52.9 mo) | Pass | Pass (second investigator; R03) |
| CheckMate 914 A Fig 2 | DFS | Main | Standard | 0.920 (95% CI 0.710–1.190) | 0.938 (95% CI 0.722–1.218) | +0.018; within reported CI | 110/118 → 109/117 (−0.9%/−0.8%) | NR vs NR; 50.7 vs 50.6 | 24 mo: −0.2/−0.7 | 0.5 (40.2 mo) / 2.2 (48.1 mo) | Pass | Pass (second investigator; R04) |
| CheckMate 914 B Fig 2 (nivolumab) | DFS | Main | Standard | 0.870 (95% CI 0.620–1.210) | 0.834 (95% CI 0.596–1.165) | −0.036; within reported CI | 97/54 → 94/54 (−3.1%/0.0%) | NR in both (reconstruction also NR) | 18 mo: −0.3/−0.2 | 0.9 (29.7 mo) / 0.4 (24.2 mo) | Pass | Pass (second investigator; R05) |
| CheckMate 914 B Fig S2 (nivolumab + ipilimumab) | DFS | Alternative (raster) | Standard | Not reported | 1.061 (95% CI 0.731–1.540) | — | 58/54 → 57/54 (−1.7%/0.0%) | NR in both (reconstruction also NR) | Not reported | 0.3 (22.0 mo) / 0.3 (35.6 mo) | Pass | Pass (second investigator; R06) |
| S-TRAC Fig 2 | DFS | Main | Standard | 0.760 (95% CI 0.590–0.980) | 0.765 (95% CI 0.598–0.979) | +0.005; within reported CI | 113/144 → 113/143 (0.0%/−0.7%) | 81.6 vs 81.9; 67.2 vs 66.8 | 36 mo: +0.1/−0.2; 60 mo: −0.1/−0.1 | 3.5 (84.8 mo) / 1.2 (80.8 mo) | Pass | Pass (second investigator; R07) |
| ASSURE Fig 2, sunitinib vs placebo | DFS | Main | Standard | 1.020 (97.5% CI 0.850–1.230) | 1.011 (97.5% CI 0.838–1.220) | −0.009; within reported CI | 284/287 → 284/287 (0.0%/0.0%) | 70.0 vs 70.0; 79.6 vs 79.5 | 60 mo: −0.2/0.0 | 1.0 (84.6 mo) / 3.0 (96.5 mo) | Pass | Pass (second investigator; R08) |
| ASSURE Fig 2, sorafenib vs placebo | DFS | Alternative | Standard | 0.970 (97.5% CI 0.800–1.170) | 0.975 (97.5% CI 0.808–1.176) | +0.005; within reported CI | 284/287 → 285/287 (+0.4%/0.0%) | 73.4 vs 73.4; 79.6 vs 79.5 | 60 mo: −0.3/0.0 | 0.3 (79.8 mo) / 3.0 (96.5 mo) | Pass | Pass (second investigator; R09) |
| PROTECT Fig A1A | DFS | Main | Standard | 0.936 (95% CI 0.769–1.140) | 0.930 (95% CI 0.762–1.135) | −0.006; within reported CI | 194/202 → 190/198 (−2.1%/−2.0%) | NR in both (reconstruction also NR) | 12 mo: +2.8/*+3.1*; 24 mo: +1.6/−0.1; 36 mo: −1.1/+0.1; 48 mo: 0.0/0.0 | 0.4 (49.3 mo) / 0.4 (53.9 mo) | Pass under the internal-inconsistency rule (D3); 12-mo landmark outside 2 pp | Pass (second investigator; R10) |
| ATLAS Fig 1 | DFS | Main | Standard ¶ | 0.870 (95% CI 0.660–1.147) | 0.863 (95% CI 0.656–1.134) | −0.007; within reported CI | 96/107 → 98/108 (+2.1%/+0.9%) | Not reported (reconstruction NR in both) | Not reported | 0.6 (42.4 mo) / 0.7 (49.1 mo) | Pass | Pass (second investigator; R11) |
| SORCE Fig 2B (1 yr) | DFS | Main | Standard | 0.940 (95% CI 0.770–1.140) | 0.935 (95% CI 0.767–1.139) | −0.005; within reported CI | 242/167 → 242/166 (0.0%/−0.6%) | NR in both (reconstruction also NR) | 120 mo: −0.5/+1.0 | 0.4 (115.8 mo) / 1.2 (110.8 mo) | Pass | Pass (second investigator; R13) |
| SORCE Fig 2A (3 yr) | DFS | Alternative | Standard | 1.010 (95% CI 0.820–1.230) | 0.984 (95% CI 0.808–1.199) | −0.026; within reported CI | 245/167 → 244/166 (−0.4%/−0.6%) | NR in both (reconstruction also NR) | 120 mo: +1.2/+1.0 | 1.0 (113.6 mo) / 1.2 (110.8 mo) | Pass | Pass (second investigator; R12) |
| LITESPARK-022 Fig 1A (investigator) | DFS | Sensitivity | Standard | 0.720 (95% CI 0.590–0.870) | 0.700 (95% CI 0.576–0.852) | −0.020; within reported CI | 186/246 → 173/238 (−7.0%/−3.3%) * | NR in both (reconstruction also NR) | 12 mo: +0.2/+0.2; 24 mo: +0.2/+0.3; 30 mo: +0.2/+0.4 | 0.8 (33.1 mo) / 0.7 (32.9 mo) | Fail (events belzutifan) | Pass (second investigator; R14) |
| LITESPARK-022 Fig 1A (investigator) | DFS | Sensitivity | Censoring-informed (post hoc) | 0.720 (95% CI 0.590–0.870) | 0.719 (95% CI 0.594–0.871) | −0.001; within reported CI | 186/246 → 184/243 (−1.1%/−1.2%) | NR in both (reconstruction also NR) | 12 mo: +0.1/+0.2; 24 mo: +0.2/+0.2; 30 mo: | 0.8 (36.9 mo) / 0.2 (29.3 mo) | Pass | Pass (second investigator; R15) |

| Curve | Endpoint | Use | Reconstruction | Reported HR (CI) | Reconstructed HR (95% CI) | Difference in HR | Events reported → reconstructed, exp/ctrl (% difference) | Median, reported vs reconstructed (mo), exp; ctrl | Landmark survival, reconstructed minus reported (pp), exp/ctrl | Maximum deviation of reconstructed KM from digitised curve, exp/ctrl (pp, at time) | Numerical criteria | Visual check (second investigator) |
|---|---|---|---|---|---|---|---|---|---|---|---|---|
| | | | | | | | | | +0.3/+0.3 | | | |
| PROSPER Fig 2 | RFS | Sensitivity | Standard | 0.940 (95% CI 0.740–1.210) | 0.955 (95% CI 0.742–1.229) | +0.015; within reported CI | 125/133 → 118/124 (−5.6%/−6.8%) * | Not reported (reconstruction NR in both) | Not reported | 0.5 (39.2 mo) / 0.9 (42.2 mo) | Fail (events nivolumab, events surgery) | Pass (second investigator; R21) |
| PROSPER Fig 2 | RFS | Sensitivity | Censoring-informed (post hoc) | 0.940 (95% CI 0.740–1.210) | 0.938 (95% CI 0.734–1.198) | −0.002; within reported CI | 125/133 → 124/133 (−0.8%/0.0%) | Not reported (reconstruction NR in both) | Not reported | 2.1 (39.2 mo) / 5.0 (45.8 mo) | Pass | Pass (second investigator; R22) |
| LITESPARK-022 Fig 2 | OS | Descriptive | Standard | 0.780 (95% CI 0.510–1.190) | 0.765 (95% CI 0.492–1.187) | −0.015; within reported CI | 38/49 → 35/46 (−7.9%/−6.1%) * | Not reported (reconstruction NR in both) | 12 mo: 0.0/0.0; 24 mo: 0.0/0.0; 30 mo: +0.2/0.0 | 0.4 (32.8 mo) / 0.4 (32.2 mo) | Fail (events belzutifan, events placebo) | Pass (second investigator; R16) |
| LITESPARK-022 Fig 2 | OS | Descriptive | Censoring-informed (post hoc) | 0.780 (95% CI 0.510–1.190) | 0.776 (95% CI 0.508–1.185) | −0.004; within reported CI | 38/49 → 38/49 (0.0%/0.0%) | Not reported (reconstruction NR in both) | 12 mo: 0.0/0.0; 24 mo: 0.0/−0.1; 30 mo: 0.0/0.0 | 0.1 (27.7 mo) / 0.1 (32.2 mo) | Pass | Pass (second investigator; R17) |
| KEYNOTE-564 Fig 1B (Ann Oncol 2026) | OS | Secondary | Standard ¶ | 0.660 (95% CI 0.480–0.900) | 0.662 (95% CI 0.486–0.901) | +0.002; within reported CI | 68/99 → 68/99 (0.0%/0.0%) | NR in both (reconstruction also NR) | 24 mo: 0.0/0.0; 36 mo: 0.0/0.0; 48 mo: 0.0/0.0; 60 mo: 0.0/0.0; 72 mo: 0.0/0.0 | 0.1 (78.3 mo) / 0.1 (80.7 mo) | Pass | Pass (second investigator; R18) |
| ASSURE Fig 3, sunitinib vs placebo | OS | Secondary | Standard | 1.170 (97.5% CI 0.900–1.520) | 1.165 (97.5% CI 0.896–1.516) | −0.005; within reported CI | 156/141 → 153/138 (−1.9%/−2.1%) | NR vs 101.6; NR vs NR | 60 mo: +0.1/−0.2 | 3.9 (101.7 mo) / 1.3 (92.3 mo) | Pass | Pass (second investigator; R19) |
| ASSURE Fig 3, sorafenib vs placebo | OS | Secondary | Standard | 0.980 (97.5% CI 0.750–1.280) | 1.002 (97.5% CI 0.764–1.314) | +0.022; within reported CI | 138/141 → 135/138 (−2.2%/−2.1%) | NR in both (reconstruction also NR) | 60 mo: +0.2/−0.2 | 1.1 (90.7 mo) / 1.3 (92.3 mo) | Pass | Pass (second investigator; R20) |
| S-TRAC Fig 2 | DFS | Method validation only (not analysed) | Censoring-informed | 0.760 (95% CI 0.590–0.980) | 0.760 (95% CI 0.594–0.971) | 0.000; within reported CI | 113/144 → 114/145 (+0.9%/+0.7%) | 81.6 vs 84.7; 67.2 vs 64.2 | 36 mo: 0.0/−0.1; 60 mo: −0.1/−0.2 | 0.5 (84.8 mo) / 0.6 (77.2 mo) | Pass | Not reviewed (method validation only; not in the review packages; not used in any analysis) |
| IMmotion010 Fig 2A | DFS | Method validation only (not analysed) | Censoring-informed | 0.930 (95% CI 0.750–1.150) | 0.927 (95% CI 0.748–1.150) | −0.003; within reported CI | 164/168 → 164/168 (0.0%/0.0%) | 57.2 vs 57.2; 49.5 vs 49.5 | 12 mo: 0.0/−0.1; 24 mo: 0.0/−0.1; 36 mo: 0.0/−0.1 | 0.1 (2.8 mo) / 0.1 (47.9 mo) | Pass | Not reviewed (method validation only; not in the review packages; not used in any analysis) |
| SORCE Fig 2A (3 yr) | DFS | Method validation only (not analysed) | Censoring-informed | 1.010 (95% CI 0.820–1.230) | 0.984 (95% CI 0.809–1.197) | −0.026; within reported CI | 245/167 → 248/168 (+1.2%/+0.6%) | NR in both (reconstruction also NR) | 120 mo: −0.3/−0.5 | 0.5 (113.6 mo) / 0.4 (108.8 mo) | Pass | Not reviewed (method validation only; not in the review packages; not used in any analysis) |
| KEYNOTE-564 Fig 1A (NEJM 2021) | DFS | Sensitivity (SAP 9.2) | Standard ¶ | 0.680 (95% CI 0.530–0.870) | 0.684 (95% CI 0.534–0.876) | +0.004; within reported CI | 109/151 → 108/150 (−0.9%/−0.7%) | NR in both (reconstruction also NR) | 12 mo: 0.0/0.0; 24 mo: −0.1/+0.1 | 2.4 (36.0 mo) / 0.1 (29.3 mo) | Pass | Pass (second investigator; R24) |
| KEYNOTE-564 | DFS | Sensitivity | Standard ¶ | 0.630 (95% CI | 0.632 (95% CI | +0.002; within | 114/169 → | NR in both | 30 mo: | 0.4 (36.4 mo) / | Pass | Pass (second |

| Curve | Endpoint | Use | Reconstruction | Reported HR (CI) | Reconstructed HR (95% CI) | Difference in HR | Events reported → reconstructed, exp/ctrl (% difference) | Median, reported vs reconstructed (mo), exp; ctrl | Landmark survival, reconstructed minus reported (pp), exp/ctrl | Maximum deviation of reconstructed KM from digitised curve, exp/ctrl (pp, at time) | Numerical criteria | Visual check (second investigator) |
|---|---|---|---|---|---|---|---|---|---|---|---|---|
| Fig 2A (Lancet Oncol 2022) | | (SAP 9.2) | | 0.500–0.800) | 0.499–0.800) | reported CI | 115/171 (+0.9%/+1.2%) | (reconstruction also NR) | −0.1/−0.2 | 0.3 (33.2 mo) | | investigator; R25) |
| KEYNOTE-564 Fig 2A (NEJM 2024) | DFS | Sensitivity (SAP 9.2) | Standard | 0.720 (95% CI 0.590–0.870) | 0.719 (95% CI 0.589–0.877) | −0.001; within reported CI | 174/224 → 172/223 (−1.1%/−0.4%) | Not reported (reconstruction NR in both) | 12 mo: +0.1/0.0; 24 mo: 0.0/0.0; 36 mo: 0.0/0.0; 48 mo: +0.2/−0.1 | 1.4 (64.5 mo) / 0.6 (61.1 mo) | Pass | Pass (second investigator; R26) |
| PROSPER Supplemental Fig 3 (Day-1 events censored) | RFS | Sensitivity (SAP 9.3) | Standard | 0.80 (CI not reported) | 0.788 (95% CI 0.594–1.045) | −0.012 (absolute difference only; CI not reported) | 87/106 → 87/108 (0.0%/+1.9%) | Not reported (reconstruction NR in both) | Not reported | 0.4 (39.2 mo) / 0.4 (41.9 mo) | Pass | Pass (second investigator; R27) |
| ASSURE Suppl Fig 1 (clear cell), sunitinib vs placebo | DFS | Sensitivity (SAP 9.4) | Standard | 1.020 (97.5% CI 0.850–1.220) | 1.011 (97.5% CI 0.824–1.241) | −0.009; within reported CI | 220/218 → 240/240 (+9.1%/+10.1 %)* | 66.9 vs 66.9; 74.4 vs 79.6 | 60 mo: 0.0/−0.1 | 4.6 (96.9 mo) / 2.5 (93.8 mo) | Accepted after adjudication (source inconsistency; see Table S1). Automated check: Fail (events sunitinib, events placebo, median DFS placebo) | Pass (second investigator; R28) |
| ASSURE Suppl Fig 1 (clear cell), sorafenib vs placebo | DFS | Sensitivity (SAP 9.4) | Standard | 0.990 (97.5% CI 0.830–1.190) | 0.977 (97.5% CI 0.797–1.198) | −0.013; within reported CI | 227/218 → 244/240 (+7.5%/+10.1 %)* | 66.9 vs 69.7; 74.4 vs 79.6 | 60 mo: −0.1/−0.1 | 0.4 (79.8 mo) / 2.5 (93.8 mo) | Accepted after adjudication (source inconsistency; see Table S1). Automated check: Fail (events sorafenib, events placebo, median DFS sorafenib, median DFS placebo) | Pass (second investigator; R29) |
| PROTECT Fig A1B (ITT800mg) | DFS | Sensitivity (SAP 9.5) | Standard | 0.663 (95% CI 0.491–0.895) | 0.647 (95% CI 0.477–0.877) | −0.016; within reported CI | 70/101 → 70/102 (0.0%/+1.0%) | NR vs NR; 54.0 vs 53.6 | 12 mo: +1.7/+1.8; 24 mo: −0.1/+1.2; 36 mo: −0.9/−0.8; 48 mo: −0.2/−0.9; 60 mo: −0.4/−3.3* | 0.3 (54.4 mo) / 0.4 (59.9 mo) | Accepted after adjudication (source inconsistency; see Table S1). Automated check: Fail (landmark 60 mo placebo) | Pass (second investigator; R30) |
| PROTECT Fig A1C (ITTAll) | DFS | Sensitivity (SAP 9.5) | Standard | 0.842 (95% CI 0.714–0.993) | 0.834 (95% CI 0.708–0.982) | −0.008; within reported CI | 264/303 → 270/305 (+2.3%/+0.7%) | NR in both (reconstruction also NR) | 12 mo: −1.0/+2.2*; 24 mo: −0.3/+1.0; 36 mo: −1.0/−1.3; 48 mo: −1.2/−1.0;* | 0.6 (59.6 mo) / 0.6 (59.7 mo) | Accepted after adjudication (source inconsistency; see Table S1). Automated | Pass (second investigator; R31) |

| Curve | Endpoint | Use | Reconstruction | Reported HR (CI) | Reconstructed HR (95% CI) | Difference in HR | Events reported → reconstructed, exp/ctrl (% difference) | Median, reported vs reconstructed (mo), exp; ctrl | Landmark survival, reconstructed minus reported (pp), exp/ctrl | Maximum deviation of reconstructed KM from digitised curve, exp/ctrl (pp, at time) | Numerical criteria | Visual check (second investigator) |
|---|---|---|---|---|---|---|---|---|---|---|---|---|
| | | | | | | | | | *60 mo: +0.1/−3.2* | | check: Fail (landmark 12 mo placebo, landmark 60 mo placebo) | |
| ATLAS Fig 3A (highest risk, IRC) | DFS | Sensitivity (SAP 9.6) | Standard ¶ | 0.735 (95% CI 0.525–1.028) | 0.728 (95% CI 0.521–1.018) | −0.007; within reported CI | 63/74 → 64/74 (+1.6%/0.0%) | Not reported (reconstruction NR in both) | Not reported | 1.4 (49.2 mo) / 1.0 (36.0 mo) | Pass | Pass (second investigator; R32) |
| ATLAS Fig 3B (highest risk, investigator) | DFS | Sensitivity (SAP 9.6) | Standard ¶ | 0.641 (95% CI 0.468–0.879) | 0.640 (95% CI 0.468–0.875) | −0.001; within reported CI | 69/89 → 70/89 (+1.4%/0.0%) | not reported vs 52.2; not reported vs 33.2 | Not reported | 2.5 (52.2 mo) / 0.4 (41.9 mo) | Pass | Pass (second investigator; R33) |
| ATLAS Fig S2 (ITT, investigator) | DFS | Sensitivity (SAP 9.6) | Standard | 0.776 (95% CI 0.599–1.005) | 0.764 (95% CI 0.588–0.993) | −0.012; within reported CI | 106/126 → 103/124 (−2.8%/−1.6%) | Not reported (reconstruction NR in both) | Not reported | 1.1 (51.9 mo) / 0.9 (46.5 mo) | Pass | Pass (second investigator; R34) |
| SORCE Fig A1 (Leibovich high risk), 1 yr | DFS | Sensitivity (SAP 9.7) | Standard | 0.920 (95% CI 0.720–1.170) | 0.914 (95% CI 0.716–1.165) | −0.006; within reported CI | not reported/not reported → 160/110 (—/—) | not reported vs 75.3; not reported vs 72.7 | Not reported | 0.9 (121.3 mo) / 2.2 (108.8 mo) | Pass | Pass (second investigator; R36) |
| SORCE Fig A1 (Leibovich high risk), 3 yr | DFS | Sensitivity (SAP 9.7) | Standard | 1.020 (95% CI 0.800–1.300) | 1.005 (95% CI 0.789–1.281) | −0.015; within reported CI | not reported/not reported → 162/110 (—/—) | not reported vs 54.0; not reported vs 72.4 | Not reported | 1.8 (113.6 mo) / 1.6 (108.8 mo) | Pass | Pass (second investigator; R35) |
| SORCE Fig A2 (clear cell), 1 yr | DFS | Sensitivity (SAP 9.7) | Standard | 0.940 (95% CI 0.750–1.160) | 0.927 (95% CI 0.747–1.150) | −0.013; within reported CI | not reported/not reported → 201/140 (—/—) | Not reported (reconstruction NR in both) | Not reported | 0.5 (115.7 mo) / 1.6 (110.8 mo) | Pass | Pass (second investigator; R38) |
| SORCE Fig A2 (clear cell), 3 yr | DFS | Sensitivity (SAP 9.7) | Standard | 0.990 (95% CI 0.800–1.230) | 0.986 (95% CI 0.796–1.221) | −0.004; within reported CI | not reported/not reported → 210/140 (—/—) | Not reported (reconstruction NR in both) | Not reported | 1.4 (113.6 mo) / 1.5 (110.8 mo) | Pass | Pass (second investigator; R37) |
| IMmotion010 Fig 3 | OS | Secondary (SAP 8.5) | Standard | 0.970 (95% CI 0.670–1.420) | 0.971 (95% CI 0.660–1.430) | +0.001; within reported CI | 54/53 → 52/51 (−3.7%/−3.8%) | NR in both (reconstruction also NR) | 36 mo: +0.2/+0.2 | 0.6 (52.0 mo) / 0.6 (52.3 mo) | Pass | Pass (second investigator; R39) |
| S-TRAC Eur Urol 2018 Fig 3 | OS | Secondary (SAP 8.5) | Standard | 0.920 (95% CI 0.660–1.280) | 0.951 (95% CI 0.680–1.331) | +0.031; within reported CI | 67/74 → 65/73 (−3.0%/−1.4%) | NR in both (reconstruction also NR) | Not reported | 1.2 (85.2 mo) / 0.7 (95.6 mo) | Pass | Pass (second investigator; R40) |
| PROTECT JCO 2017 Fig 4A (ITT600mg) | OS | Secondary (SAP 8.5) | Standard | 0.791 (95% CI 0.573–1.092) | 0.773 (95% CI 0.552–1.082) | −0.018; within reported CI | not reported/not reported → 60/78 (—/—) | NR in both (reconstruction also NR) | Not reported | 0.7 (52.8 mo) / 0.3 (50.1 mo) | Pass | Pass (second investigator; R41) |
| PROTECT Eur Urol 2021 Fig 1A (ITTAll, final) | OS | Sensitivity (raster) | Standard | 1.000 (95% CI 0.800–1.260) | 1.040 (95% CI 0.826–1.310) | +0.040; within reported CI | 145/150 → 146/143 (+0.7%/−4.7%) | NR in both (reconstruction also NR) | Not reported | 0.4 (93.1 mo) / 0.2 (91.6 mo) | Pass (numerical criteria met; the authors judged the digitisation coarse at 60–95 mo; not re-digitised; see Table S1, D8-g) | Second investigator: no discrepancy; the authors' stricter judgement is retained: did not pass the authors' visual check; shown for reference only, not used in any pooled analysis (R42) |

| Curve | Endpoint | Use | Reconstruction | Reported HR (CI) | Reconstructed HR (95% CI) | Difference in HR | Events reported → reconstructed, exp/ctrl (% difference) | Median, reported vs reconstructed (mo), exp; ctrl | Landmark survival, reconstructed minus reported (pp), exp/ctrl | Maximum deviation of reconstructed KM from digitised curve, exp/ctrl (pp, at time) | Numerical criteria | Visual check (second investigator) |
|---|---|---|---|---|---|---|---|---|---|---|---|---|
| SORCE Fig 3A (3 yr) | OS | Secondary (SAP 8.5) | Standard | 1.060 (95% CI 0.820–1.380) | 1.042 (95% CI 0.802–1.353) | −0.018; within reported CI | 143/96 → 142/93 (−0.7%/−3.1%) | NR in both (reconstruction also NR) | 120 mo: +0.8/−1.8 | 1.2 (123.2 mo) / 1.7 (104.0 mo) | Pass | Pass (second investigator; R43) |
| SORCE Fig 3B (1 yr) | OS | Secondary (SAP 8.5) | Standard | 0.920 (95% CI 0.710–1.200) | 0.905 (95% CI 0.693–1.182) | −0.015; within reported CI | 132/96 → 131/93 (−0.8%/−3.1%) | NR in both (reconstruction also NR) | 120 mo: +0.3/−1.8 | 1.3 (127.3 mo) / 1.7 (104.0 mo) | Pass | Pass (second investigator; R44) |
| CheckMate 914 A Fig S3 | OS | Secondary (SAP 8.5; raster) | Standard | Not reported | 1.196 (95% CI 0.723–1.979) | — | 33/28 → 33/28 (0.0%/0.0%) | NR in both (reconstruction also NR) | Not reported | 0.1 (44.1 mo) / 0.2 (37.6 mo) | Pass | Pass (second investigator; R45) |
| E2810 Fig 2 | DFS | Main (D9) | Standard ¶ | 0.900 (95% CI 0.600–1.340) | 0.896 (95% CI 0.602–1.333) | −0.004; within reported CI | 49/49 → 49/49 (0.0%/0.0%) | not reported vs 17.0; not reported vs 14.2 | 36 mo: 0.0/−0.4 | 0.9 (17.6 mo) / 1.4 (69.0 mo) | Pass | Pass (second investigator; R23) |
| E2810 Fig 3 | OS | Secondary (D9) | Standard ¶ | 2.550 (95% CI 1.230–5.270) | 2.135 (95% CI 1.067–4.271) | −0.415; within reported CI | 24/12 → 24/12 (0.0%/0.0%) | NR vs 79.8; NR vs NR | 36 mo: 0.0/0.0 | 1.2 (79.8 mo) / 0.9 (70.3 mo) | OS accepted after adjudication (reported HR 2.55 vs HR 2.11 derived exactly from the curve; reported model not stated — see Table S1, D9). Automated check: Fail (HR pazopanib vs placebo); deaths match and every drop is one death, so the HR difference reflects the published (unstated, possibly stratified) model rather than reconstruction error | Pass (second investigator; R46) |

HRs in this table are shown to three decimals so that small differences are visible; elsewhere they are rounded to two decimals. Reported HRs and CIs are those of the publication (97.5% CIs for ASSURE); reconstructed HRs are from unstratified Cox models with 95% CIs (97.5% for ASSURE). * Criterion not met. ¶ Points added at censoring marks (`curve_points_at_censor`; S-M2). "Maximum deviation" compares the reconstructed KM curve with the digitised curve, not with the published estimates; large deviations occurred only in tails with few patients at risk. Event totals used as references: LITESPARK-022 Fig 1B events as printed on the figure (BICR: 205/252, which differ from the investigator counts of 186/246); SORCE arm B 242 (CONSORT diagram; 245 in the text); CheckMate 914 part A 228 in total (final publication); PROTECT recurrences plus deaths summed by the extractor from the Eur Urol 2021 supplementary table; ATLAS events printed on the figure. S-TRAC medians were reported in years and converted to months (× 12). For ASSURE OS (sunitinib vs placebo), the reconstructed median in the sunitinib arm was reached at 101.6 months, after the end of the number-at-risk table at 96 months, whereas the publication reported that median OS was not reached in any arm. For the PROSPER censoring-informed reconstruction, the event-count check is not independent of the input (S-M3). The three rows marked "Method validation only" are validation reconstructions that were not used in any analysis (J23). Rows from "KEYNOTE-564 Fig 1A (NEJM 2021)" to "CheckMate 914 A Fig S3" are curves added under deviation D8; the last two rows are E2810 (D9). For E2810 OS the HR item failed (published 2.55 vs reconstructed unstratified HR; death counts match exactly); OS accepted after adjudication (reported HR 2.55 vs HR 2.11 derived exactly from the curve; reported model not stated — see Table S1, D9). For ASSURE clear cell and PROTECT Fig A1B/A1C, the automated check failed and the curves were accepted after adjudication (Table S1, D8-c and D8-d); the automated result is shown after the decision. For the final PROTECT OS curve the numerical criteria were met, but it did not pass the authors' visual check; shown for reference only, not used in any pooled analysis (not re-digitised; Table S1, D8-g). For PROSPER Supplemental Fig 3 and CheckMate 914 part A OS, no CI (PROSPER) or no HR (CheckMate 914 part A) was reported, so only the absolute HR difference, or no HR comparison, was possible. For the SORCE subsets, event counts, landmarks and medians were not reported. Visual check: all 46 overlays in the two review packages (curve numbers R01–R46 in the last column) were checked independently by the second investigator, who found no discrepancy (review completed 14 October 2026; S-M4); for R42 the authors' stricter judgement is retained. The three method-validation reconstructions were not in the review packages.

#### Supplementary Table S6. Sensitivity analyses of the class-level pooling

Source: class-level pooling output table of the SAP-completion run (the version 1 analyses, identical to the version 1 pooling output, plus the substitution analyses and the modified Hartung-Knapp columns) and, for the LITESPARK-022 rows, the version 1 pooling output; per-comparison inputs in the pooling input tables and Supplementary Tables S10 and S16–S18.

| Analysis | Class | Landmark | k | Comparisons | Pooled post/on-treatment HR ratio, ordinary HK (95% CI) | P | Modified HK 95% CI (q* = max(1, q); post hoc) | P, modified HK | q | τ² | I² (%) | P (Q test) |
|---|---|---|---|---|---|---|---|---|---|---|---|---|
| Main analysis (SAP 8.6) | VEGFR-TKI | main | 6 | S-TRAC; ASSURE, sunitinib; PROTECT; ATLAS; SORCE, sorafenib 1 yr; E2810 (M1 NED after metastasectomy) | 1.81 (1.29–2.53) | 0.006 | 1.29–2.54 | 0.007 | 0.981 | 0.045 | 43 | 0.116 |
| ASSURE: sorafenib replaces sunitinib (SAP 9, item 8) | VEGFR-TKI | main | 6 | S-TRAC; ASSURE, sorafenib; PROTECT; ATLAS; SORCE, sorafenib 1 yr; E2810 (M1 NED after metastasectomy) | 1.84 (1.33–2.54) | 0.005 | 1.33–2.54 | 0.005 | 1.013 | 0.034 | 37 | 0.160 |
| SORCE: 3 yr replaces 1 yr, landmark 36 mo (SAP 9, item 8) | VEGFR-TKI | main | 6 | S-TRAC; ASSURE, sunitinib; PROTECT; ATLAS; SORCE, sorafenib 3 yr; E2810 (M1 NED after metastasectomy) | 1.63 (1.13–2.36) | 0.019 | 1.12–2.37 | 0.020 | 0.964 | 0.064 | 51 | 0.068 |
| Both TKI replacements (SAP 9, item 8) | VEGFR-TKI | main | 6 | S-TRAC; ASSURE, sorafenib; PROTECT; ATLAS; | 1.66 (1.15–2.38) | 0.016 | 1.15–2.39 | 0.017 | 0.971 | 0.059 | 49 | 0.080 |

| Analysis | Class | Landmark | k | Comparisons | Pooled post/on-treatment HR ratio, ordinary HK (95% CI) | P | Modified HK 95% CI (q* = max(1, q); post hoc) | P, modified HK | q | τ² | I² (%) | P (Q test) |
|---|---|---|---|---|---|---|---|---|---|---|---|---|
| | | | | SORCE, sorafenib 3 yr; E2810 (M1 NED after metastasectomy) | | | | | | | | |
| Later (sensitivity) landmarks (SAP 9, item 1; D6) | VEGFR-TKI | sens | 6 | S-TRAC; ASSURE, sunitinib; PROTECT; ATLAS; SORCE, sorafenib 1 yr; E2810 (M1 NED after metastasectomy) | 1.85 (1.44–2.37) | 0.001 | 1.36–2.52 | 0.004 | 0.654 | 0.022 | 4 | 0.388 |
| Excluding E2810 (M1 NED population; post hoc, D9) | VEGFR-TKI | main | 5 | S-TRAC; ASSURE, sunitinib; PROTECT; ATLAS; SORCE, sorafenib 1 yr | 1.78 (1.17–2.72) | 0.019 | 1.17–2.72 | 0.019 | 1.069 | 0.057 | 54 | 0.070 |
| ASSURE clear cell, sunitinib replaces ASSURE, sunitinib (SAP 9, item 4) | VEGFR-TKI | main | 6 | S-TRAC; ASSURE clear cell, sunitinib; PROTECT; ATLAS; SORCE, sorafenib 1 yr; E2810 (M1 NED after metastasectomy) | 1.90 (1.40–2.58) | 0.003 | 1.40–2.58 | 0.003 | 1.070 | 0.020 | 28 | 0.224 |
| PROTECT, ITT 800 mg replaces PROTECT (SAP 9, item 5) | VEGFR-TKI | main | 6 | S-TRAC; ASSURE, sunitinib; PROTECT, ITT 800 mg; ATLAS; SORCE, sorafenib 1 yr; E2810 (M1 NED after metastasectomy) | 1.63 (1.25–2.14) | 0.006 | 1.24–2.15 | 0.006 | 0.968 | 0.005 | 5 | 0.385 |
| PROTECT, ITT all randomised replaces PROTECT (SAP 9, item 5) | VEGFR-TKI | main | 6 | S-TRAC; ASSURE, sunitinib; PROTECT, ITT all randomised; ATLAS; SORCE, sorafenib 1 yr; E2810 (M1 NED after | 1.65 (1.31–2.09) | 0.003 | 1.31–2.09 | 0.003 | 1.033 | 0.000 | 3 | 0.396 |

| Analysis | Class | Landmark | k | Comparisons | Pooled post/on-treatment HR ratio, ordinary HK (95% CI) | P | Modified HK 95% CI (q* = max(1, q); post hoc) | P, modified HK | q | τ² | I² (%) | P (Q test) |
|---|---|---|---|---|---|---|---|---|---|---|---|---|
| | | | | metastasectomy) | | | | | | | | |
| ATLAS highest risk, IRC replaces ATLAS (SAP 9, item 6) | VEGFR-TKI | main | 6 | S-TRAC; ASSURE, sunitinib; PROTECT; ATLAS highest risk, IRC; SORCE, sorafenib 1 yr; E2810 (M1 NED after metastasectomy) | 1.85 (1.36–2.53) | 0.004 | 1.35–2.54 | 0.004 | 0.957 | 0.029 | 32 | 0.195 |
| ATLAS highest risk, investigator replaces ATLAS (SAP 9, item 6) | VEGFR-TKI | main | 6 | S-TRAC; ASSURE, sunitinib; PROTECT; ATLAS highest risk, investigator; SORCE, sorafenib 1 yr; E2810 (M1 NED after metastasectomy) | 1.96 (1.55–2.48) | <0.001 | 1.47–2.62 | 0.002 | 0.663 | 0.017 | 0 | 0.429 |
| ATLAS ITT, investigator replaces ATLAS (SAP 9, item 6) | VEGFR-TKI | main | 6 | S-TRAC; ASSURE, sunitinib; PROTECT; ATLAS ITT, investigator; SORCE, sorafenib 1 yr; E2810 (M1 NED after metastasectomy) | 1.94 (1.54–2.44) | <0.001 | 1.47–2.56 | 0.002 | 0.694 | 0.014 | 0 | 0.434 |
| SORCE Leibovich high risk, sorafenib 1 yr replaces SORCE, sorafenib 1 yr (SAP 9, item 7) | VEGFR-TKI | main | 6 | S-TRAC; ASSURE, sunitinib; PROTECT; ATLAS; SORCE Leibovich high risk, sorafenib 1 yr; E2810 (M1 NED after metastasectomy) | 1.92 (1.26–2.92) | 0.010 | 1.26–2.94 | 0.011 | 0.977 | 0.095 | 60 | 0.028 |
| SORCE clear cell, sorafenib 1 yr replaces SORCE, sorafenib 1 yr (SAP 9, item 7) | VEGFR-TKI | main | 6 | S-TRAC; ASSURE, sunitinib; PROTECT; ATLAS; SORCE clear | 1.85 (1.29–2.65) | 0.007 | 1.28–2.66 | 0.008 | 0.969 | 0.057 | 49 | 0.082 |

| Analysis | Class | Landmark | k | Comparisons | Pooled post/on-treatment HR ratio, ordinary HK (95% CI) | P | Modified HK 95% CI (q* = max(1, q); post hoc) | P, modified HK | q | τ² | I² (%) | P (Q test) |
|---|---|---|---|---|---|---|---|---|---|---|---|---|
| | | | | cell, sorafenib 1 yr; E2810 (M1 NED after metastasectomy) | | | | | | | | |
| Main analysis (SAP 8.6) | ICI | main | 4 | KEYNOTE-564; IMmotion010; CheckMate 914 part A; CheckMate 914 part B, nivolumab | 1.42 (1.22–1.64) | 0.005 | 0.96–2.09 | 0.064 | 0.146 | 0.000 | 0 | 0.932 |
| CheckMate 914 B: nivolumab + ipilimumab (raster curve) replaces nivolumab (SAP 9, item 8) | ICI | main | 4 | KEYNOTE-564; IMmotion010; CheckMate 914 part A; CheckMate 914 part B, nivolumab + ipilimumab | 1.39 (1.12–1.72) | 0.017 | 0.94–2.05 | 0.077 | 0.307 | 0.000 | 0 | 0.820 |
| Later (sensitivity) landmarks (SAP 9, item 1; D6) | ICI | sens | 4 | KEYNOTE-564; IMmotion010; CheckMate 914 part A; CheckMate 914 part B, nivolumab | 1.39 (0.96–2.02) | 0.065 | 0.92–2.11 | 0.083 | 0.809 | 0.006 | 0 | 0.435 |
| Adding PROSPER, standard reconstruction (failed QC; D4-A) (SAP 9, item 3) | ICI | main | 5 | KEYNOTE-564; IMmotion010; CheckMate 914 part A; CheckMate 914 part B, nivolumab; PROSPER, standard reconstruction | 1.37 (1.18–1.60) | 0.004 | 1.00–1.88 | 0.049 | 0.229 | 0.000 | 0 | 0.922 |
| Adding PROSPER, censoring-informed reconstruction (post hoc method; D4-C) (SAP 9, item 3) | ICI | main | 5 | KEYNOTE-564; IMmotion010; CheckMate 914 part A; CheckMate 914 part B, nivolumab; PROSPER, censoring-informed reconstruction | 1.36 (1.14–1.61) | 0.008 | 0.99–1.85 | 0.053 | 0.312 | 0.000 | 0 | 0.870 |
| KEYNOTE-564, NEJM 2021 (cut-off Dec 2020) | ICI | main | 4 | KEYNOTE-564, NEJM 2021 (cut-off Dec 2020); | 1.47 (1.06–2.02) | 0.033 | 0.95–2.25 | 0.066 | 0.564 | 0.000 | 0 | 0.638 |

| Analysis | Class | Landmark | k | Comparisons | Pooled post/on-treatment HR ratio, ordinary HK (95% CI) | P | Modified HK 95% CI (q* = max(1, q); post hoc) | P, modified HK | q | τ² | I² (%) | P (Q test) |
|---|---|---|---|---|---|---|---|---|---|---|---|---|
| replaces KEYNOTE-564 (SAP 9, item 2) | | | | IMmotion010; CheckMate 914 part A; CheckMate 914 part B, nivolumab | | | | | | | | |
| KEYNOTE-564, Lancet Oncol 2022 (cut-off Jun 2021) replaces KEYNOTE-564 (SAP 9, item 2) | ICI | main | 4 | KEYNOTE-564, Lancet Oncol 2022 (cut-off Jun 2021); IMmotion010; CheckMate 914 part A; CheckMate 914 part B, nivolumab | 1.34 (1.32–1.36) | <0.001 | 0.88–2.03 | 0.115 | 0.001 | 0.000 | 0 | 1.000 |
| KEYNOTE-564, NEJM 2024 (cut-off Sep 2023) replaces KEYNOTE-564 (SAP 9, item 2) | ICI | main | 4 | KEYNOTE-564, NEJM 2024 (cut-off Sep 2023); IMmotion010; CheckMate 914 part A; CheckMate 914 part B, nivolumab | 1.42 (1.21–1.66) | 0.006 | 0.96–2.10 | 0.066 | 0.159 | 0.000 | 0 | 0.924 |
| Adding PROSPER, Day-1 events censored (Suppl Fig 3) (SAP 9, item 3) | ICI | main | 5 | KEYNOTE-564; IMmotion010; CheckMate 914 part A; CheckMate 914 part B, nivolumab; PROSPER, Day-1 events censored (Suppl Fig 3) | 1.40 (1.26–1.56) | <0.001 | 1.03–1.92 | 0.040 | 0.121 | 0.000 | 0 | 0.975 |
| LITESPARK-022 within-trial ratio, not pooled (descriptive) | HIF-2α | main | 1 | LITESPARK-022, BICR (Fig 1B) | 2.10 (1.44–3.08) | — | — | — | — | — | — | — |
| LITESPARK-022 within-trial ratio, not pooled (descriptive) | HIF-2α | sens | 1 | LITESPARK-022, BICR (Fig 1B) | 2.05 (1.36–3.08) | — | — | — | — | — | — | — |
| LITESPARK-022 within-trial ratio, not pooled (descriptive) | HIF-2α | main | 1 | LITESPARK-022, investigator (Fig 1A), standard reconstruction | 1.80 (1.21–2.68) | — | — | — | — | — | — | — |
| LITESPARK-022 within-trial | HIF-2α | sens | 1 | LITESPARK-022, | 1.70 (1.14–2.53) | — | — | — | — | — | — | — |

| Analysis | Class | Landmark | k | Comparisons | Pooled post/on-treatment HR ratio, ordinary HK (95% CI) | P | Modified HK 95% CI (q* = max(1, q); post hoc) | P, modified HK | q | τ² | I² (%) | P (Q test) |
|---|---|---|---|---|---|---|---|---|---|---|---|---|
| ratio, not pooled (descriptive) | | | | investigator (Fig 1A), standard reconstruction | | | | | | | | |
| LITESPARK-022 within-trial ratio, not pooled (descriptive) | HIF-2α | main | 1 | LITESPARK-022, investigator (Fig 1A), censoring-informed reconstruction | 1.83 (1.24–2.70) | — | — | — | — | — | — | — |
| LITESPARK-022 within-trial ratio, not pooled (descriptive) | HIF-2α | sens | 1 | LITESPARK-022, investigator (Fig 1A), censoring-informed reconstruction | 1.80 (1.22–2.66) | — | — | — | — | — | — | — |

Pooled values are ratios of post-treatment to on-treatment HRs from random-effects meta-analysis (REML, Hartung-Knapp); all pooled analyses from version 1 were re-run and reproduced exactly. “main” and “sens” in the Landmark column denote the main and sensitivity landmarks of each comparison (Supplementary Table S3). With the later (sensitivity) landmarks, the pooled ICI ratio was 1.39 (0.96–2.02; P = 0.065), and its CI included 1. With the modified HK correction, the main VEGFR-TKI pooled ratio was 1.81 (1.29–2.54; P = 0.007; q = 0.981) and the main ICI pooled ratio 1.42 (0.96–2.09; P = 0.064; q = 0.146). Across all substitution and addition analyses at the main landmarks, the pooled VEGFR-TKI ratio ranged from 1.63 to 1.96 (main analysis 1.81) and the pooled ICI ratio from 1.34 to 1.47 (main analysis 1.42); the ordinary HK CI included 1 in 0 of 12 VEGFR-TKI and 0 of 7 ICI analyses, and the modified HK CI in 0 of 12 and 5 of 7. Counting also the pooling with the later (sensitivity) landmarks, the modified HK CI included 1 in 6 of 8 ICI and 0 of 13 VEGFR-TKI sensitivity analyses. At the main landmarks, the 95% CI of the LITESPARK-022 BICR ratio (2.10; 95% CI 1.44–3.08) included the pooled VEGFR-TKI point estimate (1.81) but not the pooled ICI point estimate (1.42), which lay below its lower limit; the 95% CI of the investigator-assessed ratio (censoring-informed reconstruction; 1.83; 95% CI 1.24–2.70) included both pooled point estimates (VEGFR-TKI 1.81; ICI 1.42). With the sensitivity landmarks, the 95% CI of the LITESPARK-022 BICR ratio (2.05; 95% CI 1.36–3.08) included both pooled point estimates (VEGFR-TKI 1.85; ICI 1.39); the 95% CI of the investigator-assessed ratio (censoring-informed reconstruction; 1.80; 95% CI 1.22–2.66) included both pooled point estimates (VEGFR-TKI 1.85; ICI 1.39). These statements describe where a single-trial interval lies relative to pooled point estimates; no comparison or test across trials was performed. Modified HK = Hartung-Knapp variance with q replaced by q* = max(1, q) (post hoc; K5); when q ≥ 1 the modified interval equals the ordinary one. Rows labelled “replaces” substitute one curve added under deviation D8 for the main curve of that trial (SAP section 9, items 2 and 4–7); curves marked as accepted after adjudication are described in Table S1. The LITESPARK-022 ratios are shown for description only; LITESPARK-022 was not pooled and was not formally compared with either class. From version 3, every VEGFR-TKI pooled analysis includes E2810 (D9) except the row “Excluding E2810” (post hoc), which reproduces the version 2 main VEGFR-TKI result.

#### Supplementary Table S7. Overall survival

Sources: analysis output tables (primary RMST, selected flexible parametric models and feasibility, and the corresponding SAP-completion tables and curve list); reported HRs from the QC reports.

##### A. RMST difference and flexible parametric HR(t)

| Comparison | Role | Reported OS HR (CI) | Reconstructed deaths, exp/ctrl | Primary τ (mo) | ΔRMST at primary τ, mo (95% CI) | RMST ratio (95% CI) | ΔRMST at 24 mo (95% CI) | Flexible parametric HR(t) at maximum feasible τ, AIC-selected [spec] | HR(t) at same time, fixed df 3/tvc 2 | Note |
|---|---|---|---|---|---|---|---|---|---|---|
| LITESPARK-022, standard reconstruction | Descriptive | 0.78 (95% CI 0.51–1.19) | 35/46 | 24 | 0.04 (−0.18 to 0.25) | 1.002 (0.992–1.011) | — (primary τ = 24) | 0.56 (0.25–1.26) at 30 mo [df 2/tvc 1] | 0.41 (0.14–1.17) | Standard reconstruction failed the event- |

| Comparison | Role | Reported OS HR (CI) | Reconstructed deaths, exp/ctrl | Primary τ (mo) | ΔRMST at primary τ, mo (95% CI) | RMST ratio (95% CI) | ΔRMST at 24 mo (95% CI) | Flexible parametric HR(t) at maximum feasible τ, AIC-selected [spec] | HR(t) at same time, fixed df 3/tvc 2 | Note |
|---|---|---|---|---|---|---|---|---|---|---|
| | | | | | | | | | | count criterion (D2); descriptive only |
| LITESPARK-022, censoring-informed reconstruction | Descriptive | 0.78 (95% CI 0.51–1.19) | 38/49 | 24 | 0.04 (−0.18 to 0.25) | 1.002 (0.993–1.011) | — (primary τ = 24) | 0.61 (0.29–1.29) at 30 mo [df 2/tvc 1] | 0.50 (0.20–1.25) | Censoring-informed reconstruction (post hoc; D2); descriptive only |
| KEYNOTE-564 | Main | 0.66 (95% CI 0.48–0.90) | 68/99 | 36 | 0.60 (−0.01 to 1.20) | 1.017 (1.000–1.035) | 0.17 (−0.13 to 0.47) | 0.70 (0.43–1.15) at 75 mo [df 2/tvc 1] | 0.87 (0.46–1.63) | |
| ASSURE, sunitinib | Main | 1.17 (97.5% CI 0.90–1.52) | 153/138 | 36 | −0.33 (−1.05 to 0.39) | 0.990 (0.970–1.012) | −0.14 (−0.50 to 0.22) | 1.21 (0.83–1.75) at 84 mo [df 4/tvc 1] | 1.25 (0.79–1.98) | |
| ASSURE, sorafenib | Main | 0.98 (97.5% CI 0.75–1.28) | 135/138 | 36 | 0.12 (−0.56 to 0.80) | 1.004 (0.984–1.024) | 0.10 (−0.24 to 0.44) | 1.06 (0.70–1.61) at 84 mo [df 2/tvc 1] | 1.02 (0.61–1.71) | |
| IMmotion010 | Main | 0.97 (95% CI 0.67–1.42) | 52/51 | 36 | 0.49 (−0.25 to 1.24) | 1.014 (0.993–1.036) | 0.32 (−0.05 to 0.68) | 1.38 (0.76–2.51) at 54 mo [df 1/tvc 1] | 1.20 (0.54–2.63) | |
| S-TRAC (updated analysis) | Main | 0.92 (95% CI 0.66–1.28) | 65/73 | 36 | −0.02 (−0.84 to 0.81) | 1.000 (0.976–1.024) | 0.03 (−0.35 to 0.42) | 0.87 (0.53–1.42) at 84 mo [df 1/tvc 1] | 0.79 (0.41–1.54) | |
| PROTECT, ITT 600 mg (interim) | Main | 0.79 (95% CI 0.57–1.09) | 60/78 | 36 | 0.55 (0.01–1.10) | 1.016 (1.000–1.032) | 0.20 (−0.02 to 0.41) | 1.02 (0.56–1.85) at 54 mo [df 4/tvc 1] | 1.02 (0.50–2.07) | Deaths per arm not reported in the publication (reported total compared in Table S5) |
| PROTECT, ITT all (final; Eur Urol 2021) [did not pass the authors' visual check; shown for reference only, not used in any pooled analysis] | Sensitivity | 1.00 (95% CI 0.80–1.26) | 146/143 | 36 | 0.60 (0.08–1.13) | 1.018 (1.002–1.033) | 0.32 (0.07–0.56) | 3.77 (1.91–7.41) at 90 mo [df 3/tvc 2] | 3.77 (1.91–7.41) | Did not pass the authors' visual check; shown for reference only, not used in any pooled analysis (K7); sensitivity curve |
| SORCE, sorafenib 3 yr | Main | 1.06 (95% CI 0.82–1.38) | 142/93 | 36 | 0.06 (−0.62 to 0.75) | 1.002 (0.982–1.022) | −0.01 (−0.35 to 0.33) | 1.08 (0.70–1.66) at 96 mo [df 2/tvc 1] | 1.09 (0.66–1.83) | |
| SORCE, sorafenib 1 yr | Main | 0.92 (95% CI 0.71–1.20) | 131/93 | 36 | 0.48 (−0.15 to 1.11) | 1.014 (0.996–1.033) | 0.23 (−0.07 to 0.53) | 0.79 (0.53–1.19) at 96 mo [df 5/tvc 2] | 0.85 (0.50–1.43) | |
| CheckMate 914 part A | Main | Not reported | 33/28 | 36 | −0.44 (−1.17 to 0.28) | 0.987 (0.967–1.008) | −0.23 (−0.62 to 0.16) | 1.40 (0.56–3.50) at 45 mo [df 3/tvc 2] | 1.40 (0.56–3.50) | No OS HR reported; raster figure |
| E2810 [OS accepted after adjudication (reported HR 2.55 vs HR 2.11 derived exactly from the curve; reported model not stated — see Table S1, D9)] | Main | 2.55 (95% CI 1.23–5.27) | 24/12 | 36 | −1.32 (−3.46 to 0.82) | 0.962 (0.903–1.025) | −0.30 (−1.39 to 0.79) | 1.85 (0.67–5.05) at 72 mo [df 1/tvc 1] | 1.88 (0.56–6.29) | OS accepted after adjudication (reported HR 2.55 vs HR 2.11 derived exactly from the curve; reported model not stated — see Table S1, D9) |

OS was analysed with RMST differences and flexible parametric HR(t) (SAP 8.5); piecewise HRs are given for description only (K4). LITESPARK-022 OS is from the first interim analysis (87 deaths) and is descriptive only (D2). Reported HRs: 95% CI except ASSURE (97.5% CI). Rows from IMmotion010 to CheckMate 914 part A were added under deviation D8; E2810 (last row) was added under D9; OS accepted after adjudication (reported HR 2.55 vs HR 2.11 derived exactly from the curve; reported model not stated — see Table S1, D9). ATLAS and CheckMate 914 part B published no OS Kaplan-Meier curve (main text and supplements checked page by page; deviations log, D8), so OS could not be analysed as specified in SAP 8.5; reported values are given here for completeness. ATLAS (interim analysis, ITT): 28 vs 26 deaths (axitinib vs placebo); reported OS HR 1.026 (0.600–1.756), P = 0.9246. CheckMate 914 part B: 19/411 deaths with nivolumab vs 8/208 with placebo (8/206 with nivolumab plus ipilimumab); OS HR not reported (immature). PROSPER (sensitivity trial): the OS curve (Supplemental Fig 2) was not reconstructed: raster resolution insufficient (768×480 px; overlapping curves); reported values described: 48/404 vs 40/415 deaths (Supplemental Fig 2) and OS HR 1.28 (0.84–1.95) (main text) (Table S1, D8-g).

##### B. Piecewise HRs for OS (descriptive)

Source: analysis output table of piecewise Cox models (SAP-completion run).

| Comparison | Landmark (mo) | Deaths before landmark, exp/ctrl | Deaths after landmark, exp/ctrl | HR before landmark (95% CI) | HR after landmark (95% CI) | Ratio (95% CI) |
|---|---|---|---|---|---|---|
| KEYNOTE-564 | 12 (main) | 7/10 | 61/89 | 0.70 (0.27–1.85) | 0.66 (0.47–0.91) | 0.93 (0.34–2.59) |
| KEYNOTE-564 | 15 (sensitivity) | 7/12 | 61/87 | 0.59 (0.23–1.49) | 0.67 (0.48–0.93) | 1.15 (0.43–3.08) |
| ASSURE, sunitinib | 12 (main) | 20/21 | 133/117 | 0.97 (0.53–1.80) | 1.20 (0.94–1.54) | 1.23 (0.64–2.39) |
| ASSURE, sunitinib | 15 (sensitivity) | 35/29 | 118/109 | 1.24 (0.76–2.03) | 1.14 (0.88–1.49) | 0.92 (0.53–1.61) |
| ASSURE, sorafenib | 12 (main) | 14/21 | 121/117 | 0.68 (0.34–1.33) | 1.06 (0.82–1.37) | 1.57 (0.76–3.23) |
| ASSURE, sorafenib | 15 (sensitivity) | 23/29 | 112/109 | 0.81 (0.47–1.39) | 1.05 (0.81–1.37) | 1.31 (0.71–2.40) |
| LITESPARK-022, censoring-informed reconstruction | 12 (main) | 16/13 | 22/36 | 1.23 (0.59–2.56) | 0.61 (0.36–1.04) | 0.50 (0.20–1.22) |
| LITESPARK-022, censoring-informed reconstruction | 15 (sensitivity) | 19/22 | 19/27 | 0.87 (0.47–1.60) | 0.70 (0.39–1.26) | 0.81 (0.35–1.89) |
| LITESPARK-022, standard reconstruction | 12 (main) | 16/13 | 19/33 | 1.23 (0.59–2.56) | 0.58 (0.33–1.02) | 0.47 (0.19–1.18) |
| LITESPARK-022, standard reconstruction | 15 (sensitivity) | 19/22 | 16/24 | 0.87 (0.47–1.60) | 0.67 (0.36–1.26) | 0.77 (0.32–1.87) |
| IMmotion010 | 12 (main) | 4/10 | 48/41 | 0.39 (0.12–1.24) | 1.11 (0.73–1.69) | 2.87 (0.84–9.85) |
| IMmotion010 | 15 (sensitivity) | 8/15 | 44/36 | 0.51 (0.22–1.21) | 1.16 (0.75–1.80) | 2.25 (0.86–5.91) |
| S-TRAC (updated analysis) | 12 (main) | 3/5 | 62/68 | 0.61 (0.15–2.57) | 0.98 (0.69–1.38) | 1.59 (0.36–6.94) |
| S-TRAC (updated analysis) | 15 (sensitivity) | 7/6 | 58/67 | 1.20 (0.40–3.58) | 0.93 (0.65–1.32) | 0.77 (0.25–2.43) |
| PROTECT, ITT 600 mg (interim) | 12 (main) | 2/4 | 58/74 | 0.51 (0.09–2.78) | 0.79 (0.56–1.11) | 1.55 (0.27–8.74) |
| PROTECT, ITT 600 mg (interim) | 15 (sensitivity) | 8/13 | 52/65 | 0.62 (0.26–1.51) | 0.80 (0.56–1.16) | 1.28 (0.50–3.33) |
| PROTECT, ITT all (final; Eur Urol 2021) [did not pass the authors' visual check; shown for reference only, not used in any pooled analysis] | 12 (main) | 7/12 | 139/131 | 0.60 (0.24–1.53) | 1.08 (0.85–1.37) | 1.80 (0.69–4.70) |
| PROTECT, ITT all (final; Eur Urol 2021) [did not pass the authors' visual check; shown for reference only, not used in any pooled analysis] | 15 (sensitivity) | 7/25 | 139/118 | 0.29 (0.12–0.67) | 1.20 (0.94–1.53) | 4.16 (1.74–9.96) |
| SORCE, sorafenib 3 yr | 36 (main) | 63/45 | 79/48 | 0.95 (0.65–1.40) | 1.12 (0.79–1.61) | 1.18 (0.70–2.00) |
| SORCE, sorafenib 3 yr | 12 (sensitivity) | 13/11 | 129/82 | 0.80 (0.36–1.79) | 1.07 (0.81–1.42) | 1.34 (0.57–3.13) |
| SORCE, sorafenib 1 yr | 12 (main) | 5/11 | 126/82 | 0.30 (0.11–0.88) | 0.98 (0.74–1.30) | 3.23 (1.08–9.65) |
| SORCE, sorafenib 1 yr | 15 (sensitivity) | 8/13 | 123/80 | 0.41 (0.17–0.99) | 0.98 (0.74–1.31) | 2.39 (0.95–6.04) |
| CheckMate 914 part A | 6 (main) | 6/1 | 27/27 | 6.07 (0.73–50.45) | 1.01 (0.60–1.73) | 0.17 (0.02–1.48) |
| CheckMate 914 part A | 9 (sensitivity) | 10/4 | 23/24 | 2.54 (0.80–8.11) | 0.97 (0.55–1.72) | 0.38 (0.10–1.39) |
| E2810 [OS accepted after adjudication (reported HR 2.55 vs HR 2.11 derived exactly from the curve; reported model not stated — see Table S1, | 12 (main) | 1/2 | 23/10 | 0.50 (0.05–5.52) | 2.47 (1.17–5.19) | 4.94 (0.40–60.92) |

| Comparison | Landmark (mo) | Deaths before landmark, exp/ctrl | Deaths after landmark, exp/ctrl | HR before landmark (95% CI) | HR after landmark (95% CI) | Ratio (95% CI) |
|---|---|---|---|---|---|---|
| D9)] | | | | | | |
| E2810 [OS accepted after adjudication (reported HR 2.55 vs HR 2.11 derived exactly from the curve; reported model not stated — see Table S1, D9)] | 15 (sensitivity) | 3/2 | 21/10 | 1.51 (0.25–9.01) | 2.26 (1.07–4.81) | 1.50 (0.22–10.48) |

Landmarks are those of the trial's DFS analysis; values are descriptive only (K4).

#### Supplementary Table S8. Restricted mean disease-free survival time at 24 months and at the primary truncation time

Source: analysis output table of primary RMST results.

| Comparison | Role | τ (mo) | RMST, experimental arm (95% CI) | RMST, control arm (95% CI) | ΔRMST, mo (95% CI) | P | RMST ratio (95% CI) |
|---|---|---|---|---|---|---|---|
| LITESPARK-022, BICR (Fig 1B) | Main | 24 (primary) | 21.36 (20.98–21.74) | 20.02 (19.54–20.49) | 1.34 (0.73–1.95) | <0.001 | 1.067 (1.036–1.099) |
| KEYNOTE-564 | Main | 36 (primary) | 30.05 (29.07–31.03) | 26.83 (25.68–27.99) | 3.21 (1.70–4.73) | <0.001 | 1.120 (1.061–1.182) |
| KEYNOTE-564 | Main | 24 | 21.02 (20.45–21.59) | 19.04 (18.34–19.74) | 1.98 (1.08–2.88) | <0.001 | 1.104 (1.055–1.156) |
| IMmotion010 | Main | 36 (primary) | 26.90 (25.61–28.19) | 26.06 (24.70–27.42) | 0.84 (−1.03 to 2.71) | 0.378 | 1.032 (0.962–1.108) |
| IMmotion010 | Main | 24 | 19.34 (18.57–20.12) | 18.64 (17.81–19.48) | 0.70 (−0.44 to 1.84) | 0.228 | 1.038 (0.977–1.102) |
| CheckMate 914 part A | Main | 36 (primary) | 29.08 (27.90–30.27) | 28.50 (27.28–29.72) | 0.58 (−1.12 to 2.28) | 0.503 | 1.020 (0.962–1.083) |
| CheckMate 914 part A | Main | 24 | 20.48 (19.79–21.17) | 20.04 (19.32–20.76) | 0.44 (−0.56 to 1.44) | 0.384 | 1.022 (0.973–1.074) |
| CheckMate 914 part B, nivolumab | Main | 24 (primary) | 20.53 (19.87–21.20) | 19.69 (18.63–20.74) | 0.85 (−0.40 to 2.09) | 0.182 | 1.043 (0.980–1.110) |
| CheckMate 914 part B, nivolumab + ipilimumab | Alternative | 24 (primary) | 19.45 (18.38–20.52) | 19.68 (18.63–20.74) | −0.23 (−1.73 to 1.27) | 0.764 | 0.988 (0.915–1.067) |
| S-TRAC | Main | 36 (primary) | 28.91 (27.59–30.23) | 26.63 (25.14–28.12) | 2.28 (0.29–4.27) | 0.025 | 1.086 (1.010–1.167) |
| S-TRAC | Main | 24 | 20.71 (19.97–21.45) | 19.09 (18.18–20.00) | 1.62 (0.44–2.79) | 0.007 | 1.085 (1.022–1.151) |
| ASSURE, sunitinib | Main | 36 (primary) | 28.09 (27.15–29.03) | 27.65 (26.66–28.63) | 0.44 (−0.92 to 1.80) | 0.527 | 1.016 (0.967–1.067) |
| ASSURE, sunitinib | Main | 24 | 20.16 (19.61–20.70) | 19.61 (19.02–20.21) | 0.54 (−0.27 to 1.35) | 0.189 | 1.028 (0.987–1.070) |
| ASSURE, sorafenib | Alternative | 36 (primary) | 28.66 (27.76–29.57) | 27.65 (26.66–28.63) | 1.02 (−0.32 to 2.35) | 0.136 | 1.037 (0.989–1.087) |
| ASSURE, sorafenib | Alternative | 24 | 20.47 (19.95–21.00) | 19.61 (19.02–20.21) | 0.86 (0.07–1.65) | 0.033 | 1.044 (1.003–1.086) |
| PROTECT | Main | 36 (primary) | 29.03 (28.06–30.00) | 27.13 (26.03–28.22) | 1.91 (0.44–3.37) | 0.011 | 1.070 (1.016–1.128) |
| PROTECT | Main | 24 | 20.74 (20.20–21.28) | 19.29 (18.63–19.94) | 1.45 (0.60–2.30) | <0.001 | 1.075 (1.030–1.122) |
| ATLAS | Main | 36 (primary) | 28.82 (27.52–30.12) | 27.57 (26.18–28.97) | 1.25 (−0.66 to 3.16) | 0.198 | 1.045 (0.977–1.119) |
| ATLAS | Main | 24 | 20.22 (19.45–21.00) | 19.40 (18.56–20.23) | 0.83 (−0.32 to 1.97) | 0.156 | 1.043 (0.984–1.105) |
| SORCE, sorafenib 1 yr | Main | 36 (primary) | 30.66 (29.88–31.44) | 29.10 (27.97–30.22) | 1.57 (0.20–2.94) | 0.025 | 1.054 (1.006–1.104) |
| SORCE, sorafenib 1 yr | Main | 24 | 21.56 (21.12–21.99) | 20.35 (19.68–21.01) | 1.21 (0.41–2.00) | 0.003 | 1.059 (1.019–1.101) |
| SORCE, sorafenib 3 yr | Alternative | 36 (primary) | 30.17 (29.33–31.02) | 29.10 (27.97–30.22) | 1.08 (−0.33 to 2.48) | 0.134 | 1.037 (0.989–1.088) |
| SORCE, sorafenib 3 yr | Alternative | 24 | 21.09 (20.60–21.58) | 20.35 (19.68–21.02) | 0.74 (−0.08 to 1.56) | 0.078 | 1.036 (0.996–1.079) |
| E2810 (M1 NED after metastasectomy) | Main | 36 (primary) | 20.06 (17.11–23.00) | 17.91 (14.64–21.18) | 2.15 (−2.25 to 6.55) | 0.338 | 1.120 (0.886–1.416) |
| E2810 (M1 NED after metastasectomy) | Main | 24 | 16.32 (14.56–18.07) | 14.62 (12.37–16.88) | 1.69 (−1.16 to 4.55) | 0.245 | 1.116 (0.925–1.346) |
| LITESPARK-022, investigator (Fig 1A), standard reconstruction | Sensitivity | 24 (primary) | 21.98 (21.65–22.32) | 20.69 (20.26–21.12) | 1.29 (0.75–1.84) | <0.001 | 1.063 (1.036–1.090) |
| LITESPARK-022, investigator (Fig 1A), censoring-informed reconstruction | Sensitivity | 24 (primary) | 21.98 (21.65–22.31) | 20.69 (20.26–21.11) | 1.29 (0.75–1.83) | <0.001 | 1.062 (1.036–1.090) |
| PROSPER, standard reconstruction | Sensitivity | 36 (primary) | 26.56 (25.10–28.02) | 26.15 (24.69–27.61) | 0.41 (−1.65 to 2.48) | 0.697 | 1.016 (0.939–1.099) |
| PROSPER, standard | Sensitivity | 24 | 18.56 (17.64–19.48) | 18.24 (17.33–19.14) | 0.32 (−0.97 to 1.61) | 0.626 | 1.018 (0.949–1.092) |

| Comparison | Role | τ (mo) | RMST, experimental arm (95% CI) | RMST, control arm (95% CI) | ΔRMST, mo (95% CI) | P | RMST ratio (95% CI) |
|---|---|---|---|---|---|---|---|
| reconstruction | | | | | | | |
| PROSPER, censoring-informed reconstruction | Sensitivity | 36 (primary) | 26.35 (24.89–27.81) | 25.85 (24.39–27.31) | 0.50 (−1.56 to 2.56) | 0.635 | 1.019 (0.942–1.103) |
| PROSPER, censoring-informed reconstruction | Sensitivity | 24 | 18.50 (17.58–19.42) | 18.12 (17.22–19.03) | 0.37 (−0.92 to 1.66) | 0.572 | 1.021 (0.951–1.095) |
| SORCE, sorafenib 3 yr vs 1 yr | Exploratory | 36 (primary) | 30.17 (29.33–31.02) | 30.66 (29.88–31.44) | −0.49 (−1.64 to 0.66) | 0.403 | 0.984 (0.948–1.022) |
| SORCE, sorafenib 3 yr vs 1 yr | Exploratory | 24 | 21.09 (20.60–21.58) | 21.56 (21.12–21.99) | −0.47 (−1.11 to 0.18) | 0.160 | 0.978 (0.949–1.009) |

RMST in months; ΔRMST = experimental minus control; asymptotic CIs from `survRM2::rmst2`. For every comparison with a primary τ of 36 months, the row marked "(primary)" gives ΔRMST at 36 months with its 95% CI; no 36-month value is given for LITESPARK-022 (all 3 DFS curves) or CheckMate 914 part B (both comparisons), whose primary τ was 24 months because fewer than 10% of an arm were at risk at 36 months (Supplementary Table S3). For PROSPER the endpoint is RFS. The SORCE 3-year vs 1-year comparison is exploratory (SAP 9A; J15), with the 1-year arm as the reference.

### Supplementary Table S9. ΔRMST over the truncation time and post-treatment increment

Source: analysis output tables (RMST curves, RMST increments and bootstrap increments).

##### A. ΔRMST (months; pointwise asymptotic 95% CI) at selected truncation times

| Comparison | Role | τ = 12 | τ = 24 | τ = 36 | τ = 48 | τ = 60 | Maximum feasible τ |
|---|---|---|---|---|---|---|---|
| LITESPARK-022, BICR (Fig 1B) | Main | 0.54 (0.31–0.76) | 1.34 (0.73–1.95) | — | — | — | 1.65 (0.82–2.48) [τ = 30] |
| KEYNOTE-564 | Main | 0.72 (0.38–1.06) | 1.98 (1.08–2.88) | 3.21 (1.70–4.73) | 4.31 (2.16–6.46) | 5.44 (2.64–8.24) | 6.74 (3.11–10.37) [τ = 75] |
| IMmotion010 | Main | 0.39 (−0.06 to 0.83) | 0.70 (−0.44 to 1.84) | 0.84 (−1.03 to 2.71) | 0.78 (−1.85 to 3.41) | — | 0.78 (−1.85 to 3.41) [τ = 48] |
| CheckMate 914 part A | Main | 0.24 (−0.11 to 0.60) | 0.44 (−0.56 to 1.44) | 0.58 (−1.12 to 2.28) | — | — | 0.56 (−1.32 to 2.45) [τ = 39] |
| CheckMate 914 part B, nivolumab | Main | 0.34 (−0.10 to 0.79) | 0.85 (−0.40 to 2.09) | — | — | — | 1.00 (−0.47 to 2.46) [τ = 27] |
| CheckMate 914 part B, nivolumab + ipilimumab | Alternative | −0.03 (−0.58 to 0.52) | −0.23 (−1.73 to 1.27) | — | — | — | −0.24 (−2.00 to 1.51) [τ = 27] |
| S-TRAC | Main | 0.81 (0.35–1.26) | 1.62 (0.44–2.79) | 2.28 (0.29–4.27) | 3.11 (0.24–5.98) | 3.99 (0.22–7.76) | 4.87 (0.20–9.55) [τ = 72] |
| ASSURE, sunitinib | Main | 0.31 (0.00–0.63) | 0.54 (−0.27 to 1.35) | 0.44 (−0.92 to 1.80) | 0.24 (−1.70 to 2.19) | −0.06 (−2.60 to 2.48) | −0.29 (−3.44 to 2.86) [τ = 72] |
| ASSURE, sorafenib | Alternative | 0.41 (0.11–0.72) | 0.86 (0.07–1.65) | 1.02 (−0.32 to 2.35) | 0.98 (−0.93 to 2.89) | 0.65 (−1.85 to 3.15) | 0.40 (−2.70 to 3.50) [τ = 72] |
| PROTECT | Main | 0.66 (0.35–0.96) | 1.45 (0.60–2.30) | 1.91 (0.44–3.37) | 1.95 (−0.16 to 4.06) | — | 1.95 (−0.16 to 4.06) [τ = 48] |
| ATLAS | Main | 0.31 (−0.14 to 0.77) | 0.83 (−0.32 to 1.97) | 1.25 (−0.66 to 3.16) | — | — | 1.25 (−0.66 to 3.16) [τ = 36] |
| SORCE, sorafenib 1 yr | Main | 0.54 (0.26–0.82) | 1.21 (0.41–2.00) | 1.57 (0.20–2.94) | 1.68 (−0.30 to 3.67) | 1.78 (−0.83 to 4.39) | 2.12 (−2.42 to 6.66) [τ = 96] |
| SORCE, sorafenib 3 yr | Alternative | 0.32 (0.02–0.61) | 0.74 (−0.08 to 1.56) | 1.08 (−0.33 to 2.48) | 1.00 (−1.02 to 3.02) | 0.72 (−1.93 to 3.37) | 0.40 (−4.19 to 5.00) [τ = 96] |
| E2810 (M1 NED after metastasectomy) | Main | 1.61 (0.44–2.77) | 1.69 (−1.16 to 4.55) | 2.15 (−2.25 to 6.55) | 2.29 (−3.59 to 8.18) | — | 2.29 (−3.59 to 8.18) [τ = 48] |
| LITESPARK-022, investigator (Fig 1A), standard reconstruction | Sensitivity | 0.45 (0.27–0.64) | 1.29 (0.75–1.84) | — | — | — | 1.71 (0.96–2.47) [τ = 30] |
| LITESPARK-022, investigator (Fig 1A), censoring-informed reconstruction | Sensitivity | 0.45 (0.26–0.63) | 1.29 (0.75–1.83) | — | — | — | 1.71 (0.96–2.47) [τ = 30] |

| Comparison | Role | τ = 12 | τ = 24 | τ = 36 | τ = 48 | τ = 60 | Maximum feasible τ |
|---|---|---|---|---|---|---|---|
| PROSPER, standard reconstruction | Sensitivity | 0.00 (−0.58 to 0.58) | 0.32 (−0.97 to 1.61) | 0.41 (−1.65 to 2.48) | — | — | 0.41 (−1.65 to 2.48) [τ = 36] |
| PROSPER, censoring-informed reconstruction | Sensitivity | 0.02 (−0.56 to 0.60) | 0.37 (−0.92 to 1.66) | 0.50 (−1.56 to 2.56) | — | — | 0.50 (−1.56 to 2.56) [τ = 36] |

"—" = beyond the maximum feasible τ. The full curves (every 3 months) are in the analysis output table of RMST curves and in Figure 1 of the manuscript and Supplementary Figure S1.

##### B. Post-treatment increment (main landmark to maximum feasible τ)

| Comparison | Role | Landmark (mo) | Maximum feasible τ (mo) | Years after landmark | ΔRMST at landmark (asymptotic 95% CI) | ΔRMST at maximum τ (asymptotic 95% CI) | Post-treatment increment, mo (bootstrap 95% CI) | Increment per year, mo/yr (bootstrap 95% CI) | Proportion of resamples with increment > 0 | Failed resamples |
|---|---|---|---|---|---|---|---|---|---|---|
| LITESPARK-022, BICR (Fig 1B) | Main | 12 | 30 | 1.50 | 0.54 (0.31–0.76) | 1.65 (0.82–2.48) | 1.12 (0.48–1.78) | 0.74 (0.32–1.19) | 1.000 | 0 |
| KEYNOTE-564 | Main | 12 | 75 | 5.25 | 0.72 (0.38–1.06) | 6.74 (3.11–10.37) | 6.02 (2.62–9.33) | 1.15 (0.50–1.78) | 1.000 | 0 |
| IMmotion010 | Main | 12 | 48 | 3.00 | 0.39 (−0.06 to 0.83) | 0.78 (−1.85 to 3.41) | 0.40 (−1.94 to 2.72) | 0.13 (−0.65 to 0.91) | 0.618 | 0 |
| CheckMate 914 part A | Main | 6 | 39 | 2.75 | 0.03 (−0.07 to 0.12) | 0.56 (−1.32 to 2.45) | 0.54 (−1.33 to 2.36) | 0.20 (−0.48 to 0.86) | 0.700 | 0 |
| CheckMate 914 part B, nivolumab | Main | 6 | 27 | 1.75 | 0.09 (−0.03 to 0.21) | 1.00 (−0.47 to 2.46) | 0.91 (−0.49 to 2.22) | 0.52 (−0.28 to 1.27) | 0.898 | 0 |
| CheckMate 914 part B, nivolumab + ipilimumab | Alternative | 6 | 27 | 1.75 | −0.02 (−0.17 to 0.14) | −0.24 (−2.00 to 1.51) | −0.23 (−2.01 to 1.42) | −0.13 (−1.15 to 0.81) | 0.390 | 0 |
| S-TRAC | Main | 12 | 72 | 5.00 | 0.81 (0.35–1.26) | 4.87 (0.20–9.55) | 4.07 (−0.20 to 8.43) | 0.81 (−0.04 to 1.69) | 0.967 | 0 |
| ASSURE, sunitinib | Main | 12 | 72 | 5.00 | 0.31 (0.00–0.63) | −0.29 (−3.44 to 2.86) | −0.61 (−3.46 to 2.40) | −0.12 (−0.69 to 0.48) | 0.333 | 0 |
| ASSURE, sorafenib | Alternative | 12 | 72 | 5.00 | 0.41 (0.11–0.72) | 0.40 (−2.70 to 3.50) | −0.01 (−2.89 to 2.94) | 0.00 (−0.58 to 0.59) | 0.496 | 0 |
| PROTECT | Main | 12 | 48 | 3.00 | 0.66 (0.35–0.96) | 1.95 (−0.16 to 4.06) | 1.29 (−0.62 to 3.21) | 0.43 (−0.21 to 1.07) | 0.891 | 0 |
| ATLAS | Main | 12 | 36 | 2.00 | 0.31 (−0.14 to 0.77) | 1.25 (−0.66 to 3.16) | 0.94 (−0.60 to 2.47) | 0.47 (−0.30 to 1.23) | 0.890 | 0 |
| SORCE, sorafenib 1 yr | Main | 12 | 96 | 7.00 | 0.54 (0.26–0.82) | 2.12 (−2.42 to 6.66) | 1.58 (−2.83 to 6.02) | 0.23 (−0.40 to 0.86) | 0.762 | 0 |
| SORCE, sorafenib 3 yr | Alternative | 36 | 96 | 5.00 | 1.08 (−0.33 to 2.48) | 0.40 (−4.19 to 5.00) | −0.67 (−4.00 to 2.91) | −0.13 (−0.80 to 0.58) | 0.332 | 0 |
| E2810 (M1 NED after metastasectomy) | Main | 12 | 48 | 3.00 | 1.61 (0.44–2.77) | 2.29 (−3.59 to 8.18) | 0.69 (−4.51 to 5.51) | 0.23 (−1.50 to 1.84) | 0.581 | 0 |
| LITESPARK-022, investigator (Fig 1A), standard reconstruction | Sensitivity | 12 | 30 | 1.50 | 0.45 (0.27–0.64) | 1.71 (0.96–2.47) | 1.26 (0.65–1.89) | 0.84 (0.43–1.26) | 1.000 | 0 |
| LITESPARK-022, investigator (Fig 1A), censoring-informed reconstruction | Sensitivity | 12 | 30 | 1.50 | 0.45 (0.26–0.63) | 1.71 (0.96–2.47) | 1.27 (0.65–1.89) | 0.84 (0.44–1.26) | 1.000 | 0 |
| PROSPER, standard reconstruction | Sensitivity | 12 | 36 | 2.00 | 0.00 (−0.58 to 0.58) | 0.41 (−1.65 to 2.48) | 0.41 (−1.11 to 1.95) | 0.20 (−0.55 to 0.97) | 0.691 | 0 |
| PROSPER, censoring-informed reconstruction | Sensitivity | 12 | 36 | 2.00 | 0.02 (−0.56 to 0.60) | 0.50 (−1.56 to 2.56) | 0.48 (−1.05 to 1.98) | 0.24 (−0.53 to 0.99) | 0.718 | 0 |

Increment = ΔRMST at the maximum feasible τ minus ΔRMST at the main landmark; increment per year = increment ÷ years after the landmark (SAP 7.2). CIs of the increment and the increment per year are bootstrap percentile intervals (B = 2000; post hoc, D5); CIs of ΔRMST at the landmark and at the maximum τ are asymptotic.

### Supplementary Table S10. Piecewise Cox models

Source: analysis output table of piecewise Cox models.

| Comparison | Role | Landmark (mo) | Events before landmark, exp/ctrl | Events after landmark, exp/ctrl | On-treatment HR (95% CI) | Post-treatment HR (95% CI) | Post/on-treatment HR ratio (95% CI) | P (interaction) |
|---|---|---|---|---|---|---|---|---|
| LITESPARK-022, BICR (Fig 1B) | Main | 12 (main) | 95/166 | 104/84 | 0.55 (0.43–0.71) | 1.16 (0.87–1.54) | 2.10 (1.44–3.08) | <0.001 |
| LITESPARK-022, BICR (Fig 1B) | Main | 15 (sensitivity) | 121/191 | 78/59 | 0.61 (0.48–0.76) | 1.24 (0.88–1.74) | 2.05 (1.36–3.08) | <0.001 |
| KEYNOTE-564 | Main | 12 (main) | 69/118 | 118/121 | 0.56 (0.42–0.75) | 0.88 (0.68–1.13) | 1.57 (1.06–2.32) | 0.023 |
| KEYNOTE-564 | Main | 15 (sensitivity) | 78/131 | 109/108 | 0.57 (0.43–0.75) | 0.91 (0.70–1.18) | 1.60 (1.09–2.35) | 0.017 |
| IMmotion010 | Main | 12 (main) | 85/98 | 79/68 | 0.83 (0.62–1.11) | 1.10 (0.79–1.52) | 1.32 (0.85–2.04) | 0.210 |
| IMmotion010 | Main | 15 (sensitivity) | 99/102 | 65/64 | 0.93 (0.71–1.23) | 0.96 (0.68–1.35) | 1.03 (0.66–1.60) | 0.902 |
| CheckMate 914 part A | Main | 6 (main) | 42/54 | 67/63 | 0.79 (0.53–1.18) | 1.06 (0.75–1.50) | 1.34 (0.79–2.28) | 0.275 |
| CheckMate 914 part A | Main | 9 (sensitivity) | 54/73 | 55/44 | 0.75 (0.53–1.07) | 1.25 (0.84–1.86) | 1.66 (0.98–2.83) | 0.060 |
| CheckMate 914 part B, nivolumab | Main | 6 (main) | 41/28 | 53/26 | 0.72 (0.44–1.16) | 0.96 (0.60–1.53) | 1.34 (0.68–2.62) | 0.398 |
| CheckMate 914 part B, nivolumab | Main | 9 (sensitivity) | 58/38 | 36/16 | 0.74 (0.49–1.12) | 1.05 (0.58–1.88) | 1.41 (0.69–2.88) | 0.351 |
| CheckMate 914 part B, nivolumab + ipilimumab | Alternative | 6 (main) | 29/28 | 28/26 | 1.03 (0.61–1.73) | 1.10 (0.64–1.87) | 1.07 (0.51–2.25) | 0.858 |
| CheckMate 914 part B, nivolumab + ipilimumab | Alternative | 9 (sensitivity) | 37/38 | 20/16 | 0.98 (0.62–1.53) | 1.27 (0.66–2.44) | 1.30 (0.58–2.88) | 0.523 |
| S-TRAC | Main | 12 (main) | 35/67 | 78/76 | 0.51 (0.34–0.76) | 0.99 (0.73–1.36) | 1.97 (1.17–3.30) | 0.010 |
| S-TRAC | Main | 15 (sensitivity) | 45/75 | 68/68 | 0.58 (0.40–0.84) | 0.97 (0.69–1.35) | 1.66 (1.01–2.74) | 0.046 |
| ASSURE, sunitinib | Main | 12 (main) | 100/123 | 184/164 | 0.80 (0.62–1.05) | 1.17 (0.95–1.45) | 1.45 (1.04–2.04) | 0.030 |
| ASSURE, sunitinib | Main | 15 (sensitivity) | 127/148 | 157/139 | 0.85 (0.67–1.08) | 1.19 (0.94–1.49) | 1.40 (1.00–1.94) | 0.047 |
| ASSURE, sorafenib | Alternative | 12 (main) | 94/123 | 191/164 | 0.74 (0.57–0.97) | 1.15 (0.94–1.42) | 1.56 (1.11–2.19) | 0.011 |
| ASSURE, sorafenib | Alternative | 15 (sensitivity) | 118/148 | 167/139 | 0.77 (0.61–0.98) | 1.20 (0.95–1.50) | 1.55 (1.11–2.16) | 0.009 |
| PROTECT | Main | 12 (main) | 63/111 | 127/87 | 0.55 (0.40–0.75) | 1.42 (1.08–1.86) | 2.59 (1.71–3.91) | <0.001 |
| PROTECT | Main | 15 (sensitivity) | 88/133 | 102/65 | 0.63 (0.48–0.83) | 1.54 (1.13–2.11) | 2.43 (1.61–3.67) | <0.001 |
| ATLAS | Main | 12 (main) | 66/74 | 32/34 | 0.86 (0.61–1.19) | 0.88 (0.54–1.42) | 1.03 (0.57–1.85) | 0.930 |
| ATLAS | Main | 36 (sensitivity) | 93/104 | 5/4 | 0.85 (0.64–1.12) | 1.37 (0.37–5.13) | 1.62 (0.42–6.26) | 0.483 |
| SORCE, sorafenib 1 yr | Main | 12 (main) | 73/78 | 169/88 | 0.59 (0.43–0.82) | 1.24 (0.96–1.61) | 2.09 (1.39–3.15) | <0.001 |
| SORCE, sorafenib 1 yr | Main | 15 (sensitivity) | 86/88 | 156/78 | 0.62 (0.46–0.83) | 1.30 (0.99–1.70) | 2.10 (1.40–3.14) | <0.001 |
| SORCE, sorafenib 3 yr | Alternative | 36 (main) | 171/120 | 73/46 | 0.93 (0.74–1.17) | 1.13 (0.78–1.64) | 1.22 (0.79–1.89) | 0.370 |
| SORCE, sorafenib 3 yr | Alternative | 12 (sensitivity) | 89/78 | 155/88 | 0.74 (0.55–1.01) | 1.20 (0.92–1.56) | 1.61 (1.08–2.41) | 0.020 |
| E2810 (M1 NED after metastasectomy) | Main | 12 (main) | 20/27 | 29/22 | 0.61 (0.34–1.08) | 1.29 (0.74–2.25) | 2.12 (0.95–4.75) | 0.067 |
| E2810 (M1 NED after metastasectomy) | Main | 15 (sensitivity) | 24/31 | 25/18 | 0.63 (0.37–1.08) | 1.40 (0.76–2.59) | 2.21 (0.98–4.98) | 0.055 |
| LITESPARK-022, investigator (Fig 1A), standard reconstruction | Sensitivity | 12 (main) | 70/132 | 103/106 | 0.52 (0.39–0.69) | 0.93 (0.71–1.22) | 1.80 (1.21–2.68) | 0.004 |
| LITESPARK-022, investigator (Fig 1A), standard reconstruction | Sensitivity | 15 (sensitivity) | 92/157 | 81/81 | 0.57 (0.44–0.73) | 0.96 (0.71–1.31) | 1.70 (1.14–2.53) | 0.010 |
| LITESPARK-022, investigator (Fig 1A), censoring-informed reconstruction | Sensitivity | 12 (main) | 71/132 | 113/111 | 0.52 (0.39–0.69) | 0.95 (0.73–1.24) | 1.83 (1.24–2.70) | 0.002 |
| LITESPARK-022, investigator (Fig 1A), censoring-informed | Sensitivity | 15 (sensitivity) | 92/158 | 92/85 | 0.56 (0.43–0.72) | 1.01 (0.75–1.36) | 1.80 (1.22–2.66) | 0.003 |

| Comparison | Role | Landmark (mo) | Events before landmark, exp/ctrl | Events after landmark, exp/ctrl | On-treatment HR (95% CI) | Post-treatment HR (95% CI) | Post/on-treatment HR ratio (95% CI) | P (interaction) |
|---|---|---|---|---|---|---|---|---|
| reconstruction | | | | | | | | |
| PROSPER, standard reconstruction | Sensitivity | 12 (main) | 88/98 | 30/26 | 0.93 (0.70–1.24) | 1.05 (0.62–1.77) | 1.13 (0.62–2.05) | 0.693 |
| PROSPER, standard reconstruction | Sensitivity | 10 (sensitivity) | 86/95 | 32/29 | 0.94 (0.70–1.26) | 1.01 (0.61–1.67) | 1.07 (0.60–1.92) | 0.809 |
| PROSPER, censoring-informed reconstruction | Sensitivity | 12 (main) | 89/100 | 35/33 | 0.92 (0.69–1.23) | 0.99 (0.61–1.59) | 1.07 (0.62–1.87) | 0.803 |
| PROSPER, censoring-informed reconstruction | Sensitivity | 10 (sensitivity) | 87/96 | 37/37 | 0.94 (0.70–1.26) | 0.93 (0.59–1.47) | 1.00 (0.58–1.71) | 0.986 |
| SORCE, sorafenib 3 yr vs 1 yr | Exploratory | 36 (main) | 171/168 | 73/74 | 1.05 (0.84–1.29) | 1.08 (0.78–1.49) | 1.03 (0.70–1.52) | 0.883 |
| SORCE, sorafenib 3 yr vs 1 yr | Exploratory | 12 (sensitivity) | 89/73 | 155/169 | 1.25 (0.92–1.71) | 0.97 (0.78–1.20) | 0.77 (0.53–1.13) | 0.178 |

On-treatment HR = HR before the landmark; post-treatment HR = HR after the landmark among patients still event free; ratio = post-treatment HR ÷ on-treatment HR (exponentiated interaction term), with the Wald P value of the interaction. Post-treatment HRs are conditional on being event free at the landmark and are subject to survivor selection.

#### Supplementary Table S11. Kaplan-Meier disease-free survival differences at 12–60 months

Source: analysis output table of Kaplan-Meier differences.

| Comparison | Role | 12 mo | 24 mo | 36 mo | 48 mo | 60 mo |
|---|---|---|---|---|---|---|
| LITESPARK-022, BICR (Fig 1B) | Main | 89.4 vs 81.6; 7.7 (4.5–11.0) | 77.3 vs 72.4; 4.9 (0.7–9.1) | — | — | — |
| KEYNOTE-564 | Main | 85.6 vs 76.1; 9.5 (4.6–14.4) | 78.3 vs 67.2; 11.1 (5.5–16.7) | 72.5 vs 62.7; 9.8 (3.8–15.7) | 65.1 vs 56.5; 8.6 (2.4–14.8) | 60.8 vs 52.1; 8.7 (2.3–15.1) |
| IMmotion010 | Main | 77.4 vs 74.0; 3.4 (−2.8 to 9.5) | 67.3 vs 64.9; 2.4 (−4.4 to 9.2) | 59.4 vs 59.1; 0.2 (−6.9 to 7.3) | 54.3 vs 52.8; 1.5 (−6.5 to 9.6) | — |
| CheckMate 914 part A | Main | 82.2 vs 79.9; 2.3 (−3.2 to 7.9) | 75.8 vs 73.3; 2.5 (−3.8 to 8.8) | 68.1 vs 67.8; 0.3 (−7.2 to 7.7) | — | — |
| CheckMate 914 part B, nivolumab | Main | 83.0 vs 78.2; 4.8 (−2.1 to 11.7) | 73.9 vs 71.7; 2.3 (−6.1 to 10.7) | — | — | — |
| CheckMate 914 part B, nivolumab + ipilimumab | Alternative | 77.3 vs 80.3; −2.9 (−11.0 to 5.2) | 69.3 vs 69.5; −0.2 (−10.2 to 9.8) | — | — | — |
| S-TRAC | Main | 87.7 vs 77.4; 10.2 (4.1–16.4) | 71.3 vs 67.3; 4.0 (−3.7 to 11.7) | 65.0 vs 59.3; 5.7 (−2.5 to 13.9) | 62.0 vs 54.5; 7.5 (−0.9 to 15.9) | 59.2 vs 51.2; 8.0 (−0.6 to 16.6) |
| ASSURE, sunitinib | Main | 84.0 vs 80.6; 3.3 (−0.9 to 7.5) | 70.1 vs 70.1; 0.0 (−5.2 to 5.1) | 62.7 vs 63.7; −1.0 (−6.5 to 4.4) | 57.6 vs 59.8; −2.2 (−7.9 to 3.4) | 54.1 vs 56.4; −2.3 (−8.2 to 3.5) |
| ASSURE, sorafenib | Alternative | 85.1 vs 80.6; 4.4 (0.3–8.6) | 72.3 vs 70.1; 2.3 (−2.8 to 7.3) | 65.1 vs 63.7; 1.5 (−3.9 to 6.9) | 58.2 vs 59.8; −1.7 (−7.3 to 3.9) | 53.7 vs 56.4; −2.6 (−8.5 to 3.2) |
| PROTECT | Main | 87.8 vs 79.1; 8.7 (4.2–13.2) | 72.6 vs 67.9; 4.7 (−0.9 to 10.3) | 65.9 vs 64.1; 1.8 (−4.1 to 7.7) | 61.0 vs 61.0; 0.1 (−6.3 to 6.4) | — |
| ATLAS | Main | 80.9 vs 78.6; 2.3 (−3.7 to 8.3) | 75.4 vs 70.2; 5.2 (−1.7 to 12.2) | 69.1 vs 66.1; 3.1 (−4.8 to 10.9) | — | — |
| SORCE, sorafenib 1 yr | Main | 88.5 vs 81.8; 6.8 (2.3–11.2) | 79.5 vs 74.7; 4.8 (−0.4 to 10.0) | 73.3 vs 71.8; 1.5 (−4.0 to 7.0) | 70.5 vs 69.8; 0.7 (−5.0 to 6.3) | 67.3 vs 66.7; 0.6 (−5.2 to 6.5) |
| SORCE, sorafenib 3 yr | Alternative | 86.0 vs 81.8; 4.2 (−0.4 to 8.7) | 78.3 vs 74.7; 3.6 (−1.6 to 8.9) | 72.7 vs 71.8; 0.9 (−4.6 to 6.4) | 68.0 vs 69.8; −1.8 (−7.5 to 3.9) | 64.5 vs 66.7; −2.1 (−8.0 to 3.8) |
| E2810 (M1 NED after metastasectomy) | Main | 67.9 vs 56.5; 11.4 (−5.6 to 28.4) | 35.1 vs 37.2; −2.0 (−19.4 to 15.4) | 27.4 vs 21.5; 5.9 (−10.1 to 22.0) | 21.4 vs 21.5; −0.1 (−15.6 to 15.4) | — |
| LITESPARK-022, investigator (Fig 1A), standard reconstruction | Sensitivity | 92.1 vs 85.4; 6.7 (3.8–9.7) | 80.9 vs 74.0; 6.9 (2.8–11.0) | — | — | — |
| LITESPARK-022, investigator (Fig 1A), censoring-informed reconstruction | Sensitivity | 92.0 vs 85.4; 6.7 (3.7–9.6) | 80.9 vs 73.9; 7.0 (2.9–11.1) | — | — | — |
| PROSPER, standard reconstruction | Sensitivity | 76.3 vs 74.1; 2.1 (−4.1 to 8.3) | 70.0 vs 66.6; 3.4 (−3.6 to 10.3) | 63.5 vs 65.1; −1.6 (−9.5 to 6.3) | — | — |
| PROSPER, censoring-informed reconstruction | Sensitivity | 76.0 vs 73.6; 2.4 (−3.9 to 8.6) | 69.0 vs 65.3; 3.6 (−3.4 to 10.6) | 61.9 vs 63.6; −1.7 (−9.5 to 6.2) | — | — |

Each cell: KM DFS (%) in the experimental vs control arm; difference in pp (experimental minus control; Greenwood 95% CI). “—” = time point not feasible under the 10% rule. The KM differences at 30 months for LITESPARK-022 are given in Supplementary Tables S13 and S14.

### Supplementary Table S12. Proportional hazards tests and flexible parametric time-varying HRs

Source: analysis output tables (proportional hazards tests and selected flexible parametric models); all fitted models with their AIC values, and HR(t) every 0.25 months, are in further analysis output tables.

| Comparison | Role | Whole-period Cox HR (95% CI) | Schoenfeld χ² | P | PH rejected (P < 0.05) | AIC-selected model (baseline df/tvc df) | Converged models | ΔAIC to second-best model | HR(t) at landmark, AIC-selected | HR(t) at maximum τ, AIC-selected | HR(t) at landmark, fixed df 3/tvc 2 | HR(t) at maximum τ, fixed df 3/tvc 2 |
|---|---|---|---|---|---|---|---|---|---|---|---|---|
| LITESPARK-022, BICR (Fig 1B) | Main | 0.76 (0.63–0.91) | 15.81 | <0.001 | Yes | df 4/tvc 2 | 15/15 | 1.12 | 0.75 (0.61–0.92) [12 mo] | 2.03 (1.18–3.48) [30 mo] | 0.80 (0.64–0.99) | 1.68 (1.10–2.56) |
| KEYNOTE-564 | Main | 0.72 (0.60–0.88) | 6.54 | 0.011 | Yes | df 4/tvc 1 | 15/15 | 0.99 | 0.73 (0.60–0.89) [12 mo] | 1.03 (0.75–1.43) [75 mo] | 0.72 (0.53–0.99) | 0.99 (0.67–1.47) |
| IMmotion010 | Main | 0.94 (0.76–1.17) | 2.87 | 0.090 | No | df 5/tvc 1 | 15/15 | 0.20 | 1.03 (0.81–1.31) [12 mo] | 1.25 (0.86–1.81) [48 mo] | 1.20 (0.85–1.71) | 1.01 (0.65–1.59) |
| CheckMate 914 part A | Main | 0.94 (0.72–1.22) | 1.73 | 0.188 | No | df 5/tvc 1 | 15/15 | 1.79 | 0.82 (0.59–1.14) [6 mo] | 1.21 (0.77–1.90) [39 mo] | 0.80 (0.55–1.16) | 1.36 (0.73–2.56) |
| CheckMate 914 part B, nivolumab | Main | 0.83 (0.60–1.16) | 0.35 | 0.552 | No | df 5/tvc 1 | 15/15 | 1.99 | 0.74 (0.50–1.09) [6 mo] | 1.14 (0.57–2.28) [27 mo] | 0.83 (0.52–1.32) | 1.02 (0.40–2.60) |
| CheckMate 914 part B, nivolumab + ipilimumab | Alternative | 1.06 (0.73–1.54) | 0.36 | 0.551 | No | df 5/tvc 1 | 15/15 | 2.00 | 1.06 (0.71–1.60) [6 mo] | 1.09 (0.47–2.54) [27 mo] | 1.11 (0.67–1.84) | 1.08 (0.38–3.09) |
| S-TRAC | Main | 0.77 (0.60–0.98) | 2.40 | 0.121 | No | df 2/tvc 3 | 15/15 | 2.00 | 0.92 (0.67–1.27) [12 mo] | 0.68 (0.36–1.27) [72 mo] | 0.68 (0.51–0.91) | 1.15 (0.66–1.99) |
| ASSURE, sunitinib | Main | 1.01 (0.86–1.19) | 1.96 | 0.161 | No | df 5/tvc 1 | 15/15 | 0.15 | 0.99 (0.83–1.17) [12 mo] | 1.22 (0.94–1.58) [72 mo] | 1.00 (0.81–1.22) | 1.23 (0.86–1.77) |
| ASSURE, sorafenib | Alternative | 0.97 (0.83–1.15) | 3.79 | 0.052 | No | df 5/tvc 1 | 15/15 | 0.18 | 0.93 (0.78–1.10) [12 mo] | 1.31 (1.00–1.71) [72 mo] | 1.02 (0.82–1.26) | 1.17 (0.81–1.67) |
| PROTECT | Main | 0.93 (0.76–1.14) | 24.63 | <0.001 | Yes | df 5/tvc 2 | 14/15 | 1.40 | 0.94 (0.75–1.19) [12 mo] | 2.74 (1.55–4.84) [48 mo] | 0.78 (0.63–0.97) | 2.99 (1.70–5.27) |
| ATLAS | Main | 0.86 (0.66–1.13) | 1.93 | 0.165 | No | df 5/tvc 1 | 15/15 | 0.07 | 0.96 (0.69–1.33) [12 mo] | 1.07 (0.67–1.71) [36 mo] | 0.78 (0.57–1.06) | 1.58 (0.83–3.00) |
| SORCE, sorafenib 1 yr | Main | 0.93 (0.77–1.14) | 9.43 | 0.002 | Yes | df 5/tvc 3 | 14/15 | 4.14 | 0.89 (0.69–1.16) [12 mo] | 0.93 (0.60–1.42) [96 mo] | 0.84 (0.65–1.07) | 1.45 (0.92–2.28) |
| SORCE, sorafenib 3 yr | Alternative | 0.98 (0.81–1.20) | 3.21 | 0.073 | No | df 5/tvc 1 | 15/15 | 1.29 | 1.13 (0.87–1.46) [36 mo] | 1.26 (0.90–1.76) [96 mo] | 1.06 (0.84–1.34) | 1.29 (0.82–2.04) |
| E2810 (M1 NED after metastasectomy) | Main | 0.90 (0.60–1.33) | 4.17 | 0.041 | Yes | df 5/tvc 3 | 15/15 | 1.29 | 1.25 (0.69–2.27) [12 mo] | 0.81 (0.30–2.20) [48 mo] | 1.00 (0.58–1.71) | 1.29 (0.50–3.34) |
| LITESPARK-022, investigator (Fig 1A), standard reconstruction | Sensitivity | 0.70 (0.58–0.85) | 10.26 | 0.001 | Yes | df 4/tvc 1 | 15/15 | 1.13 | 0.76 (0.62–0.93) [12 mo] | 1.10 (0.77–1.57) [30 mo] | 0.72 (0.56–0.93) | 1.17 (0.78–1.75) |
| LITESPARK-022, investigator (Fig 1A), censoring-informed reconstruction | Sensitivity | 0.72 (0.59–0.87) | 12.55 | <0.001 | Yes | df 4/tvc 1 | 15/15 | 0.37 | 0.79 (0.64–0.96) [12 mo] | 1.12 (0.80–1.57) [30 mo] | 0.71 (0.55–0.92) | 1.24 (0.84–1.83) |
| PROSPER, standard reconstruction | Sensitivity | 0.96 (0.74–1.23) | 0.42 | 0.519 | No | df 3/tvc 1 | 7/15 | 1.47 | 0.90 (0.68–1.19) [12 mo] | 0.88 (0.66–1.19) [36 mo] | Not estimable | Not estimable |

| Comparison | Role | Whole-period Cox HR (95% CI) | Schoenfeld χ² | P | PH rejected (P < 0.05) | AIC-selected model (baseline df/tvc df) | Converged models | ΔAIC to second-best model | HR(t) at landmark, AIC-selected | HR(t) at maximum τ, AIC-selected | HR(t) at landmark, fixed df 3/tvc 2 | HR(t) at maximum τ, fixed df 3/tvc 2 |
|---|---|---|---|---|---|---|---|---|---|---|---|---|
| PROSPER, censoring-informed reconstruction | Sensitivity | 0.94 (0.73–1.20) | 0.54 | 0.463 | No | df 2/tvc 1 | 5/15 | 1.12 | 0.89 (0.67–1.18) [12 mo] | 0.86 (0.63–1.18) [36 mo] | Not estimable | Not estimable |

The whole-period Cox HR is the model used for the Schoenfeld test and is shown for reference only. “Converged models” = models out of 15 that met the convergence criteria (J11). For PROSPER, the fixed df 3/tvc 2 model could not be estimated for either reconstruction. For LITESPARK-022, the maximum feasible τ is 30 months, so “HR(t) at maximum τ” is the HR(t) at 30 months.

#### Supplementary Table S13. Window-matched post-treatment analysis (post hoc, D7)

Source: analysis output table of the window-matched increment; KEYNOTE-564 NEJM 2021 and Lancet Oncol 2022 cut-offs: separate output table (added post hoc on 14 October 2026; bootstrap seed set before each curve, K1).

| Comparison | Class | Role | Window (mo) | ΔRMST at window start (bootstrap 95% CI) | ΔRMST at window end (bootstrap 95% CI) | Increment, mo (bootstrap 95% CI) | Increment per year, mo/yr (bootstrap 95% CI) | Proportion of resamples with increment > 0 | KM DFS at start, exp vs ctrl (%); difference, pp (Greenwood 95% CI) | KM DFS at end, exp vs ctrl (%); difference, pp (Greenwood 95% CI) | Change in difference, pp (bootstrap 95% CI) |
|---|---|---|---|---|---|---|---|---|---|---|---|
| LITESPARK-022, BICR (Fig 1B) | HIF-2α | Main | 12→30 | 0.54 (0.31–0.76) | 1.65 (0.85–2.48) | 1.12 (0.48–1.78) | 0.74 (0.32–1.19) | 1.000 | 89.4 vs 81.6; 7.7 (4.5–11.0) | 73.2 vs 69.1; 4.0 (−1.0 to 9.1) | −3.7 (−8.1 to 0.8) |
| LITESPARK-022, investigator (Fig 1A), standard reconstruction | HIF-2α | Sensitivity | 12→30 | 0.45 (0.27–0.64) | 1.71 (0.94–2.50) | 1.26 (0.65–1.89) | 0.84 (0.43–1.26) | 1.000 | 92.1 vs 85.4; 6.7 (3.8–9.7) | 76.0 vs 69.0; 7.0 (1.8–12.1) | 0.2 (−4.3 to 4.8) |
| LITESPARK-022, investigator (Fig 1A), censoring-informed reconstruction | HIF-2α | Sensitivity | 12→30 | 0.45 (0.26–0.63) | 1.71 (0.94–2.45) | 1.27 (0.65–1.89) | 0.84 (0.44–1.26) | 1.000 | 92.0 vs 85.4; 6.7 (3.7–9.6) | 76.1 vs 68.9; 7.2 (2.3–12.1) | 0.6 (−4.0 to 5.0) |
| KEYNOTE-564 | ICI | Main | 12→30 | 0.72 (0.39–1.05) | 2.62 (1.40–3.81) | 1.90 (0.95–2.80) | 1.26 (0.63–1.87) | 1.000 | 85.6 vs 76.1; 9.5 (4.6–14.4) | 74.9 vs 64.9; 10.1 (4.3–15.8) | 0.5 (−3.4 to 4.7) |
| KEYNOTE-564, NEJM 2021 (cut-off Dec 2020) [added 14 Oct 2026; per-curve seed, K1] | ICI | Sensitivity | 12→30 | 0.71 (0.37–1.04) | 2.29 (1.03–3.53) | 1.57 (0.60–2.55) | 1.05 (0.40–1.70) | 1.000 | 85.7 vs 76.2; 9.6 (4.7–14.5) | 72.1 vs 66.1; 6.0 (−1.2 to 13.1) | −3.6 (−9.4 to 2.1) |
| KEYNOTE-564, Lancet Oncol 2022 (cut-off Jun 2021) [added 14 Oct 2026; per-curve seed, K1] | ICI | Sensitivity | 12→30 | 0.72 (0.35–1.05) | 2.58 (1.33–3.76) | 1.87 (0.91–2.79) | 1.25 (0.61–1.86) | 1.000 | 85.2 vs 75.9; 9.3 (4.4–14.3) | 75.1 vs 65.3; 9.9 (3.8–15.9) | 0.5 (−3.6 to 4.8) |
| IMmotion010 | ICI | Main | 12→30 | 0.39 (−0.05 to 0.82) | 0.79 (−0.73 to 2.29) | 0.40 (−0.72 to 1.52) | 0.27 (−0.48 to 1.01) | 0.748 | 77.4 vs 74.0; 3.4 (−2.8 to 9.5) | 62.3 vs 61.5; 0.8 (−6.3 to 7.8) | −2.6 (−7.6 to 2.3) |
| CheckMate 914 part A | ICI | Main | 6→24 | 0.03 (−0.07 to 0.12) | 0.44 (−0.55 to 1.44) | 0.42 (−0.53 to 1.37) | 0.28 (−0.35 to 0.91) | 0.799 | 89.0 vs 86.3; 2.7 (−1.9 to 7.3) | 75.8 vs 73.3; 2.5 (−3.8 to 8.8) | −0.2 (−5.2 to 4.7) |
| CheckMate 914 part B, nivolumab | ICI | Main | 6→24 | 0.09 (−0.03 to 0.22) | 0.85 (−0.41 to 2.09) | 0.76 (−0.41 to 1.89) | 0.51 (−0.27 to 1.26) | 0.898 | 89.7 vs 85.9; 3.8 (−1.9 to 9.5) | 73.9 vs 71.7; 2.3 (−6.1 to 10.7) | −1.5 (−8.2 to 5.3) |

| Comparison | Class | Role | Window (mo) | ΔRMST at window start (bootstrap 95% CI) | ΔRMST at window end (bootstrap 95% CI) | Increment, mo (bootstrap 95% CI) | Increment per year, mo/yr (bootstrap 95% CI) | Proportion of resamples with increment > 0 | KM DFS at start, exp vs ctrl (%); difference, pp (Greenwood 95% CI) | KM DFS at end, exp vs ctrl (%); difference, pp (Greenwood 95% CI) | Change in difference, pp (bootstrap 95% CI) |
|---|---|---|---|---|---|---|---|---|---|---|---|
| CheckMate 914 part B, nivolumab + ipilimumab | ICI | Alternative | 6→24 | −0.02 (−0.17 to 0.13) | −0.23 (−1.83 to 1.27) | −0.21 (−1.74 to 1.19) | −0.14 (−1.16 to 0.79) | 0.374 | 85.2 vs 85.9; −0.6 (−7.6 to 6.3) | 69.3 vs 69.5; −0.2 (−10.2 to 9.8) | 0.4 (−8.3 to 8.9) |
| PROSPER, standard reconstruction | ICI | Sensitivity | 12→30 | 0.00 (−0.60 to 0.58) | 0.44 (−1.21 to 2.01) | 0.43 (−0.68 to 1.56) | 0.29 (−0.45 to 1.04) | 0.758 | 76.3 vs 74.1; 2.1 (−4.1 to 8.3) | 66.6 vs 66.0; 0.6 (−6.8 to 7.9) | −1.6 (−6.6 to 3.5) |
| PROSPER, censoring-informed reconstruction | ICI | Sensitivity | 12→30 | 0.02 (−0.57 to 0.57) | 0.51 (−1.12 to 2.12) | 0.50 (−0.63 to 1.63) | 0.33 (−0.42 to 1.09) | 0.787 | 76.0 vs 73.6; 2.4 (−3.9 to 8.6) | 65.4 vs 64.3; 1.1 (−6.3 to 8.5) | −1.3 (−6.6 to 3.7) |
| S-TRAC | VEGFR-TKI | Main | 12→30 | 0.81 (0.35–1.26) | 1.92 (0.40–3.48) | 1.12 (−0.06 to 2.31) | 0.74 (−0.04 to 1.54) | 0.969 | 87.7 vs 77.4; 10.2 (4.1–16.4) | 68.8 vs 62.8; 6.0 (−2.0 to 14.0) | −4.2 (−10.8 to 2.3) |
| ASSURE, sunitinib | VEGFR-TKI | Main | 12→30 | 0.31 (0.00–0.61) | 0.49 (−0.56 to 1.53) | 0.17 (−0.63 to 0.99) | 0.11 (−0.42 to 0.66) | 0.664 | 84.0 vs 80.6; 3.3 (−0.9 to 7.5) | 66.1 vs 66.8; −0.7 (−6.0 to 4.6) | −4.0 (−8.1 to 0.3) |
| ASSURE, sorafenib | VEGFR-TKI | Alternative | 12→30 | 0.41 (0.09–0.72) | 0.94 (−0.10 to 2.02) | 0.53 (−0.28 to 1.39) | 0.35 (−0.19 to 0.93) | 0.888 | 85.1 vs 80.6; 4.4 (0.3–8.6) | 67.5 vs 66.8; 0.6 (−4.6 to 5.9) | −3.8 (−7.8 to 0.5) |
| PROTECT | VEGFR-TKI | Main | 12→30 | 0.66 (0.34–0.96) | 1.68 (0.51–2.83) | 1.02 (0.08–1.94) | 0.68 (0.06–1.29) | 0.984 | 87.8 vs 79.1; 8.7 (4.2–13.2) | 68.8 vs 65.1; 3.7 (−2.1 to 9.5) | −5.0 (−9.7 to −0.2) |
| ATLAS | VEGFR-TKI | Main | 12→30 | 0.31 (−0.15 to 0.77) | 1.06 (−0.46 to 2.59) | 0.75 (−0.39 to 1.88) | 0.50 (−0.26 to 1.25) | 0.909 | 80.9 vs 78.6; 2.3 (−3.7 to 8.3) | 70.5 vs 67.5; 3.0 (−4.5 to 10.6) | 0.7 (−4.7 to 6.2) |
| SORCE, sorafenib 1 yr | VEGFR-TKI | Main | 12→30 | 0.54 (0.28–0.82) | 1.43 (0.38–2.50) | 0.89 (0.06–1.74) | 0.59 (0.04–1.16) | 0.981 | 88.5 vs 81.8; 6.8 (2.3–11.2) | 75.4 vs 72.0; 3.4 (−2.0 to 8.8) | −3.4 (−7.4 to 0.3) |
| SORCE, sorafenib 3 yr | VEGFR-TKI | Alternative | 36→54 | 1.08 (−0.27 to 2.56) | 0.90 (−1.38 to 3.33) | −0.18 (−1.15 to 0.88) | −0.12 (−0.77 to 0.59) | 0.343 | 72.7 vs 71.8; 0.9 (−4.6 to 6.4) | 66.0 vs 68.8; −2.8 (−8.6 to 3.0) | −3.7 (−6.3 to −1.1) |
| E2810 (M1 NED after metastasectomy ) | VEGFR-TKI | Main | 12→30 | 1.61 (0.48–2.80) | 1.88 (−1.57 to 5.47) | 0.27 (−2.46 to 2.95) | 0.18 (−1.64 to 1.97) | 0.562 | 67.9 vs 56.5; 11.4 (−5.6 to 28.4) | 31.3 vs 27.7; 3.6 (−13.2 to 20.3) | −7.9 (−25.5 to 9.1) |

Window = main landmark to 18 months after the landmark. Increment per year = increment ÷ 1.5. KM DFS values in %, differences in pp (experimental minus control). Bootstrap: B = 2000, resampling within arms, percentile CIs; no resample failed. Class is used for grouping only; no pooling or test across trials.

### Supplementary Table S14. LITESPARK-022: central review versus investigator assessment

Sources: analysis output tables (feasibility, proportional hazards tests, primary RMST, RMST curves, piecewise HRs, Kaplan-Meier differences, window-matched increment, bootstrap increments and selected flexible parametric models).

| Measure | BICR (Fig 1B); standard reconstruction; main analysis | Investigator (Fig 1A); standard reconstruction (failed event-count criterion) | Investigator (Fig 1A); censoring-informed reconstruction (post hoc) |
|---|---|---|---|
| Reconstructed events, exp/ctrl | 199/250 | 173/238 | 184/243 |
| Whole-period Cox HR (95% CI) | 0.76 (0.63–0.91) | 0.70 (0.58–0.85) | 0.72 (0.59–0.87) |
| ΔRMST at 24 mo, mo (95% CI) | 1.34 (0.73–1.95) | 1.29 (0.75–1.84) | 1.29 (0.75–1.83) |
| RMST ratio at 24 mo (95% CI) | 1.067 (1.036–1.099) | 1.063 (1.036–1.090) | 1.062 (1.036–1.090) |
| ΔRMST at 30 mo (maximum feasible τ), mo (95% CI) | 1.65 (0.82–2.48) | 1.71 (0.96–2.47) | 1.71 (0.96–2.47) |
| On-treatment HR, landmark 12 mo (95% CI) | 0.55 (0.43–0.71) | 0.52 (0.39–0.69) | 0.52 (0.39–0.69) |
| Post-treatment HR, landmark 12 mo (95% CI) | 1.16 (0.87–1.54) | 0.93 (0.71–1.22) | 0.95 (0.73–1.24) |
| Post/on-treatment HR ratio, landmark 12 mo (95% CI) | 2.10 (1.44–3.08) | 1.80 (1.21–2.68) | 1.83 (1.24–2.70) |
| On-treatment HR, landmark 15 mo (95% CI) | 0.61 (0.48–0.76) | 0.57 (0.44–0.73) | 0.56 (0.43–0.72) |
| Post-treatment HR, landmark 15 mo (95% CI) | 1.24 (0.88–1.74) | 0.96 (0.71–1.31) | 1.01 (0.75–1.36) |

| Measure | BICR (Fig 1B); standard reconstruction; main analysis | Investigator (Fig 1A); standard reconstruction (failed event-count criterion) | Investigator (Fig 1A); censoring-informed reconstruction (post hoc) |
|---|---|---|---|
| Post/on-treatment HR ratio, landmark 15 mo (95% CI) | 2.05 (1.36–3.08) | 1.70 (1.14–2.53) | 1.80 (1.22–2.66) |
| KM DFS difference at 12 mo, pp (95% CI) | 7.7 (4.5–11.0) | 6.7 (3.8–9.7) | 6.7 (3.7–9.6) |
| KM DFS difference at 24 mo, pp (95% CI) | 4.9 (0.7–9.1) | 6.9 (2.8–11.0) | 7.0 (2.9–11.1) |
| KM DFS difference at 30 mo, pp (95% CI) | 4.0 (−1.0 to 9.1) | 7.0 (1.8–12.1) | 7.2 (2.3–12.1) |
| Change in KM DFS difference, 12→30 mo, pp (bootstrap 95% CI) (D7) | −3.7 (−8.1 to 0.8) | 0.2 (−4.3 to 4.8) | 0.6 (−4.0 to 5.0) |
| Post-treatment ΔRMST increment per year, 12→30 mo (bootstrap 95% CI) (D5) | 0.74 (0.32–1.19) | 0.84 (0.43–1.26) | 0.84 (0.44–1.26) |
| Schoenfeld test P | <0.001 | 0.001 | <0.001 |
| Flexible parametric HR(t) at 12 mo, AIC-selected [spec] | 0.75 (0.61–0.92) [df 4/tvc 2] | 0.76 (0.62–0.93) [df 4/tvc 1] | 0.79 (0.64–0.96) [df 4/tvc 1] |
| Flexible parametric HR(t) at 30 mo, AIC-selected | 2.03 (1.18–3.48) | 1.10 (0.77–1.57) | 1.12 (0.80–1.57) |
| Flexible parametric HR(t) at 30 mo, fixed df 3/tvc 2 | 1.68 (1.10–2.56) | 1.17 (0.78–1.75) | 1.24 (0.84–1.83) |

All three versions come from the same patients (NEJM 2026, first interim analysis); the control arm received pembrolizumab plus placebo. The investigator-assessed curve is the trial's primary DFS assessment; its standard reconstruction failed the event-count criterion and the censoring-informed reconstruction (post hoc) met all numerical criteria (Supplementary Table S5).

### Supplementary Table S15. SORCE, 3 years versus 1 year of sorafenib (exploratory)

Source: analysis output tables (primary RMST and piecewise HRs; SORCE 3 years vs 1 year comparison).

| Measure (sorafenib 3 yr, Fig 2A, vs sorafenib 1 yr, Fig 2B; reference = 1 yr) | Estimate |
|---|---|
| ΔRMST at 36 mo (95% CI); RMST ratio | −0.49 (−1.64 to 0.66); 0.984 (0.948–1.022) |
| ΔRMST at 24 mo (95% CI); RMST ratio | −0.47 (−1.11 to 0.18); 0.978 (0.949–1.009) |
| Piecewise Cox, landmark 36 mo (main): on-treatment HR; post-treatment HR; ratio | 1.05 (0.84–1.29); 1.08 (0.78–1.49); 1.03 (0.70–1.52) |
| Piecewise Cox, landmark 12 mo (sensitivity): on-treatment HR; post-treatment HR; ratio | 1.25 (0.92–1.71); 0.97 (0.78–1.20); 0.77 (0.53–1.13) |

Prespecified as exploratory and reported only in the supplement (SAP 9A item 1). The two sorafenib arms were reconstructed from separate figures (Fig 2A and Fig 2B); the actual median drug exposure was similar in the two arms, so this comparison is not used as a main argument.

### Supplementary Table S16. KEYNOTE-564: five-year analysis and earlier data cut-offs (SAP section 9, item 2)

Sources: analysis output tables (existing tables for the five-year curve; SAP-completion tables for the earlier cut-offs).

| Measure | Ann Oncol 2026, 5-year analysis (main analysis) | NEJM 2024 (cut-off Sep 2023) | Lancet Oncol 2022 (cut-off Jun 2021) | NEJM 2021 (cut-off Dec 2020) |
|---|---|---|---|---|
| Reconstructed events, exp/ctrl | 187/239 | 172/223 | 115/171 | 108/150 |
| Whole-period Cox HR (95% CI) | 0.72 (0.60–0.88) | 0.72 (0.59–0.88) | 0.63 (0.50–0.80) | 0.68 (0.53–0.88) |
| Primary τ / maximum feasible τ (mo) | 36 / 75 | 36 / 60 | 24 / 33 | 24 / 30 |
| ΔRMST at primary τ, mo (95% CI) | 3.21 (1.70–4.73) | 3.18 (1.66–4.70) | 1.98 (1.07–2.89) | 1.86 (0.95–2.77) |
| RMST ratio at primary τ (95% CI) | 1.120 (1.061–1.182) | 1.119 (1.060–1.181) | 1.104 (1.055–1.157) | 1.097 (1.048–1.149) |
| ΔRMST at 24 mo, mo (95% CI) | 1.98 (1.08–2.88) | 1.98 (1.08–2.88) | 1.98 (1.07–2.89) | 1.86 (0.95–2.77) |
| ΔRMST at maximum feasible τ, mo (95% CI) | 6.74 (3.11–10.37) | 5.31 (2.50–8.11) | 2.85 (1.47–4.23) | 2.29 (1.05–3.52) |
| Post-treatment increment per year, mo/yr (bootstrap 95% CI) | 1.15 (0.50–1.78) | 1.15 (0.53–1.80) | 1.22 (0.56–1.84) | 1.05 (0.40–1.70) |
| Window-matched increment per year, 12→30 mo, mo/yr (bootstrap 95% CI) (D7 method; post hoc) | 1.26 (0.63–1.87) | Not computed | 1.25 (0.61–1.86) | 1.05 (0.40–1.70) |
| KM DFS difference at window end (30 mo), pp (Greenwood 95% CI) | 10.1 (4.3–15.8) | Not computed | 9.9 (3.8–15.9) | 6.0 (−1.2 to 13.1) |
| Change in KM DFS difference, 12→30 mo, pp (bootstrap 95% CI) (D7 method; post hoc) | 0.5 (−3.4 to 4.7) | Not computed | 0.5 (−3.6 to 4.8) | −3.6 (−9.4 to 2.1) |
| Landmark (main landmark), mo | 12 | 12 | 12 | 12 |
| Events before main landmark, exp/ctrl | 69/118 | 69/118 | 71/119 | 68/117 |

| Measure | Ann Oncol 2026, 5-year analysis (main analysis) | NEJM 2024 (cut-off Sep 2023) | Lancet Oncol 2022 (cut-off Jun 2021) | NEJM 2021 (cut-off Dec 2020) |
|---|---|---|---|---|
| Events after main landmark, exp/ctrl | 118/121 | 103/105 | 44/52 | 40/33 |
| On-treatment HR, main landmark (95% CI) | 0.56 (0.42–0.75) | 0.56 (0.42–0.75) | 0.57 (0.43–0.77) | 0.56 (0.41–0.75) |
| Post-treatment HR, main landmark (95% CI) | 0.88 (0.68–1.13) | 0.89 (0.68–1.17) | 0.77 (0.51–1.15) | 1.12 (0.71–1.78) |
| Post/on-treatment HR ratio, main landmark (95% CI) | 1.57 (1.06–2.32) | 1.59 (1.06–2.38) | 1.35 (0.82–2.21) | 2.02 (1.16–3.49) |
| Landmark (sensitivity landmark), mo | 15 | 15 | 15 | 15 |
| Events before sensitivity landmark, exp/ctrl | 78/131 | 78/131 | 79/131 | 77/128 |
| Events after sensitivity landmark, exp/ctrl | 109/108 | 94/92 | 36/40 | 31/22 |
| On-treatment HR, sensitivity landmark (95% CI) | 0.57 (0.43–0.75) | 0.57 (0.43–0.75) | 0.57 (0.43–0.76) | 0.57 (0.43–0.76) |
| Post-treatment HR, sensitivity landmark (95% CI) | 0.91 (0.70–1.18) | 0.93 (0.69–1.23) | 0.82 (0.52–1.28) | 1.31 (0.76–2.26) |
| Post/on-treatment HR ratio, sensitivity landmark (95% CI) | 1.60 (1.09–2.35) | 1.63 (1.09–2.43) | 1.42 (0.84–2.42) | 2.27 (1.23–4.21) |
| KM DFS difference at 12 mo, pp (95% CI) | 9.5 (4.6–14.4) | 9.5 (4.6–14.4) | 9.3 (4.4–14.3) | 9.6 (4.7–14.5) |
| KM DFS difference at 24 mo, pp (95% CI) | 11.1 (5.5–16.7) | 11.1 (5.5–16.6) | 10.9 (5.2–16.5) | 9.0 (3.0–15.0) |
| KM DFS difference at 36 mo, pp (95% CI) | 9.8 (3.8–15.7) | 9.5 (3.6–15.4) | — | — |
| KM DFS difference at 48 mo, pp (95% CI) | 8.6 (2.4–14.8) | 8.6 (2.3–14.8) | — | — |
| KM DFS difference at 60 mo, pp (95% CI) | 8.7 (2.3–15.1) | 9.3 (2.2–16.4) | — | — |
| Schoenfeld test P | 0.011 | 0.009 | 0.049 | 0.002 |
| Flexible parametric HR(t) at main landmark, AIC-selected [spec] | 0.73 (0.60–0.89) [df 4/tvc 1] | 0.72 (0.58–0.89) [df 5/tvc 1] | 0.70 (0.53–0.91) [df 5/tvc 1] | 0.91 (0.67–1.24) [df 5/tvc 1] |
| Flexible parametric HR(t) at maximum feasible τ, AIC-selected | 1.03 (0.75–1.43) | 1.01 (0.74–1.38) | 0.88 (0.55–1.40) | 1.36 (0.80–2.31) |
| Flexible parametric HR(t) at maximum feasible τ, fixed df 3/tvc 2 | 0.99 (0.67–1.47) | 1.00 (0.67–1.49) | 1.12 (0.62–2.03) | 1.48 (0.75–2.92) |

All four curves are investigator-assessed DFS in the same ITT population; earlier cut-offs have shorter follow-up and therefore smaller maximum feasible τ, so per-year increments are averaged over different post-treatment windows. “—” = time point not feasible under the 10% rule. Window-matched rows (D7 method, landmark to landmark + 18 months; post hoc): five-year curve from the window-matched increment table; NEJM 2021 and Lancet Oncol 2022 cut-offs from the separate post hoc output table (added 14 October 2026); “Not computed” = the window-matched analysis was not run for the NEJM 2024 curve. Events before and after each landmark (experimental/control) are those of the piecewise Cox models (SAP-completion run for the earlier cut-offs). Substitution of each earlier curve in the ICI pooling is shown in Table S6.

#### Supplementary Table S17. PROSPER: main-text curve and curve with Day-1 events censored (SAP section 9, item 3)

Sources: analysis output tables (existing tables for Fig 2; SAP-completion tables for Supplemental Fig 3).

| Measure | Fig 2, standard reconstruction (failed QC; D4-A) | Fig 2, censoring-informed reconstruction (post hoc method; D4-C) | Supplemental Fig 3, Day-1 events censored (passed QC; SAP 9 item 3) |
|---|---|---|---|
| Reconstructed events, exp/ctrl | 118/124 | 124/133 | 87/108 |
| Whole-period Cox HR (95% CI) | 0.96 (0.74–1.23) | 0.94 (0.73–1.20) | 0.79 (0.59–1.05) |
| Primary τ / maximum feasible τ (mo) | 36 / 36 | 36 / 36 | 36 / 36 |
| ΔRMST at primary τ, mo (95% CI) | 0.41 (−1.65 to 2.48) | 0.50 (−1.56 to 2.56) | 1.59 (−0.28 to 3.46) |
| RMST ratio at primary τ (95% CI) | 1.016 (0.939–1.099) | 1.019 (0.942–1.103) | 1.057 (0.990–1.129) |
| ΔRMST at 24 mo, mo (95% CI) | 0.32 (−0.97 to 1.61) | 0.37 (−0.92 to 1.66) | 1.12 (0.02–2.22) |
| ΔRMST at maximum feasible τ, mo (95% CI) | 0.41 (−1.65 to 2.48) | 0.50 (−1.56 to 2.56) | 1.59 (−0.28 to 3.46) |
| Post-treatment increment per year, mo/yr (bootstrap 95% CI) | 0.20 (−0.55 to 0.97) | 0.24 (−0.53 to 0.99) | 0.58 (−0.21 to 1.41) |
| Landmark (main landmark), mo | 12 | 12 | 12 |
| On-treatment HR, main landmark (95% CI) | 0.93 (0.70–1.24) | 0.92 (0.69–1.23) | 0.71 (0.50–1.02) |
| Post-treatment HR, main landmark (95% CI) | 1.05 (0.62–1.77) | 0.99 (0.61–1.59) | 0.94 (0.59–1.50) |
| Post/on-treatment HR ratio, main landmark (95% CI) | 1.13 (0.62–2.05) | 1.07 (0.62–1.87) | 1.32 (0.73–2.38) |
| Landmark (sensitivity landmark), mo | 10 | 10 | 10 |
| On-treatment HR, sensitivity landmark (95% CI) | 0.94 (0.70–1.26) | 0.94 (0.70–1.26) | 0.72 (0.50–1.03) |
| Post-treatment HR, sensitivity landmark (95% CI) | 1.01 (0.61–1.67) | 0.93 (0.59–1.47) | 0.92 (0.58–1.45) |

| Measure | Fig 2, standard reconstruction (failed QC; D4-A) | Fig 2, censoring-informed reconstruction (post hoc method; D4-C) | Supplemental Fig 3, Day-1 events censored (passed QC; SAP 9 item 3) |
|---|---|---|---|
| Post/on-treatment HR ratio, sensitivity landmark (95% CI) | 1.07 (0.60–1.92) | 1.00 (0.58–1.71) | 1.28 (0.71–2.29) |
| KM DFS difference at 12 mo, pp (95% CI) | 2.1 (−4.1 to 8.3) | 2.4 (−3.9 to 8.6) | 4.9 (−0.8 to 10.6) |
| KM DFS difference at 24 mo, pp (95% CI) | 3.4 (−3.6 to 10.3) | 3.6 (−3.4 to 10.6) | 6.8 (−0.2 to 13.7) |
| KM DFS difference at 36 mo, pp (95% CI) | −1.6 (−9.5 to 6.3) | −1.7 (−9.5 to 6.2) | 0.6 (−7.5 to 8.8) |
| KM DFS difference at 48 mo, pp (95% CI) | — | — | — |
| KM DFS difference at 60 mo, pp (95% CI) | — | — | — |
| Schoenfeld test P | 0.519 | 0.463 | 0.047 |
| Flexible parametric HR(t) at main landmark, AIC-selected [spec] | 0.90 (0.68–1.19) [df 3/tvc 1] | 0.89 (0.67–1.18) [df 2/tvc 1] | 0.88 (0.63–1.21) [df 5/tvc 1] |
| Flexible parametric HR(t) at maximum feasible τ, AIC-selected | 0.88 (0.66–1.19) | 0.86 (0.63–1.18) | 1.07 (0.62–1.85) |
| Flexible parametric HR(t) at maximum feasible τ, fixed df 3/tvc 2 | Not estimable | Not estimable | 0.96 (0.47–1.96) |

Endpoint RFS; control arm observation. The Day-1-censored curve passed the quality checks and is therefore reported separately, as prespecified; the standard reconstruction of Fig 2 failed the event-count criterion (D4-A) and the censoring-informed reconstruction is a method outside the plan (D4-C).

### Supplementary Table S18. ASSURE, PROTECT, ATLAS and SORCE: subsets, populations and assessment methods (SAP section 9, items 4–7)

Sources: existing tables for the main and alternative curves; SAP-completion tables for the added curves. † Accepted after adjudication (source inconsistency; see Table S1, D8-c and D8-d).

##### A. RMST and post-treatment increment

| Comparison | n, exp/ctrl | Reconstructed events, exp/ctrl | Primary τ (mo) | ΔRMST at primary τ, mo (95% CI) | RMST ratio (95% CI) | ΔRMST at 24 mo (95% CI) | Maximum feasible τ (mo) | Increment per year, mo/yr (bootstrap 95% CI) | Proportion of resamples with increment > 0 |
|---|---|---|---|---|---|---|---|---|---|
| ASSURE, sunitinib (main or alternative analysis) | 647/647 | 284/287 | 36 | 0.44 (−0.92 to 1.80) | 1.016 (0.967–1.067) | 0.54 (−0.27 to 1.35) | 72 | −0.12 (−0.69 to 0.48) | 0.333 |
| ASSURE clear cell, sunitinib † | 542/540 | 240/240 | 36 | 0.88 (−0.60 to 2.36) | 1.032 (0.979–1.088) | 0.87 (−0.01 to 1.75) | 72 | −0.08 (−0.72 to 0.57) | 0.407 |
| ASSURE, sorafenib (main or alternative analysis) | 649/647 | 285/287 | 36 | 1.02 (−0.32 to 2.35) | 1.037 (0.989–1.087) | 0.86 (0.07–1.65) | 72 | 0.00 (−0.58 to 0.59) | 0.496 |
| ASSURE clear cell, sorafenib † | 540/540 | 244/240 | 36 | 1.46 (0.01–2.91) | 1.053 (1.000–1.109) | 1.20 (0.34–2.06) | 72 | 0.02 (−0.62 to 0.65) | 0.519 |
| PROTECT (main or alternative analysis) | 571/564 | 190/198 | 36 | 1.91 (0.44–3.37) | 1.070 (1.016–1.128) | 1.45 (0.60–2.30) | 48 | 0.43 (−0.21 to 1.07) | 0.891 |
| PROTECT, ITT 800 mg † | 198/205 | 70/102 | 36 | 3.12 (0.68–5.56) | 1.122 (1.025–1.228) | 1.86 (0.43–3.30) | 60 | 1.29 (0.19–2.35) | 0.992 |
| PROTECT, ITT all randomised † | 769/769 | 270/305 | 36 | 2.27 (1.01–3.53) | 1.085 (1.037–1.136) | 1.59 (0.85–2.33) | 54 | 0.59 (0.01–1.14) | 0.978 |
| ATLAS (main or alternative analysis) | 363/361 | 98/108 | 36 | 1.25 (−0.66 to 3.16) | 1.045 (0.977–1.119) | 0.83 (−0.32 to 1.97) | 36 | 0.47 (−0.30 to 1.23) | 0.890 |
| ATLAS highest risk, IRC | 209/200 | 64/74 | 36 | 2.74 (0.05–5.43) | 1.109 (1.001–1.229) | 1.68 (0.05–3.31) | 36 | 1.07 (0.01–2.17) | 0.976 |
| ATLAS highest risk, investigator | 209/200 | 70/89 | 36 | 4.45 (1.81–7.10) | 1.192 (1.071–1.325) | 3.00 (1.44–4.55) | 36 | 1.61 (0.53–2.71) | 0.997 |
| ATLAS ITT, investigator | 363/361 | 103/124 | 36 | 2.54 (0.67–4.41) | 1.096 (1.024–1.174) | 1.84 (0.76–2.93) | 36 | 0.87 (0.12–1.68) | 0.987 |
| SORCE, sorafenib 1 yr (main or alternative analysis) | 642/430 | 242/166 | 36 | 1.57 (0.20–2.94) | 1.054 (1.006–1.104) | 1.21 (0.41–2.00) | 96 | 0.23 (−0.40 to 0.86) | 0.762 |
| SORCE Leibovich | 300/206 | 160/110 | 36 | 2.78 (0.46–5.10) | 1.112 (1.016–1.217) | 2.30 (0.92–3.69) | 96 | 0.33 (−0.63 to 1.34) | 0.750 |

| Comparison | n, exp/ctrl | Reconstructed events, exp/ctrl | Primary τ (mo) | ΔRMST at primary τ, mo (95% CI) | RMST ratio (95% CI) | ΔRMST at 24 mo (95% CI) | Maximum feasible τ (mo) | Increment per year, mo/yr (bootstrap 95% CI) | Proportion of resamples with increment > 0 |
|---|---|---|---|---|---|---|---|---|---|
| high risk, sorafenib 1 yr | | | | | | | | | |
| SORCE clear cell, sorafenib 1 yr | 534/361 | 201/140 | 36 | 1.82 (0.34–3.29) | 1.062 (1.011–1.116) | 1.34 (0.49–2.19) | 96 | 0.32 (−0.35 to 0.96) | 0.826 |
| SORCE, sorafenib 3 yr (main or alternative analysis) | 639/430 | 244/166 | 36 | 1.08 (−0.33 to 2.48) | 1.037 (0.989–1.088) | 0.74 (−0.08 to 1.56) | 96 | −0.13 (−0.80 to 0.58) | 0.332 |
| SORCE Leibovich high risk, sorafenib 3 yr | 295/206 | 162/110 | 36 | 1.36 (−1.03 to 3.75) | 1.054 (0.960–1.158) | 1.17 (−0.26 to 2.61) | 96 | −0.43 (−1.45 to 0.62) | 0.201 |
| SORCE clear cell, sorafenib 3 yr | 550/361 | 210/140 | 36 | 1.07 (−0.45 to 2.59) | 1.037 (0.985–1.092) | 0.74 (−0.14 to 1.63) | 96 | −0.15 (−0.90 to 0.58) | 0.347 |

#### B. Piecewise Cox models

| Comparison | Landmark (mo) | Events before landmark, exp/ctrl | Events after landmark, exp/ctrl | On-treatment HR (95% CI) | Post-treatment HR (95% CI) | Post/on-treatment HR ratio (95% CI) | P (interaction) |
|---|---|---|---|---|---|---|---|
| ASSURE, sunitinib (main or alternative analysis) | 12 (main) | 100/123 | 184/164 | 0.80 (0.62–1.05) | 1.17 (0.95–1.45) | 1.45 (1.04–2.04) | 0.030 |
| ASSURE, sunitinib (main or alternative analysis) | 15 (sensitivity) | 127/148 | 157/139 | 0.85 (0.67–1.08) | 1.19 (0.94–1.49) | 1.40 (1.00–1.94) | 0.047 |
| ASSURE clear cell, sunitinib † | 12 (main) | 77/104 | 163/136 | 0.72 (0.54–0.97) | 1.24 (0.99–1.56) | 1.72 (1.19–2.50) | 0.004 |
| ASSURE clear cell, sunitinib † | 15 (sensitivity) | 102/125 | 138/115 | 0.79 (0.61–1.03) | 1.26 (0.98–1.61) | 1.58 (1.10–2.27) | 0.013 |
| ASSURE, sorafenib (main or alternative analysis) | 12 (main) | 94/123 | 191/164 | 0.74 (0.57–0.97) | 1.15 (0.94–1.42) | 1.56 (1.11–2.19) | 0.011 |
| ASSURE, sorafenib (main or alternative analysis) | 15 (sensitivity) | 118/148 | 167/139 | 0.77 (0.61–0.98) | 1.20 (0.95–1.50) | 1.55 (1.11–2.16) | 0.009 |
| ASSURE clear cell, sorafenib † | 12 (main) | 70/104 | 174/136 | 0.64 (0.47–0.87) | 1.23 (0.99–1.55) | 1.92 (1.32–2.80) | <0.001 |
| ASSURE clear cell, sorafenib † | 15 (sensitivity) | 92/125 | 152/115 | 0.70 (0.53–0.91) | 1.28 (1.01–1.64) | 1.84 (1.28–2.64) | 0.001 |
| PROTECT (main or alternative analysis) | 12 (main) | 63/111 | 127/87 | 0.55 (0.40–0.75) | 1.42 (1.08–1.86) | 2.59 (1.71–3.91) | <0.001 |
| PROTECT (main or alternative analysis) | 15 (sensitivity) | 88/133 | 102/65 | 0.63 (0.48–0.83) | 1.54 (1.13–2.11) | 2.43 (1.61–3.67) | <0.001 |
| PROTECT, ITT 800 mg † | 12 (main) | 27/51 | 43/51 | 0.52 (0.33–0.83) | 0.77 (0.51–1.16) | 1.49 (0.80–2.76) | 0.209 |
| PROTECT, ITT 800 mg † | 15 (sensitivity) | 36/58 | 34/44 | 0.60 (0.40–0.91) | 0.70 (0.45–1.10) | 1.17 (0.63–2.15) | 0.619 |
| PROTECT, ITT all randomised † | 12 (main) | 114/168 | 156/137 | 0.64 (0.51–0.81) | 1.07 (0.85–1.34) | 1.66 (1.19–2.31) | 0.003 |
| PROTECT, ITT all randomised † | 15 (sensitivity) | 130/192 | 140/113 | 0.64 (0.51–0.80) | 1.17 (0.91–1.49) | 1.83 (1.31–2.55) | <0.001 |
| ATLAS (main or alternative analysis) | 12 (main) | 66/74 | 32/34 | 0.86 (0.61–1.19) | 0.88 (0.54–1.42) | 1.03 (0.57–1.85) | 0.930 |
| ATLAS (main or alternative analysis) | 36 (sensitivity) | 93/104 | 5/4 | 0.85 (0.64–1.12) | 1.37 (0.37–5.13) | 1.62 (0.42–6.26) | 0.483 |
| ATLAS highest risk, IRC | 12 (main) | 42/52 | 22/22 | 0.72 (0.48–1.08) | 0.76 (0.42–1.37) | 1.05 (0.51–2.16) | 0.884 |
| ATLAS highest risk, IRC | 36 (sensitivity) | 62/74 | 2/0 | 0.71 (0.51–0.99) | Not estimable (0 events in one arm and period) | Not estimable (0 events in one arm and period) | NA |
| ATLAS highest risk, investigator | 12 (main) | 35/62 | 35/27 | 0.48 (0.32–0.72) | 0.99 (0.60–1.64) | 2.07 (1.08–3.98) | 0.028 |
| ATLAS highest risk, investigator | 36 (sensitivity) | 66/88 | 4/1 | 0.61 (0.45–0.85) | 2.63 (0.29–23.58) | 4.28 (0.47–39.29) | 0.199 |
| ATLAS ITT, investigator | 12 (main) | 51/81 | 52/43 | 0.59 (0.41–0.83) | 1.10 (0.73–1.64) | 1.87 (1.09–3.19) | 0.022 |
| ATLAS ITT, investigator | 36 (sensitivity) | 99/120 | 4/4 | 0.75 (0.57–0.98) | 1.42 (0.35–5.72) | 1.90 (0.46–7.84) | 0.376 |
| SORCE, sorafenib 1 yr | 12 (main) | 73/78 | 169/88 | 0.59 (0.43–0.82) | 1.24 (0.96–1.61) | 2.09 (1.39–3.15) | <0.001 |

| Comparison | Landmark (mo) | Events before landmark, exp/ctrl | Events after landmark, exp/ctrl | On-treatment HR (95% CI) | Post-treatment HR (95% CI) | Post/on-treatment HR ratio (95% CI) | P (interaction) |
|---|---|---|---|---|---|---|---|
| (main or alternative analysis) | | | | | | | |
| SORCE, sorafenib 1 yr (main or alternative analysis) | 15 (sensitivity) | 86/88 | 156/78 | 0.62 (0.46–0.83) | 1.30 (0.99–1.70) | 2.10 (1.40–3.14) | <0.001 |
| SORCE Leibovich high risk, sorafenib 1 yr | 12 (main) | 54/67 | 106/43 | 0.50 (0.35–0.72) | 1.56 (1.10–2.23) | 3.11 (1.88–5.15) | <0.001 |
| SORCE Leibovich high risk, sorafenib 1 yr | 15 (sensitivity) | 62/73 | 98/37 | 0.52 (0.37–0.74) | 1.70 (1.16–2.48) | 3.23 (1.95–5.37) | <0.001 |
| SORCE clear cell, sorafenib 1 yr | 12 (main) | 54/65 | 147/75 | 0.53 (0.37–0.76) | 1.27 (0.96–1.68) | 2.39 (1.52–3.78) | <0.001 |
| SORCE clear cell, sorafenib 1 yr | 15 (sensitivity) | 66/74 | 135/66 | 0.57 (0.41–0.79) | 1.33 (0.99–1.79) | 2.35 (1.51–3.66) | <0.001 |
| SORCE, sorafenib 3 yr (main or alternative analysis) | 36 (main) | 171/120 | 73/46 | 0.93 (0.74–1.17) | 1.13 (0.78–1.64) | 1.22 (0.79–1.89) | 0.370 |
| SORCE, sorafenib 3 yr (main or alternative analysis) | 12 (sensitivity) | 89/78 | 155/88 | 0.74 (0.55–1.01) | 1.20 (0.92–1.56) | 1.61 (1.08–2.41) | 0.020 |
| SORCE Leibovich high risk, sorafenib 3 yr | 36 (main) | 124/86 | 38/24 | 0.95 (0.72–1.25) | 1.22 (0.73–2.03) | 1.28 (0.72–2.29) | 0.403 |
| SORCE Leibovich high risk, sorafenib 3 yr | 12 (sensitivity) | 71/66 | 91/44 | 0.71 (0.51–0.99) | 1.47 (1.02–2.11) | 2.08 (1.27–3.40) | 0.004 |
| SORCE clear cell, sorafenib 3 yr | 36 (main) | 148/101 | 62/39 | 0.93 (0.72–1.20) | 1.13 (0.76–1.69) | 1.21 (0.76–1.95) | 0.425 |
| SORCE clear cell, sorafenib 3 yr | 12 (sensitivity) | 72/65 | 138/75 | 0.71 (0.51–0.99) | 1.23 (0.93–1.63) | 1.74 (1.12–2.70) | 0.013 |

#### C. Proportional hazards and flexible parametric HR(t)

| Comparison | Whole-period Cox HR (95% CI) | Schoenfeld P | AIC-selected model | Converged models | HR(t) at landmark, AIC-selected | HR(t) at maximum τ, AIC-selected | HR(t) at maximum τ, fixed df 3/tvc 2 |
|---|---|---|---|---|---|---|---|
| ASSURE, sunitinib (main or alternative analysis) | 1.01 (0.86–1.19) | 0.161 | df 5/tvc 1 | 15/15 | 0.99 (0.83–1.17) [12 mo] | 1.22 (0.94–1.58) [72 mo] | 1.23 (0.86–1.77) |
| ASSURE clear cell, sunitinib † | 1.01 (0.85–1.21) | 0.018 | df 5/tvc 1 | 15/15 | 0.97 (0.81–1.17) [12 mo] | 1.29 (0.98–1.69) [72 mo] | 1.46 (1.03–2.06) |
| ASSURE, sorafenib (main or alternative analysis) | 0.97 (0.83–1.15) | 0.052 | df 5/tvc 1 | 15/15 | 0.93 (0.78–1.10) [12 mo] | 1.31 (1.00–1.71) [72 mo] | 1.17 (0.81–1.67) |
| ASSURE clear cell, sorafenib † | 0.98 (0.82–1.17) | 0.005 | df 4/tvc 3 | 15/15 | 1.08 (0.84–1.39) [12 mo] | 1.03 (0.65–1.63) [72 mo] | 1.48 (1.02–2.14) |
| PROTECT (main or alternative analysis) | 0.93 (0.76–1.14) | <0.001 | df 5/tvc 2 | 14/15 | 0.94 (0.75–1.19) [12 mo] | 2.74 (1.55–4.84) [48 mo] | 2.99 (1.70–5.27) |
| PROTECT, ITT 800 mg † | 0.65 (0.48–0.88) | 0.412 | df 5/tvc 1 | 11/15 | 0.64 (0.47–0.87) [12 mo] | 0.68 (0.34–1.36) [60 mo] | Not estimable |
| PROTECT, ITT all randomised † | 0.83 (0.71–0.98) | <0.001 | df 5/tvc 3 | 13/15 | 1.02 (0.76–1.36) [12 mo] | 1.29 (0.77–2.18) [54 mo] | 1.64 (1.09–2.49) |
| ATLAS (main or alternative analysis) | 0.86 (0.66–1.13) | 0.165 | df 5/tvc 1 | 15/15 | 0.96 (0.69–1.33) [12 mo] | 1.07 (0.67–1.71) [36 mo] | 1.58 (0.83–3.00) |
| ATLAS highest risk, IRC | 0.73 (0.52–1.02) | 0.320 | df 5/tvc 3 | 15/15 | 0.86 (0.53–1.41) [12 mo] | 1.09 (0.45–2.65) [36 mo] | 1.78 (0.80–3.95) |
| ATLAS highest risk, investigator | 0.64 (0.47–0.87) | 0.002 | df 5/tvc 1 | 15/15 | 0.81 (0.56–1.16) [12 mo] | 1.92 (0.79–4.65) [36 mo] | 1.34 (0.54–3.33) |
| ATLAS ITT, investigator | 0.76 (0.59–0.99) | 0.001 | df 5/tvc 1 | 13/15 | 1.06 (0.74–1.51) [12 mo] | 1.83 (1.01–3.33) [36 mo] | 1.26 (0.63–2.51) |
| SORCE, sorafenib 1 yr (main or alternative analysis) | 0.93 (0.77–1.14) | 0.002 | df 5/tvc 3 | 14/15 | 0.89 (0.69–1.16) [12 mo] | 0.93 (0.60–1.42) [96 mo] | 1.45 (0.92–2.28) |
| SORCE Leibovich high risk, sorafenib 1 yr | 0.91 (0.72–1.16) | <0.001 | df 5/tvc 3 | 13/15 | 0.95 (0.67–1.35) [12 mo] | 1.14 (0.64–2.04) [96 mo] | 1.97 (1.06–3.64) |
| SORCE clear cell, sorafenib 1 yr | 0.93 (0.75–1.15) | 0.001 | df 5/tvc 3 | 14/15 | 0.80 (0.60–1.06) [12 mo] | 1.03 (0.66–1.61) [96 mo] | 1.74 (1.06–2.85) |

| Comparison | Whole-period Cox HR (95% CI) | Schoenfeld P | AIC-selected model | Converged models | HR(t) at landmark, AIC-selected | HR(t) at maximum τ, AIC-selected | HR(t) at maximum τ, fixed df 3/tvc 2 |
|---|---|---|---|---|---|---|---|
| SORCE, sorafenib 3 yr (main or alternative analysis) | 0.98 (0.81–1.20) | 0.073 | df 5/tvc 1 | 15/15 | 1.13 (0.87–1.46) [36 mo] | 1.26 (0.90–1.76) [96 mo] | 1.29 (0.82–2.04) |
| SORCE Leibovich high risk, sorafenib 3 yr | 1.01 (0.79–1.28) | 0.021 | df 5/tvc 1 | 14/15 | 1.40 (0.94–2.09) [36 mo] | 1.49 (0.99–2.26) [96 mo] | 1.52 (0.83–2.79) |
| SORCE clear cell, sorafenib 3 yr | 0.99 (0.80–1.22) | 0.104 | df 5/tvc 2 | 14/15 | 1.14 (0.86–1.53) [36 mo] | 1.33 (0.88–2.02) [96 mo] | 1.39 (0.84–2.30) |

#### D. Kaplan-Meier DFS (%) in the experimental vs control arm; difference, pp (Greenwood 95% CI)

| Comparison | 12 mo | 24 mo | 36 mo | 48 mo | 60 mo |
|---|---|---|---|---|---|
| ASSURE, sunitinib (main or alternative analysis) | 84.0 vs 80.6; 3.3 (−0.9 to 7.5) | 70.1 vs 70.1; 0.0 (−5.2 to 5.1) | 62.7 vs 63.7; −1.0 (−6.5 to 4.4) | 57.6 vs 59.8; −2.2 (−7.9 to 3.4) | 54.1 vs 56.4; −2.3 (−8.2 to 3.5) |
| ASSURE clear cell, sunitinib † | 85.2 vs 80.4; 4.8 (0.2–9.4) | 70.8 vs 69.8; 1.0 (−4.7 to 6.6) | 62.5 vs 62.9; −0.4 (−6.4 to 5.6) | 57.5 vs 59.6; −2.0 (−8.2 to 4.2) | 53.4 vs 55.9; −2.6 (−9.0 to 3.9) |
| ASSURE, sorafenib (main or alternative analysis) | 85.1 vs 80.6; 4.4 (0.3–8.6) | 72.3 vs 70.1; 2.3 (−2.8 to 7.3) | 65.1 vs 63.7; 1.5 (−3.9 to 6.9) | 58.2 vs 59.8; −1.7 (−7.3 to 3.9) | 53.7 vs 56.4; −2.6 (−8.5 to 3.2) |
| ASSURE clear cell, sorafenib † | 86.6 vs 80.4; 6.3 (1.8–10.7) | 73.0 vs 69.8; 3.2 (−2.3 to 8.7) | 64.8 vs 62.9; 1.9 (−4.0 to 7.9) | 57.4 vs 59.6; −2.2 (−8.4 to 4.0) | 52.6 vs 55.9; −3.3 (−9.7 to 3.1) |
| PROTECT (main or alternative analysis) | 87.8 vs 79.1; 8.7 (4.2–13.2) | 72.6 vs 67.9; 4.7 (−0.9 to 10.3) | 65.9 vs 64.1; 1.8 (−4.1 to 7.7) | 61.0 vs 61.0; 0.1 (−6.3 to 6.4) | — |
| PROTECT, ITT 800 mg † | 85.7 vs 74.8; 10.9 (3.1–18.7) | 71.9 vs 63.2; 8.6 (−0.7 to 18.0) | 65.1 vs 55.2; 10.0 (0.1–19.8) | 62.8 vs 50.1; 12.7 (2.7–22.7) | 60.6 vs 44.7; 15.9 (5.1–26.7) |
| PROTECT, ITT all randomised † | 84.0 vs 77.2; 6.8 (2.7–10.9) | 71.7 vs 67.0; 4.7 (−0.1 to 9.5) | 65.0 vs 60.7; 4.3 (−0.8 to 9.3) | 60.8 vs 57.0; 3.8 (−1.6 to 9.2) | — |
| ATLAS (main or alternative analysis) | 80.9 vs 78.6; 2.3 (−3.7 to 8.3) | 75.4 vs 70.2; 5.2 (−1.7 to 12.2) | 69.1 vs 66.1; 3.1 (−4.8 to 10.9) | — | — |
| ATLAS highest risk, IRC | 79.0 vs 72.6; 6.4 (−2.1 to 14.9) | 72.4 vs 62.2; 10.2 (0.4–20.0) | 62.2 vs 52.4; 9.8 (−2.8 to 22.3) | — | — |
| ATLAS highest risk, investigator | 82.4 vs 67.9; 14.5 (6.1–23.0) | 67.7 vs 54.2; 13.4 (3.3–23.5) | 60.2 vs 49.4; 10.8 (−0.8 to 22.5) | — | — |
| ATLAS ITT, investigator | 85.1 vs 76.7; 8.5 (2.6–14.3) | 73.4 vs 65.0; 8.4 (1.2–15.7) | 65.2 vs 61.0; 4.2 (−4.2 to 12.7) | — | — |
| SORCE, sorafenib 1 yr (main or alternative analysis) | 88.5 vs 81.8; 6.8 (2.3–11.2) | 79.5 vs 74.7; 4.8 (−0.4 to 10.0) | 73.3 vs 71.8; 1.5 (−4.0 to 7.0) | 70.5 vs 69.8; 0.7 (−5.0 to 6.3) | 67.3 vs 66.7; 0.6 (−5.2 to 6.5) |
| SORCE Leibovich high risk, sorafenib 1 yr | 81.9 vs 67.5; 14.4 (6.6–22.1) | 68.5 vs 59.6; 8.9 (0.4–17.4) | 58.9 vs 57.7; 1.2 (−7.6 to 10.0) | 56.4 vs 55.7; 0.7 (−8.1 to 9.6) | 52.7 vs 51.9; 0.8 (−8.2 to 9.8) |
| SORCE clear cell, sorafenib 1 yr | 89.8 vs 81.9; 7.9 (3.1–12.6) | 80.2 vs 74.9; 5.4 (−0.3 to 11.0) | 74.6 vs 71.7; 2.8 (−3.1 to 8.8) | 71.6 vs 69.7; 1.9 (−4.3 to 8.0) | 68.1 vs 66.9; 1.2 (−5.2 to 7.6) |
| SORCE, sorafenib 3 yr (main or alternative analysis) | 86.0 vs 81.8; 4.2 (−0.4 to 8.7) | 78.3 vs 74.7; 3.6 (−1.6 to 8.9) | 72.7 vs 71.8; 0.9 (−4.6 to 6.4) | 68.0 vs 69.8; −1.8 (−7.5 to 3.9) | 64.5 vs 66.7; −2.1 (−8.0 to 3.8) |
| SORCE Leibovich high risk, sorafenib 3 yr | 75.8 vs 67.9; 7.9 (−0.2 to 15.9) | 64.4 vs 60.1; 4.3 (−4.3 to 13.0) | 57.4 vs 58.1; −0.7 (−9.5 to 8.1) | 51.6 vs 56.1; −4.4 (−13.4 to 4.5) | 48.1 vs 51.7; −3.6 (−12.6 to 5.4) |
| SORCE clear cell, sorafenib 3 yr | 86.8 vs 81.9; 4.9 (0.0–9.8) | 78.3 vs 74.9; 3.4 (−2.3 to 9.1) | 72.5 vs 71.7; 0.8 (−5.2 to 6.8) | 67.8 vs 69.7; −1.8 (−8.0 to 4.4) | 64.2 vs 66.9; −2.6 (−9.1 to 3.8) |

Increment CIs are bootstrap percentile intervals (B = 2000; K1); other CIs are asymptotic. “—” = time point not feasible under the 10% rule. For ATLAS highest risk (IRC) no placebo event occurred after 36 months, so the piecewise model at the 36-month sensitivity landmark is not estimable.

### Supplementary Figures

All figures were produced by the analysis code or the reconstruction code. In version 2, Figures S1–S9 were redrawn from the same data with the same plotting functions, with readable panel labels instead of internal identifiers; the version 1 figures are unchanged.

**Figure S1.** ΔRMST(τ) curves for the alternative comparisons sharing a control arm (CheckMate 914 part B nivolumab plus ipilimumab, ASSURE sorafenib, SORCE 3 years) and for the sensitivity curves (LITESPARK-022 investigator-assessed DFS, standard and censoring-informed reconstructions; PROSPER RFS, standard and censoring-informed reconstructions). Shaded band, pointwise 95% CI; dashed vertical line, main landmark; solid grey vertical line, primary τ.

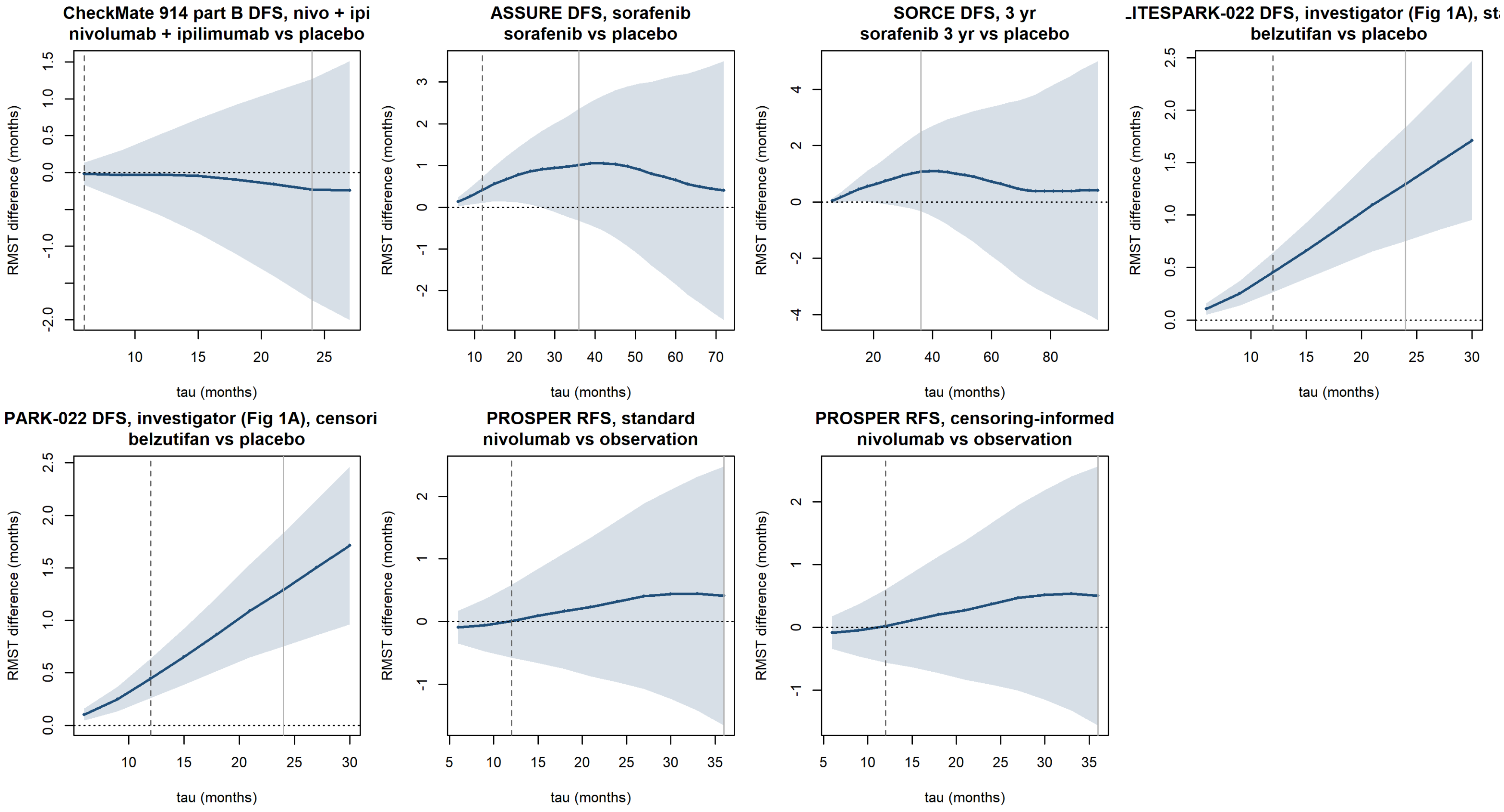

**Figure S2.** Time-varying HR(t) from flexible parametric models, main comparisons. Solid line and band, AIC-selected model (specification in the panel title) with 95% CI; dashed line, prespecified model with baseline df 3 and time-varying df 2; dashed vertical line, main landmark.

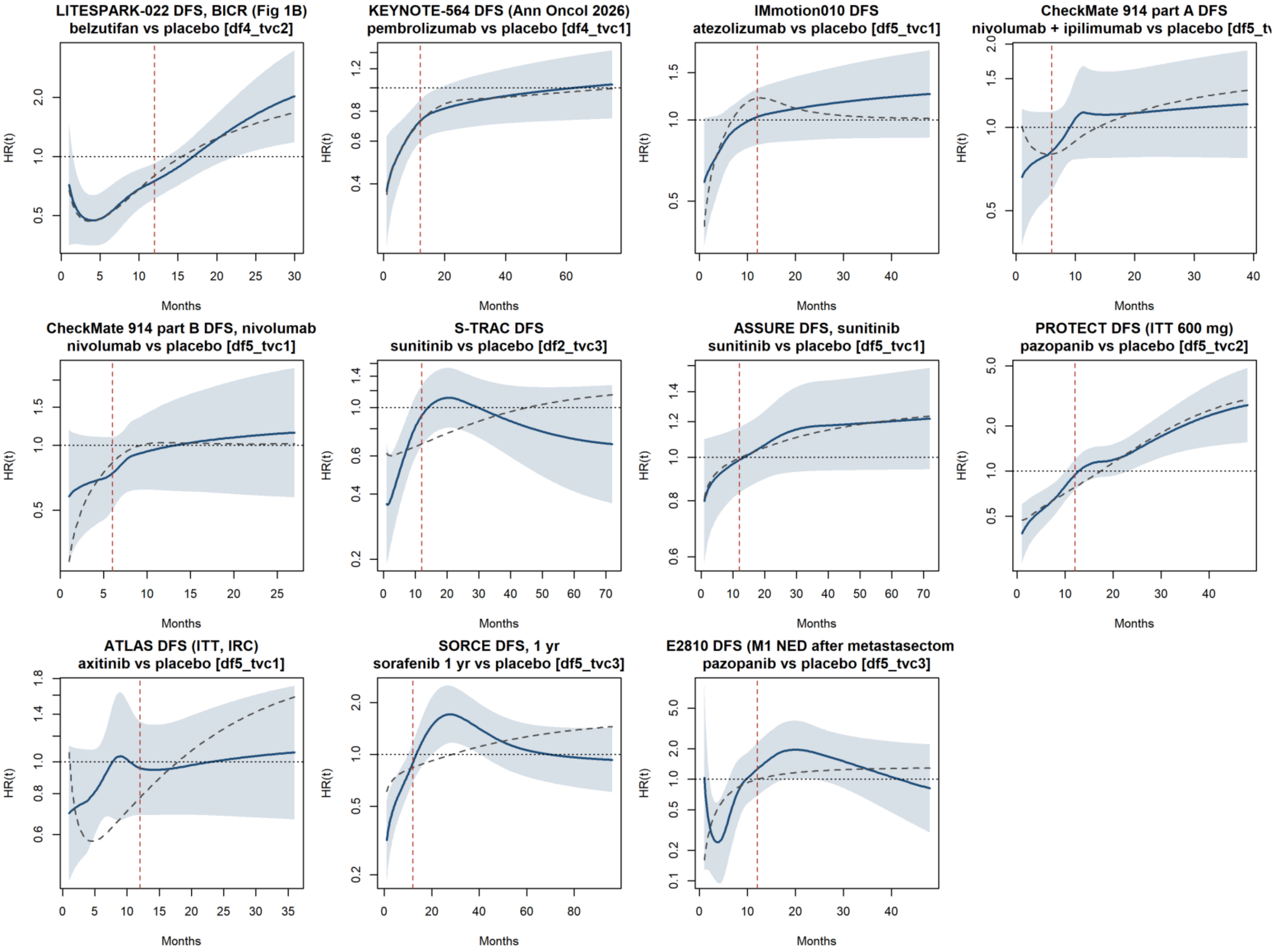

**Figure S3.** Time-varying HR(t), alternative comparisons and sensitivity curves (as in Figure S2).

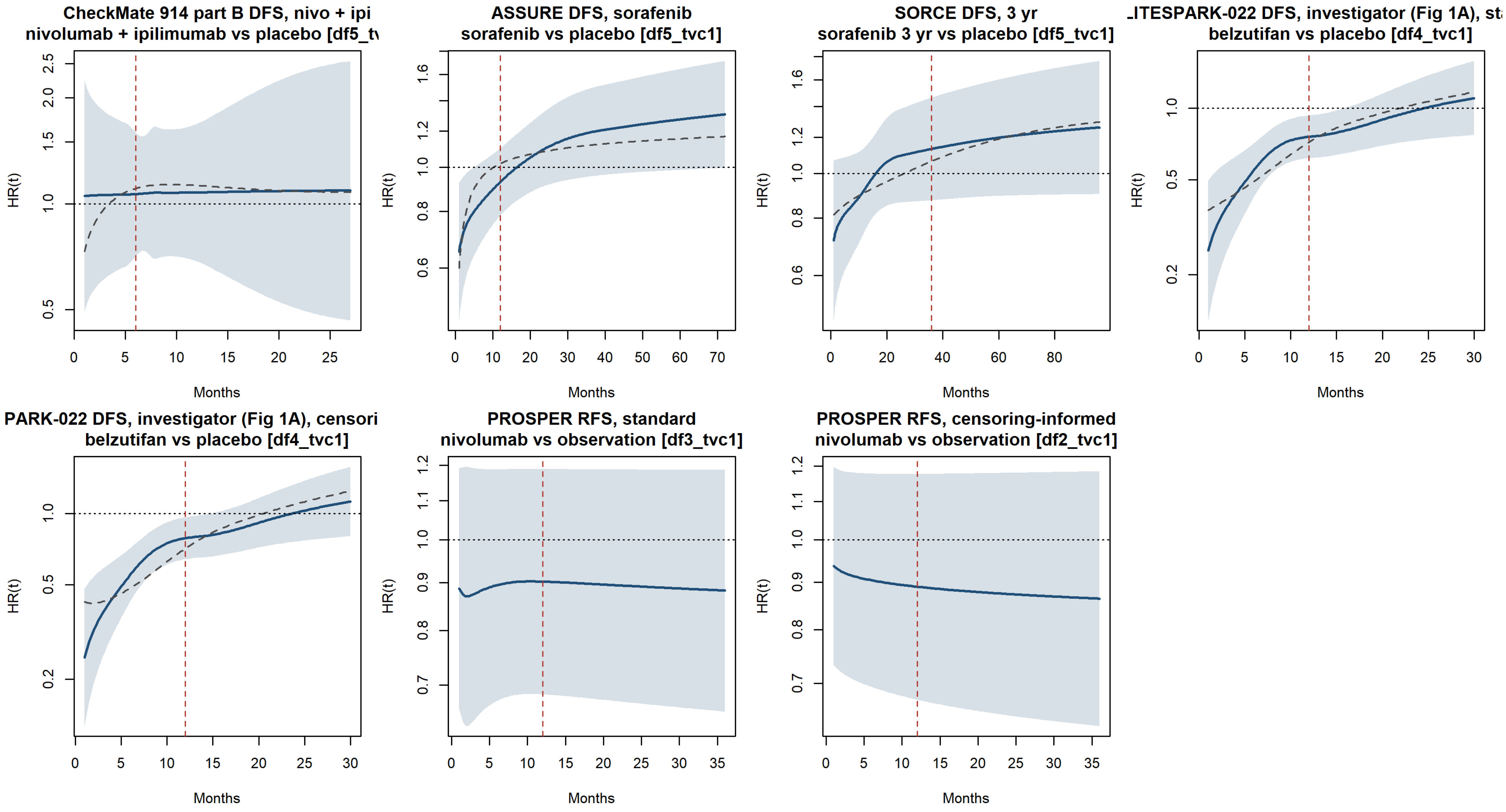

**Figure S4.** Time-varying HR(t) for overall survival (KEYNOTE-564; ASSURE sunitinib and sorafenib; LITESPARK-022, standard and censoring-informed reconstructions, descriptive).

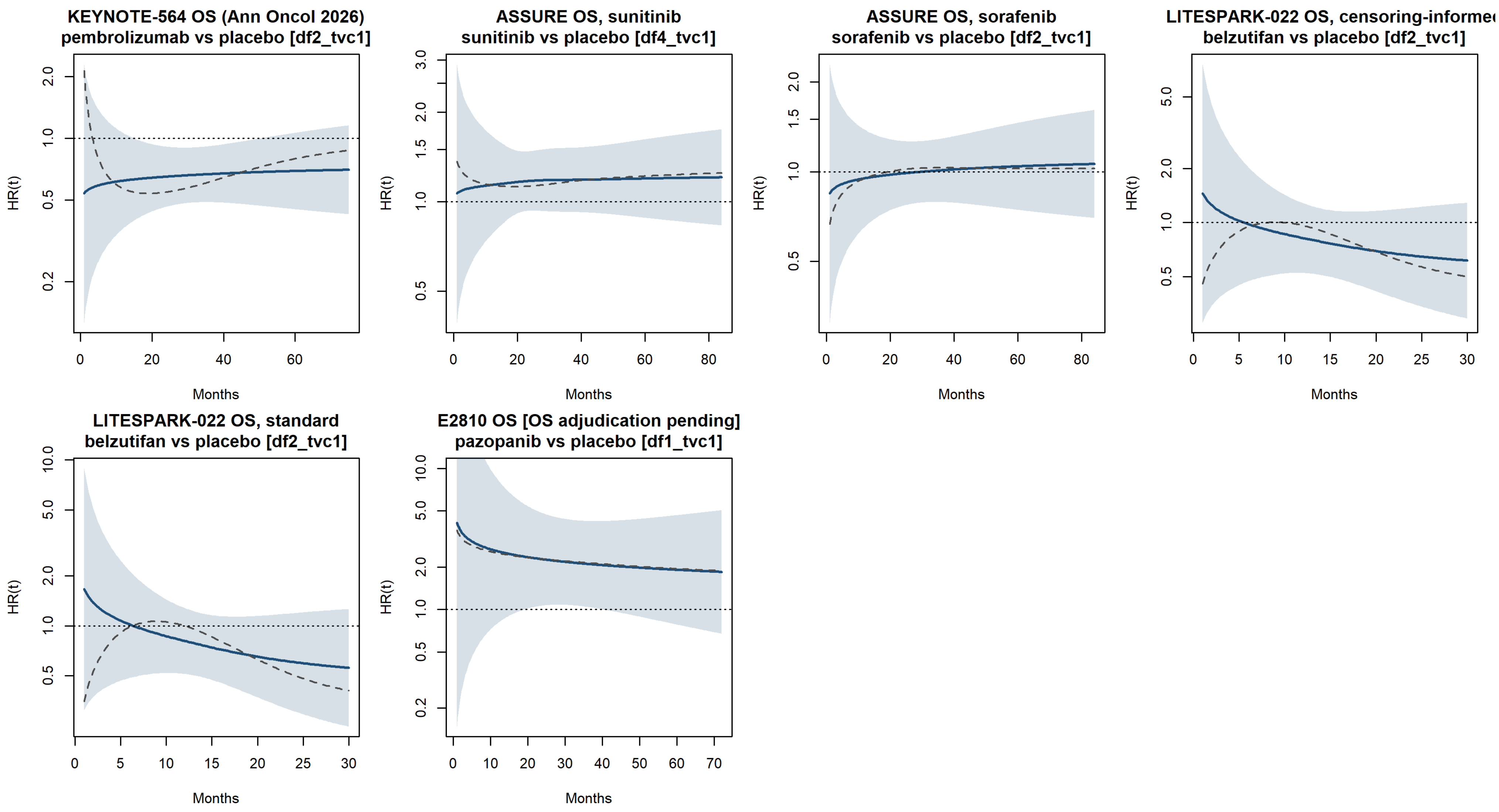

**Figure S5.** Log(−log) survival plots, main comparisons.

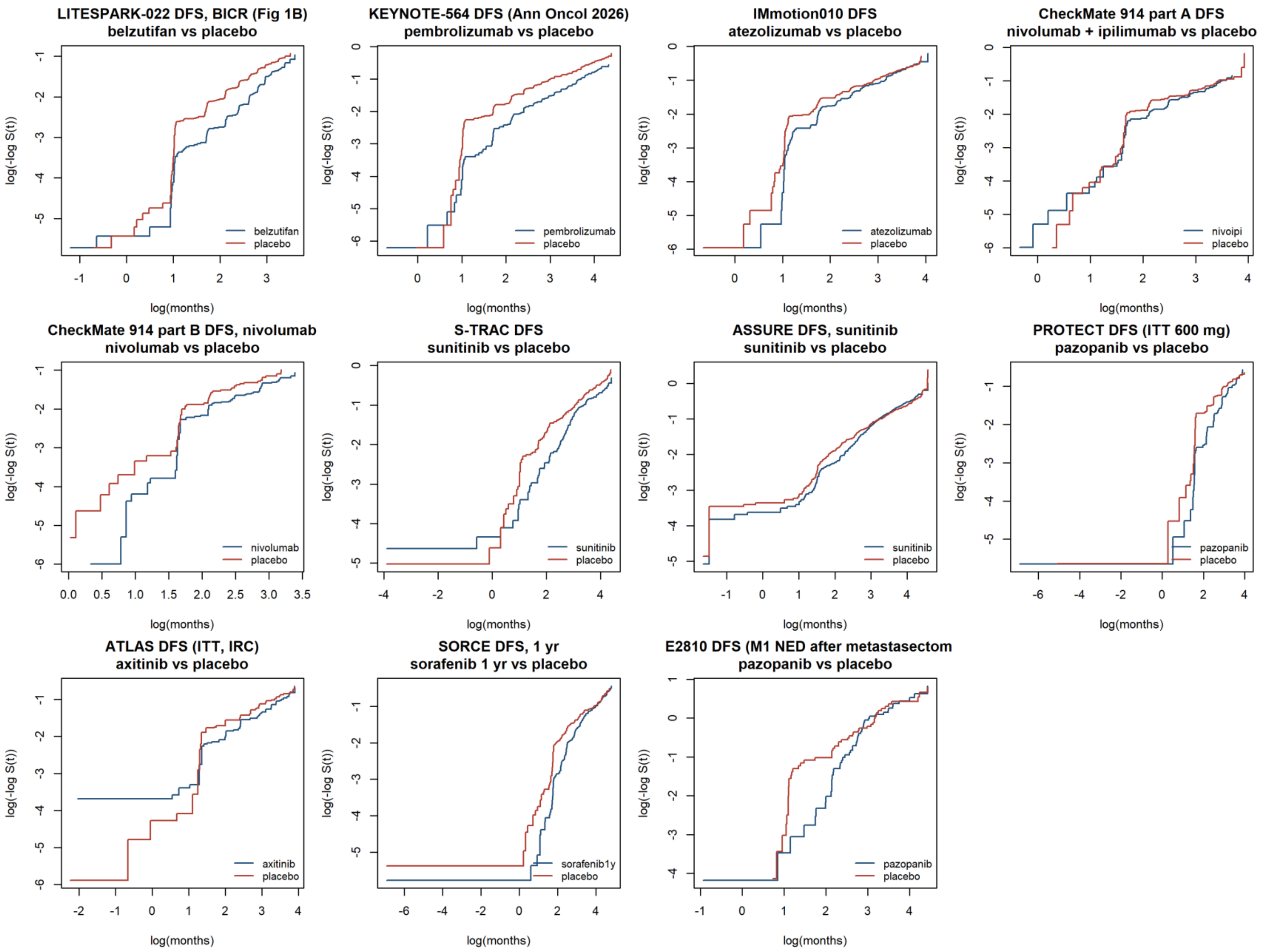

**Figure S6.** Log(−log) survival plots, alternative comparisons and sensitivity curves.

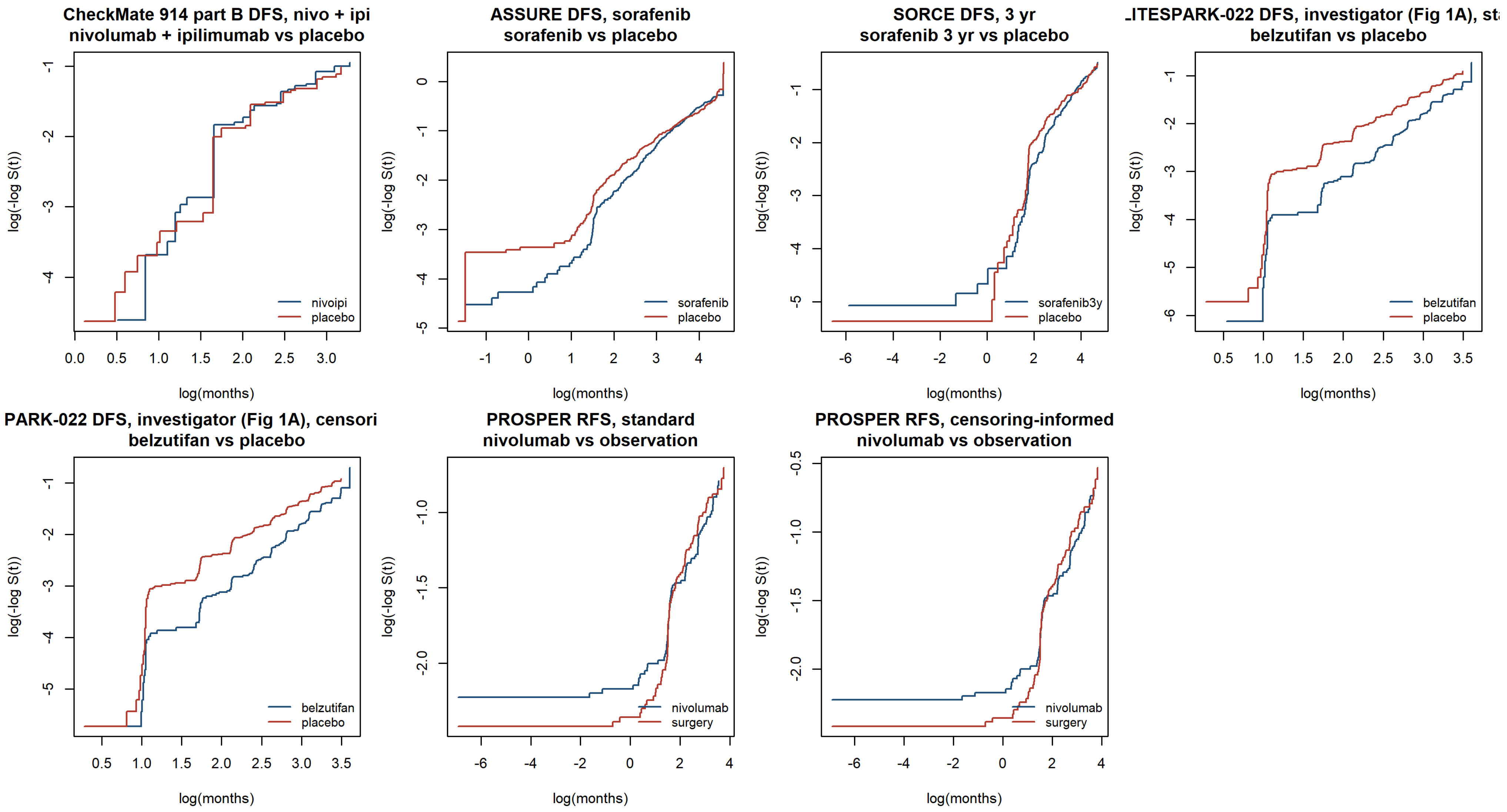

**Figure S7.** Smoothed hazard functions by arm (`muhaz`), main comparisons, from 0 to the maximum feasible τ.

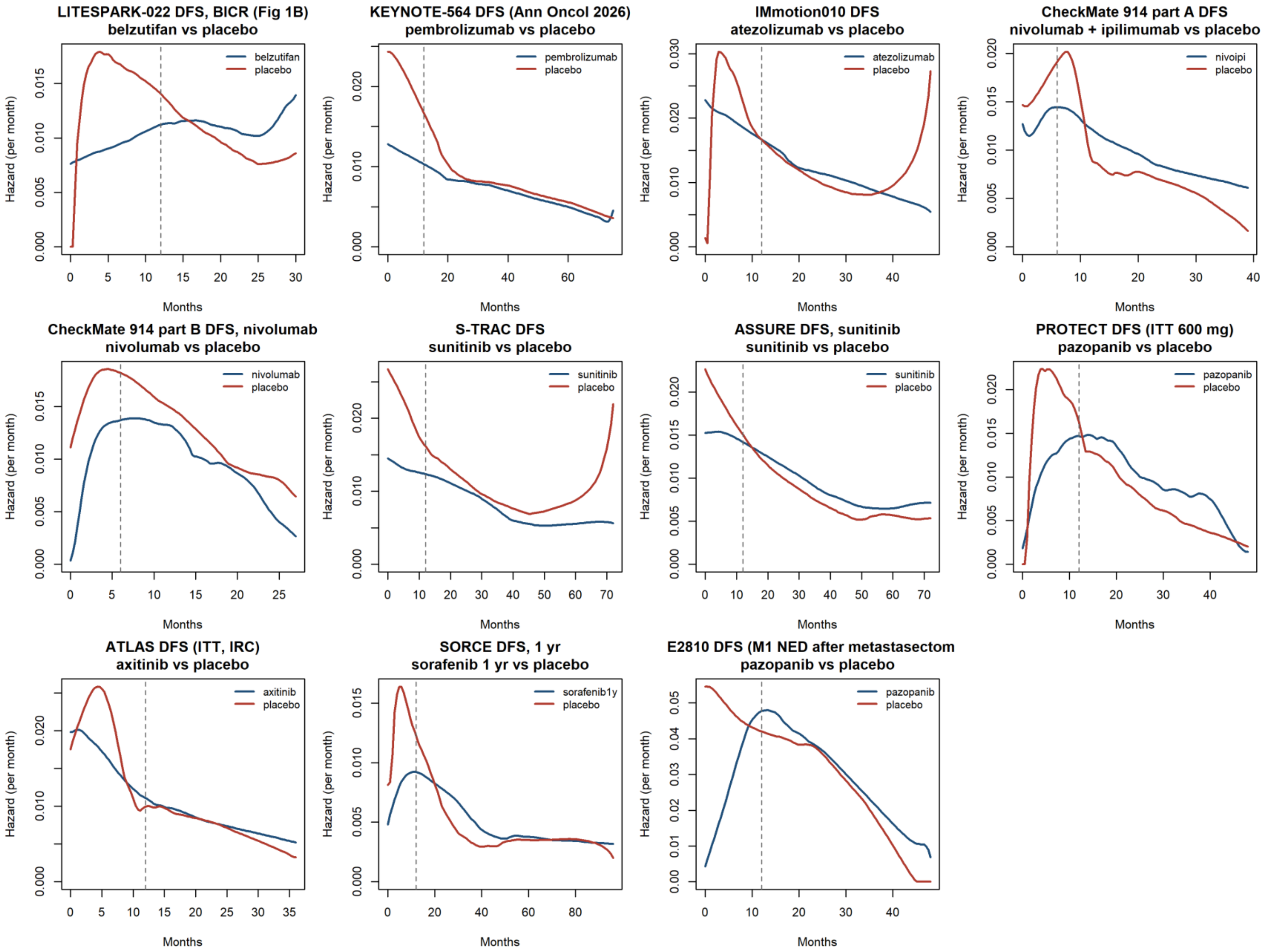

**Figure S8.** Smoothed hazard functions by arm, alternative comparisons and sensitivity curves.

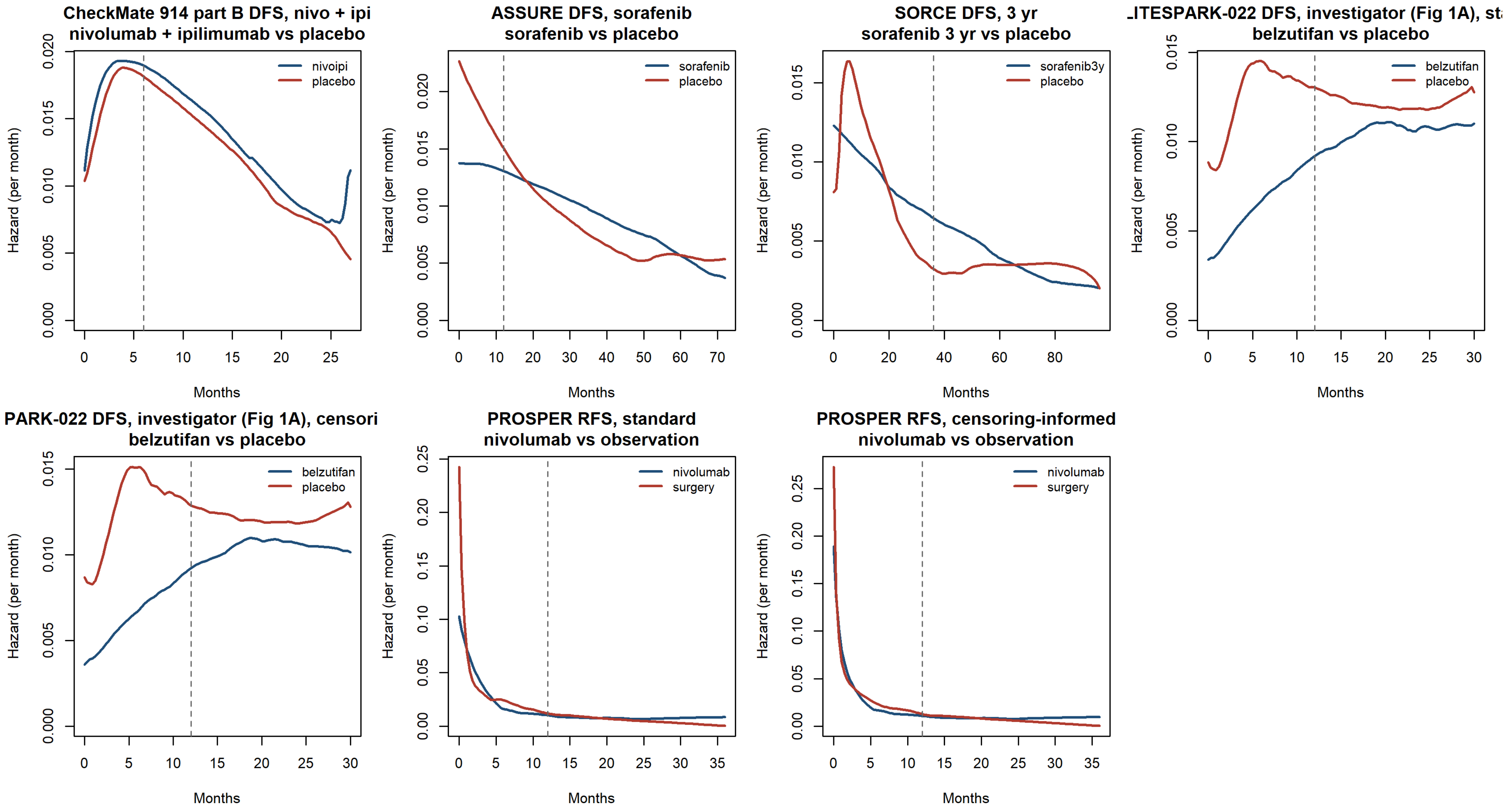

**Figure S9.** Window-matched analysis for all comparisons, including the alternative comparisons and PROSPER (post hoc, D7). (A) ΔRMST increment per year over the 18 months after the landmark, with bootstrap 95% CI; (B) KM DFS difference at the start and end of the window, with Greenwood 95% CI. Comparisons are grouped by class; LITESPARK-022 is shown in red and sensitivity curves with open symbols.

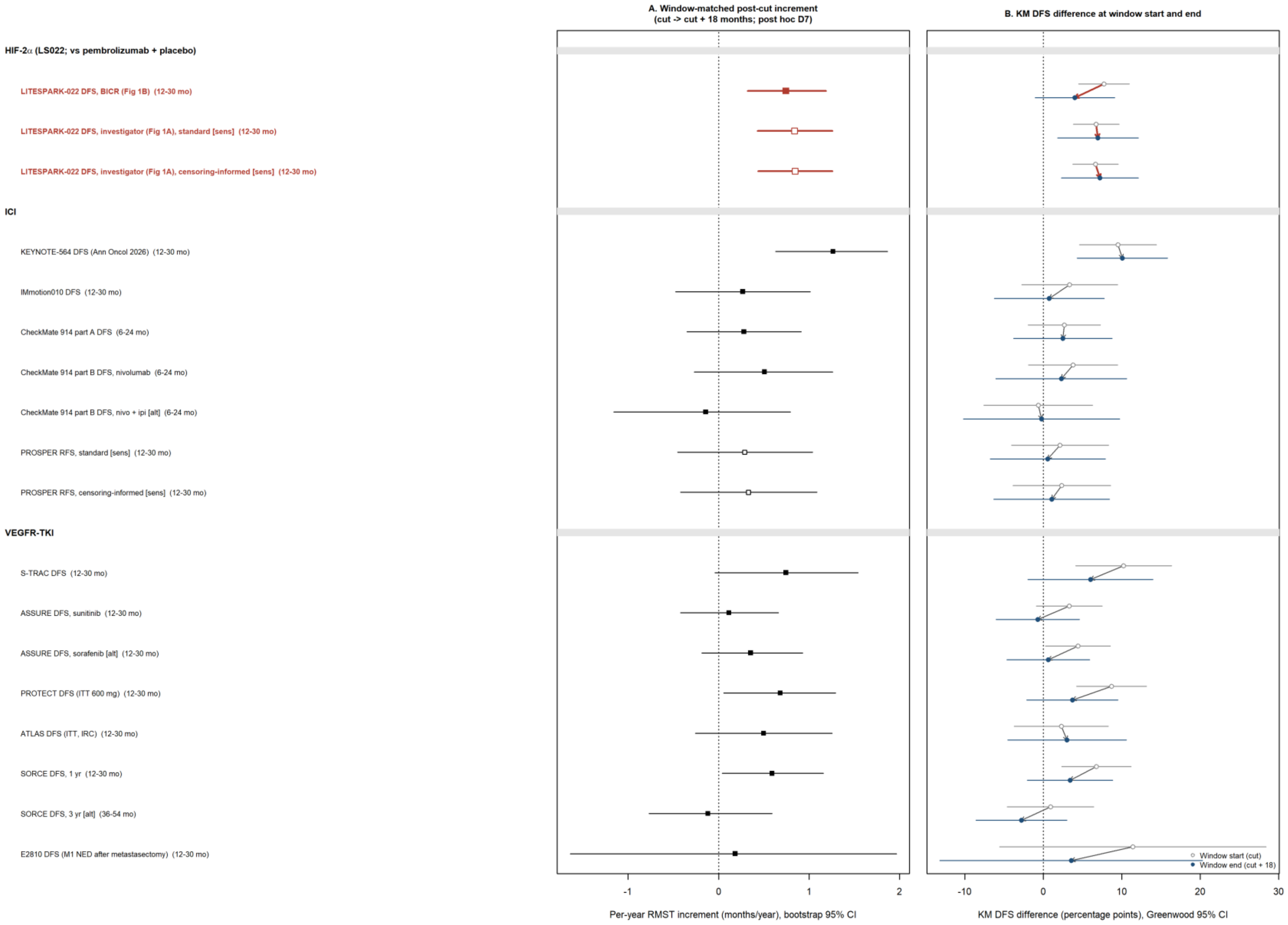

**Figure S10.** Reconstruction overlays: KM curve from the reconstructed IPD (thin line) drawn over the digitised curve (thick line), one panel per curve. Every overlay listed was checked independently by the second investigator, who found no discrepancy (review completed 14 October 2026; S-M4 and Table S5); for panel (ao), the authors' stricter judgement is retained. Each panel is shown below with its label.

**Figure S10a.** LITESPARK-022 Fig 1B.

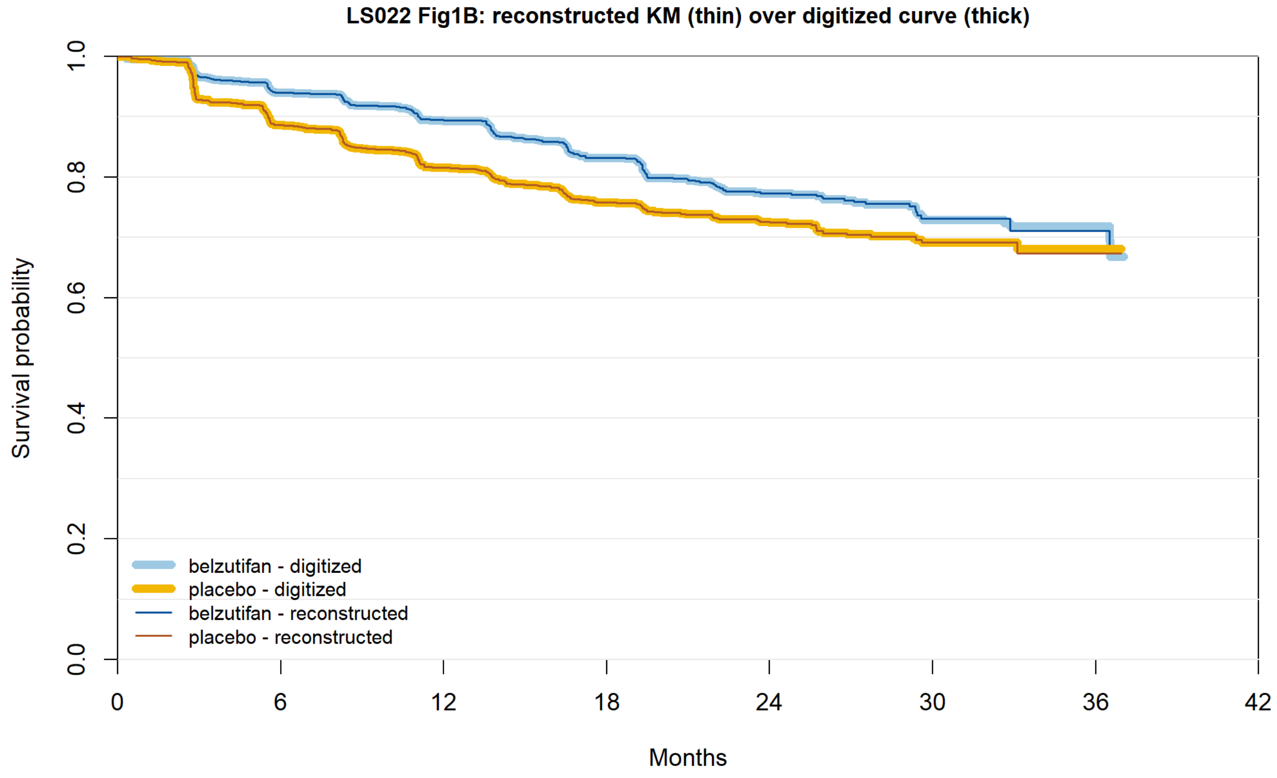


| | | | | | | | | |
|---|---|---|---|---|---|---|---|---|
| belzutifan reported | 921 | 837 | 786 | 643 | 424 | 141 | 34 | 0 |
| belzutifan recon. | 921 | 837 | 786 | 643 | 424 | 141 | 34 | 0 |
| placebo reported | 920 | 800 | 727 | 590 | 404 | 138 | 37 | 0 |
| placebo recon. | 920 | 800 | 727 | 590 | 404 | 138 | 37 | 0 |

**Figure S10b.** LITESPARK-022 Fig 1A, standard.

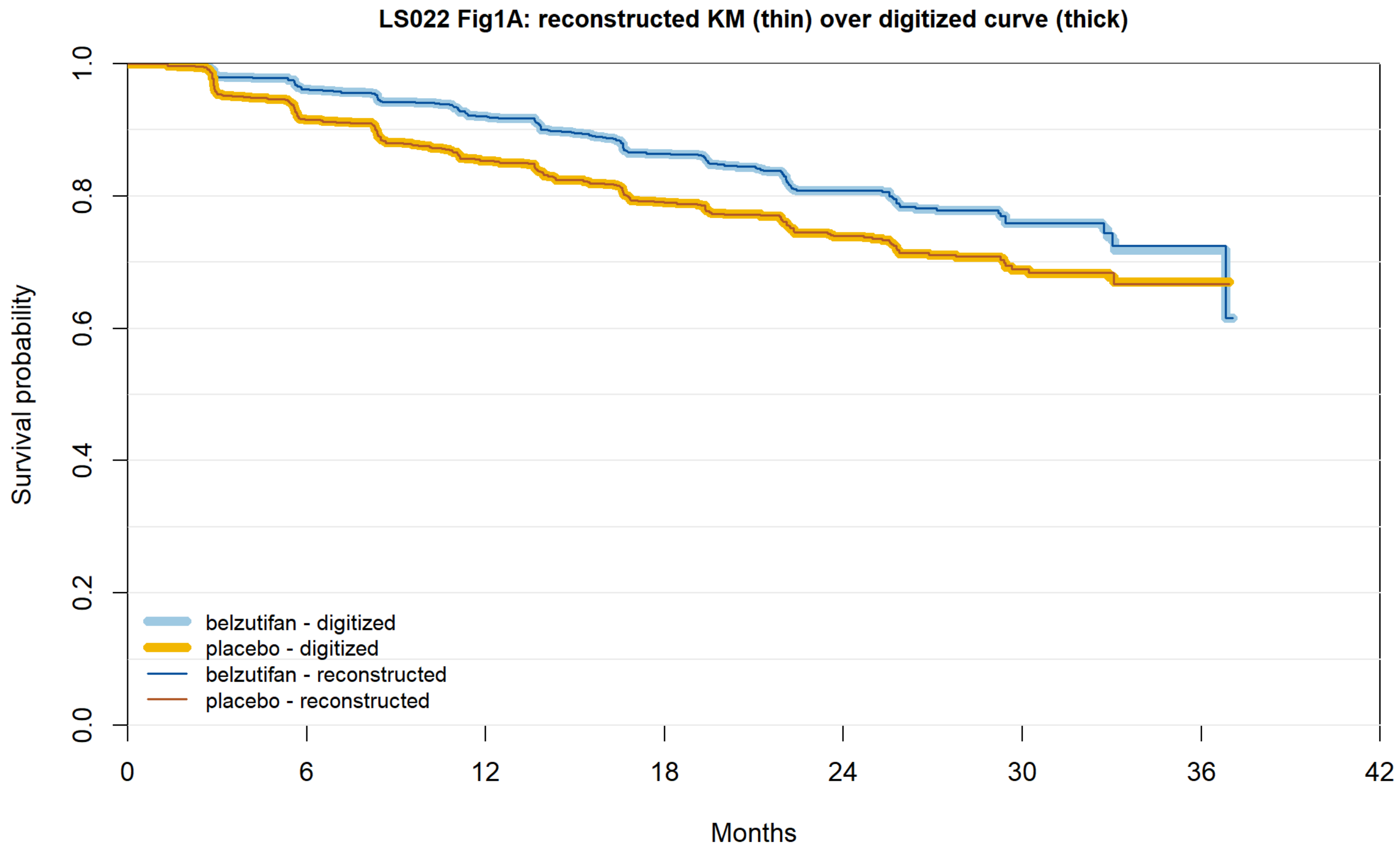


| | | | | | | | | |
|---|---|---|---|---|---|---|---|---|
| belzutifan reported | 921 | 850 | 805 | 669 | 451 | 153 | 35 | 0 |
| belzutifan recon. | 921 | 850 | 805 | 669 | 451 | 153 | 35 | 0 |
| placebo reported | 920 | 823 | 759 | 619 | 417 | 140 | 38 | 0 |
| placebo recon. | 920 | 823 | 759 | 619 | 417 | 140 | 38 | 0 |

**Figure S10c.** LITESPARK-022 Fig 1A, censoring-informed.

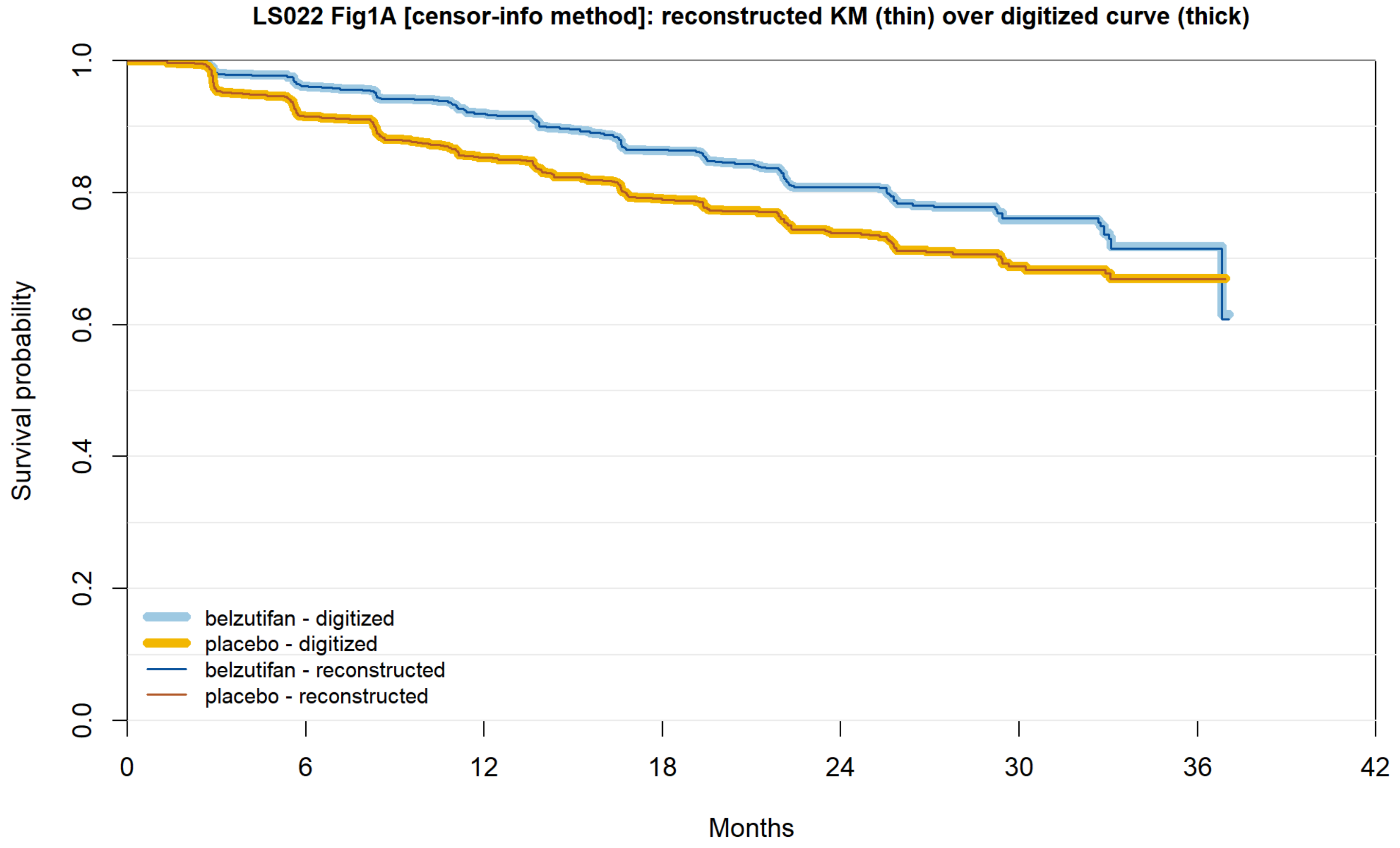


| | | | | | | | | |
|---|---|---|---|---|---|---|---|---|
| belzutifan reported | 921 | 850 | 805 | 669 | 451 | 153 | 35 | 0 |
| belzutifan recon. | 921 | 850 | 805 | 669 | 451 | 153 | 35 | 0 |
| placebo reported | 920 | 823 | 759 | 619 | 417 | 140 | 38 | 0 |
| placebo recon. | 920 | 823 | 759 | 619 | 417 | 140 | 38 | 0 |

**Figure S10d.** LITESPARK-022 Fig 2 (OS), standard.

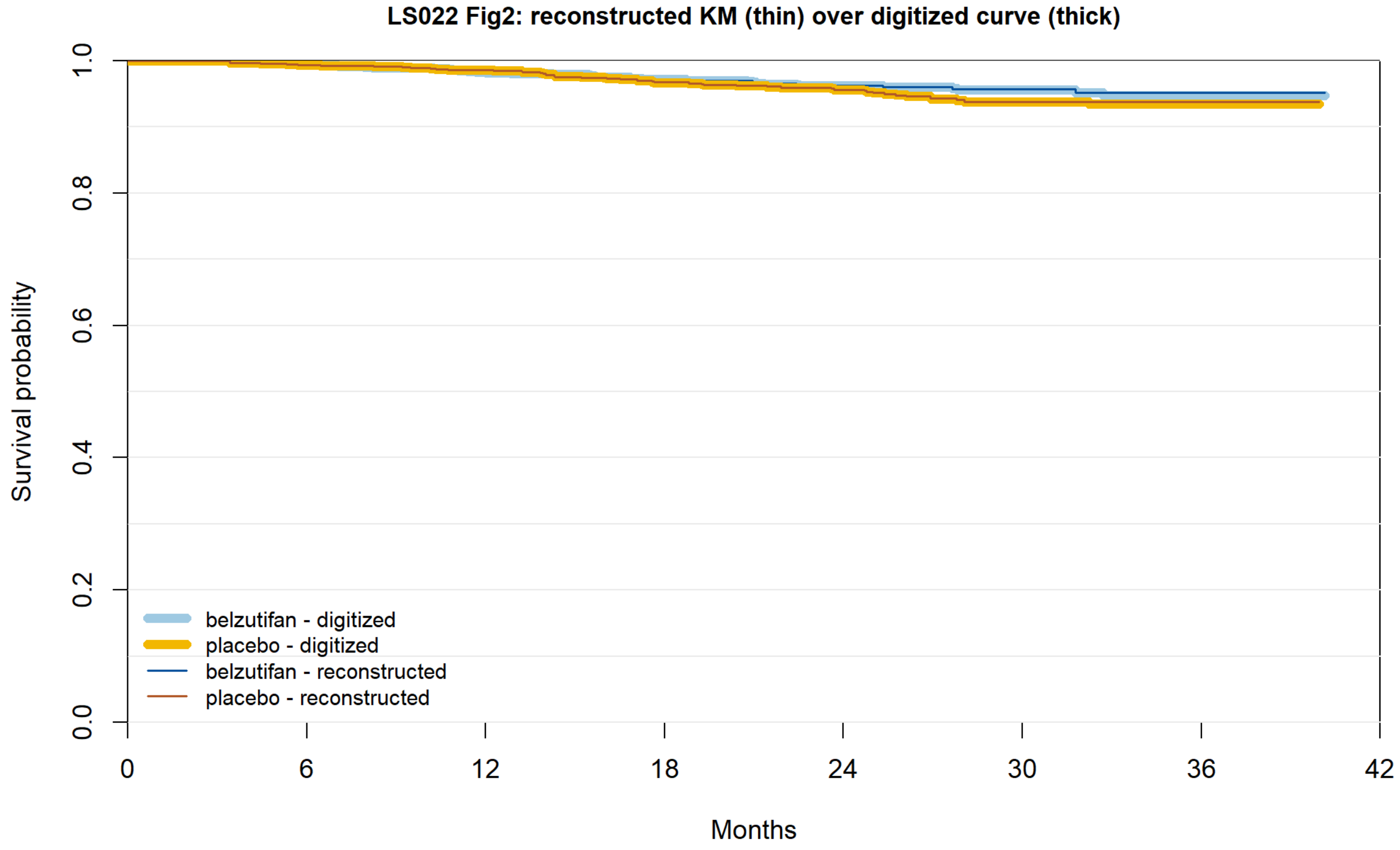


| | | | | | | | | |
|---|---|---|---|---|---|---|---|---|
| belzutifan reported | 921 | 914 | 900 | 805 | 631 | 319 | 70 | 0 |
| belzutifan recon. | 921 | 914 | 900 | 805 | 631 | 319 | 70 | 0 |
| placebo reported | 920 | 914 | 907 | 807 | 638 | 319 | 69 | 0 |
| placebo recon. | 920 | 914 | 907 | 807 | 638 | 319 | 69 | 0 |

**Figure S10e.** LITESPARK-022 Fig 2 (OS), censoring-informed.

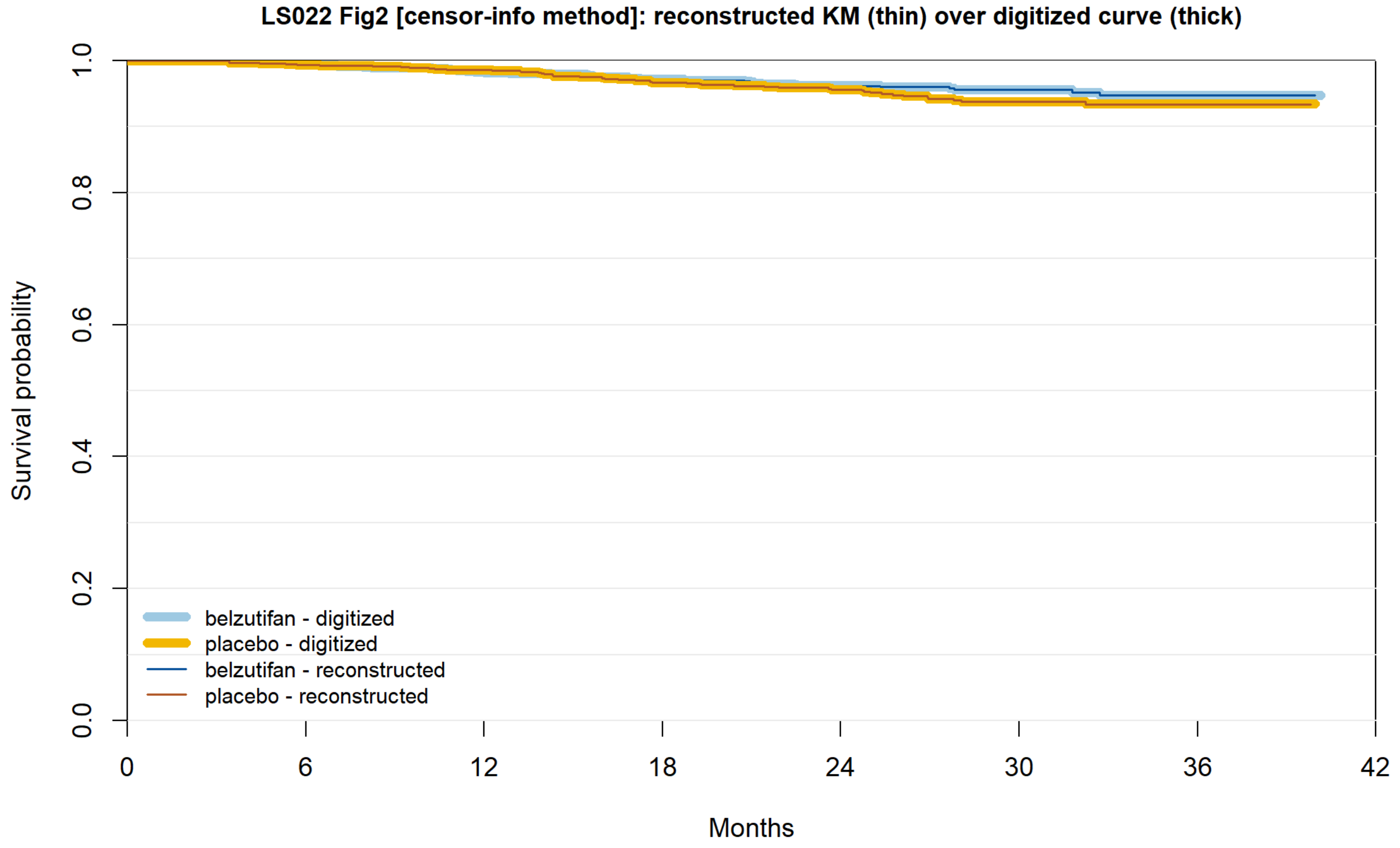


| | | | | | | | | |
|---|---|---|---|---|---|---|---|---|
| belzutifan reported | 921 | 914 | 900 | 805 | 631 | 319 | 70 | 0 |
| belzutifan recon. | 921 | 914 | 900 | 805 | 631 | 319 | 70 | 0 |
| placebo reported | 920 | 914 | 907 | 807 | 638 | 319 | 69 | 0 |
| placebo recon. | 920 | 914 | 907 | 807 | 638 | 319 | 69 | 0 |

**Figure S10f.** KEYNOTE-564 DFS.

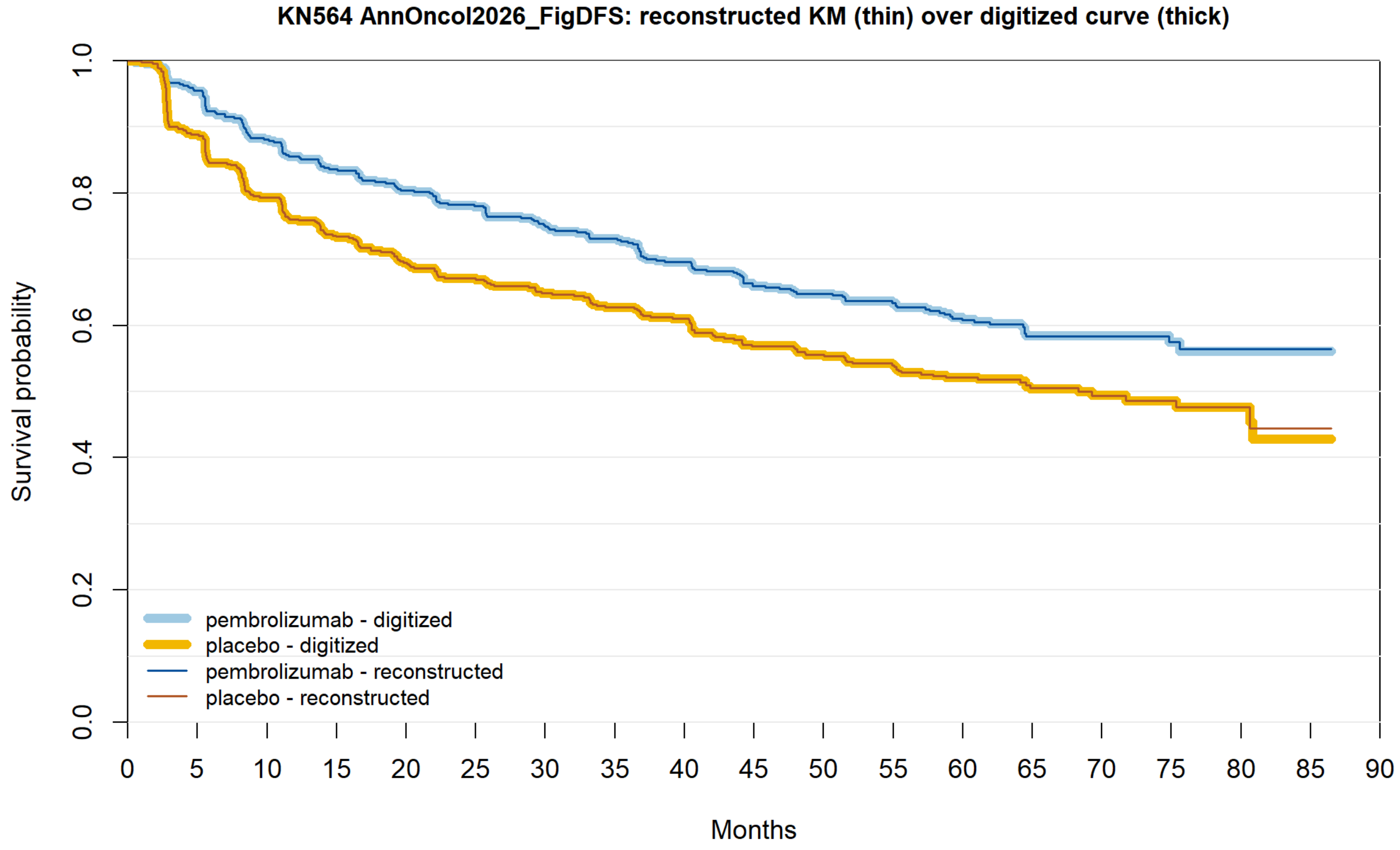


| | 0 | 5 | 10 | 15 | 20 | 25 | 30 | 35 | 40 | 45 | 50 | 55 | 60 | 65 | 70 | 75 | 80 | 85 | 90 |
|---|---|---|---|---|---|---|---|---|---|---|---|---|---|---|---|---|---|---|---|
| pembrolizumab reported | 496 | 459 | 417 | 388 | 371 | 356 | 338 | 328 | 309 | 291 | 282 | 269 | 207 | 128 | 88 | 63 | 26 | 1 | 0 |
| pembrolizumab recon. | 496 | 459 | 417 | 388 | 371 | 356 | 338 | 328 | 309 | 291 | 282 | 269 | 207 | 128 | 88 | 63 | 26 | 1 | 0 |
| placebo reported | 498 | 438 | 390 | 357 | 334 | 322 | 308 | 293 | 283 | 260 | 251 | 240 | 179 | 118 | 71 | 55 | 21 | 1 | 0 |
| placebo recon. | 498 | 438 | 390 | 357 | 334 | 322 | 308 | 293 | 283 | 260 | 251 | 240 | 179 | 118 | 71 | 55 | 21 | 1 | 0 |

**Figure S10g.** KEYNOTE-564 OS.

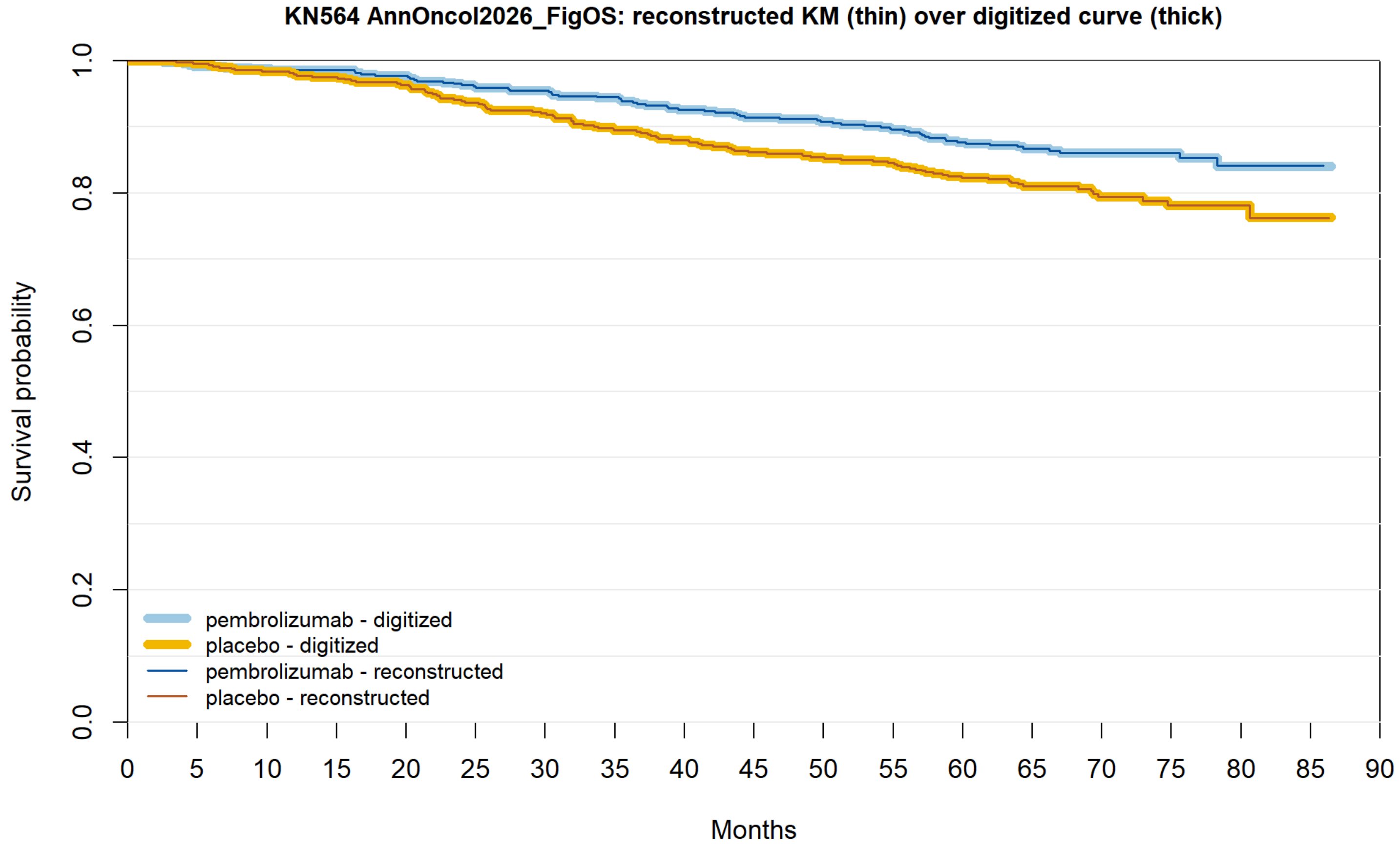


| | | | | | | | | | | | | | | | | | | |
|---|---|---|---|---|---|---|---|---|---|---|---|---|---|---|---|---|---|---|
| pembrolizumab reported | 496 | 489 | 486 | 484 | 479 | 470 | 468 | 462 | 452 | 445 | 442 | 436 | 426 | 310 | 198 | 121 | 50 | 3 | 0 |
| pembrolizumab recon. | 496 | 489 | 486 | 484 | 479 | 470 | 468 | 462 | 452 | 445 | 442 | 436 | 426 | 310 | 198 | 121 | 50 | 3 | 0 |
| placebo reported | 498 | 494 | 487 | 483 | 476 | 463 | 455 | 441 | 433 | 423 | 419 | 413 | 400 | 284 | 177 | 111 | 46 | 4 | 0 |
| placebo recon. | 498 | 494 | 487 | 483 | 476 | 463 | 455 | 441 | 433 | 423 | 419 | 413 | 400 | 284 | 177 | 111 | 46 | 4 | 0 |

**Figure S10h.** IMmotion010.

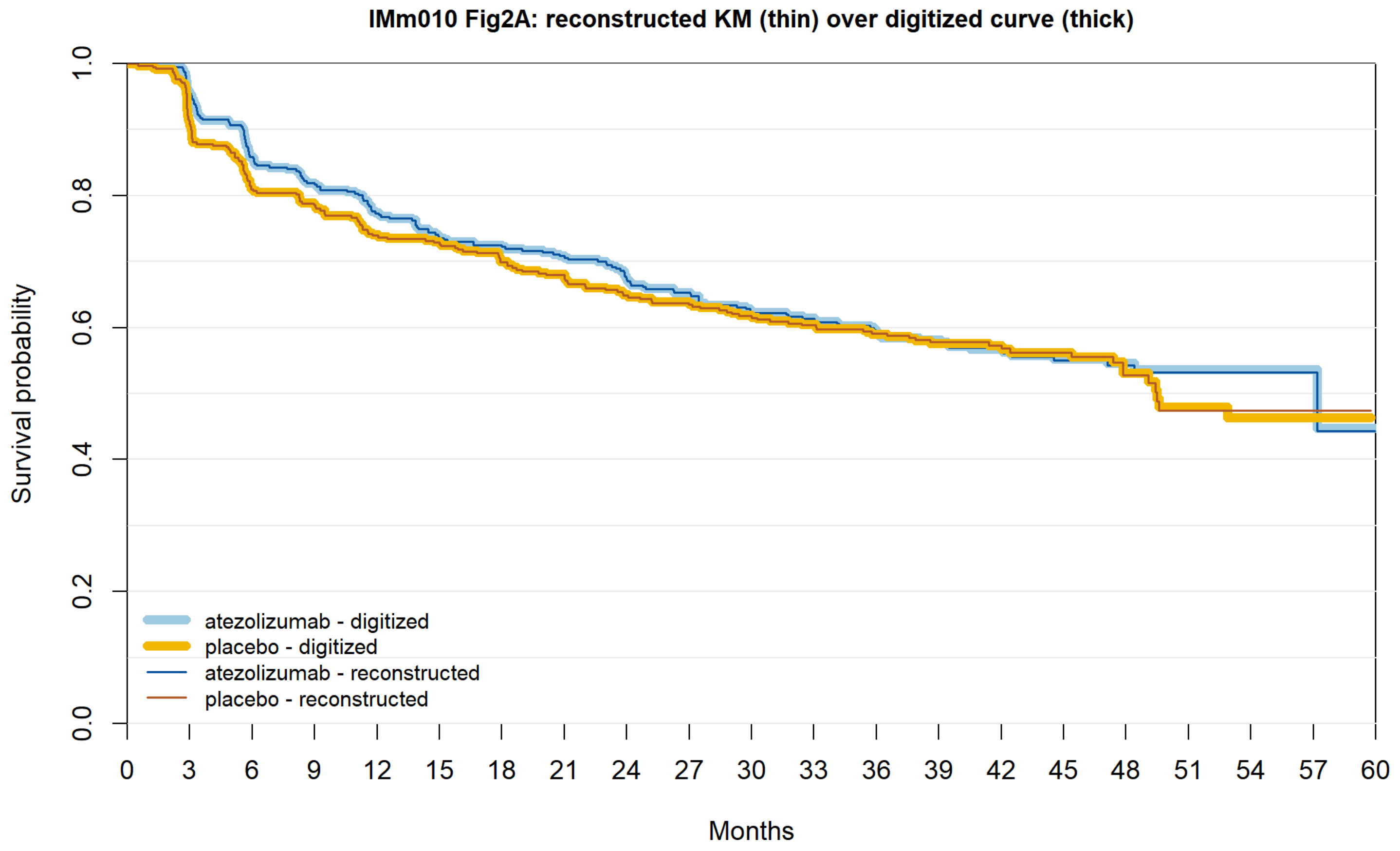


| | 0 | 3 | 6 | 9 | 12 | 15 | 18 | 21 | 24 | 27 | 30 | 33 | 36 | 39 | 42 | 45 | 48 | 51 | 54 | 57 | 60 |
|---|---|---|---|---|---|---|---|---|---|---|---|---|---|---|---|---|---|---|---|---|---|
| atezolizumab reported | 390 | 360 | 322 | 306 | 288 | 272 | 265 | 257 | 244 | 234 | 222 | 218 | 194 | 171 | 124 | 100 | 75 | 48 | 22 | 6 | 1 |
| atezolizumab recon. | 390 | 360 | 322 | 306 | 288 | 272 | 265 | 257 | 244 | 234 | 222 | 218 | 194 | 171 | 124 | 100 | 75 | 48 | 22 | 6 | 0 |
| placebo reported | 388 | 343 | 305 | 294 | 275 | 268 | 254 | 243 | 232 | 226 | 216 | 209 | 187 | 161 | 121 | 91 | 56 | 33 | 15 | 3 | |
| placebo recon. | 388 | 343 | 305 | 294 | 275 | 268 | 254 | 243 | 232 | 226 | 216 | 209 | 187 | 161 | 121 | 91 | 56 | 33 | 15 | 3 | 0 |

**Figure S10i.** CheckMate 914 part A.

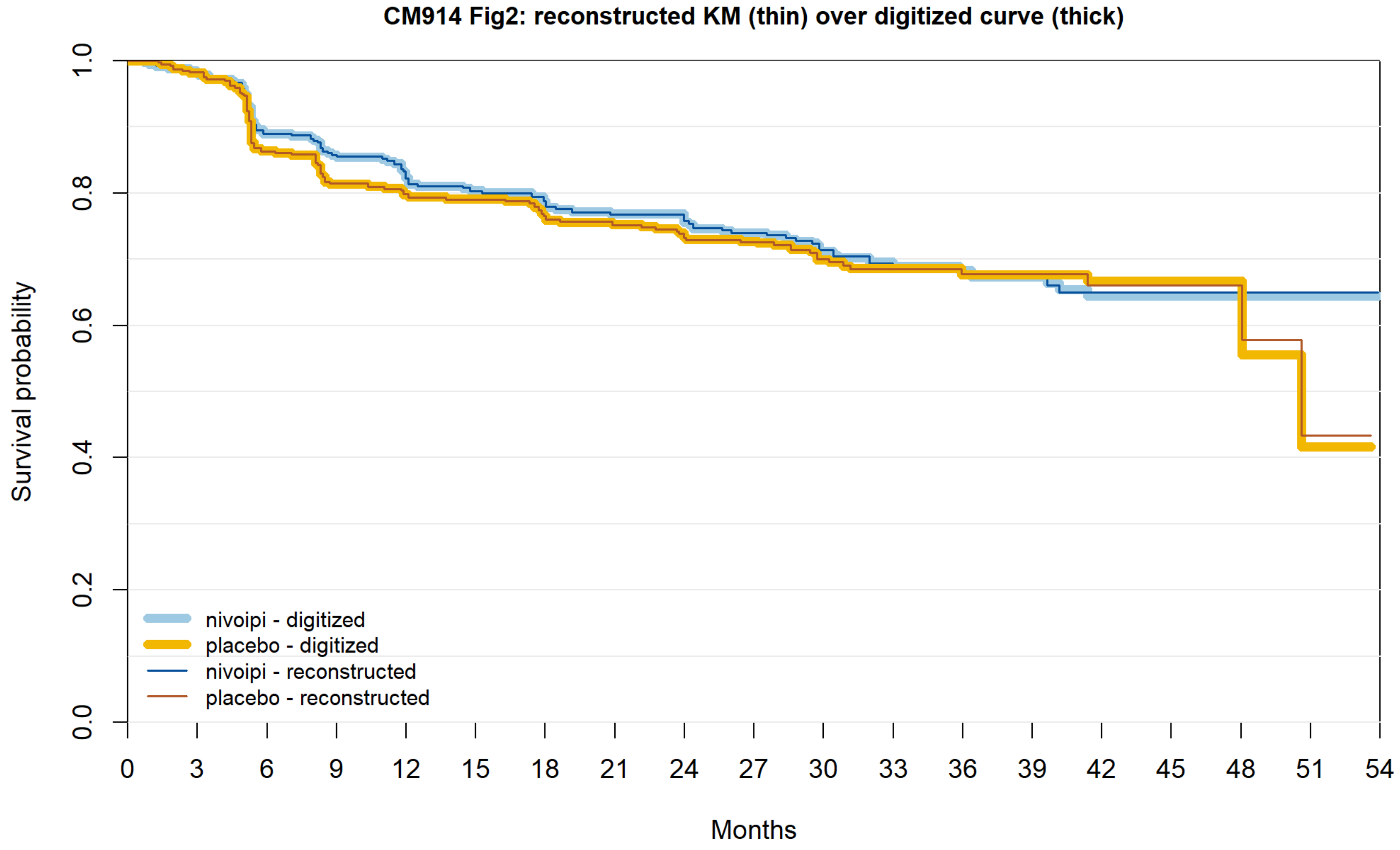


| | | | | | | | | | | | | | | | | | | | |
|---|---|---|---|---|---|---|---|---|---|---|---|---|---|---|---|---|---|---|---|
| nivoipi reported | 405 | 378 | 337 | 316 | 299 | 289 | 270 | 259 | 224 | 203 | 150 | 125 | 89 | 73 | 42 | 34 | 13 | 9 | 0 |
| nivoipi recon. | 405 | 378 | 337 | 316 | 299 | 289 | 270 | 259 | 224 | 203 | 150 | 125 | 89 | 73 | 42 | 34 | 13 | 9 | 0 |
| placebo reported | 411 | 391 | 340 | 315 | 299 | 293 | 275 | 268 | 227 | 205 | 155 | 128 | 90 | 66 | 38 | 25 | 8 | 3 | 0 |
| placebo recon. | 411 | 391 | 340 | 315 | 299 | 293 | 275 | 268 | 227 | 205 | 155 | 128 | 90 | 66 | 38 | 25 | 8 | 3 | 0 |

**Figure S10j.** CheckMate 914 part B nivolumab.

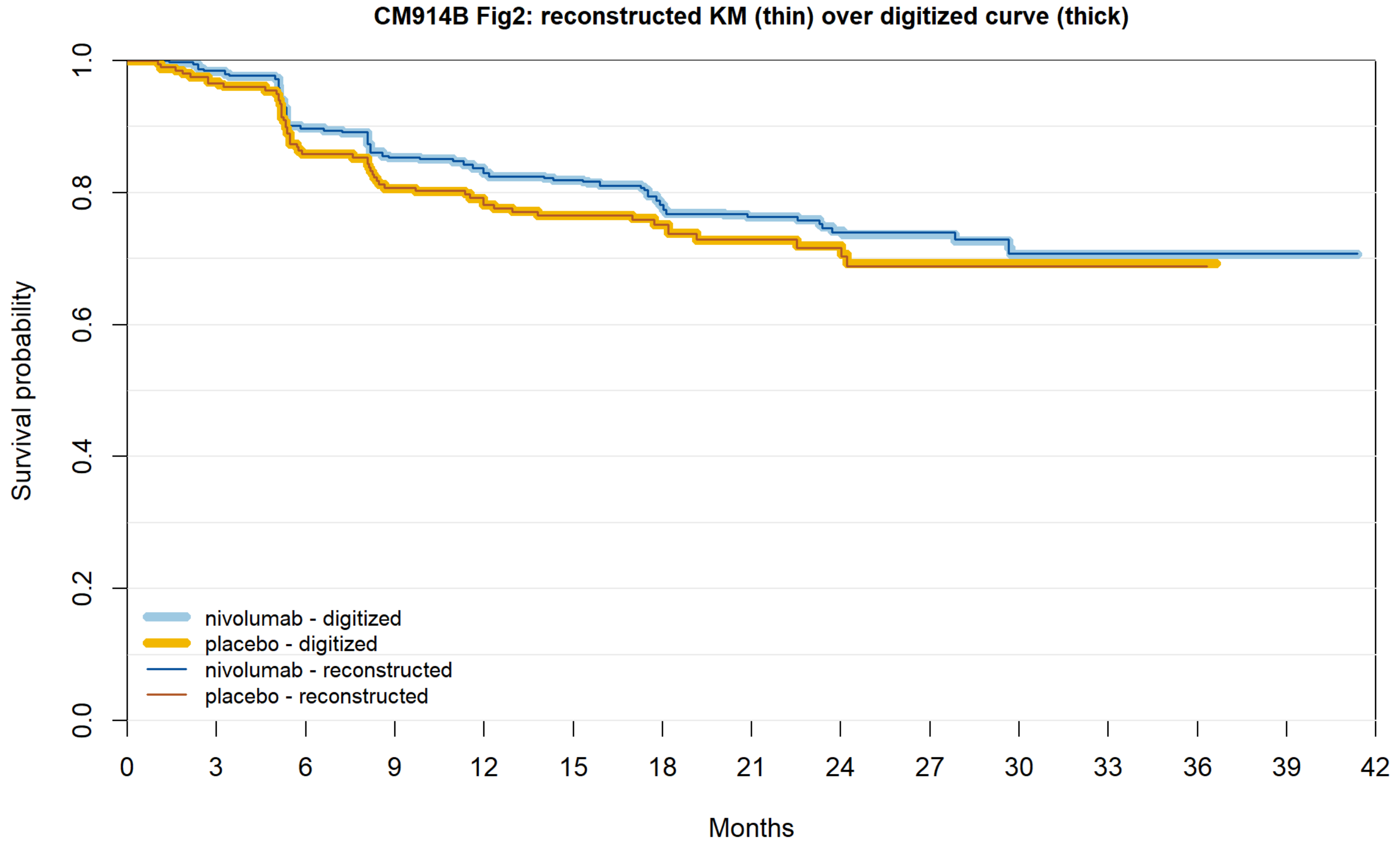


| | | | | | | | | | | | | | | | |
|---|---|---|---|---|---|---|---|---|---|---|---|---|---|---|---|
| nivolumab reported | 411 | 393 | 356 | 332 | 317 | 307 | 232 | 189 | 110 | 88 | 32 | 20 | 7 | 1 | 0 |
| nivolumab recon. | 411 | 393 | 356 | 332 | 317 | 307 | 232 | 189 | 110 | 88 | 32 | 20 | 7 | 1 | 0 |
| placebo reported | 208 | 193 | 169 | 158 | 150 | 143 | 105 | 86 | 57 | 46 | 14 | 8 | 2 | 0 | 0 |
| placebo recon. | 208 | 193 | 169 | 158 | 150 | 143 | 105 | 86 | 57 | 46 | 14 | 8 | 2 | 0 | 0 |

**Figure S10k.** CheckMate 914 part B nivolumab plus ipilimumab.

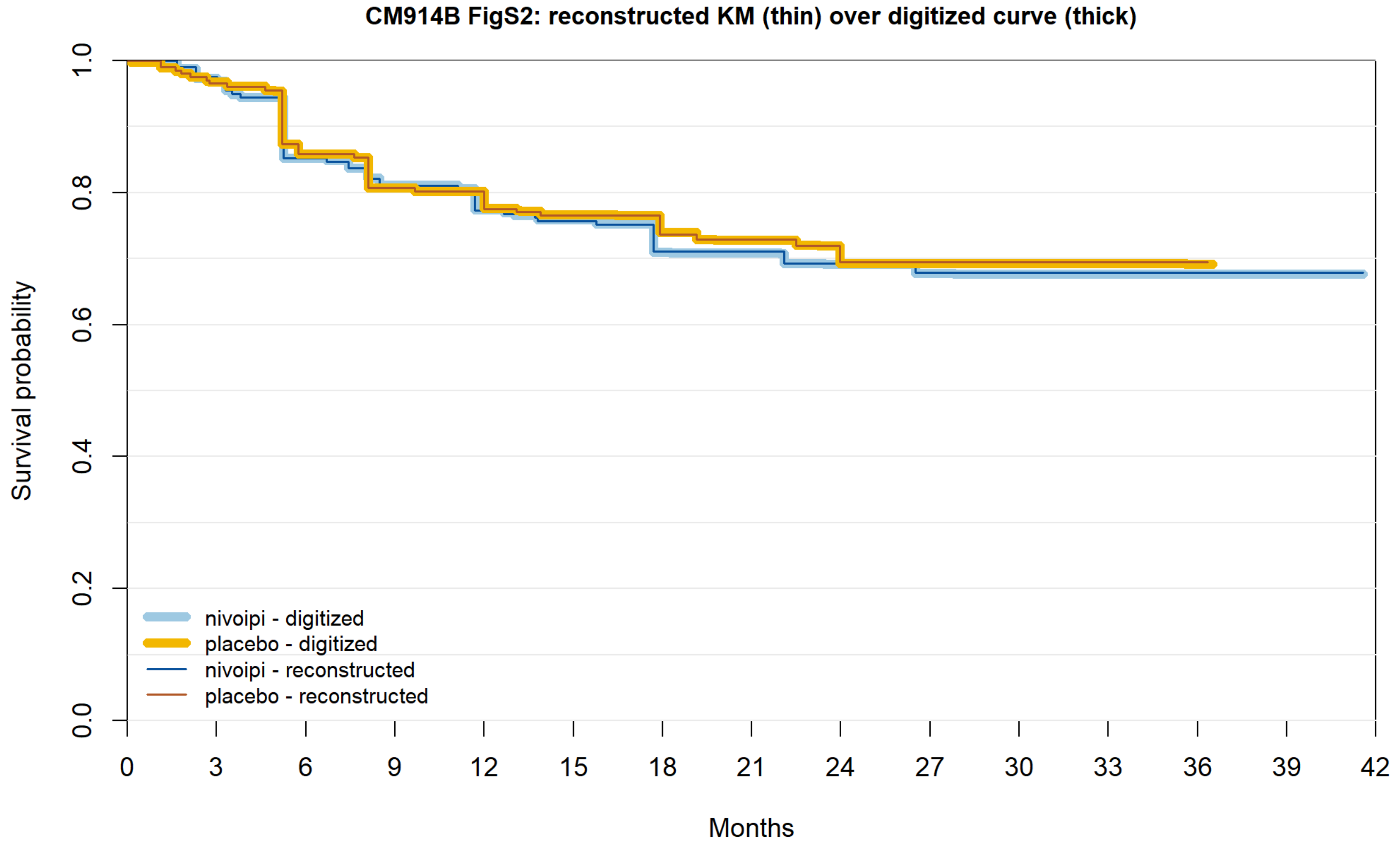


| | | | | | | | | | | | | | | | |
|---|---|---|---|---|---|---|---|---|---|---|---|---|---|---|---|
| nivoipi reported | 206 | 194 | 165 | 155 | 145 | 138 | 106 | 91 | 54 | 46 | 19 | 12 | 1 | 1 | 0 |
| nivoipi recon. | 206 | 194 | 165 | 155 | 145 | 138 | 106 | 91 | 54 | 46 | 19 | 12 | 1 | 1 | 0 |
| placebo reported | 208 | 193 | 169 | 158 | 150 | 143 | 105 | 86 | 57 | 46 | 14 | 8 | 2 | 0 | 0 |
| placebo recon. | 208 | 193 | 169 | 158 | 150 | 143 | 105 | 86 | 57 | 46 | 14 | 8 | 2 | 0 | 0 |

**Figure S10l.** S-TRAC.

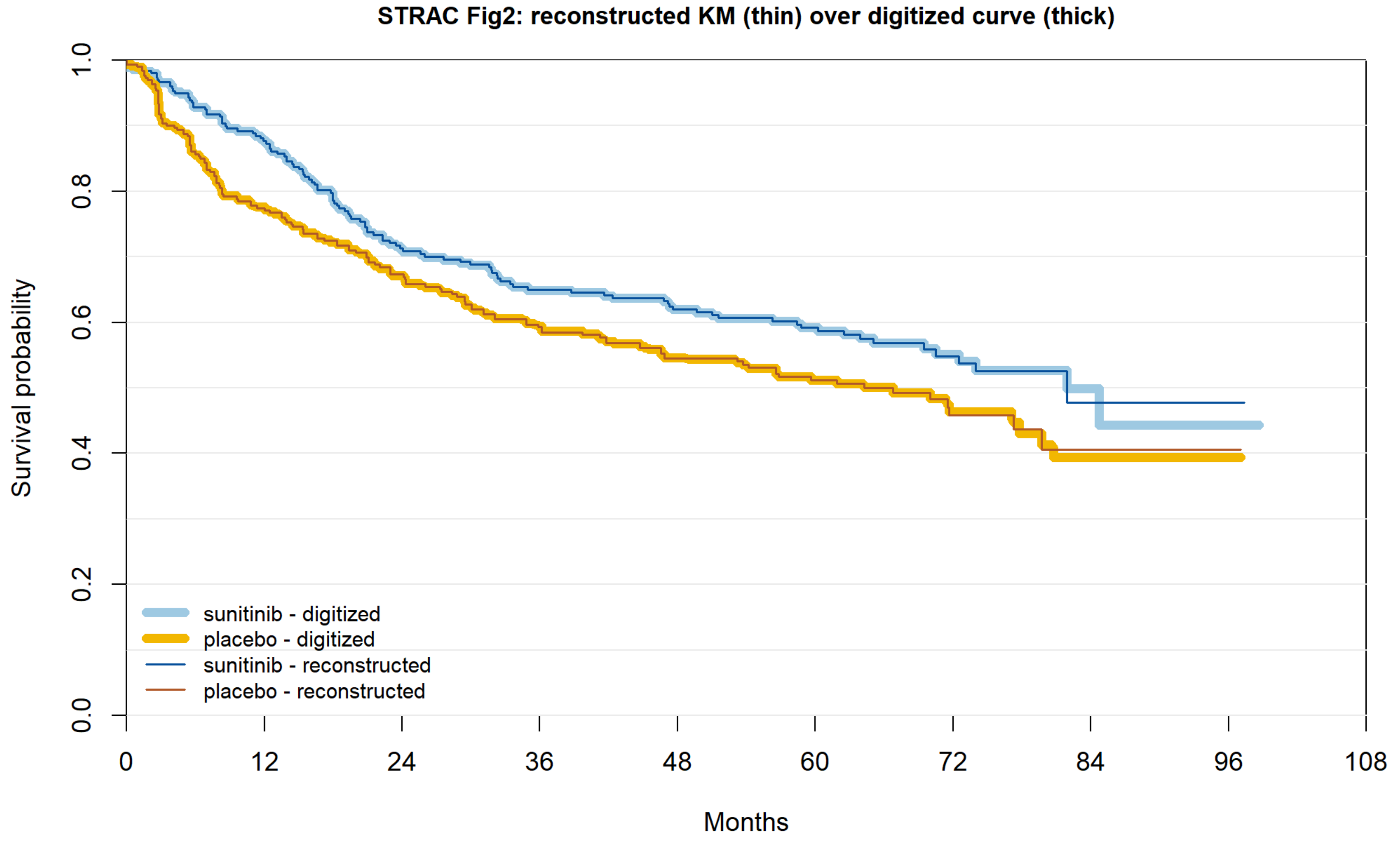


| | | | | | | | | | | |
|---|---|---|---|---|---|---|---|---|---|---|
| sunitinib reported | 309 | 225 | 173 | 153 | 144 | 119 | 53 | 10 | 3 | 0 |
| sunitinib recon. | 309 | 225 | 173 | 153 | 144 | 119 | 53 | 10 | 3 | 0 |
| placebo reported | 306 | 220 | 181 | 150 | 135 | 102 | 37 | 10 | 2 | 0 |
| placebo recon. | 306 | 220 | 181 | 150 | 135 | 102 | 37 | 10 | 2 | 0 |

**Figure S10m.** ASSURE DFS, sunitinib vs placebo.

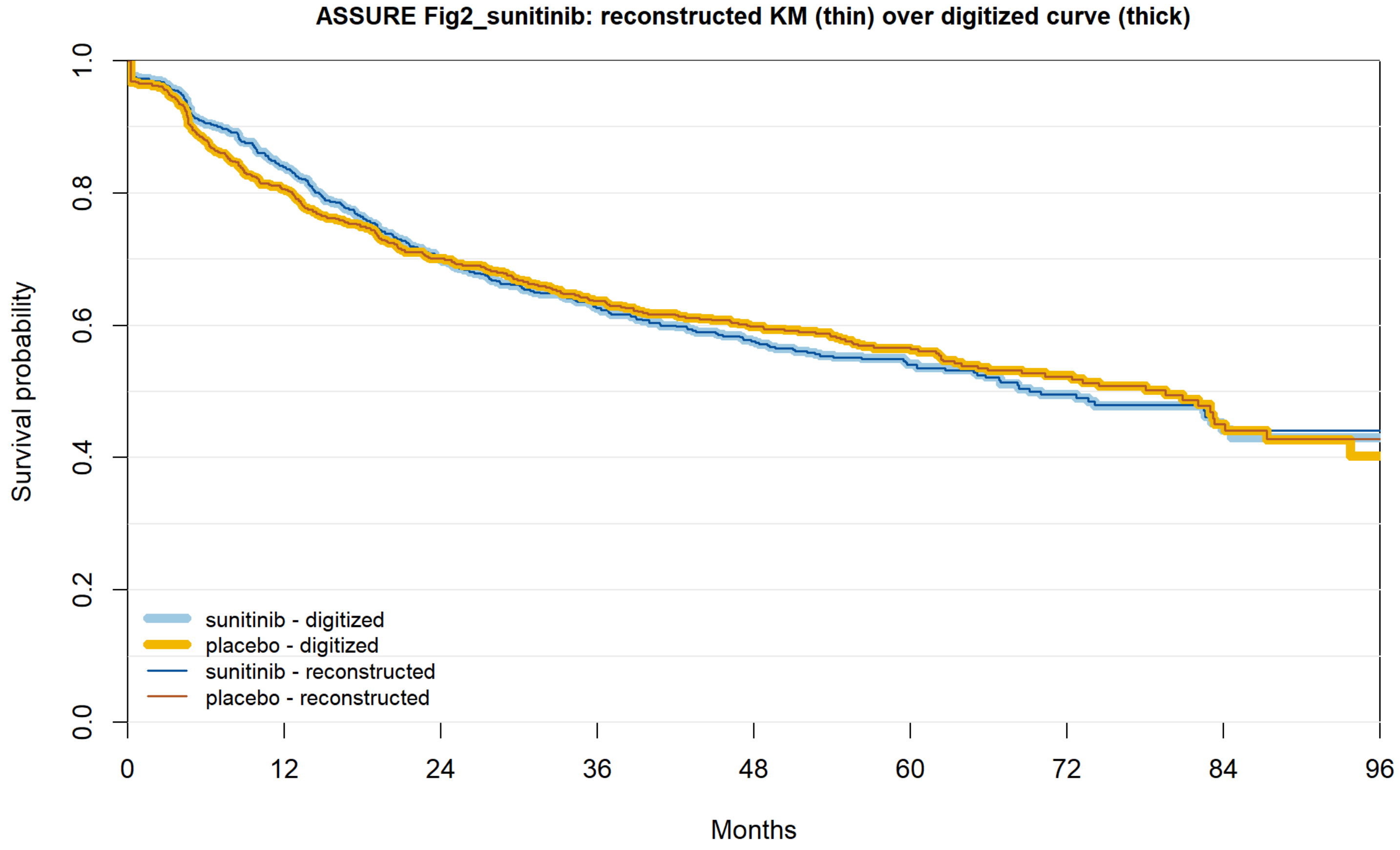


| | 0 | 12 | 24 | 36 | 48 | 60 | 72 | 84 | 96 |
|---|---|---|---|---|---|---|---|---|---|
| sunitinib reported | 647 | 500 | 397 | 338 | 279 | 194 | 102 | 42 | 7 |
| sunitinib recon. | 647 | 500 | 397 | 338 | 279 | 194 | 102 | 42 | 7 |
| placebo reported | 647 | 499 | 414 | 360 | 312 | 200 | 111 | 48 | 7 |
| placebo recon. | 647 | 499 | 414 | 360 | 312 | 200 | 111 | 48 | 7 |

**Figure S10n.** ASSURE DFS, sorafenib vs placebo.

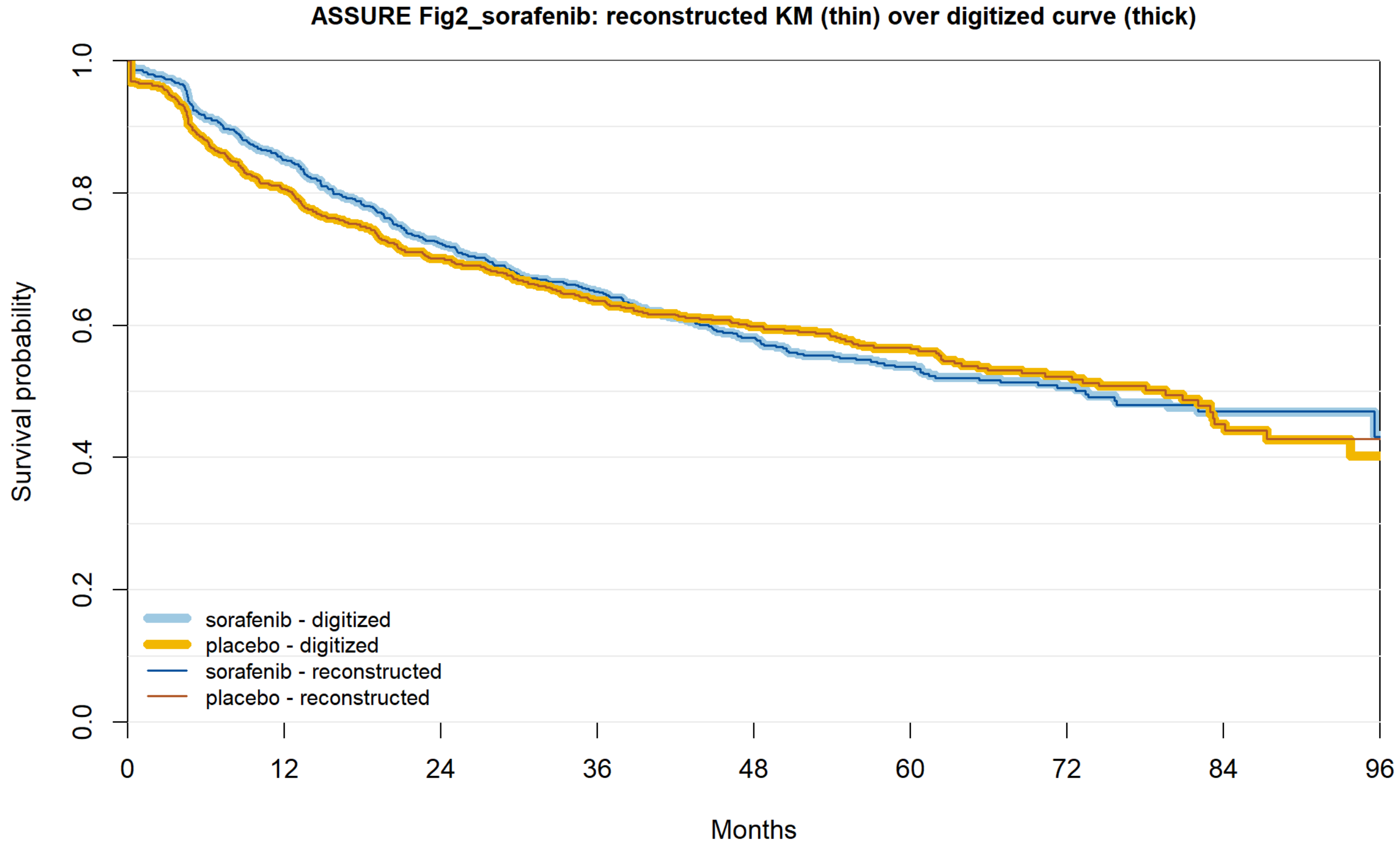


| | 0 | 12 | 24 | 36 | 48 | 60 | 72 | 84 | 96 |
|---|---|---|---|---|---|---|---|---|---|
| sorafenib reported | 649 | 517 | 423 | 357 | 297 | 199 | 114 | 48 | 11 |
| sorafenib recon. | 649 | 517 | 423 | 357 | 297 | 199 | 114 | 48 | 11 |
| placebo reported | 647 | 499 | 414 | 360 | 312 | 200 | 111 | 48 | 7 |
| placebo recon. | 647 | 499 | 414 | 360 | 312 | 200 | 111 | 48 | 7 |

**Figure S10o.** ASSURE OS, sunitinib vs placebo.

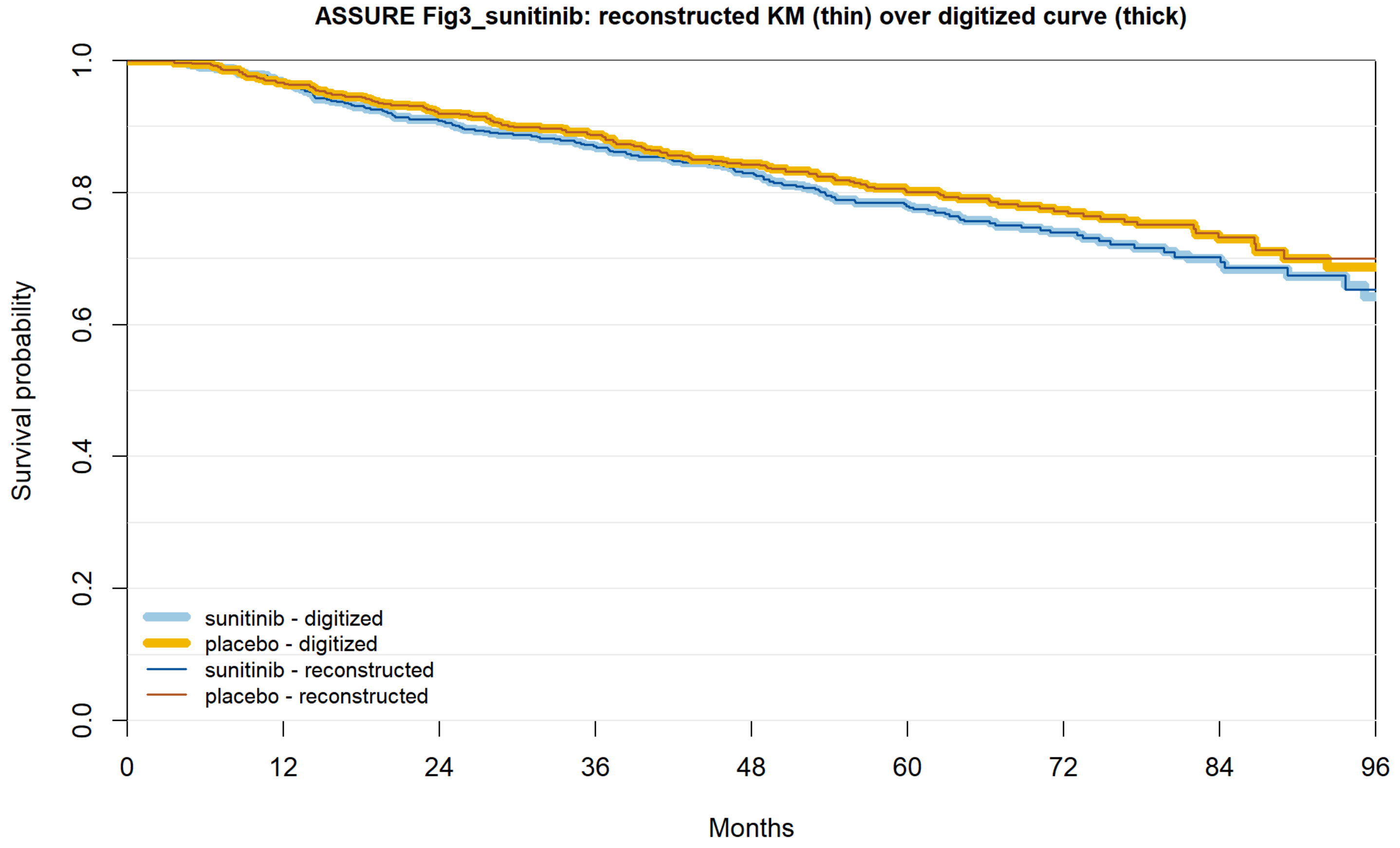


| | | | | | | | | | |
|---|---|---|---|---|---|---|---|---|---|
| sunitinib reported | 647 | 586 | 543 | 503 | 458 | 332 | 190 | 89 | 22 |
| sunitinib recon. | 647 | 586 | 543 | 503 | 458 | 332 | 190 | 89 | 22 |
| placebo reported | 647 | 606 | 569 | 533 | 482 | 349 | 211 | 104 | 22 |
| placebo recon. | 647 | 606 | 569 | 533 | 482 | 349 | 211 | 104 | 22 |

**Figure S10p.** ASSURE OS, sorafenib vs placebo.

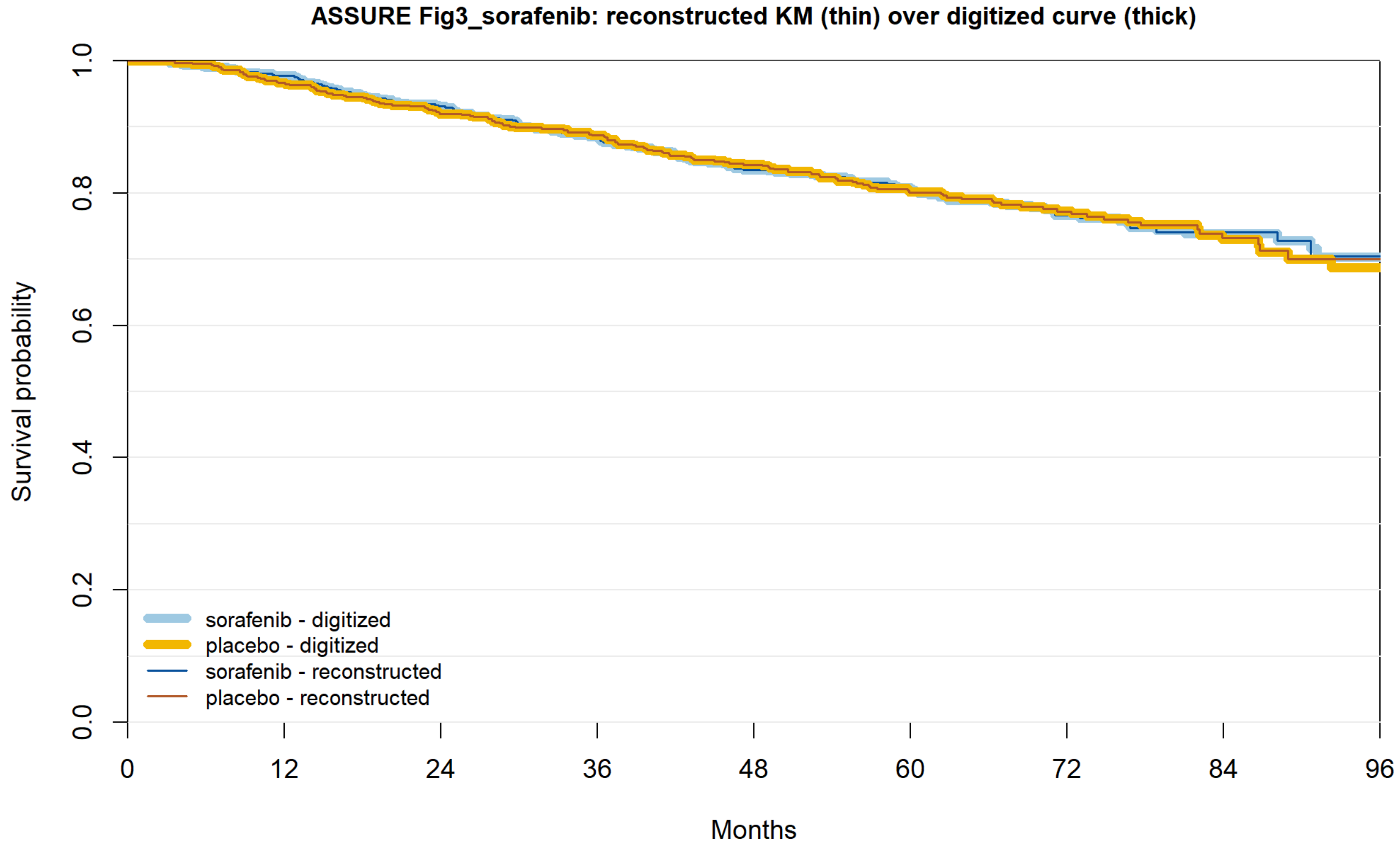


| | | | | | | | | | |
|---|---|---|---|---|---|---|---|---|---|
| sorafenib reported | 649 | 597 | 562 | 514 | 474 | 353 | 203 | 97 | 26 |
| sorafenib recon. | 649 | 597 | 562 | 514 | 474 | 353 | 203 | 97 | 26 |
| placebo reported | 647 | 606 | 569 | 533 | 482 | 349 | 211 | 104 | 22 |
| placebo recon. | 647 | 606 | 569 | 533 | 482 | 349 | 211 | 104 | 22 |

**Figure S10q.** PROTECT.

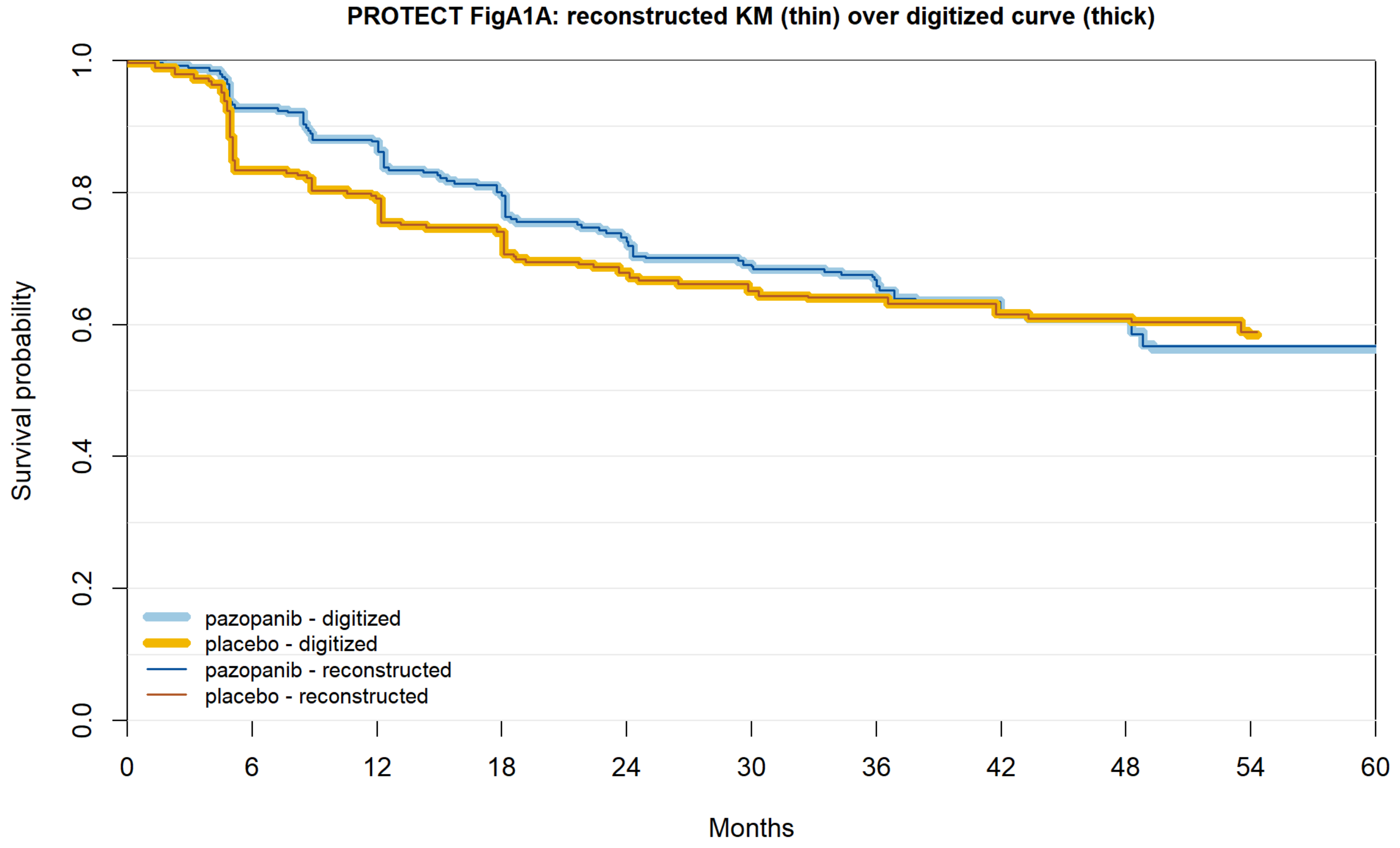


| | 0 | 6 | 12 | 18 | 24 | 30 | 36 | 42 | 48 | 54 | 60 |
|---|---|---|---|---|---|---|---|---|---|---|---|
| pazopanib reported | 571 | 482 | 423 | 383 | 346 | 330 | 282 | 194 | 115 | 30 | 0 |
| pazopanib recon. | 571 | 482 | 423 | 382 | 346 | 328 | 282 | 194 | 115 | 30 | 0 |
| placebo reported | 564 | 443 | 396 | 375 | 348 | 333 | 282 | 191 | 107 | 36 | 0 |
| placebo recon. | 564 | 443 | 396 | 371 | 340 | 326 | 282 | 191 | 107 | 36 | 0 |

**Figure S10r.** ATLAS.

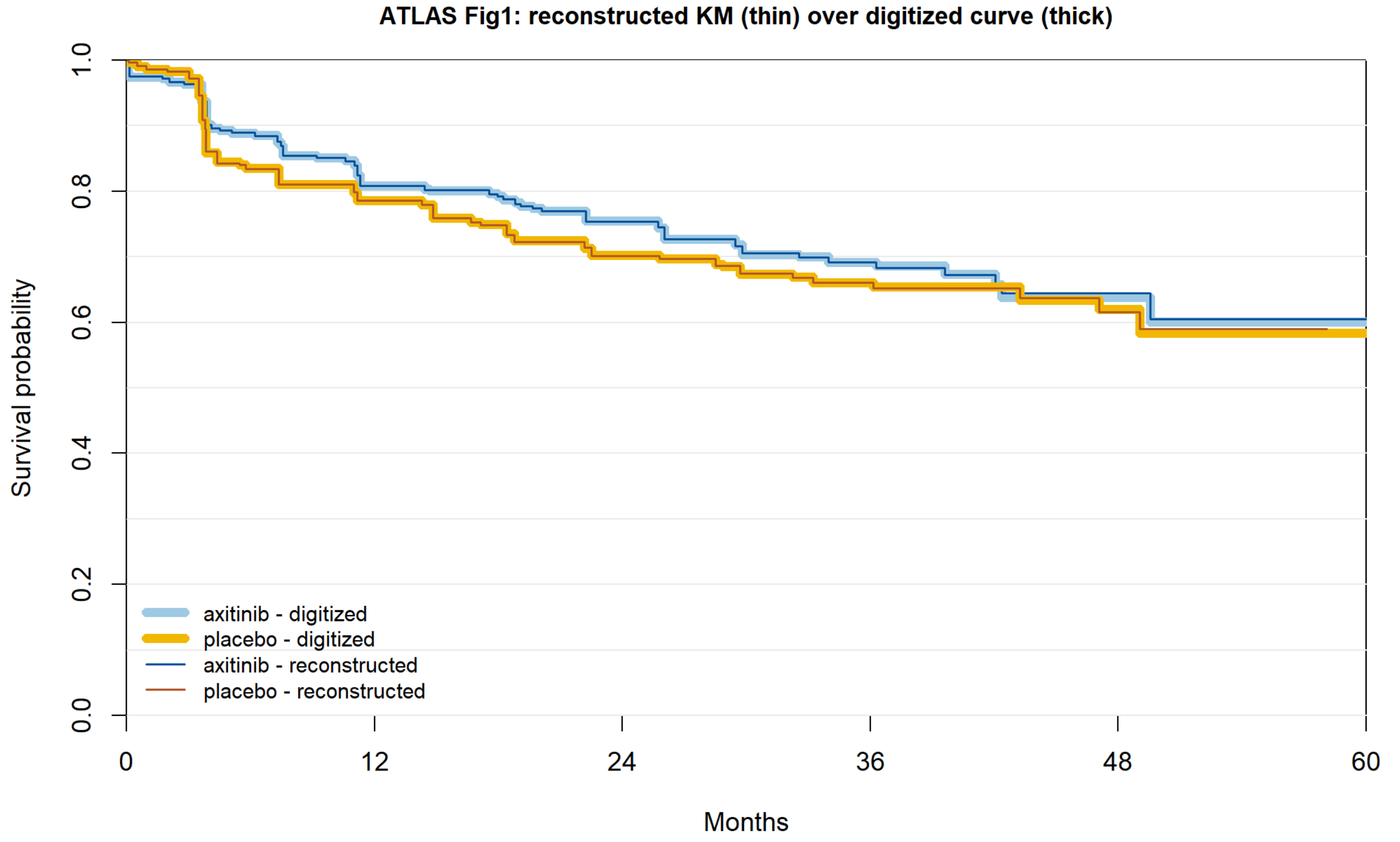


| | | | | | | |
|---|---|---|---|---|---|---|
| axitinib reported | 363 | 264 | 182 | 81 | 20 | 1 |
| axitinib recon. | 363 | 264 | 182 | 81 | 20 | 1 |
| placebo reported | 361 | 252 | 164 | 72 | 26 | 0 |
| placebo recon. | 361 | 252 | 164 | 72 | 26 | 0 |

**Figure S10s.** SORCE 1 year.

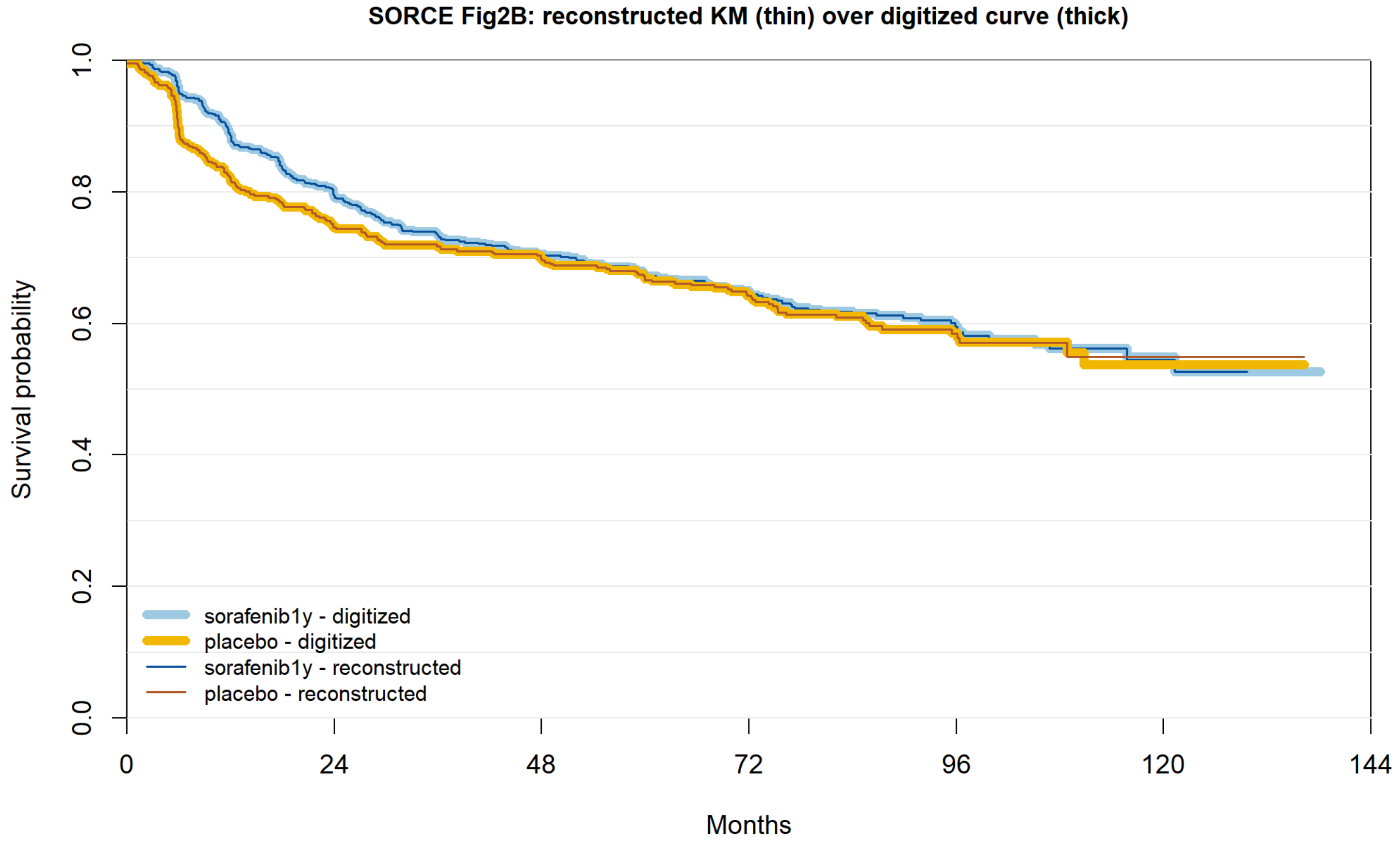


| | 0 | 24 | 48 | 72 | 96 | 120 | 144 |
|---|---|---|---|---|---|---|---|
| sorafenib1y reported | 642 | 495 | 422 | 300 | 131 | 32 | 0 |
| sorafenib1y recon. | 642 | 495 | 422 | 300 | 131 | 32 | 0 |
| placebo reported | 430 | 313 | 282 | 207 | 87 | 17 | 0 |
| placebo recon. | 430 | 313 | 282 | 207 | 87 | 17 | 0 |

**Figure S10t.** SORCE 3 years.

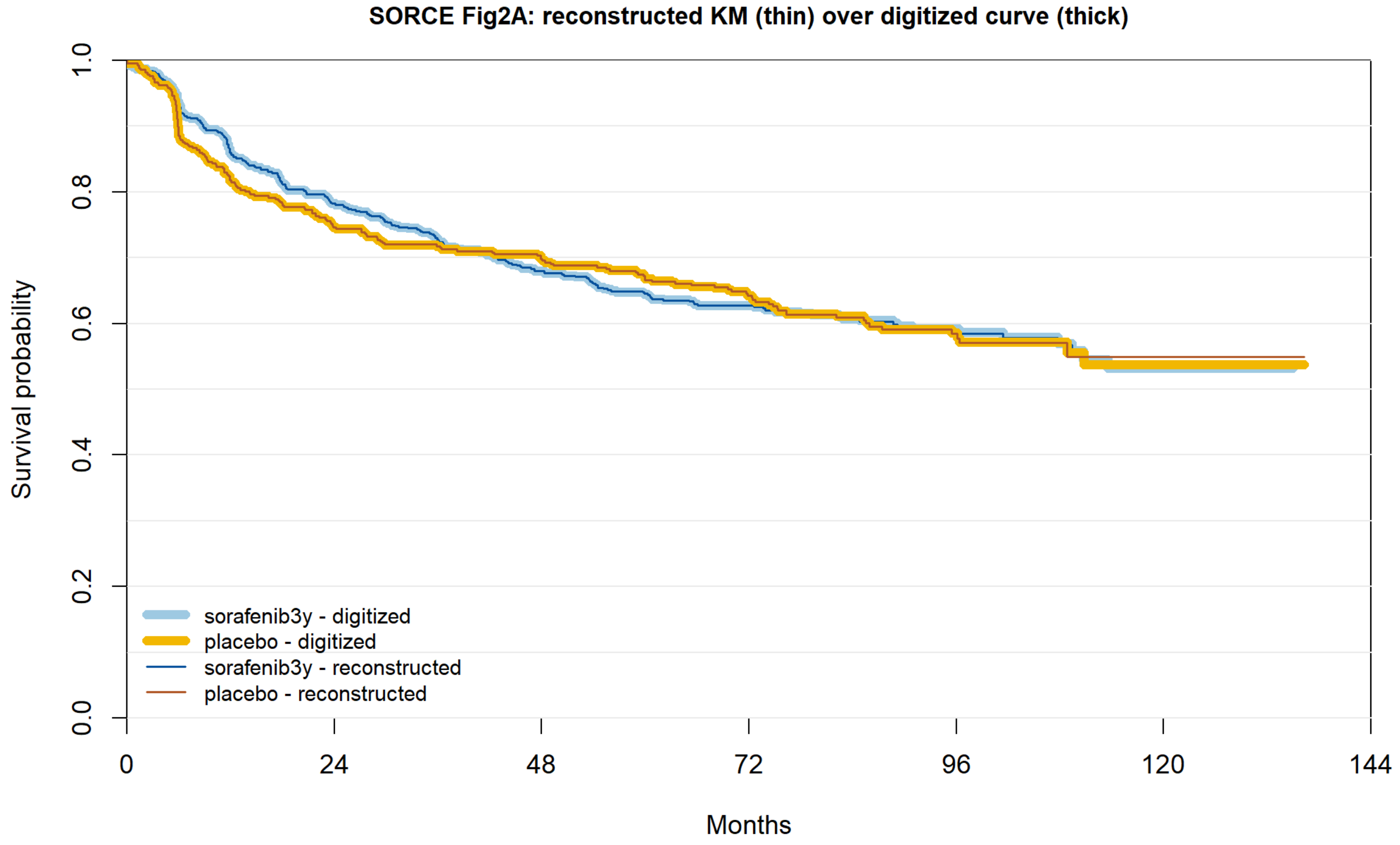


| | | | | | | | |
|---|---|---|---|---|---|---|---|
| sorafenib3y reported | 639 | 484 | 399 | 297 | 118 | 25 | 0 |
| sorafenib3y recon. | 639 | 484 | 399 | 297 | 118 | 25 | 0 |
| placebo reported | 430 | 313 | 282 | 207 | 87 | 17 | 0 |
| placebo recon. | 430 | 313 | 282 | 207 | 87 | 17 | 0 |

**Figure S10u.** PROSPER, standard.

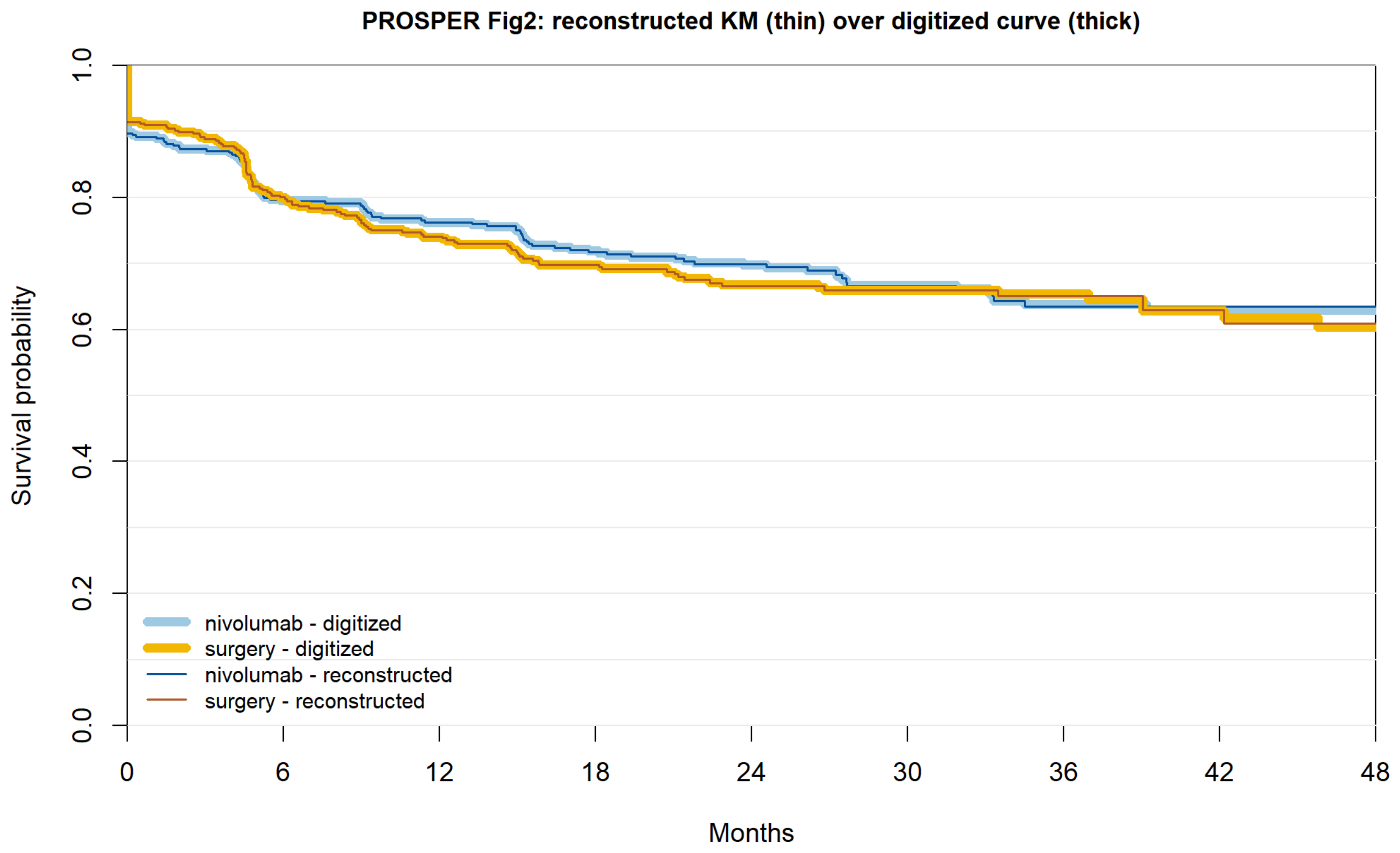


| | 0 | 6 | 12 | 18 | 24 | 30 | 36 | 42 | 48 |
|---|---|---|---|---|---|---|---|---|---|
| nivolumab reported | 381 | 286 | 264 | 232 | 149 | 111 | 76 | 42 | 25 |
| nivolumab recon. | 381 | 286 | 264 | 232 | 149 | 111 | 76 | 42 | 25 |
| surgery reported | 399 | 292 | 256 | 218 | 136 | 110 | 74 | 30 | 16 |
| surgery recon. | 399 | 292 | 256 | 218 | 136 | 110 | 74 | 30 | 16 |

**Figure S10v.** PROSPER, censoring-informed.

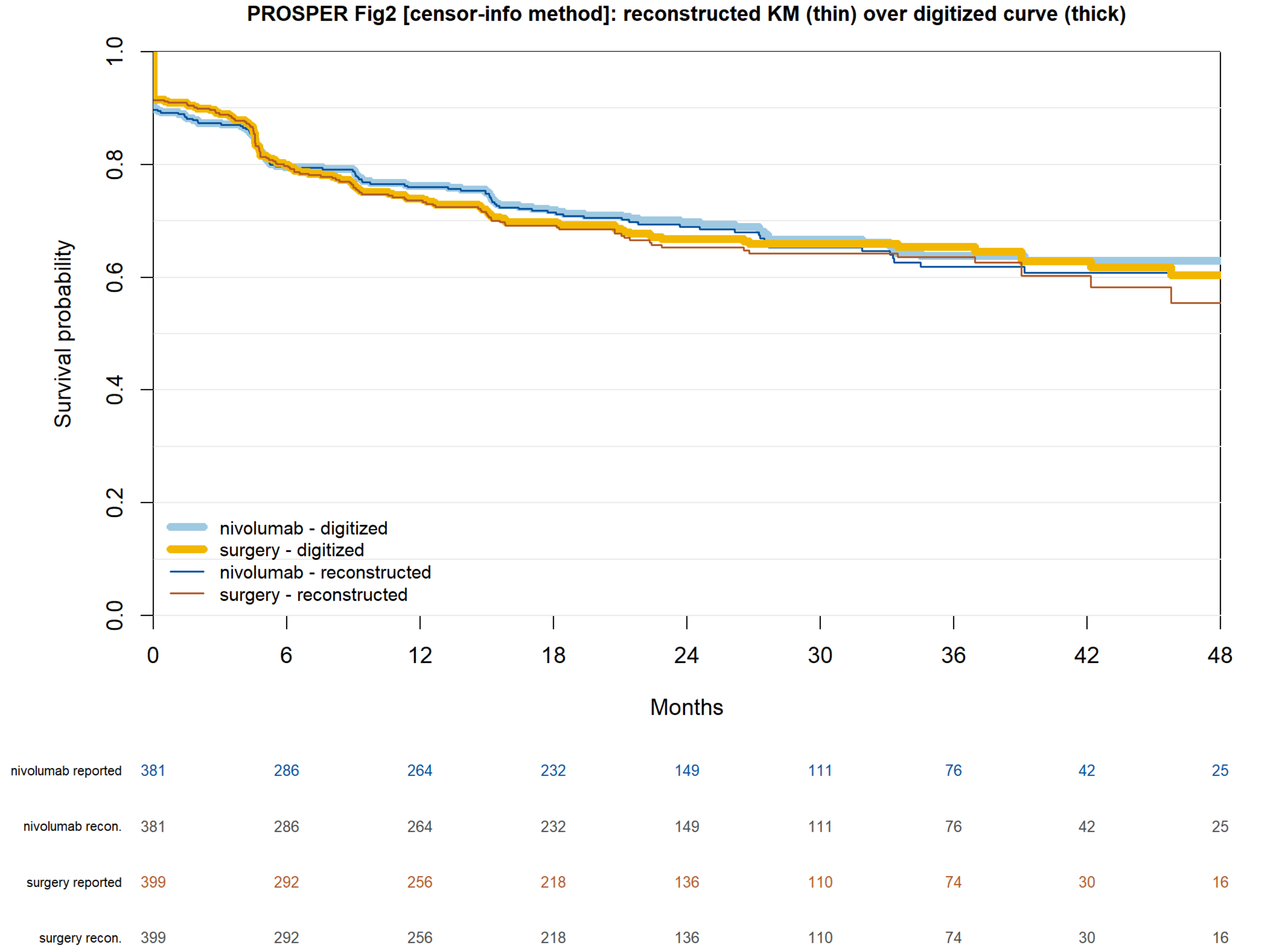

**Figure S10u2.** E2810 DFS (D9).

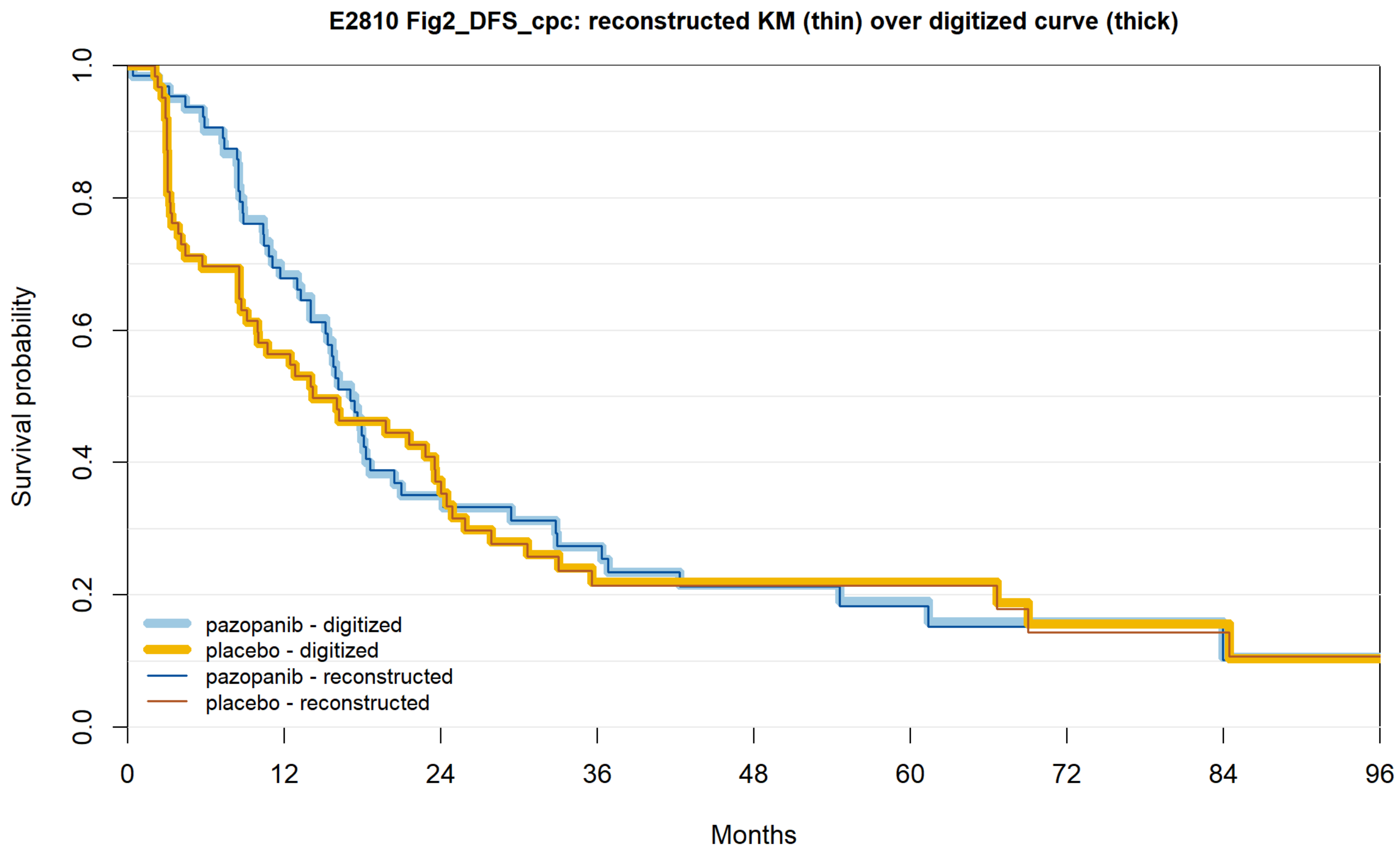


| | | | | | | | | | |
|---|---|---|---|---|---|---|---|---|---|
| pazopanib reported | 66 | 41 | 19 | 14 | 10 | 6 | 4 | 2 | 1 |
| pazopanib recon. | 66 | 41 | 19 | 14 | 10 | 6 | 4 | 2 | 1 |
| placebo reported | 63 | 34 | 20 | 10 | 8 | 7 | 4 | 4 | 1 |
| placebo recon. | 63 | 34 | 20 | 10 | 8 | 7 | 4 | 4 | 1 |

**Figure S10v2.** E2810 OS (D9; OS accepted after adjudication: reported HR 2.55 vs HR 2.11 derived exactly from the curve; reported model not stated — see Table S1, D9).

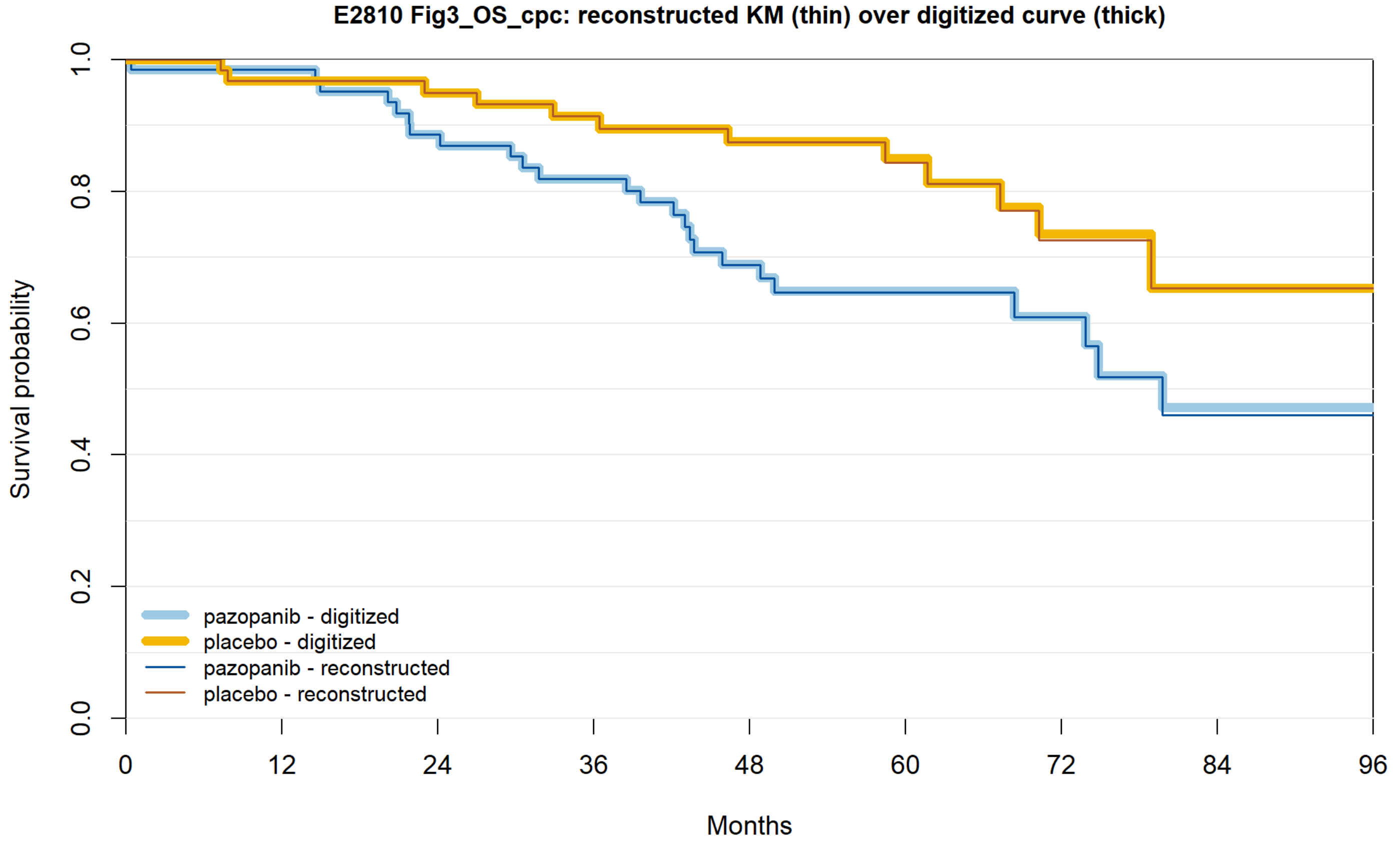


| | 0 | 12 | 24 | 36 | 48 | 60 | 72 | 84 | 96 |
|---|---|---|---|---|---|---|---|---|---|
| pazopanib reported | 66 | 60 | 54 | 48 | 34 | 23 | 14 | 6 | 1 |
| pazopanib recon. | 66 | 60 | 54 | 48 | 34 | 23 | 14 | 6 | 1 |
| placebo reported | 63 | 60 | 54 | 49 | 42 | 27 | 15 | 6 | 1 |
| placebo recon. | 63 | 60 | 54 | 49 | 42 | 27 | 15 | 6 | 1 |

Curves added under deviation D8 (Figures S10w–S10ar):

**Figure S10w.** KEYNOTE-564 NEJM 2021.

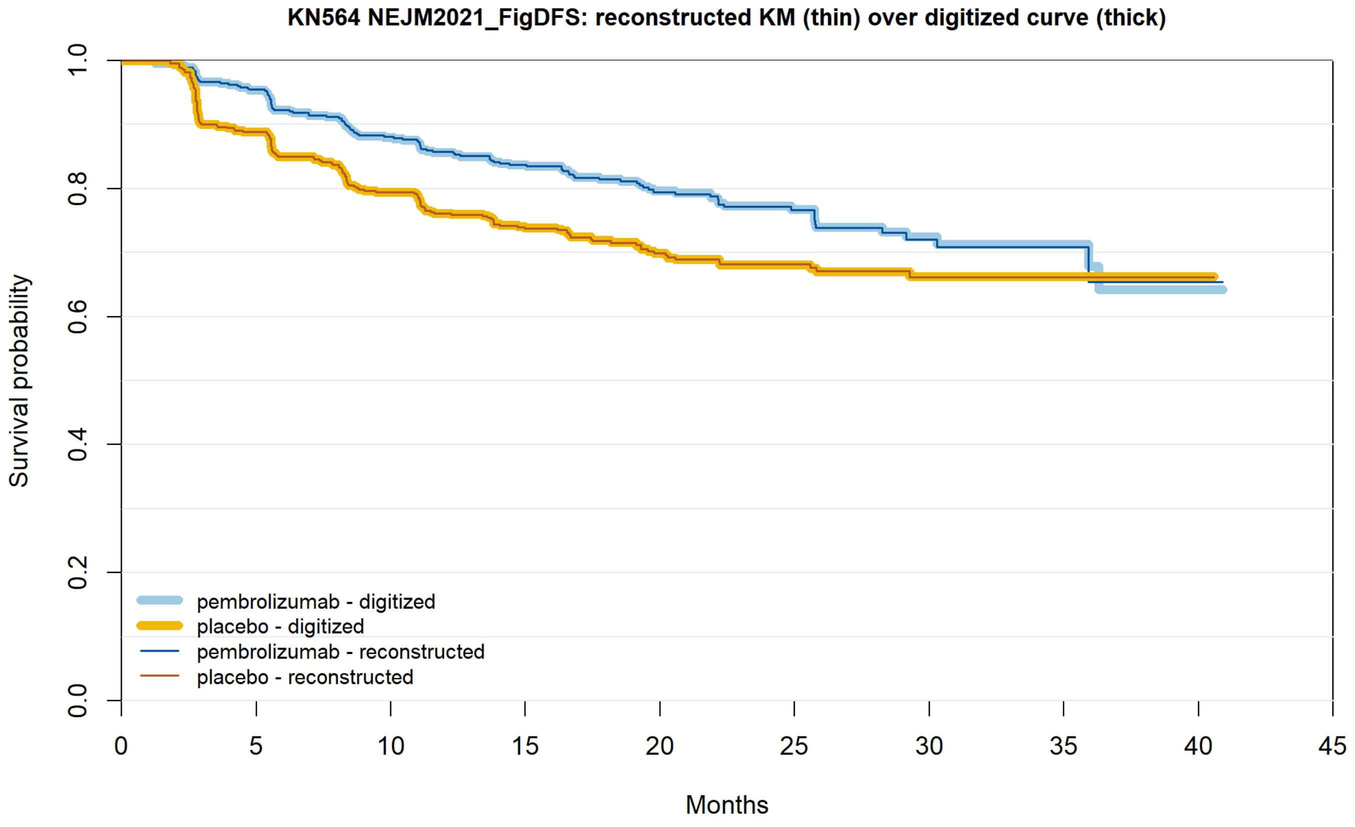


| | | | | | | | | | | |
|---|---|---|---|---|---|---|---|---|---|---|
| pembrolizumab reported | 496 | 457 | 414 | 371 | 233 | 151 | 61 | 21 | 1 | 0 |
| pembrolizumab recon. | 496 | 457 | 414 | 371 | 233 | 151 | 61 | 21 | 1 | 0 |
| placebo reported | 498 | 436 | 389 | 341 | 209 | 145 | 56 | 19 | 1 | 0 |
| placebo recon. | 498 | 436 | 389 | 341 | 209 | 145 | 56 | 19 | 1 | 0 |

**Figure S10x.** KEYNOTE-564 Lancet Oncol 2022.

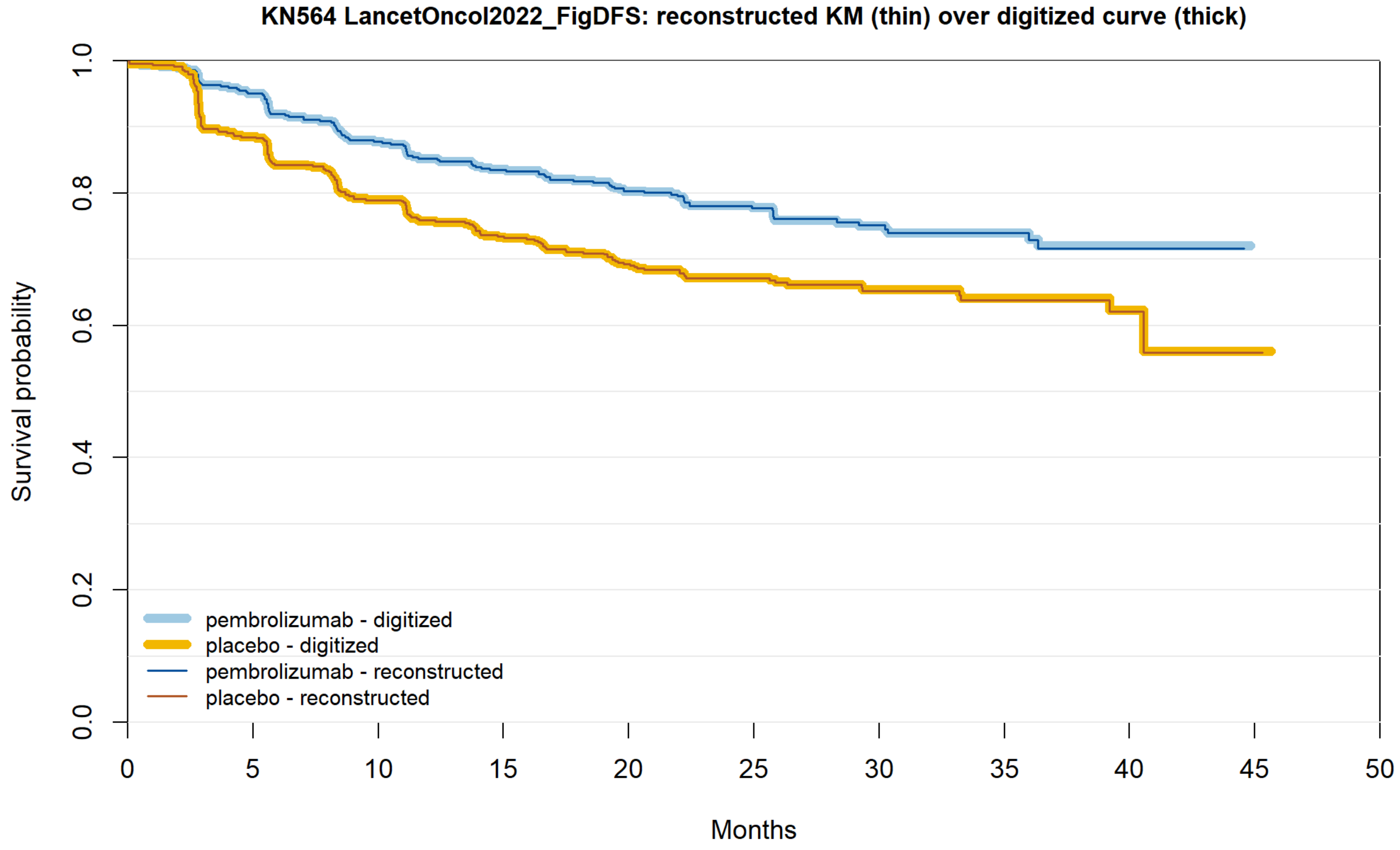


| | | | | | | | | | | | |
|---|---|---|---|---|---|---|---|---|---|---|---|
| pembrolizumab reported | 496 | 458 | 416 | 398 | 361 | 255 | 135 | 77 | 37 | 0 | 0 |
| pembrolizumab recon. | 496 | 458 | 416 | 396 | 361 | 255 | 135 | 77 | 37 | 0 | 0 |
| placebo reported | 498 | 437 | 389 | 356 | 325 | 230 | 125 | 74 | 33 | 1 | 0 |
| placebo recon. | 498 | 437 | 389 | 356 | 325 | 230 | 125 | 74 | 33 | 1 | 0 |

**Figure S10y.** KEYNOTE-564 NEJM 2024.

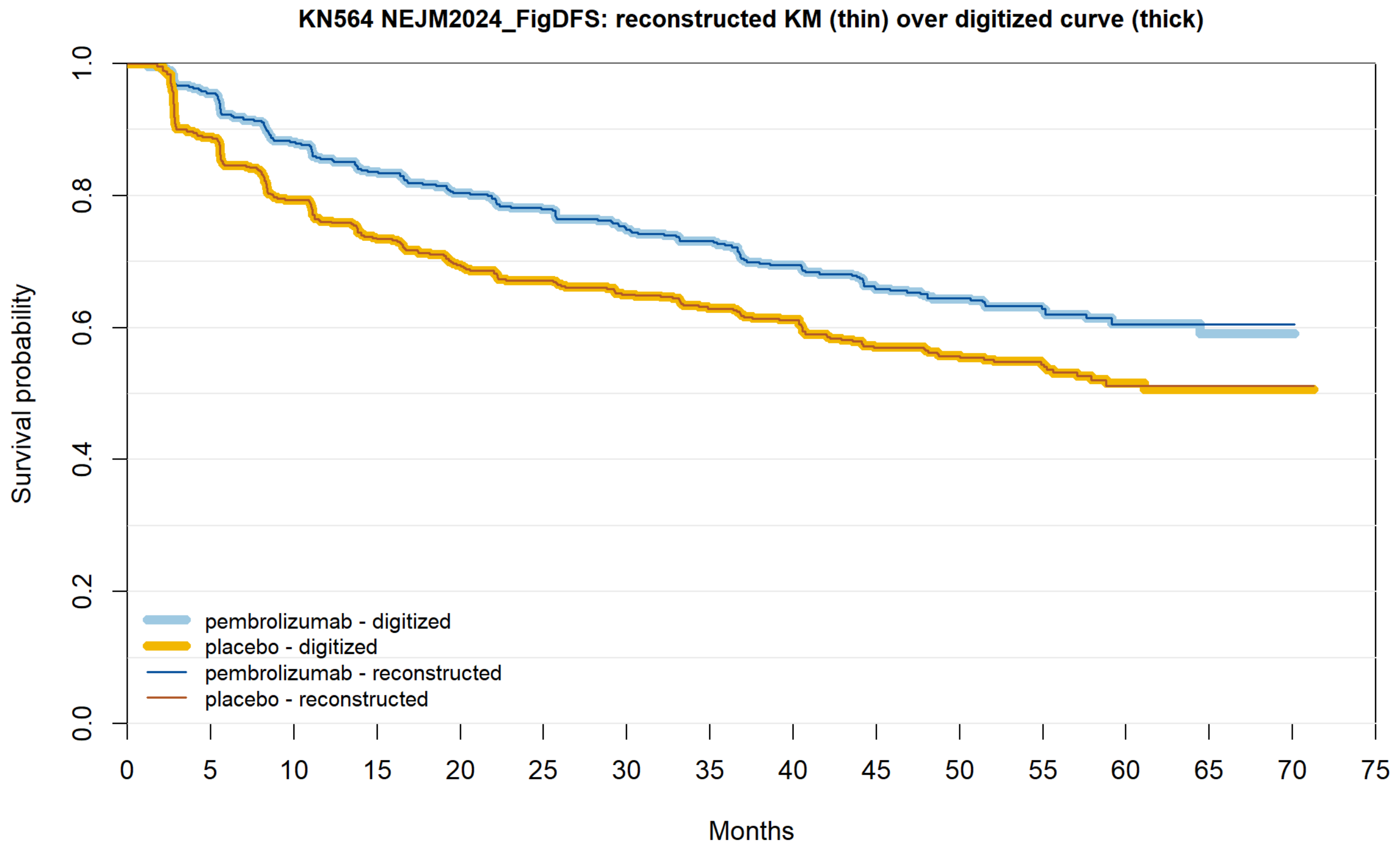


| | | | | | | | | | | | | | | | | |
|---|---|---|---|---|---|---|---|---|---|---|---|---|---|---|---|---|
| pembrolizumab reported | 496 | 458 | 416 | 388 | 370 | 355 | 337 | 327 | 307 | 284 | 221 | 160 | 65 | 19 | 5 | 0 |
| pembrolizumab recon. | 496 | 458 | 416 | 388 | 370 | 355 | 337 | 327 | 307 | 284 | 221 | 160 | 65 | 19 | 5 | 0 |
| placebo reported | 498 | 438 | 390 | 357 | 333 | 320 | 307 | 292 | 282 | 254 | 210 | 139 | 62 | 16 | 2 | 0 |
| placebo recon. | 498 | 438 | 390 | 357 | 333 | 320 | 307 | 292 | 282 | 254 | 210 | 139 | 62 | 16 | 2 | 0 |

**Figure S10z.** PROSPER Supplemental Fig 3.

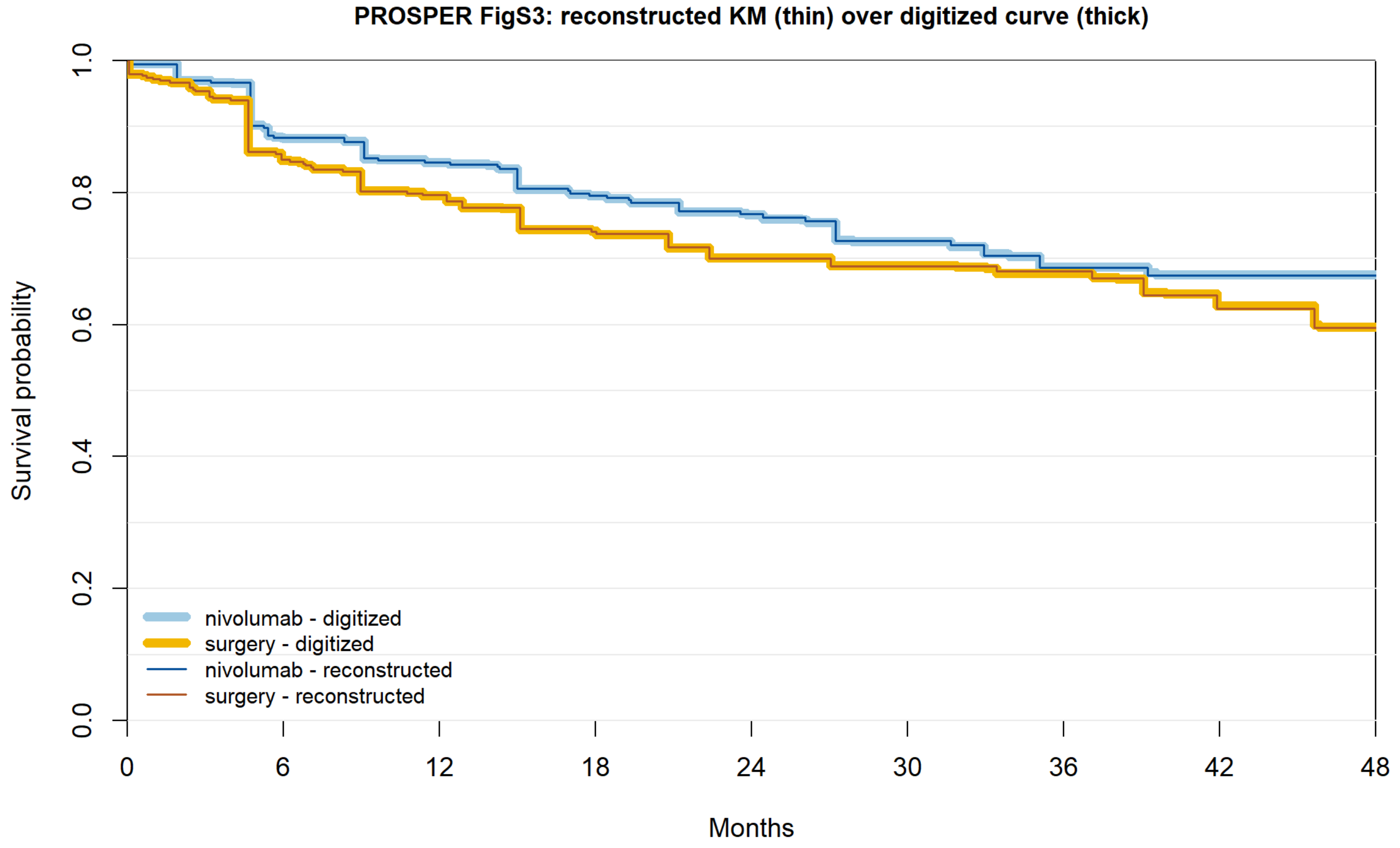


| | 0 | 6 | 12 | 18 | 24 | 30 | 36 | 42 | 48 |
|---|---|---|---|---|---|---|---|---|---|
| nivolumab reported | 381 | 286 | 264 | 232 | 149 | 111 | 76 | 42 | 25 |
| nivolumab recon. | 381 | 286 | 264 | 232 | 149 | 111 | 76 | 42 | 25 |
| surgery reported | 399 | 292 | 256 | 218 | 136 | 110 | 74 | 30 | 16 |
| surgery recon. | 399 | 292 | 256 | 218 | 136 | 110 | 74 | 30 | 16 |

**Figure S10aa.** ASSURE clear cell, sunitinib vs placebo.

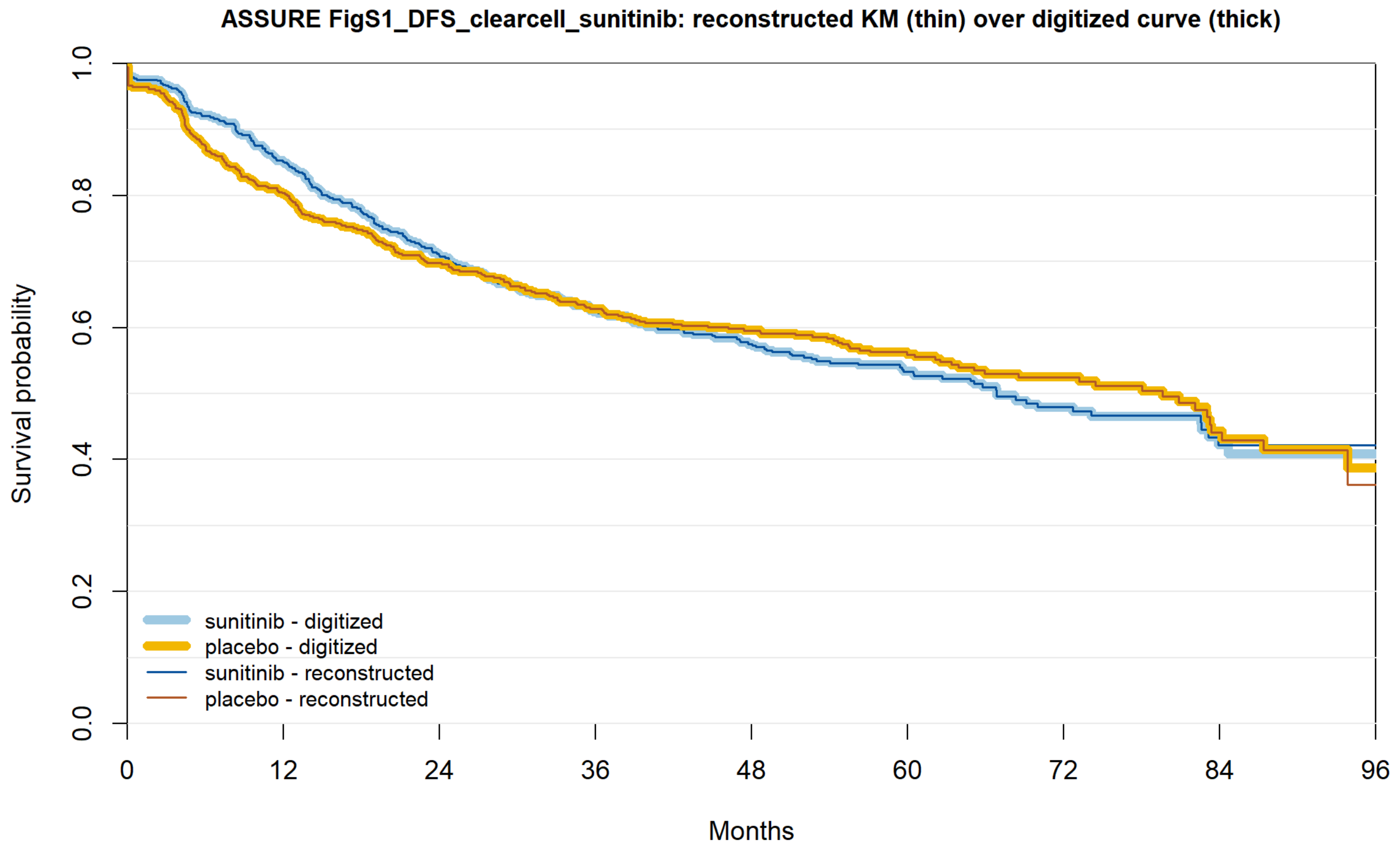


| | | | | | | | | | |
|---|---|---|---|---|---|---|---|---|---|
| sunitinib reported | 542 | 422 | 335 | 277 | 233 | 160 | 82 | 35 | 5 |
| sunitinib recon. | 542 | 422 | 335 | 277 | 233 | 160 | 82 | 35 | 5 |
| placebo reported | 540 | 413 | 340 | 293 | 258 | 161 | 91 | 38 | 7 |
| placebo recon. | 540 | 413 | 340 | 293 | 258 | 161 | 91 | 38 | 7 |

**Figure S10ab.** ASSURE clear cell, sorafenib vs placebo.

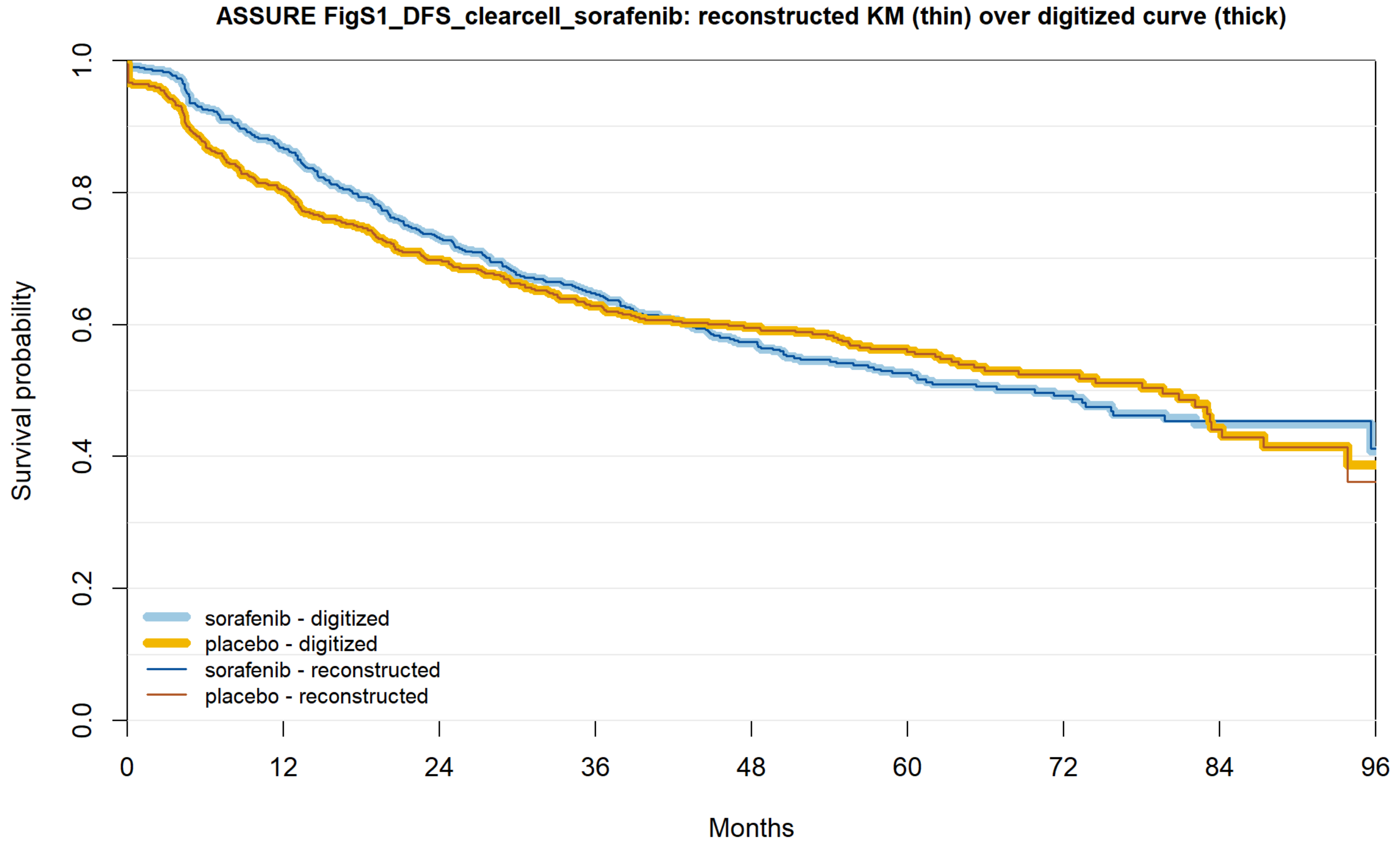


| | 0 | 12 | 24 | 36 | 48 | 60 | 72 | 84 | 96 |
|---|---|---|---|---|---|---|---|---|---|
| sorafenib reported | 540 | 440 | 356 | 298 | 246 | 166 | 96 | 39 | 10 |
| sorafenib recon. | 540 | 440 | 356 | 298 | 246 | 166 | 96 | 39 | 10 |
| placebo reported | 540 | 413 | 340 | 293 | 258 | 161 | 91 | 38 | 7 |
| placebo recon. | 540 | 413 | 340 | 293 | 258 | 161 | 91 | 38 | 7 |

**Figure S10ac.** PROTECT ITT800mg.

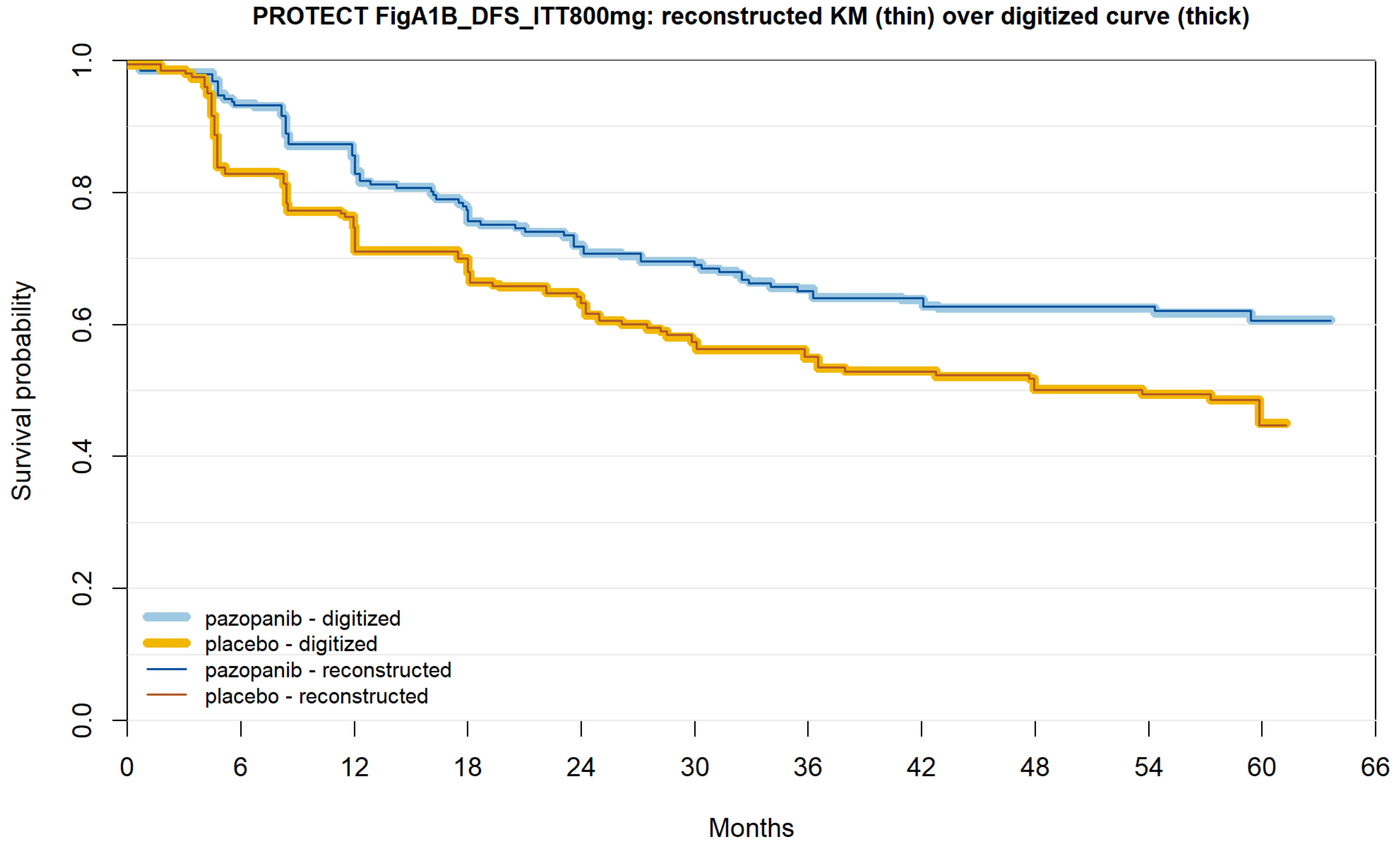


| | | | | | | | | | | | |
|---|---|---|---|---|---|---|---|---|---|---|---|
| pazopanib reported | 198 | 176 | 156 | 141 | 129 | 124 | 114 | 107 | 100 | 91 | 42 | 0 |
| pazopanib recon. | 198 | 176 | 156 | 140 | 129 | 124 | 114 | 107 | 100 | 91 | 42 | 0 |
| placebo reported | 205 | 169 | 144 | 134 | 119 | 107 | 99 | 94 | 89 | 76 | 35 | 0 |
| placebo recon. | 205 | 169 | 144 | 134 | 119 | 107 | 99 | 94 | 89 | 76 | 35 | 0 |

**Figure S10ad.** PROTECT ITTAll.

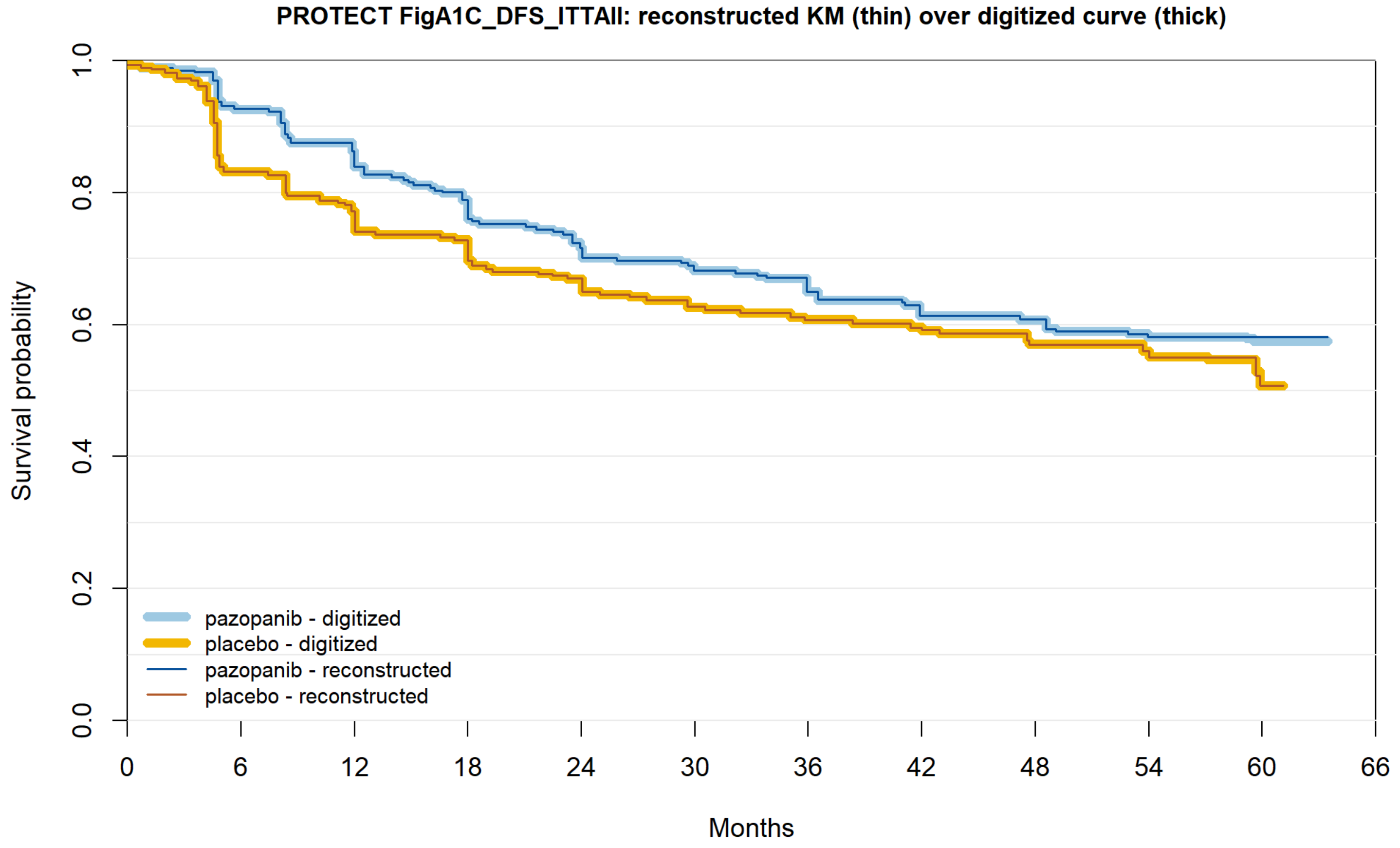


| | | | | | | | | | | | | |
|---|---|---|---|---|---|---|---|---|---|---|---|---|
| pazopanib reported | 769 | 658 | 579 | 524 | 475 | 454 | 396 | 301 | 215 | 121 | 42 | 0 |
| pazopanib recon. | 769 | 658 | 579 | 524 | 475 | 452 | 396 | 301 | 215 | 121 | 42 | 0 |
| placebo reported | 769 | 612 | 540 | 509 | 467 | 440 | 381 | 285 | 196 | 112 | 35 | 0 |
| placebo recon. | 769 | 612 | 540 | 488 | 467 | 437 | 381 | 285 | 196 | 112 | 35 | 0 |

**Figure S10ae.** ATLAS highest risk, IRC.

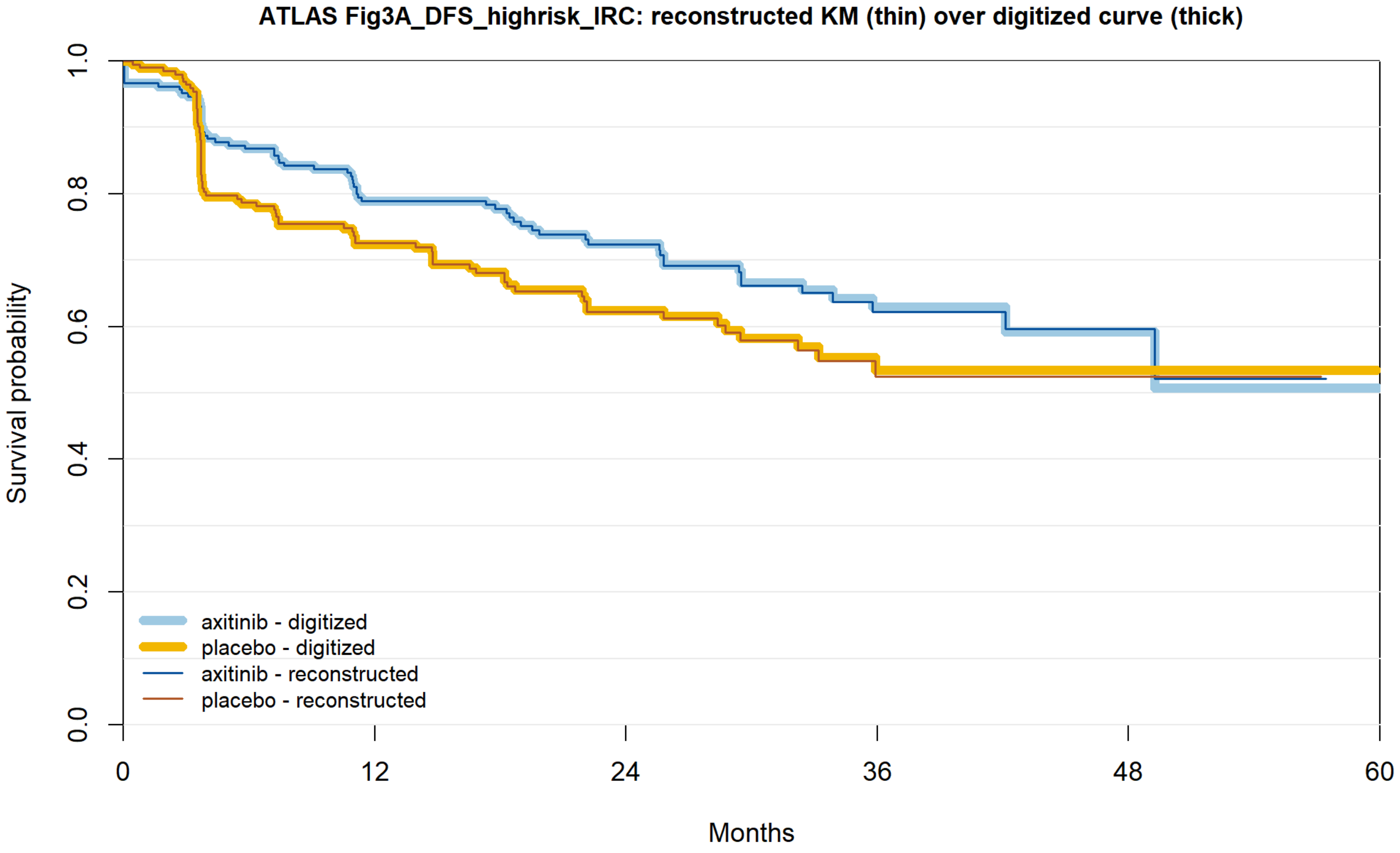


| | | | | | | |
|---|---|---|---|---|---|---|
| axitinib reported | 209 | 149 | 96 | 40 | 8 | 0 |
| axitinib recon. | 209 | 149 | 96 | 40 | 8 | 0 |
| placebo reported | 200 | 124 | 74 | 22 | 7 | 0 |
| placebo recon. | 200 | 124 | 74 | 22 | 7 | 0 |

**Figure S10af.** ATLAS highest risk, investigator.

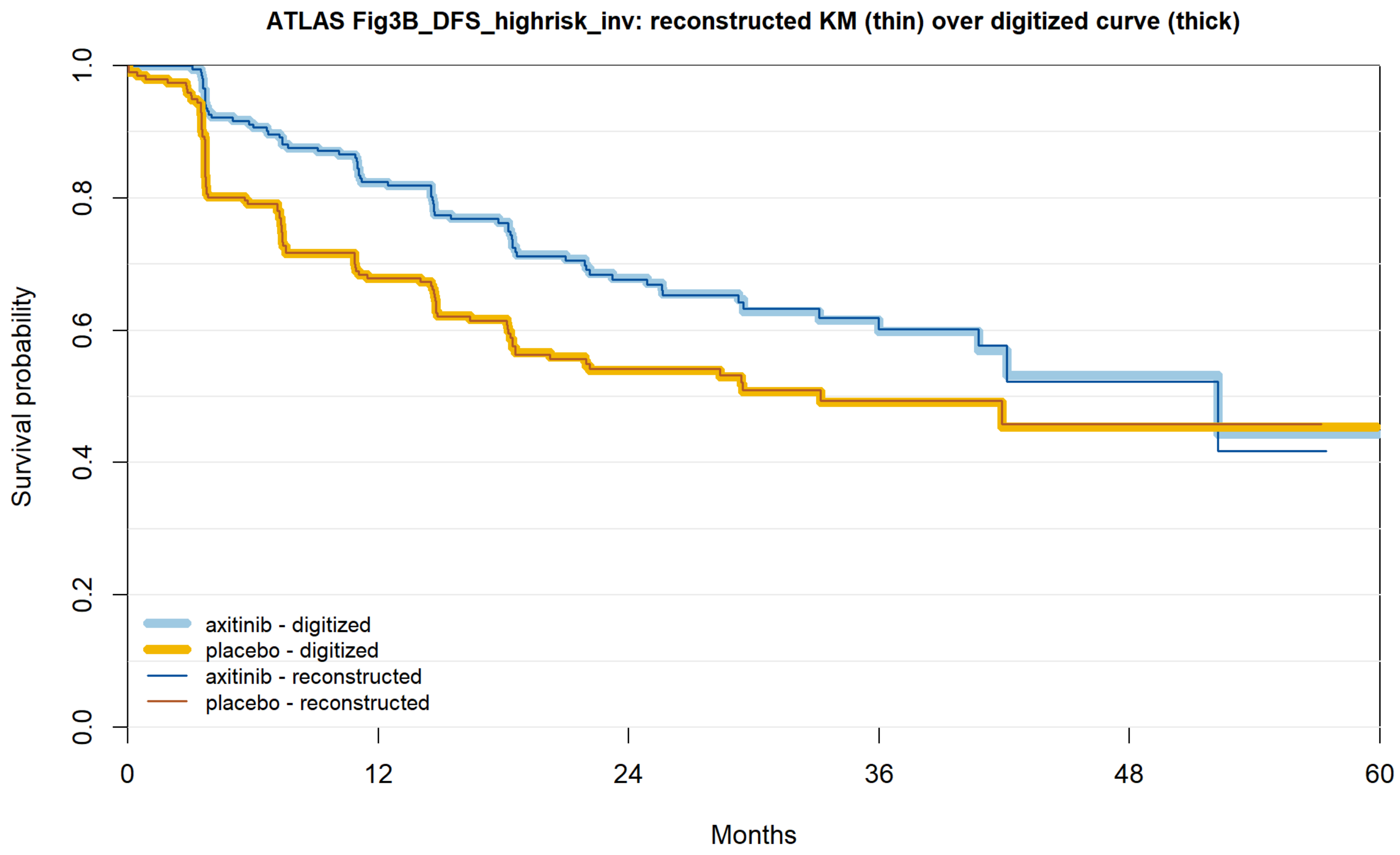


| | | | | | | |
|---|---|---|---|---|---|---|
| axitinib reported | 209 | 158 | 90 | 34 | 7 | 0 |
| axitinib recon. | 209 | 158 | 90 | 34 | 7 | 0 |
| placebo reported | 200 | 124 | 70 | 20 | 7 | 0 |
| placebo recon. | 200 | 124 | 70 | 20 | 7 | 0 |

**Figure S10ag.** ATLAS ITT, investigator.

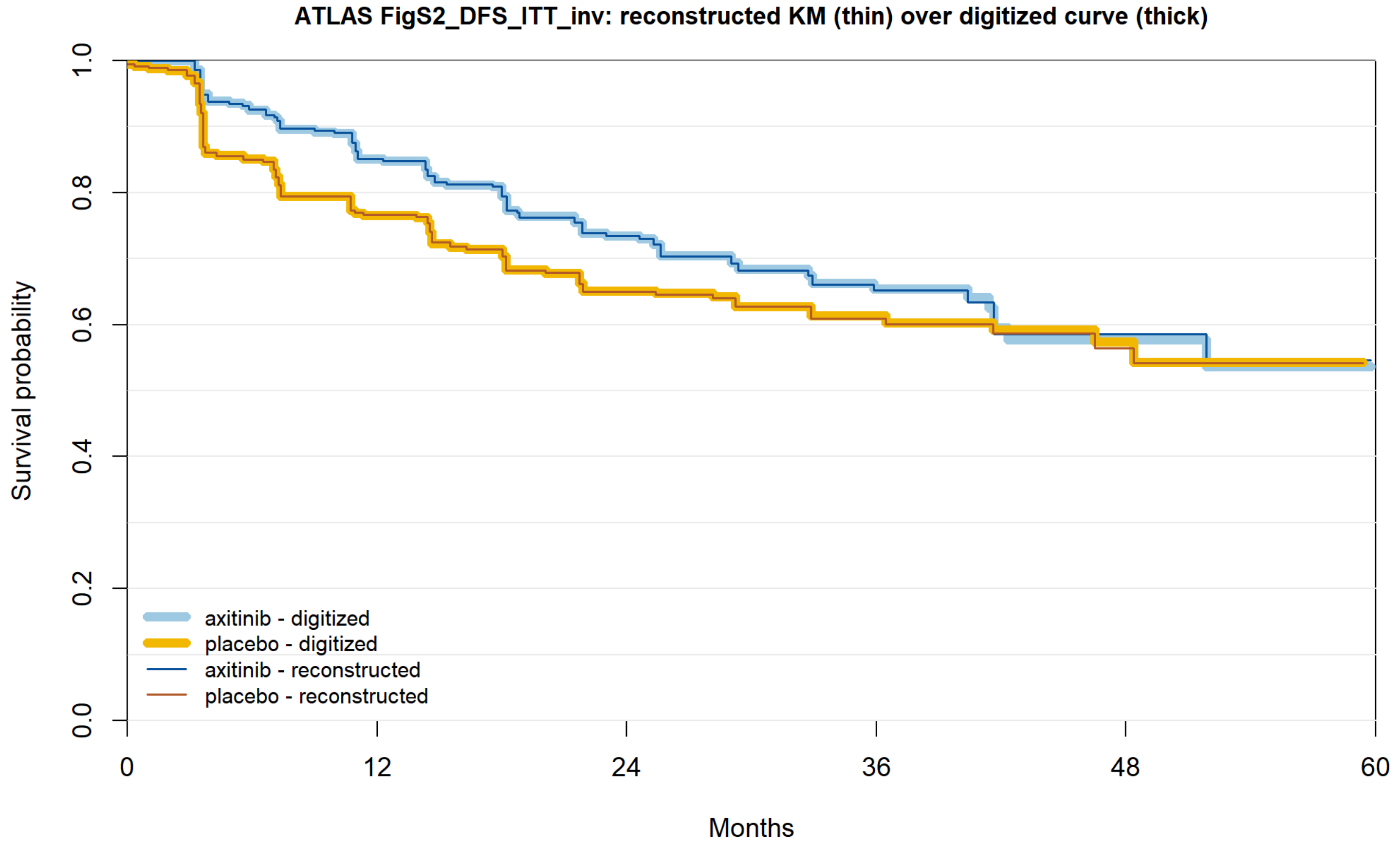


| | | | | | | |
|---|---|---|---|---|---|---|
| axitinib reported | 363 | 280 | 175 | 73 | 19 | 1 |
| axitinib recon. | 363 | 280 | 175 | 73 | 19 | 0 |
| placebo reported | 361 | 252 | 159 | 67 | 25 | 0 |
| placebo recon. | 361 | 252 | 159 | 67 | 25 | 0 |

**Figure S10ah.** SORCE Leibovich high risk, 1 year.

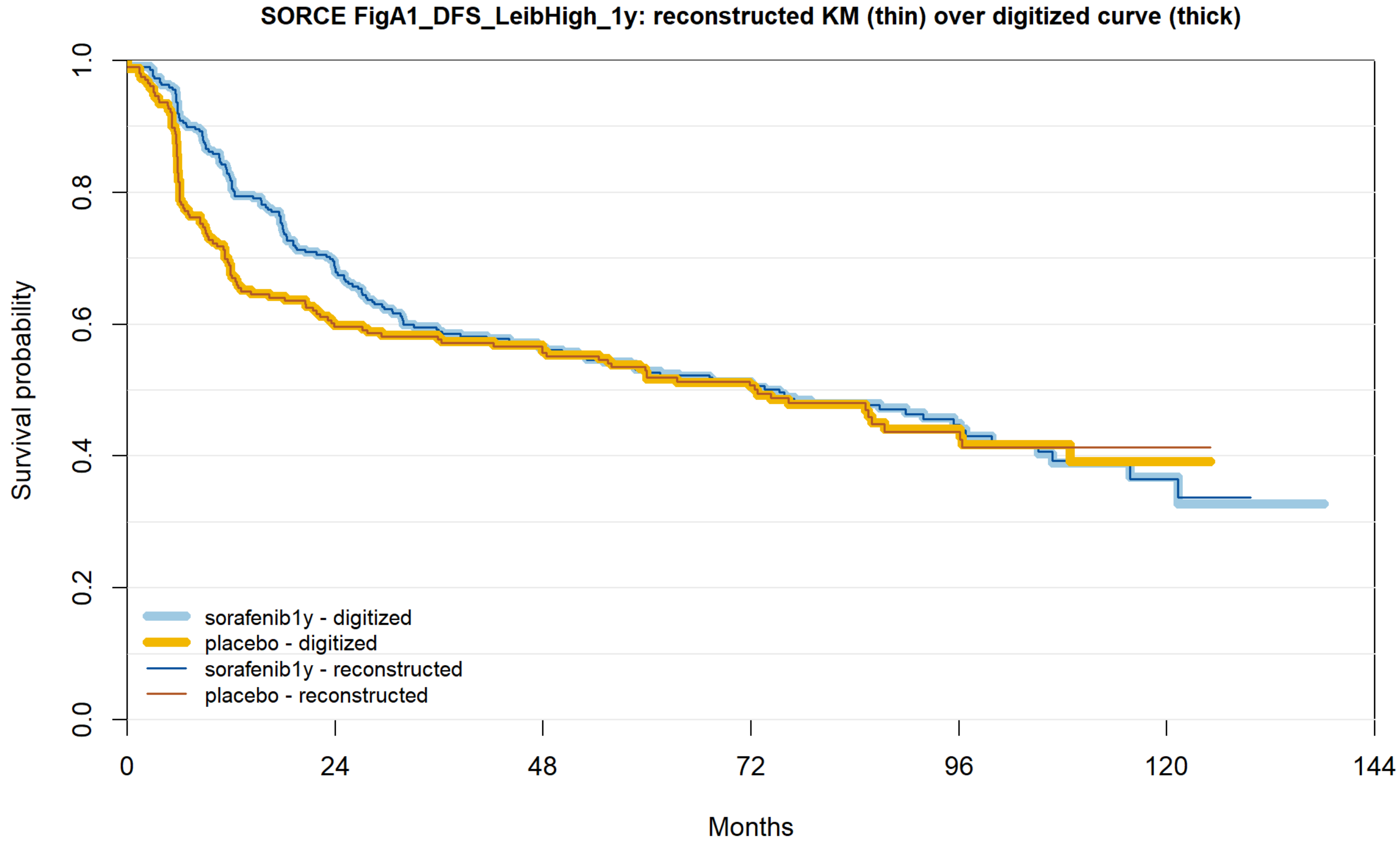


| | | | | | | | |
|---|---|---|---|---|---|---|---|
| sorafenib1y reported | 300 | 199 | 160 | 116 | 51 | 13 | 0 |
| sorafenib1y recon. | 300 | 199 | 160 | 116 | 51 | 13 | 0 |
| placebo reported | 206 | 122 | 111 | 81 | 37 | 8 | 0 |
| placebo recon. | 206 | 122 | 111 | 81 | 37 | 8 | 0 |

**Figure S10ai.** SORCE Leibovich high risk, 3 years.

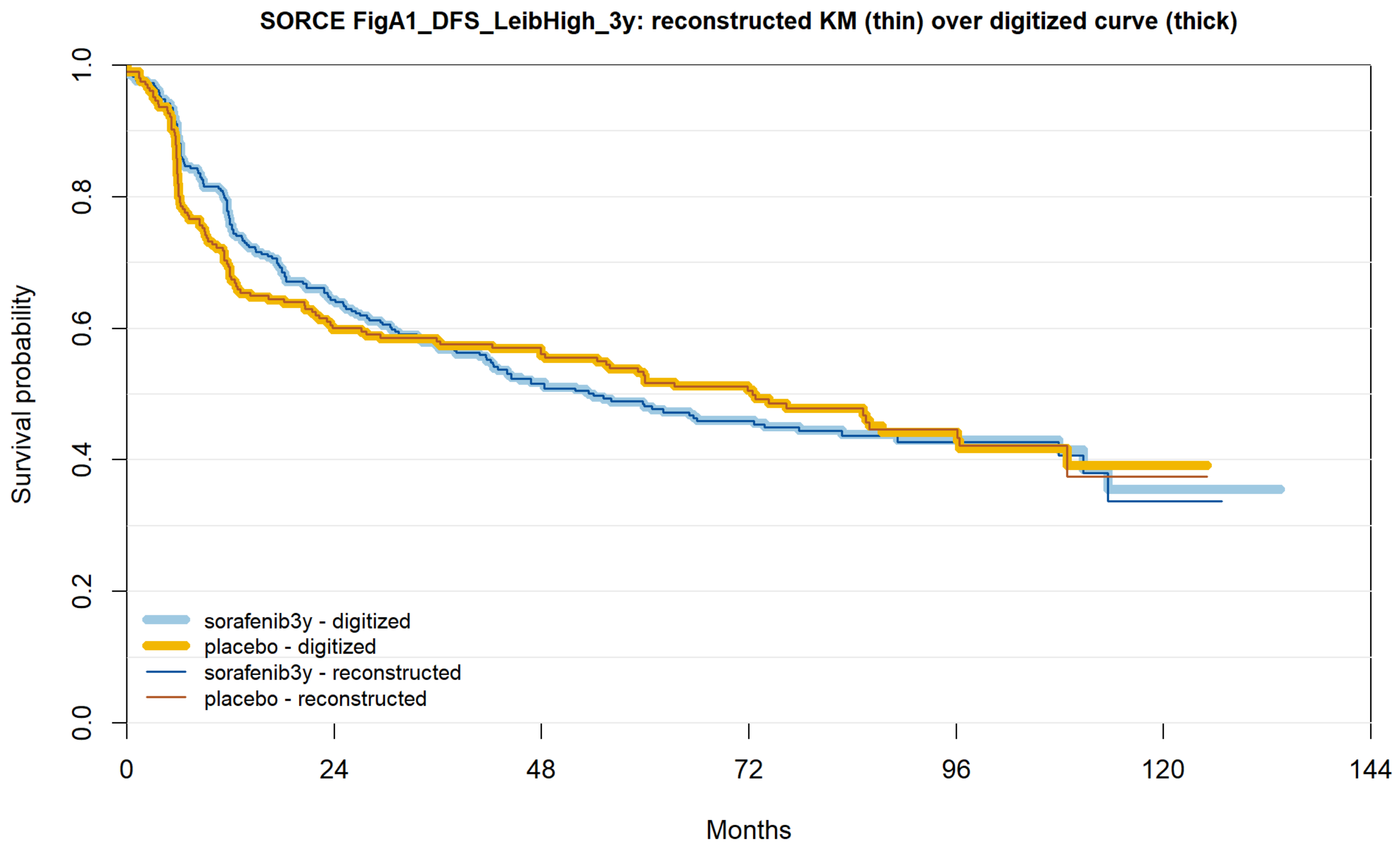


| | 0 | 24 | 48 | 72 | 96 | 120 | 144 |
|---|---|---|---|---|---|---|---|
| sorafenib3y reported | 295 | 185 | 143 | 100 | 43 | 8 | 0 |
| sorafenib3y recon. | 295 | 185 | 143 | 100 | 43 | 8 | 0 |
| placebo reported | 206 | 122 | 111 | 81 | 37 | 8 | 0 |
| placebo recon. | 206 | 122 | 111 | 81 | 37 | 8 | 0 |

**Figure S10aj.** SORCE clear cell, 1 year.

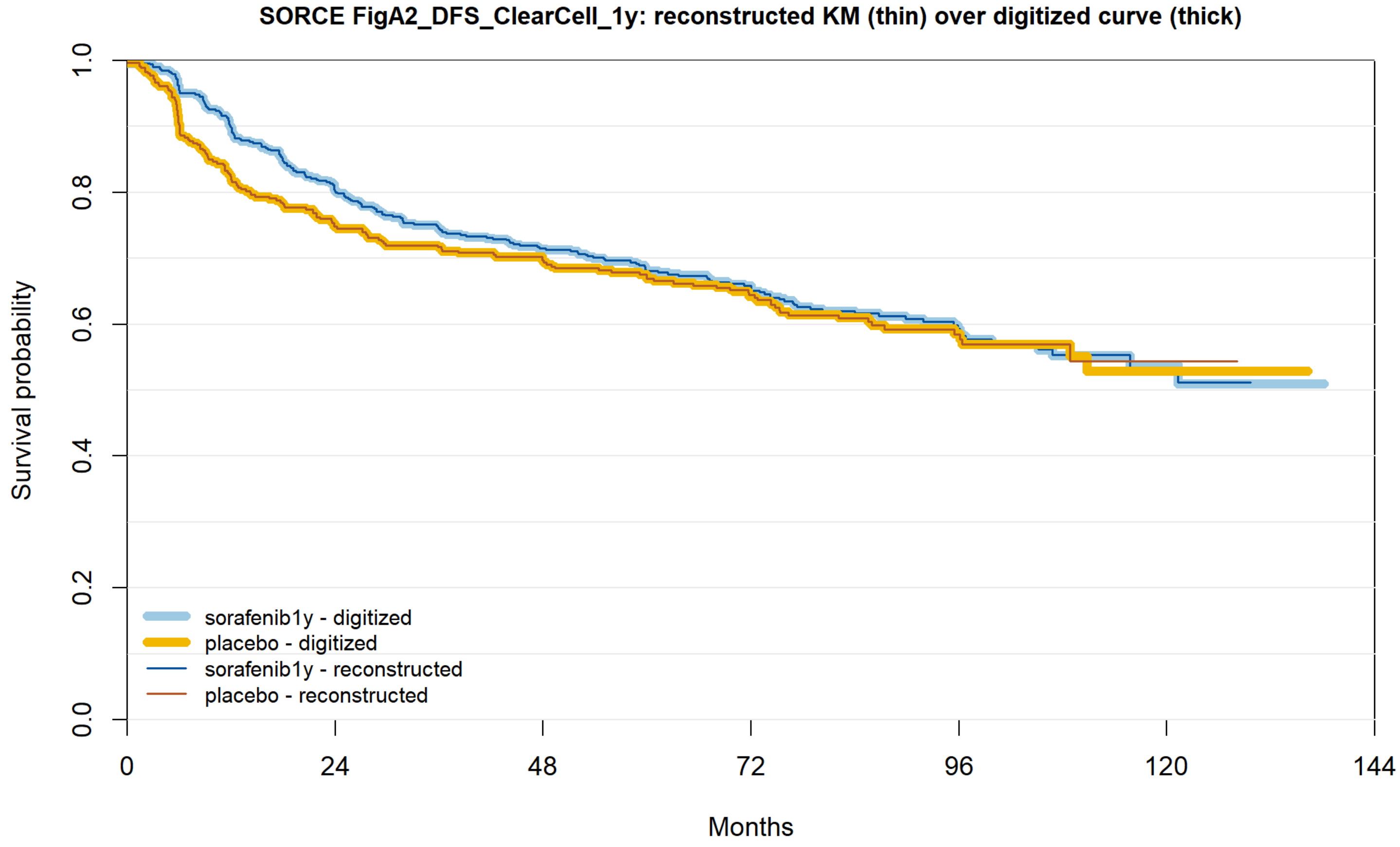


| | | | | | | | |
|---|---|---|---|---|---|---|---|
| sorafenib1y reported | 534 | 415 | 353 | 252 | 110 | 26 | 0 |
| sorafenib1y recon. | 534 | 415 | 353 | 252 | 110 | 26 | 0 |
| placebo reported | 361 | 264 | 236 | 176 | 75 | 15 | 0 |
| placebo recon. | 361 | 264 | 236 | 176 | 75 | 15 | 0 |

**Figure S10ak.** SORCE clear cell, 3 years.

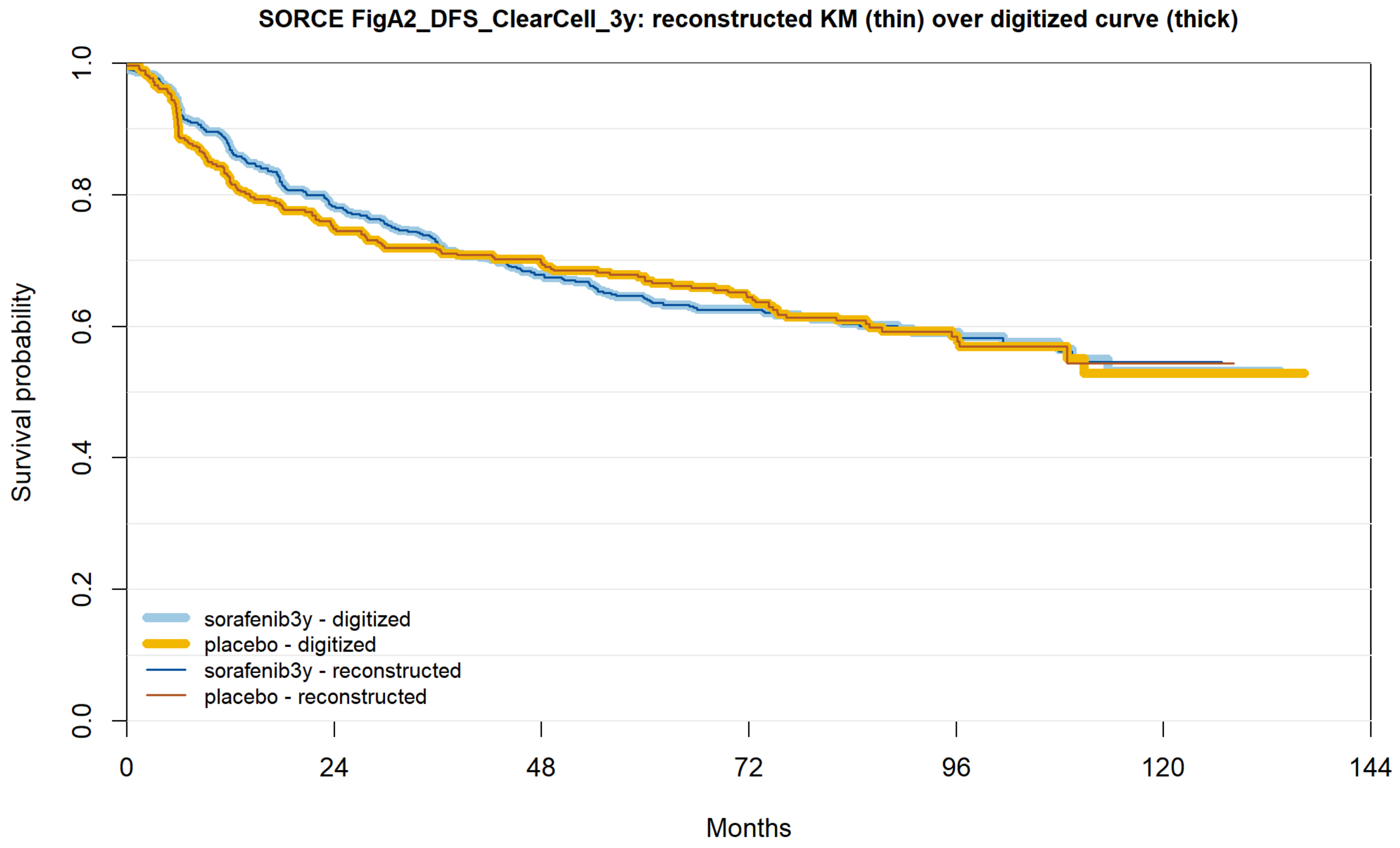


| | | | | | | | |
|---|---|---|---|---|---|---|---|
| sorafenib3y reported | 550 | 417 | 342 | 253 | 98 | 17 | 0 |
| sorafenib3y recon. | 550 | 417 | 342 | 253 | 98 | 17 | 0 |
| placebo reported | 361 | 264 | 236 | 176 | 75 | 15 | 0 |
| placebo recon. | 361 | 264 | 236 | 176 | 75 | 15 | 0 |

**Figure S10al.** IMmotion010 OS.

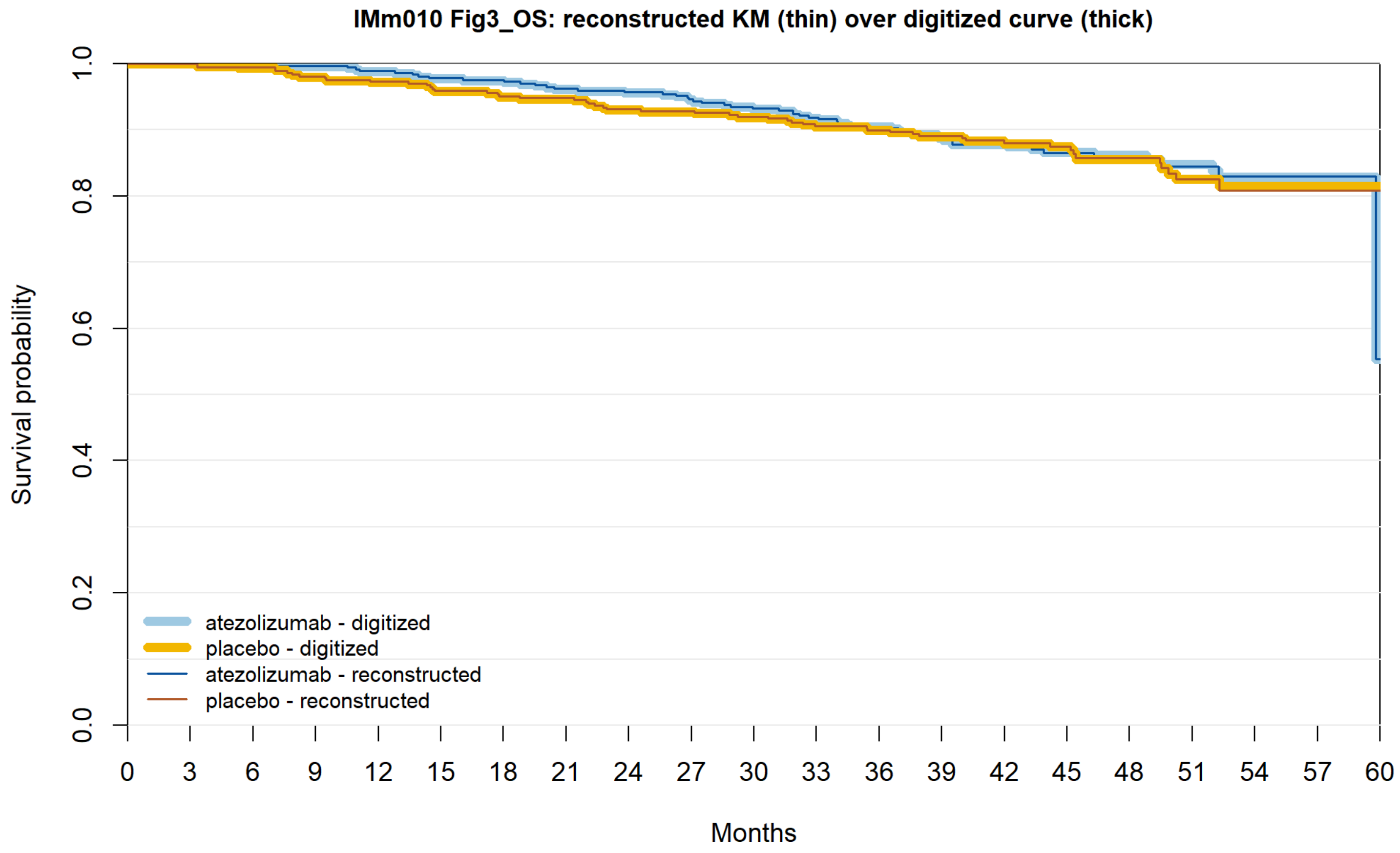


| | 0 | 3 | 6 | 9 | 12 | 15 | 18 | 21 | 24 | 27 | 30 | 33 | 36 | 39 | 42 | 45 | 48 | 51 | 54 | 57 | 60 |
|---|---|---|---|---|---|---|---|---|---|---|---|---|---|---|---|---|---|---|---|---|---|
| atezolizumab reported | 390 | 383 | 379 | 378 | 371 | 365 | 363 | 357 | 355 | 347 | 341 | 336 | 326 | 300 | 250 | 196 | 144 | 103 | 58 | 28 | 2 |
| atezolizumab recon. | 390 | 383 | 379 | 378 | 371 | 365 | 363 | 357 | 355 | 347 | 341 | 336 | 326 | 300 | 250 | 196 | 144 | 103 | 58 | 28 | 2 |
| placebo reported | 388 | 379 | 372 | 363 | 355 | 349 | 343 | 340 | 331 | 330 | 325 | 314 | 304 | 287 | 226 | 181 | 138 | 92 | 49 | 20 | 6 |
| placebo recon. | 388 | 379 | 372 | 363 | 355 | 349 | 343 | 340 | 331 | 330 | 325 | 314 | 304 | 287 | 226 | 181 | 138 | 92 | 49 | 20 | 6 |

**Figure S10am.** S-TRAC OS.

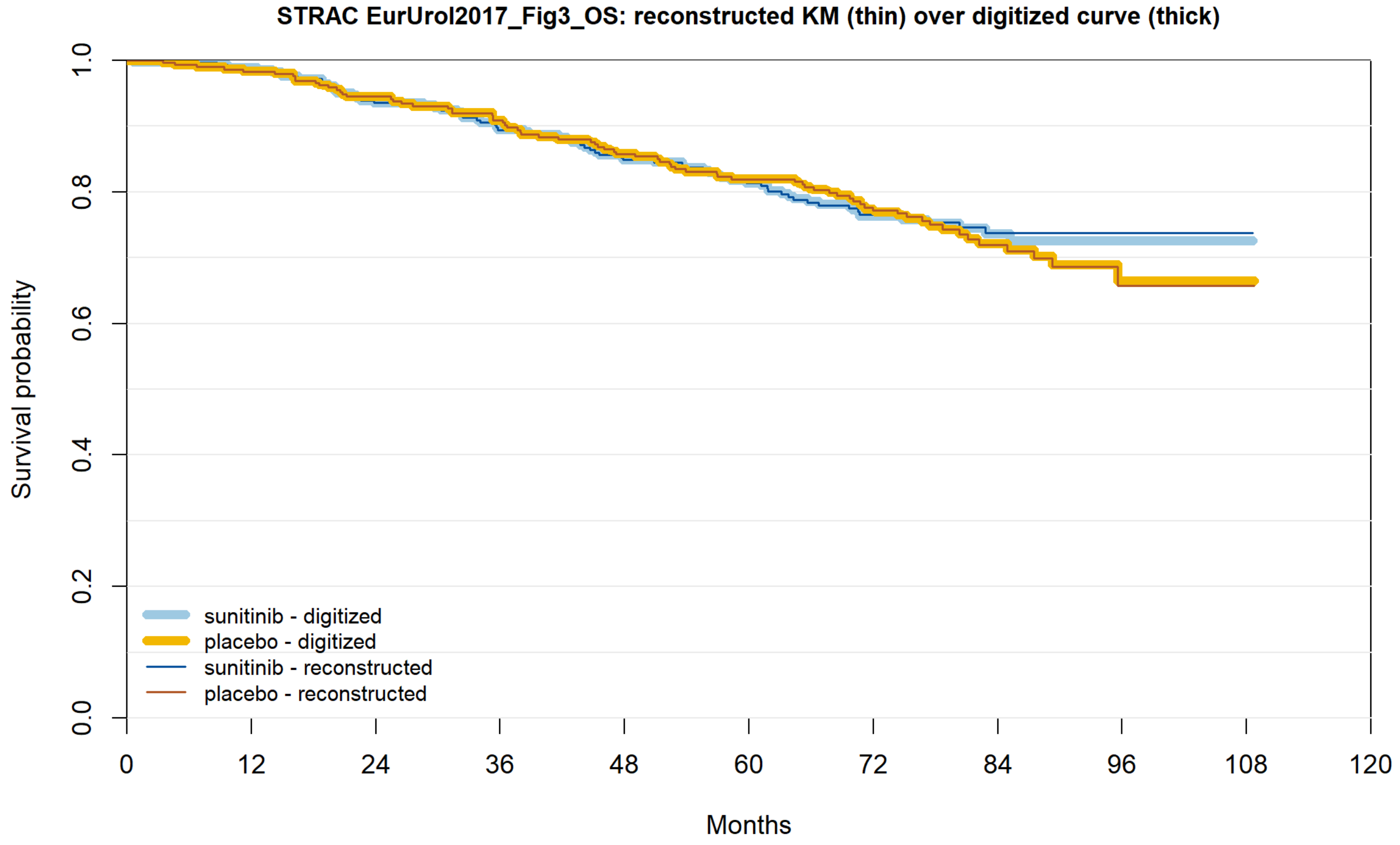


| | 0 | 12 | 24 | 36 | 48 | 60 | 72 | 84 | 96 | 108 | 120 |
|---|---|---|---|---|---|---|---|---|---|---|---|
| sunitinib reported | 309 | 278 | 258 | 236 | 222 | 205 | 160 | 82 | 16 | 1 | 0 |
| sunitinib recon. | 309 | 278 | 258 | 236 | 222 | 205 | 160 | 82 | 16 | 1 | 0 |
| placebo reported | 306 | 289 | 269 | 250 | 231 | 210 | 172 | 82 | 23 | 1 | 0 |
| placebo recon. | 306 | 289 | 269 | 250 | 231 | 210 | 172 | 82 | 23 | 1 | 0 |

**Figure S10an.** PROTECT OS (JCO 2017).

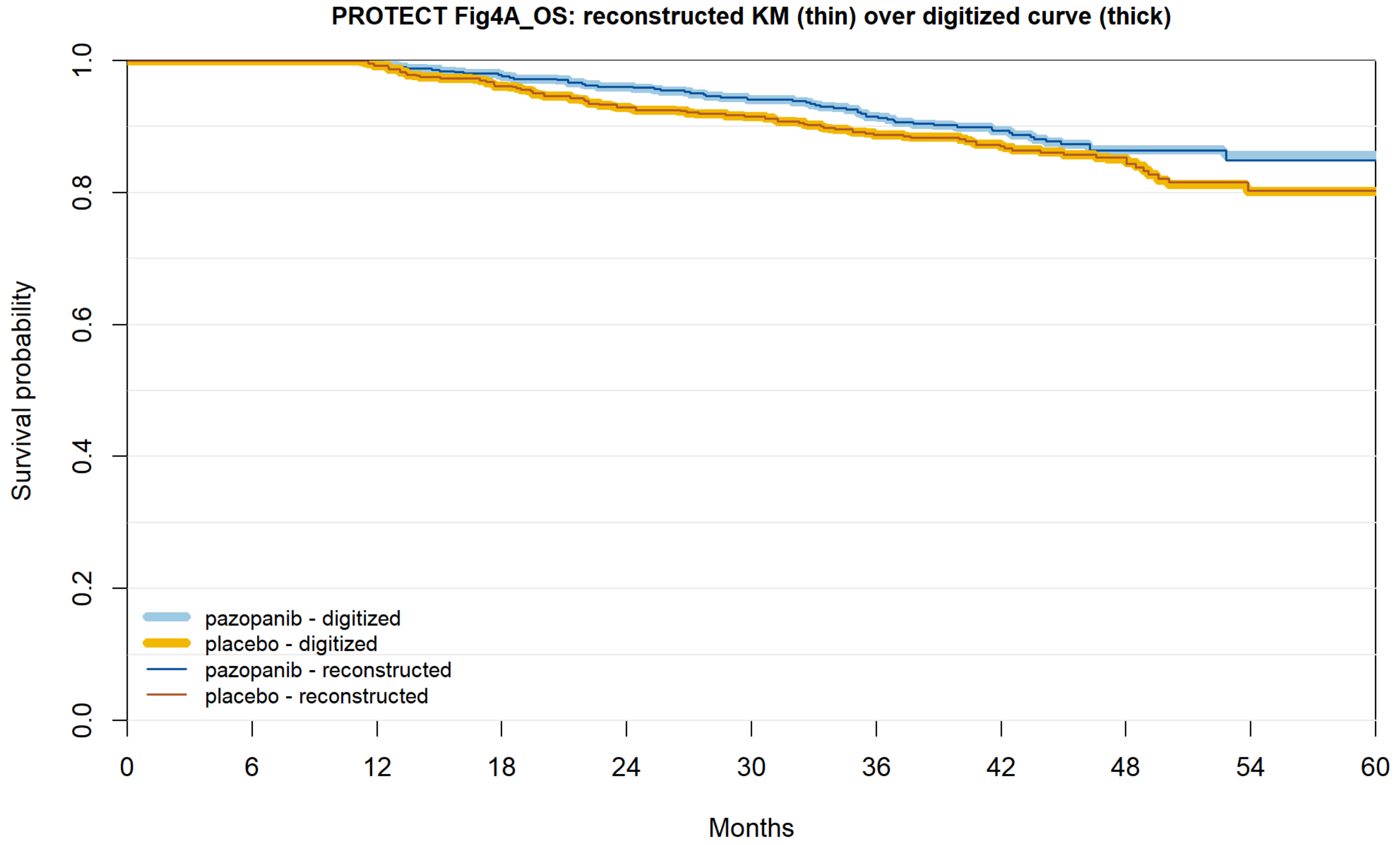


| | | | | | | | | | | | |
|---|---|---|---|---|---|---|---|---|---|---|---|
| pazopanib reported | 571 | 529 | 517 | 498 | 484 | 468 | 428 | 315 | 178 | 58 | 0 |
| pazopanib recon. | 571 | 529 | 517 | 498 | 484 | 468 | 428 | 315 | 178 | 58 | 0 |
| placebo reported | 564 | 539 | 525 | 504 | 484 | 476 | 427 | 310 | 176 | 63 | 0 |
| placebo recon. | 564 | 539 | 525 | 504 | 484 | 476 | 427 | 310 | 176 | 63 | 0 |

**Figure S10ao.** PROTECT final OS (Eur Urol 2021; did not pass the authors’ visual check; shown for reference only, not used in any pooled analysis).

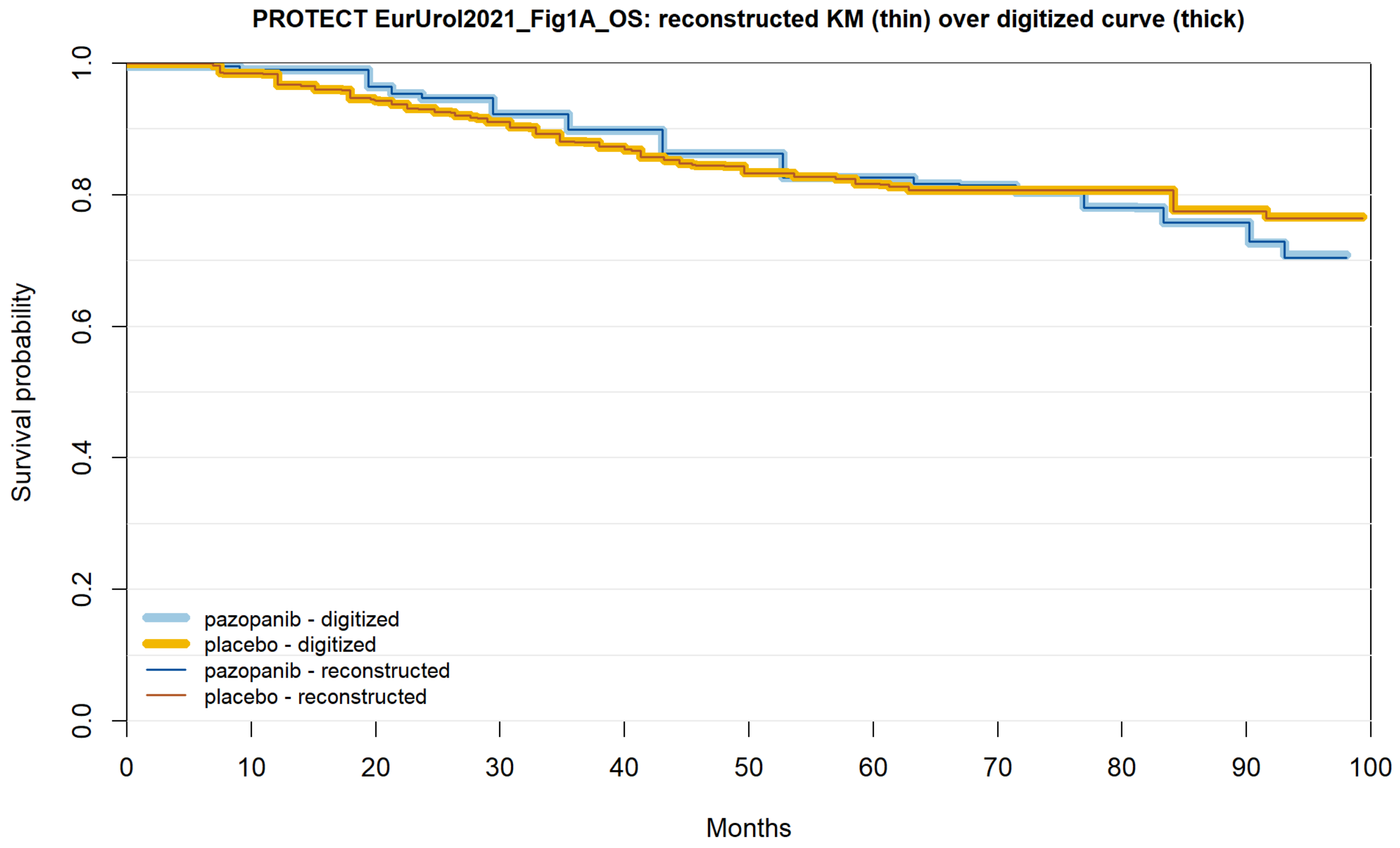


| | | | | | | | | | | | |
|---|---|---|---|---|---|---|---|---|---|---|---|
| pazopanib reported | 769 | 703 | 669 | 638 | 605 | 565 | 528 | 424 | 228 | 81 | 0 |
| pazopanib recon. | 769 | 703 | 669 | 638 | 605 | 565 | 528 | 424 | 228 | 81 | 0 |
| placebo reported | 769 | 731 | 684 | 655 | 613 | 581 | 555 | 458 | 242 | 80 | 0 |
| placebo recon. | 769 | 731 | 684 | 655 | 613 | 581 | 555 | 458 | 242 | 80 | 0 |

**Figure S10ap.** SORCE OS, 3 years.

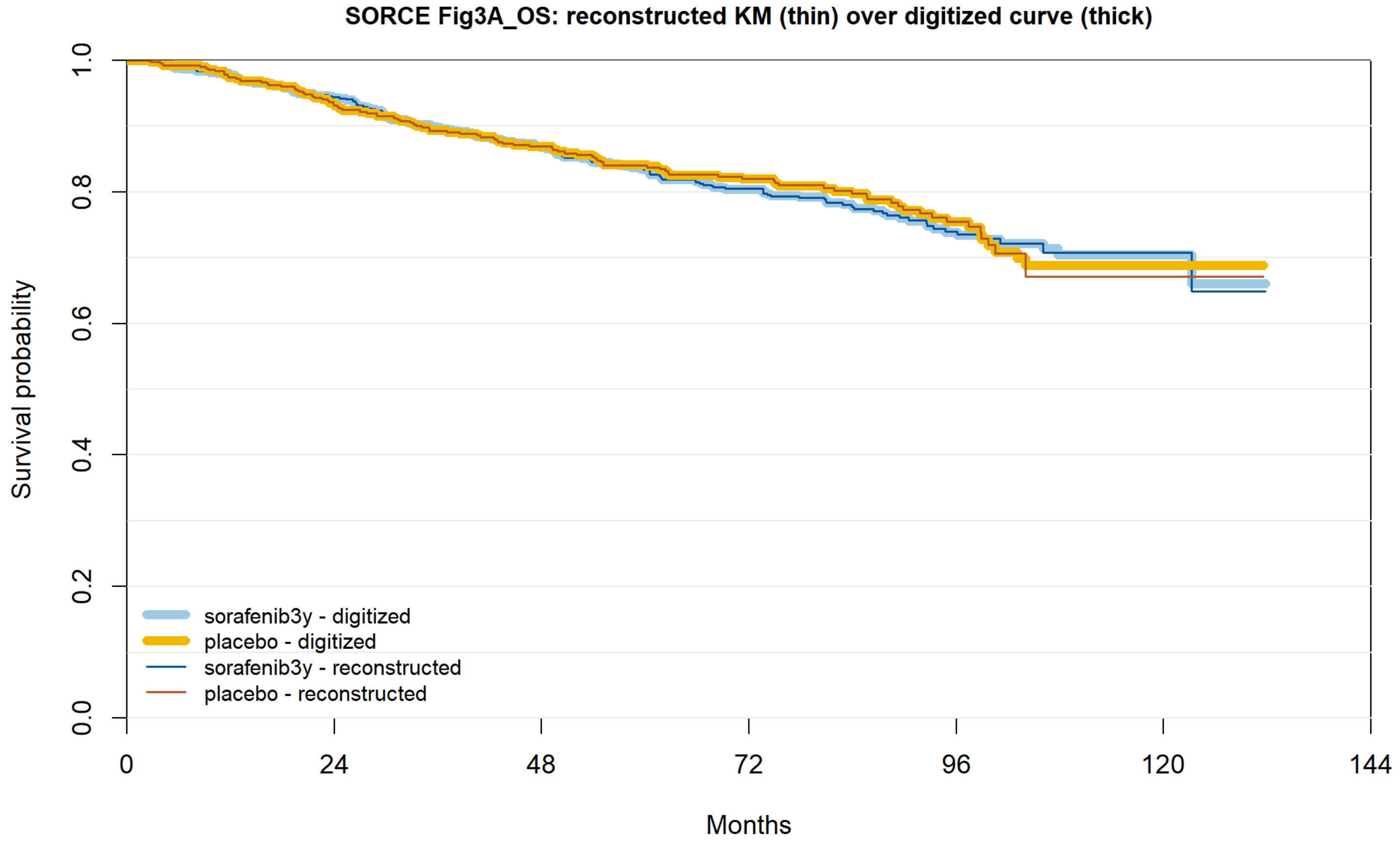


| | | | | | | | |
|---|---|---|---|---|---|---|---|
| sorafenib3y reported | 638 | 582 | 514 | 382 | 158 | 32 | 0 |
| sorafenib3y recon. | 638 | 582 | 514 | 382 | 158 | 32 | 0 |
| placebo reported | 430 | 394 | 351 | 261 | 115 | 19 | 0 |
| placebo recon. | 430 | 394 | 351 | 261 | 115 | 19 | 0 |

**Figure S10aq.** SORCE OS, 1 year.

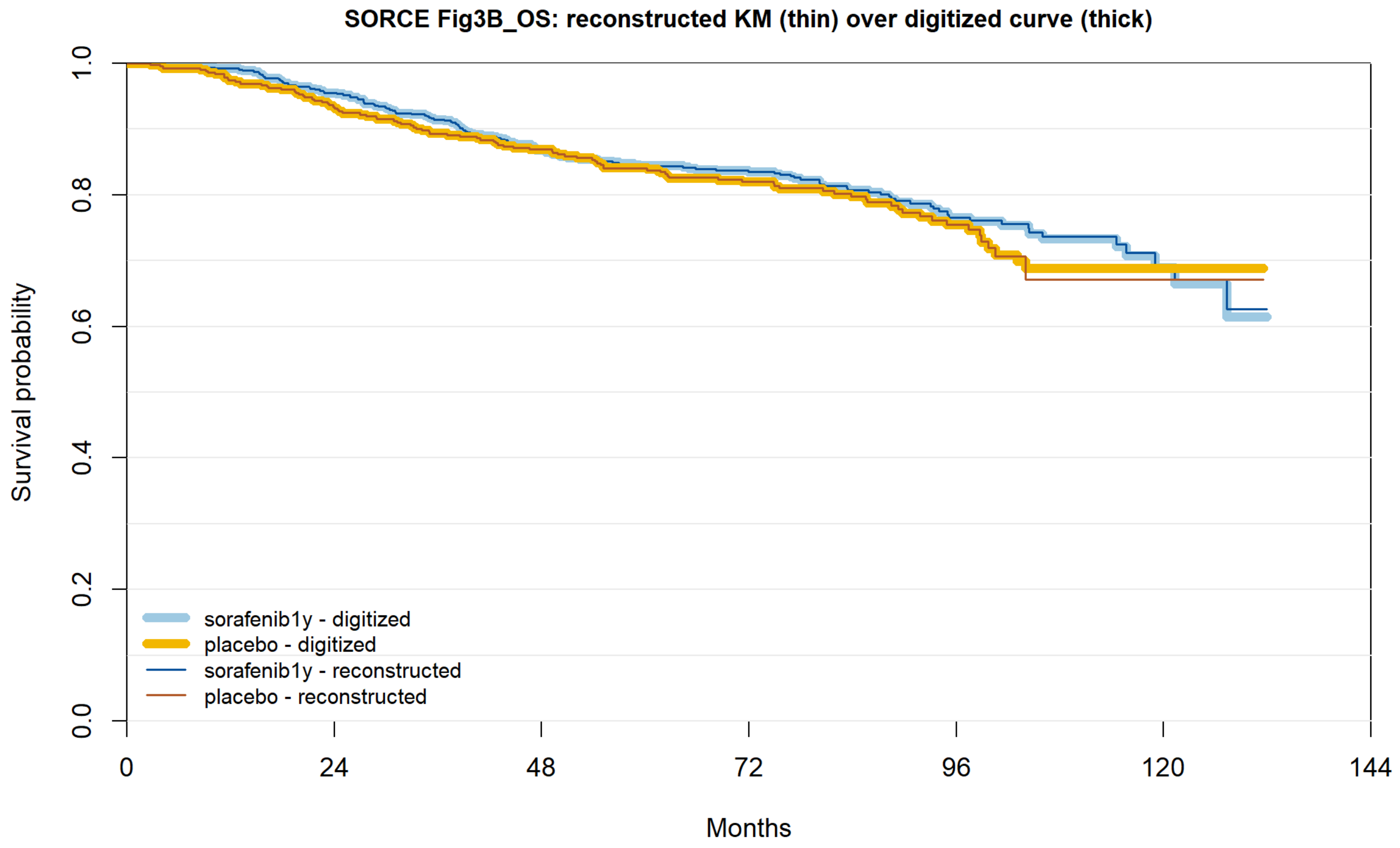


| | 0 | 24 | 48 | 72 | 96 | 120 | 144 |
|---|---|---|---|---|---|---|---|
| sorafenib1y reported | 641 | 594 | 517 | 393 | 169 | 36 | 0 |
| sorafenib1y recon. | 641 | 594 | 517 | 393 | 169 | 36 | 0 |
| placebo reported | 430 | 394 | 351 | 261 | 115 | 19 | 0 |
| placebo recon. | 430 | 394 | 351 | 261 | 115 | 19 | 0 |

**Figure S10ar.** CheckMate 914 part A OS.

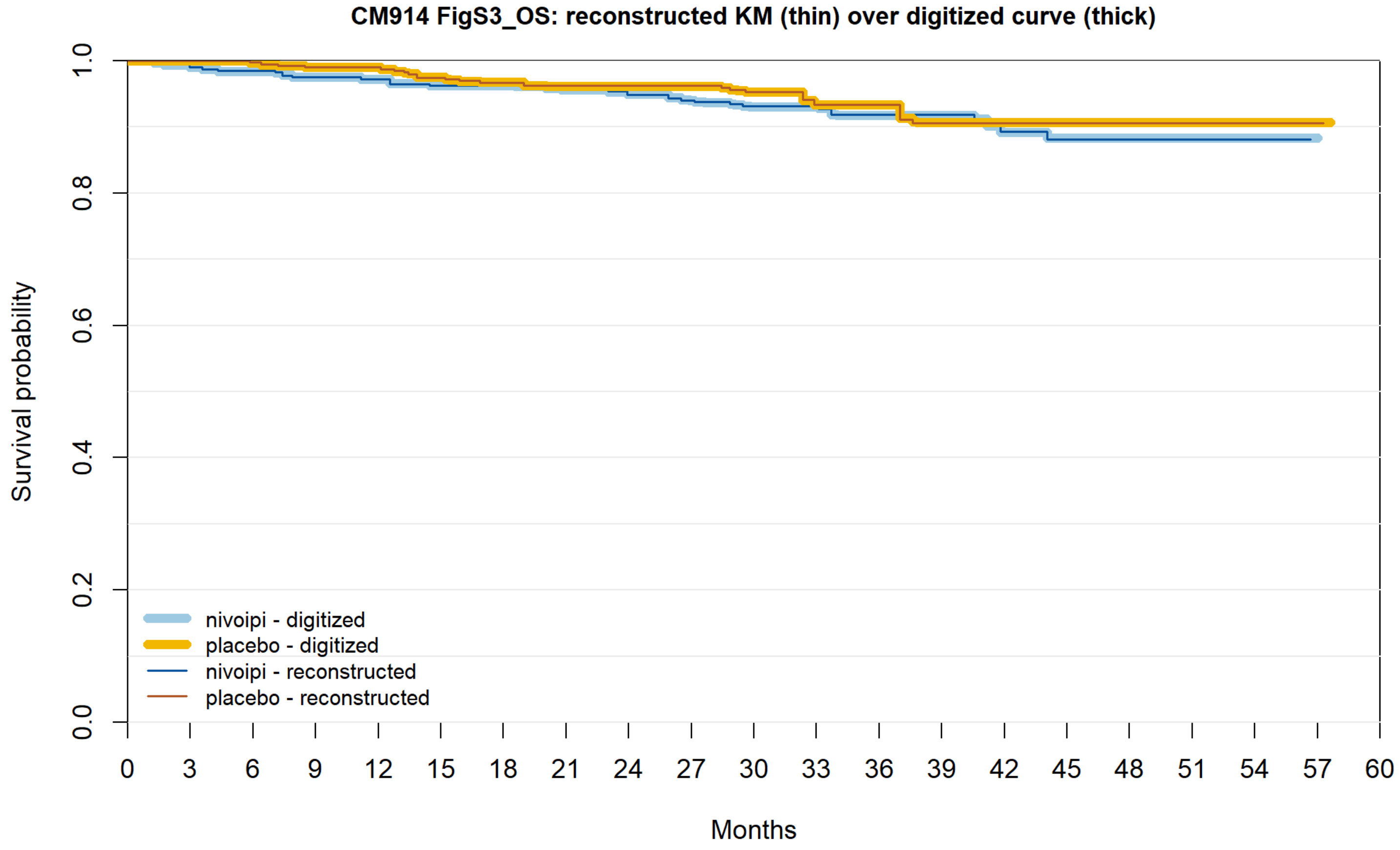


| | | | | | | | | | | | | | | | | | | | | | |
|---|---|---|---|---|---|---|---|---|---|---|---|---|---|---|---|---|---|---|---|---|---|
| nivoipi reported | 405 | 399 | 391 | 386 | 383 | 378 | 374 | 363 | 348 | 330 | 278 | 230 | 184 | 137 | 96 | 68 | 40 | 23 | 8 | 1 | 0 |
| nivoipi recon. | 405 | 399 | 391 | 386 | 383 | 378 | 374 | 363 | 348 | 330 | 278 | 230 | 184 | 137 | 96 | 68 | 40 | 23 | 8 | 0 | 0 |
| placebo reported | 411 | 403 | 400 | 396 | 395 | 386 | 381 | 372 | 361 | 343 | 292 | 237 | 184 | 138 | 98 | 66 | 36 | 22 | 10 | 1 | 0 |
| placebo recon. | 411 | 403 | 400 | 396 | 395 | 386 | 381 | 372 | 361 | 343 | 292 | 237 | 184 | 138 | 98 | 66 | 36 | 22 | 10 | 1 | 0 |

**Figure S11.** ΔRMST(τ) curves for the curves added under deviation D8 (SAP section 9, items 2–7), as in Figure S1.

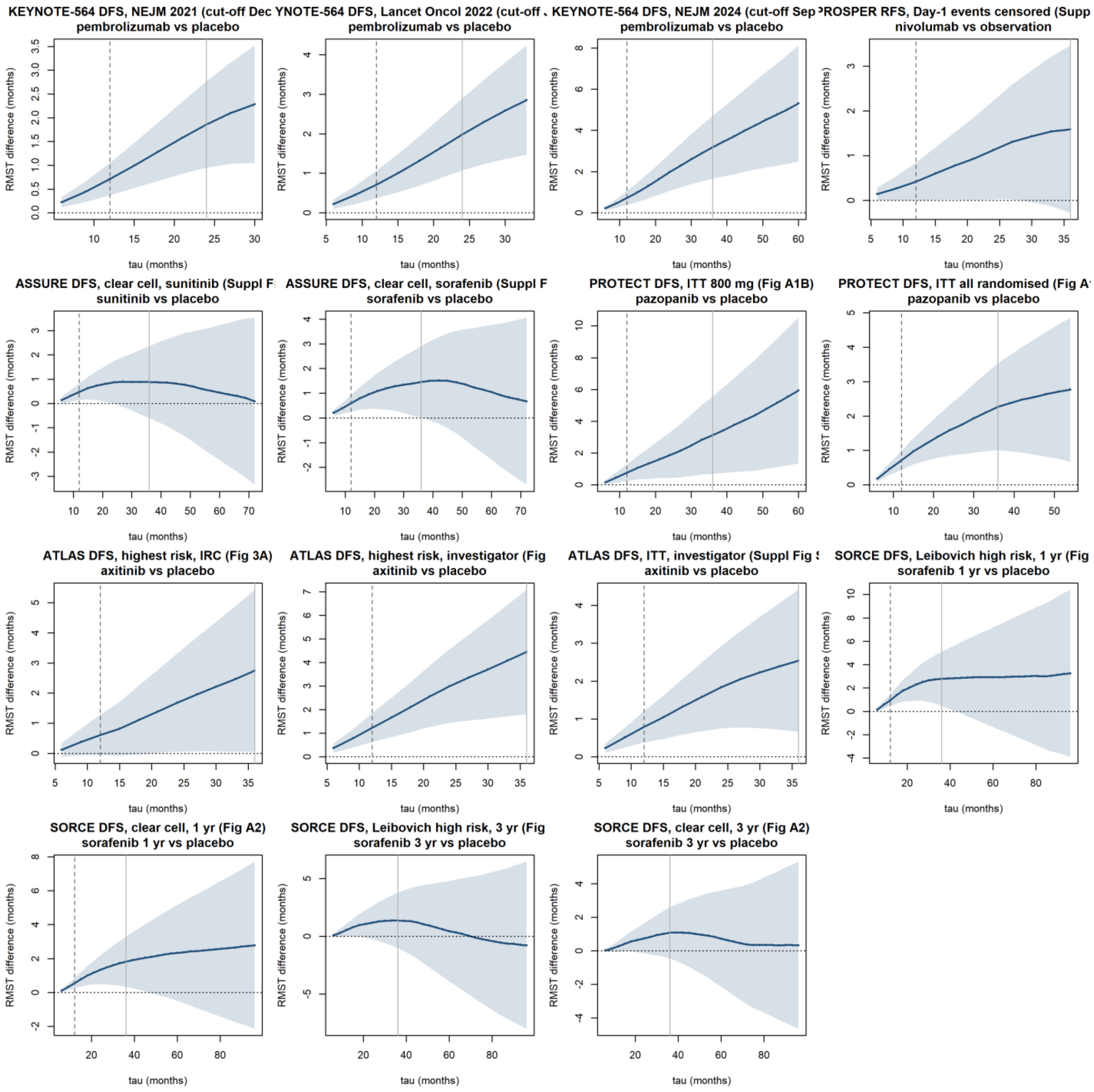

**Figure S12.** Time-varying HR(t), curves added under deviation D8 (as in Figure S2).

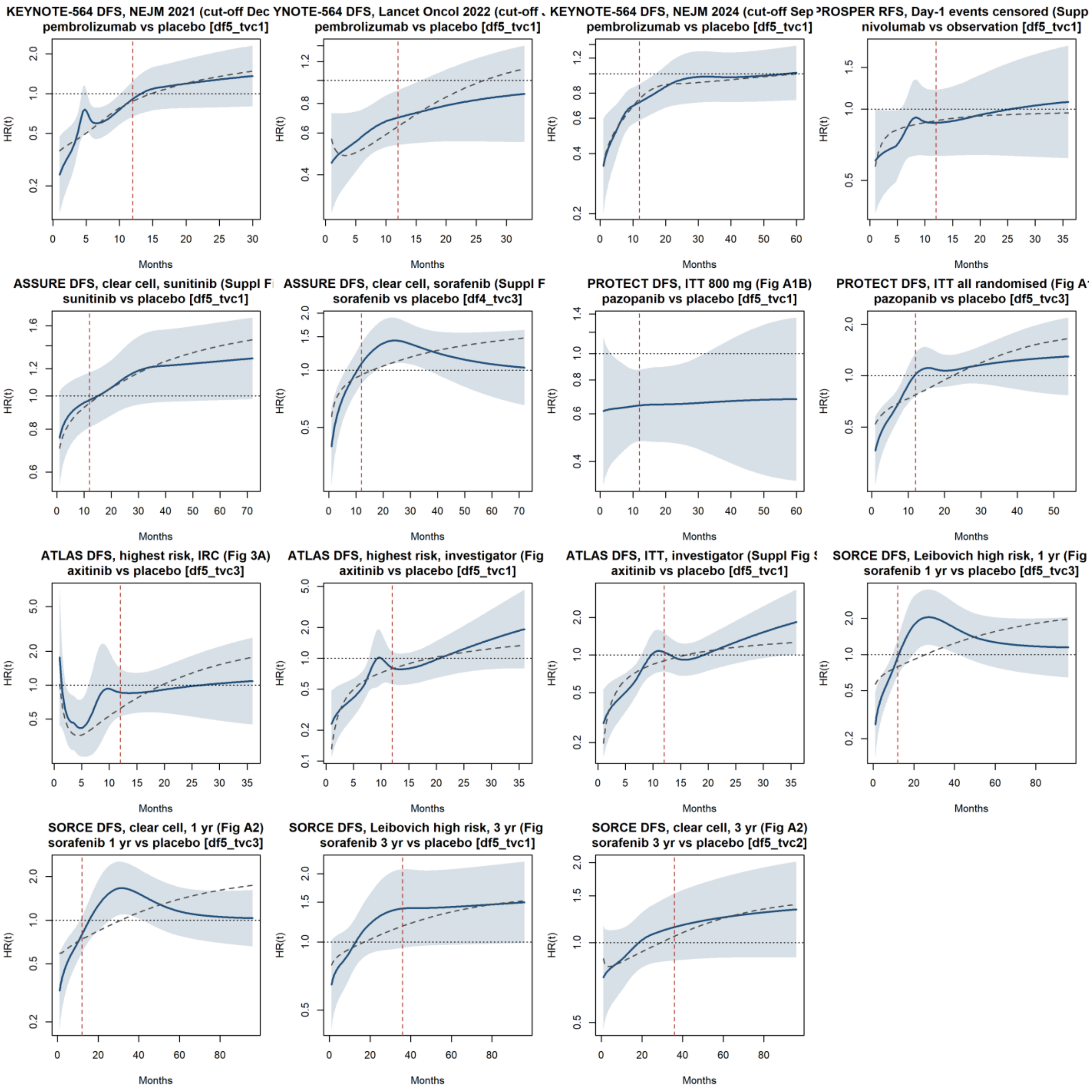

**Figure S13.** Time-varying HR(t) for overall survival, all OS curves (version 1 curves and those added under D8; the final PROTECT OS curve did not pass the authors' visual check; shown for reference only, not used in any pooled analysis).

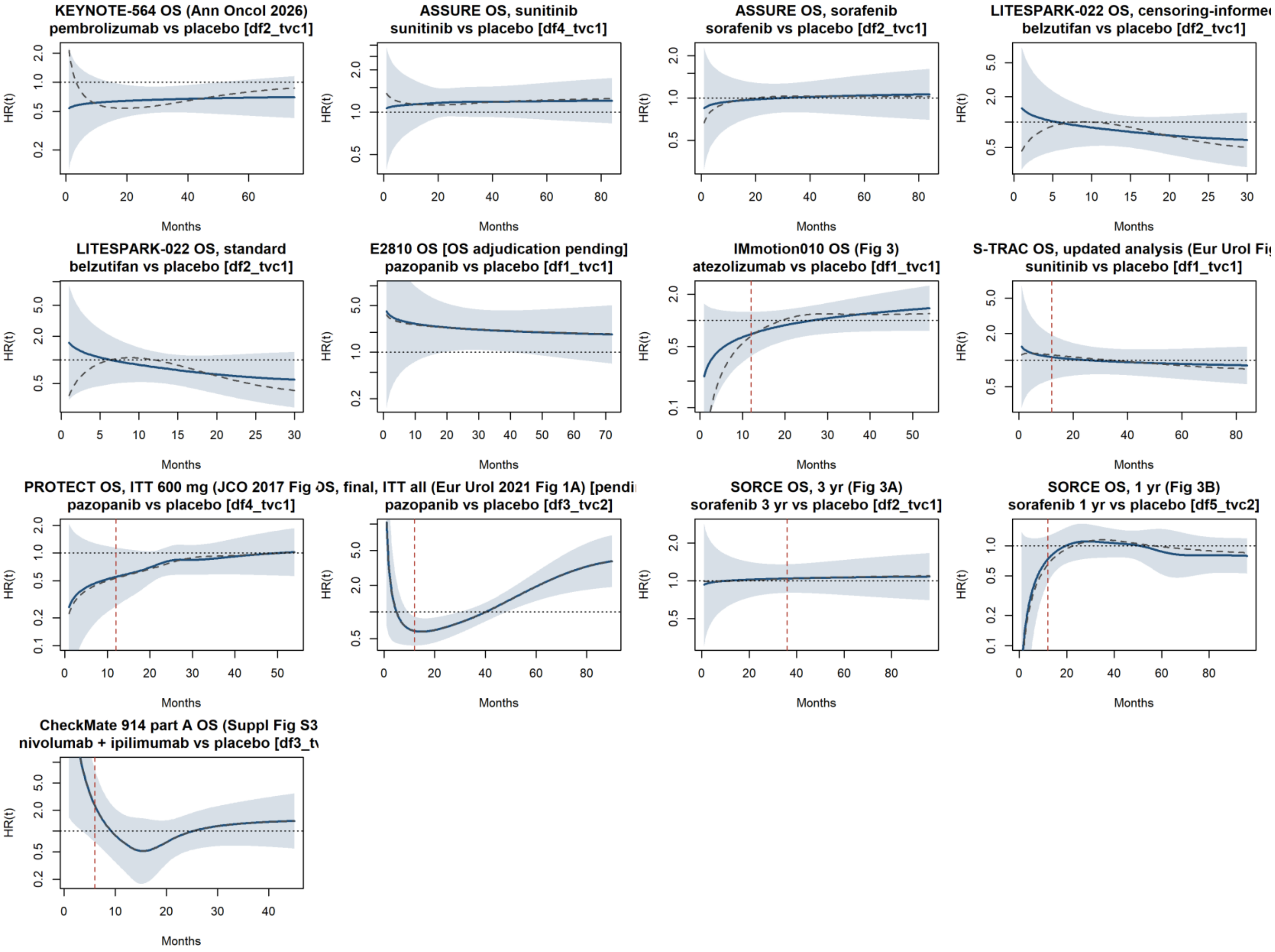

**Figure S14.** Log(−log) survival plots, curves added under deviation D8.

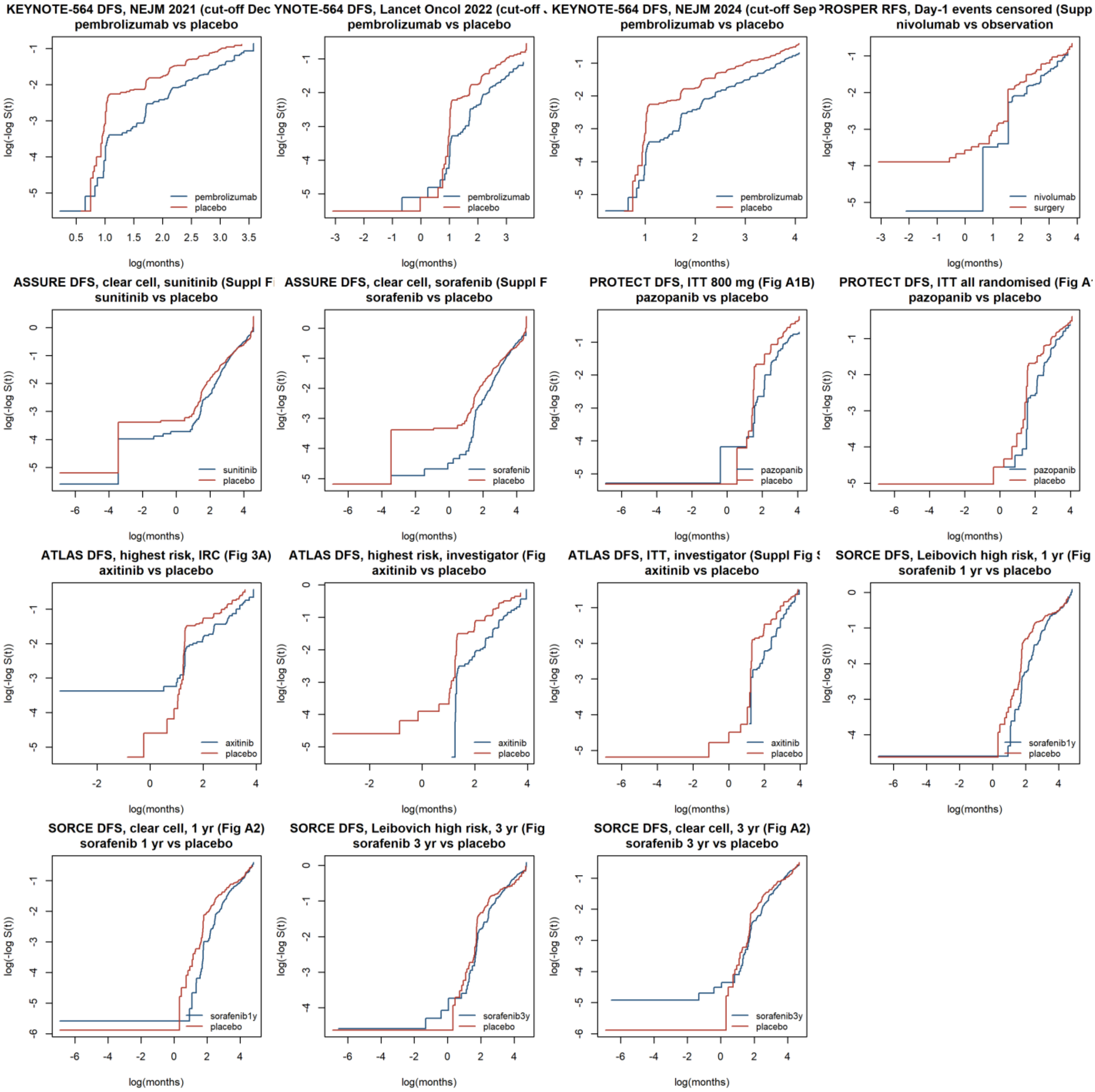

**Figure S15.** Smoothed hazard functions by arm, curves added under deviation D8.

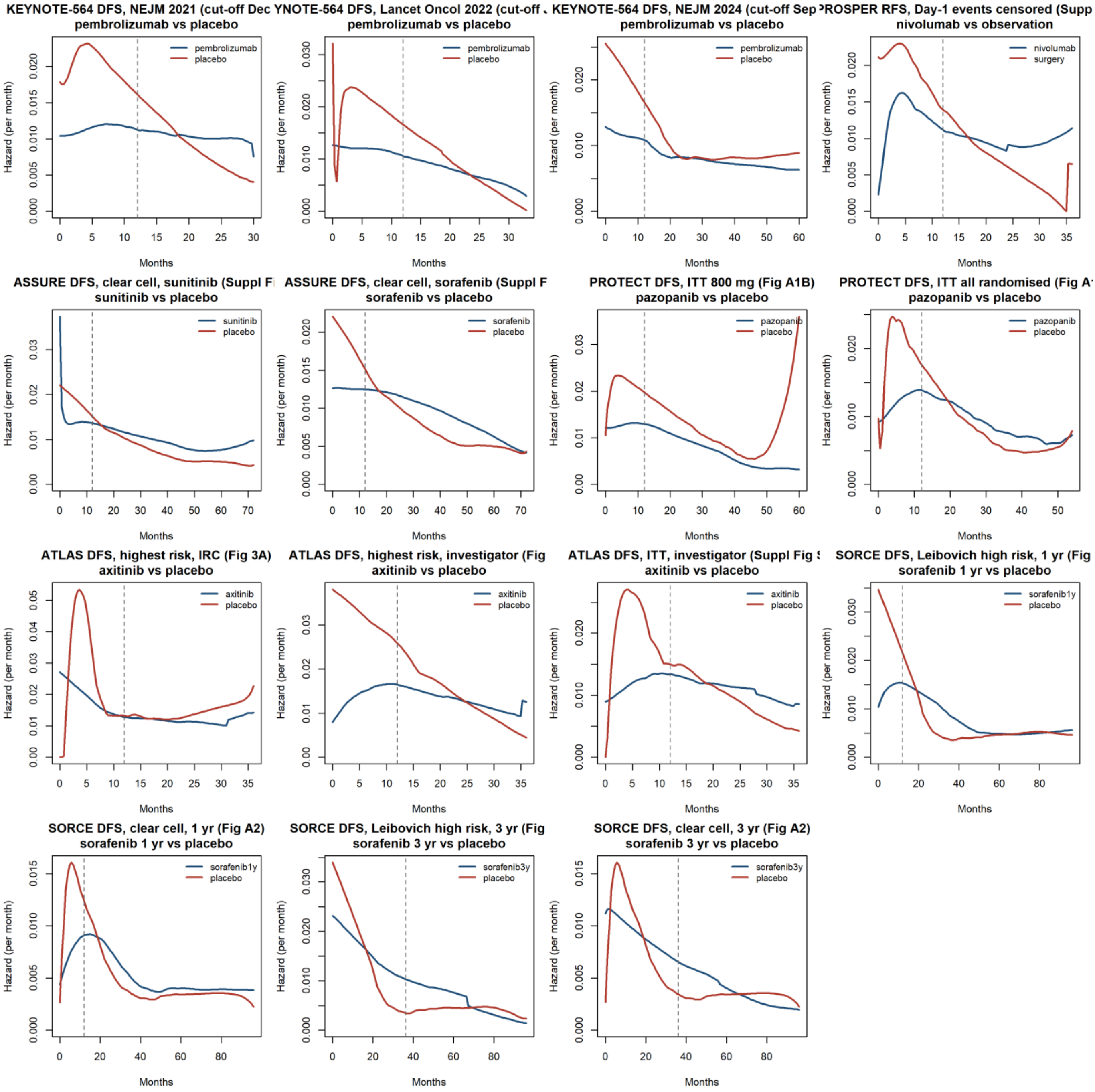

**Figure S16.** Class-level pooling: main analyses and sensitivity analyses at the main landmarks, with ordinary Hartung-Knapp (black) and modified Hartung-Knapp (grey, q* = max(1, q); post hoc) 95% CIs.

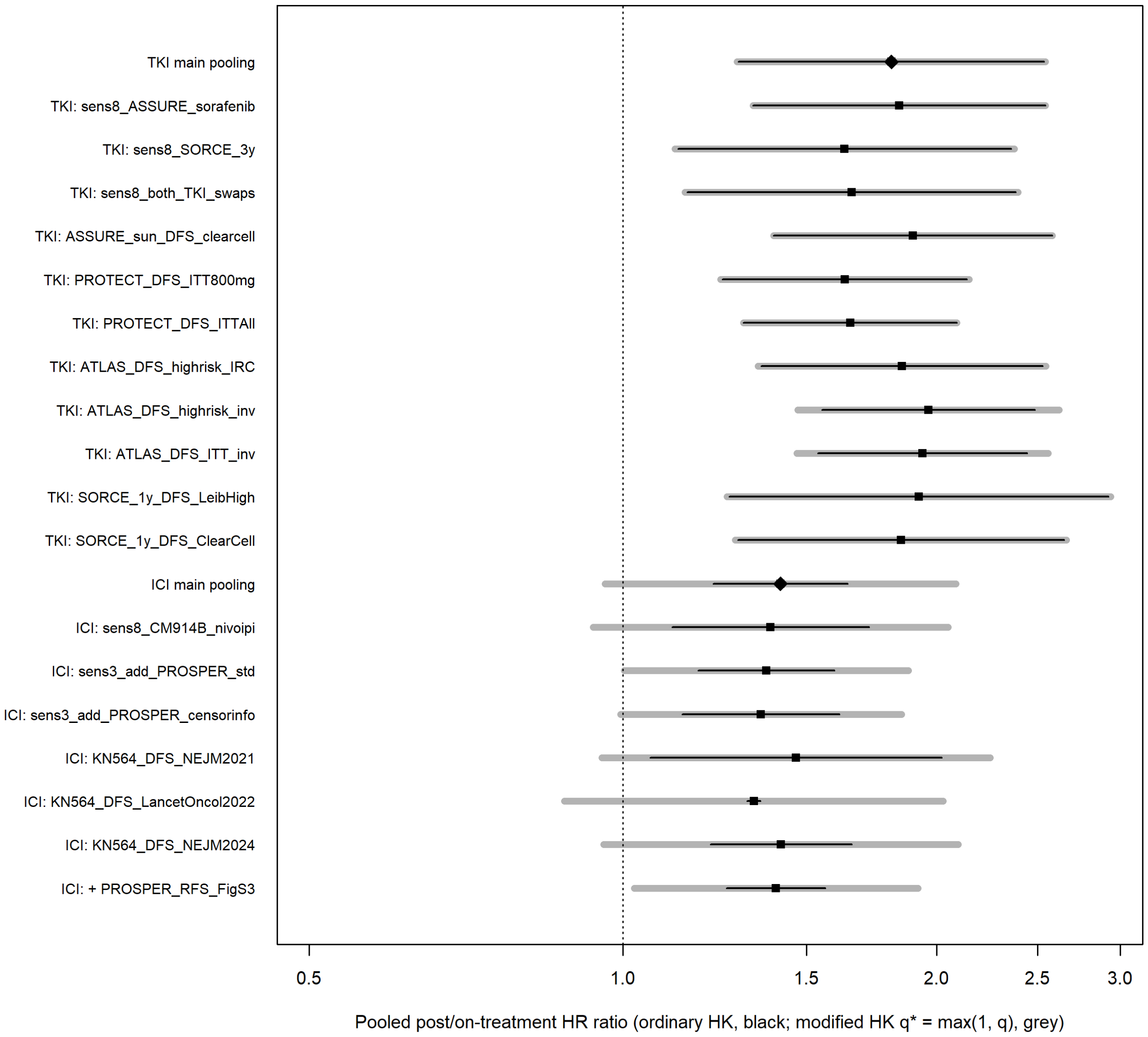

The main manuscript already shows the ΔRMST(τ) curves of the main comparisons (Fig 1), the ΔRMST forest plot (Fig 2), the piecewise HRs and the forest plot of post-treatment to on-treatment HR ratios with the class pooling (Fig 3) and the window-matched analysis of the main comparisons (Fig 4); the corresponding figures are therefore not repeated here. The digitisation overlays that show each extracted curve on the published figure reproduce copyrighted publisher figures and are not included; they are available to reviewers on request.
