## Supplementary material for "Does the Disease-free Survival Benefit of Adding Belzutifan to Adjuvant Pembrolizumab Persist Beyond the Planned Treatment Period? A Reconstructed Individual Patient Data Analysis of Phase 3 Renal Cell Carcinoma Trials": Registered Statistical Analysis Plan (OSF, 12 October 2026)

### Statistical Analysis Plan — Durability of disease-free survival benefit after adjuvant therapy for renal cell carcinoma: a reconstructed individual patient data analysis of phase III trials

**Version:** 4.0
**Date:** 12 October 2026
**Investigators:** Hao-Han Chang (Graduate Institute of Clinical Medicine, Kaohsiung Medical University; Department of Urology, Kaohsiung Medical University Hospital) and colleagues
**Status:** Final version for prospective registration on OSF, prepared before digitization of the LITESPARK-022 and KEYNOTE-564 curves.

> **Purpose.** This plan prespecifies all analytic decisions before any reconstructed data from the index trials (LITESPARK-022 and KEYNOTE-564) are generated; items that depend on documents not yet obtained will be completed only according to the rules stated here and will not be adjusted after results are seen.

---

#### 1. Background and objectives

##### 1.1 Background

In LITESPARK-022, pembrolizumab plus belzutifan improved DFS compared with pembrolizumab alone (HR 0.72, 95% CI 0.59–0.87; NEJM 2026, PMID 42384869). Belzutifan inhibits HIF-2α and, like VEGFR-TKIs, acts on the VHL–HIF–VEGF axis. In adjuvant VEGFR-TKI trials, DFS and OS were dissociated, and there are signals that the DFS benefit converged after treatment discontinuation (S-TRAC conditional DFS, PMID 31825895; curve shapes in PROTECT, SORCE and ASSURE). Published analyses of belzutifan (Cigliola 2026; Das 2026) used only the overall HR and did not address whether the benefit persists after treatment ends.

##### 1.2 Primary objective

To quantify the DFS benefit within each phase III adjuvant RCC trial using the difference in restricted mean survival time (RMST), and to describe whether the benefit continues to widen, is maintained, or converges after the end of planned treatment.

##### 1.3 Secondary objectives

1. To describe the change in benefit over time using piecewise HRs, the trajectory of the DFS difference, and flexible parametric HR(t).
2. To compare the post-treatment pattern of benefit between VEGFR-TKI and ICI trials, and to describe which class LITESPARK-022 more closely resembles.
3. To describe OS using the same methods.

---

#### 2. Study design

- Secondary analysis of published phase III RCTs, using individual patient data reconstructed from published KM curves (reconstructed IPD).
- No actual patient-level data are used, and no data are requested from trial sponsors.
- All treatment effects are estimated within trials, preserving randomization.
- Across trials, only the temporal pattern of effects is compared; **arms from different trials are not pooled**, and no Bucher or other indirect comparison is performed.
- Ethics review: not expected to be required, as only published aggregate data are used; a formal

exemption will be obtained from the institutional review board if required.

#### 3. Trial selection

##### 3.1 Inclusion criteria

1. Phase III randomized controlled trial.
2. Population with RCC after radical or partial nephrectomy (including M1 NED after resection).
3. Experimental arm receiving adjuvant or perioperative systemic therapy; control arm receiving placebo, observation, or placebo added to a common background therapy.
4. Published in full in a peer-reviewed journal (including appendices).
5. At least one KM curve of the primary efficacy endpoint with a number-at-risk table.

##### 3.2 Exclusion criteria

1. Available only as a conference abstract or poster.
2. KM curve without a number-at-risk table, or with insufficient resolution to pass the quality checks in Section 5.3.
3. Trials of treatment eras no longer in use, such as cytokines, vaccines, or monoclonal antibodies (e.g., girentuximab).

Trials not meeting the criteria are excluded directly, without requesting data from authors or sponsors, and the exclusion is reported in the manuscript.

##### 3.3 Literature search

- A PubMed search was conducted on 9–10 October 2026 (search strings will be documented in Appendix A).
- Supplementary searches will be performed in PROSPERO, medRxiv, and 2026 ASCO GU, ASCO and ESMO abstracts.
- The search process will be reported in the manuscript appendix; this study is not positioned as a systematic review.

##### 3.4 Prespecified trials and roles

| Trial | Comparison | Class | Role |
|---|---|---|---|
| LITESPARK-022 | pembro + belzutifan vs pembro + placebo | HIF-2α (added to ICI) | Index trial |
| KEYNOTE-564 | pembro vs placebo | ICI | ICI positive reference |
| IMmotion010 | atezolizumab vs placebo | ICI | ICI negative reference |
| CheckMate 914 Part A | nivo + ipi vs placebo | ICI | ICI negative reference |
| CheckMate 914 Part B | nivo vs placebo; nivo + ipi vs placebo | ICI | ICI negative reference (shared placebo) |
| S-TRAC | sunitinib vs placebo | VEGFR-TKI | TKI reference |
| ASSURE | sunitinib vs placebo; sorafenib vs placebo | VEGFR-TKI | TKI reference (shared placebo) |
| PROTECT | pazopanib 600 mg vs placebo | VEGFR-TKI | TKI reference |
| ATLAS | axitinib vs placebo | VEGFR-TKI | TKI reference |
| SORCE | sorafenib 1 year vs placebo; 3 years vs placebo | VEGFR-TKI | TKI reference (shared placebo) |
| PROSPER | perioperative nivo vs observation | ICI | Sensitivity analysis only |

EVEREST (everolimus, an mTOR inhibitor) does not belong to either class and is not included.

#### 4. Endpoints

- **Primary endpoint:** DFS as defined in each trial's primary analysis (RFS for PROSPER).
- **Secondary endpoint:** OS.
- DFS definitions differ across trials (e.g., SORCE counts only RCC deaths, ATLAS includes second primary cancers, and CheckMate 914 censors before subsequent therapy); these differences are listed in Table 1. Because all effects are estimated within trials, definitional differences do not affect within-trial comparisons, but they are listed as a limitation when cross-trial patterns are discussed.
- When the wording of the DFS definition differs between publications of the same trial, the wording in the publication containing the primary-analysis curve is used (e.g., NEJM 2024 for KEYNOTE-564).

---

#### 5. Data reconstruction

##### 5.1 Curve selection rules (prespecified)

A single primary-analysis curve is designated for each comparison, according to the following rules applied in order:

1. Use the ITT population (for PROTECT, ITT600mg, the primary analysis population of the original trial).
2. Use the assessment method of the trial's primary analysis (BICR, IRC or investigator).
3. Use the latest DFS analysis that has been published **and for which the full text is available**. If a later analysis is obtained **before** digitization of that trial begins, it replaces the earlier one; an analysis obtained after digitization has begun is used only in sensitivity analyses.
4. If multiple versions of the same figure exist, the publisher's final PDF (including corrected versions) is preferred, followed by the PMC version.

Primary-analysis curve for each comparison (confirmed against full-text publications on 12 October 2026):

| Comparison | Primary-analysis curve | Assessment | Figure type |
|---|---|---|---|
| LITESPARK-022 | NEJM 2026 Fig 1A | investigator | vector |
| KEYNOTE-564 | NEJM 2024 Fig 2A (data cutoff 15 September 2023; median follow-up 57.2 months); if the Ann Oncol 2026 five-year analysis is obtained before digitization, it will be used instead | investigator | vector |
| IMmotion010 | Lancet 2022 Fig 2A | investigator | vector |
| CheckMate 914 Part A | Lancet 2023 Fig 2 | BICR | vector |
| CheckMate 914 Part B nivo | JCO 2024 Fig 2 (PMC version, 3951 px) | BICR | raster |
| CheckMate 914 Part B nivo + ipi | JCO 2024 Data Supplement Fig S2 | BICR | raster |
| S-TRAC | NEJM 2016 Fig 2 | BICR | vector |
| ASSURE (both comparisons) | Lancet 2016 Fig 2 (PMC version, 636×375 px; publisher's main-text PDF not obtained) | not reported in the original publication | raster |
| PROTECT 600 mg | JCO 2017 Fig A1A (October 2016 follow-up analysis) | investigator | vector |
| ATLAS | Ann Oncol 2018 Fig 1 | IRC | vector |
| SORCE (both comparisons) | JCO 2020 Fig 2A, 2B | investigator | vector |
| PROSPER (sensitivity) | Lancet Oncol 2024 Fig 2 (corrected version, 24 November 2025) | investigator | vector |

OS curves are selected by the same rules (Section 8.5), **applied in the stated order: population (rule**

**1) takes precedence over analysis time point (rule 3)**. For example, the primary OS curve for PROTECT is JCO 2017 Fig 4A (ITT600mg interim analysis), whereas the final OS analysis in Eur Urol 2021 (ITTAll only) is used as a sensitivity analysis.

##### 5.2 Digitization and reconstruction

1. **Vector figures: direct coordinate extraction first.** When a curve is stored as a PDF vector path, path coordinates are read with Python PyMuPDF and converted to time and survival probability using axis tick positions, without manual point selection.
2. **Raster figures** are digitized with WebPlotDigitizer, with the axis calibration points recorded for each figure.
3. Individual patient data are reconstructed with the Guyot method (R package `IPDfromKM`), incorporating the number-at-risk table.
4. When the number censored at each time point or the total number of events is shown in the figure, these are also entered.
5. Each curve is digitized by one investigator and independently verified by a second investigator; digitization is repeated if the two differ substantially. For vector figures, the automatically extracted results are verified visually by the second investigator by overlaying them on the original figure.
6. All coordinate files, reconstructed data and code will be made publicly available on OSF or GitHub.

##### 5.3 Reconstruction quality checks (prespecified acceptance criteria)

The following quantities are recalculated from the reconstructed data and compared with the original publication:

| **Item** | **Acceptance criterion** |
|---|---|
| HR | Absolute difference between reconstructed and reported HR ≤ 0.05, and within the reported CI |
| Number of events | Within 5% of the reported value in each arm |
| Median DFS (if reported) | Difference ≤ 1 month, or within the reported CI |
| Landmark survival rates (if reported) | Difference ≤ 2 percentage points at each time point |
| Visual | No apparent deviation when the reconstructed KM curve is overlaid on the original figure |

- If a curve fails, it is re-digitized once; if it still fails, the curve is designated for sensitivity analyses only, and this is reported in the manuscript.
- When the original publication is internally inconsistent, the number-at-risk and censoring counts on the KM figure take precedence, and the discrepancy is recorded in the appendix. Known example: SORCE arm B events are reported as 245 in the main text and 242 in the CONSORT diagram (242 is used). For CheckMate 914 Part A, the discrepancy of 218 vs 228 has been resolved as 228 in the final published version.
- For trials reporting 97.5% CIs (ASSURE), the original CI is used for comparison.
- For the comparison without a reported HR (CheckMate 914 Part B nivo + ipi vs placebo), the HR item is marked "not applicable", and the check is instead based on the event counts shown in the figure (58/206 vs 54/208) and the number-at-risk table.

---

#### 6. Time parameters (prespecified)

##### 6.1 RMST truncation time τ

- **Primary τ = 36 months**; **secondary τ = 24 months**.

- Feasibility rule: in each arm, the number at risk at τ must be ≥ 10% of the arm's initial number of patients.
- If a trial does not meet the feasibility rule at 36 months, its primary τ is changed to 24 months, and this is indicated in the results. If LITESPARK-022 is changed to τ = 24 months under this rule, the abstract and main results of the manuscript will present ΔRMST at 24 months.
- If a trial does not meet the rule at 24 months either, only its maximum feasible τ is reported for that trial, and the analysis is labelled exploratory.
- **Determinations under this rule** (12 October 2026; based only on the ratios of the numbers in the number-at-risk tables of the primary-analysis curves, without inspecting the curves themselves):

| Trial | 36 months | 24 months | Primary τ |
|---|---|---|---|
| LITESPARK-022 | Not feasible (3.8%/4.1%) | Feasible (49.0%/45.3%) | **24 months** |
| CheckMate 914 Part B nivo | Not feasible (1.7%/1.0% at 36 months) | Feasible | 24 months |
| CheckMate 914 Part B nivo + ipi | Not feasible (0.5%/1.0%) | Feasible (26.2%/27.4%) | 24 months |
| KEYNOTE-564 (NEJM 2024) | Feasible (≥ 56.6%) | Feasible | 36 months |
| Other trials | Feasible (see the extraction file for each trial) | Feasible | 36 months |

These determinations were computed solely from the patient counts in the number-at-risk tables; no survival estimates read from the curves were examined.

##### 6.2 Cut points for piecewise HRs

The cut point equals the time at which planned treatment ends and is set directly in months; **it does not need to coincide with a number-at-risk time point** (a piecewise Cox model can be split at any time; the number-at-risk time points are used only for the quality checks in Section 5.3):

| Trial | Primary cut point | Sensitivity cut point | Rationale |
|---|---|---|---|
| LITESPARK-022 | 12 months | 15 months | pembro up to 9 doses q6w (last dose at approximately week 48); belzutifan up to 54 weeks; median treatment duration 12.4 months in both arms (NEJM 2026) |
| KEYNOTE-564 | 12 months | 15 months | pembro up to 17 doses q3w (approximately 1 year) |
| IMmotion010 | 12 months | 15 months | atezolizumab 16 doses q3w or 1 year, whichever occurs first; median 10.4 months |
| CheckMate 914 Part A/B | 6 months | 9 months | Planned 24 weeks, extendable to a maximum of 36 weeks |
| S-TRAC | 12 months | 15 months | 9 cycles, approximately 1 year |
| ASSURE | 12 months | 15 months | 54 weeks |
| PROTECT | 12 months | 15 months | 1 year |
| ATLAS | 12 months | 36 months | Planned duration variable, 1–3 years; the shortest duration is used as the primary cut point |
| SORCE 1 year vs placebo | 12 months | 15 months | 1 year |
| SORCE 3 years vs placebo | 36 months | 12 months | Planned 3 years, but actual median drug exposure approximately 10.7 months |
| PROSPER | 12 months | 10 months | Preoperative plus postoperative treatment approximately 9 months; 10 months is the authors' own landmark |

Sensitivity cut points are set later because recurrence is detected only at the next imaging assessment.

---

#### 7. Primary analysis

##### 7.1 RMST difference

- Within each trial, the RMST difference and 95% CI at $\tau$ = 36 months (or as determined by the rule in Section 6.1) and $\tau$ = 24 months are estimated with the R package `survRM2`.
- The RMST ratio is also reported.
- Results are presented as "an average of X additional months of disease-free survival within $\tau$ months".

##### 7.2 ΔRMST(τ) curve

- For each trial, ΔRMST and 95% pointwise CIs are calculated for $\tau$ from 6 months in 3-month steps up to the trial's maximum feasible $\tau$ (the 10% rule in Section 6.1), and plotted.
- The **post-treatment ΔRMST increment** is defined as ΔRMST(maximum feasible $\tau$) − ΔRMST(primary cut point), and is also expressed as an annual increment (increment ÷ years after the end of treatment). When the benefit persists, the increment is positive and stable; when the benefit converges, the increment approaches 0.
- This curve is the central figure addressing the primary question.

---

#### 8. Secondary analyses

##### 8.1 Piecewise HR

- Within each trial, a Cox model with a treatment × period interaction (with follow-up split at the Section 6.2 cut points using `survSplit`) is used to estimate the on-treatment HR and the post-treatment HR.
- Both period-specific HRs, the late/early HR ratio (the exponentiated interaction term) and its 95% CI are reported.
- Limitation: the post-treatment HR includes only patients who were event-free during the treatment period and is therefore subject to survivor selection bias; RMST is the primary measure, with piecewise HRs as supporting analyses.

##### 8.2 Trajectory of the DFS difference

- The difference in KM DFS between arms and its 95% CI are calculated at 12, 24, 36, 48 and 60 months (within the feasible time points of each trial).
- Whether the difference widens, is maintained or narrows after the end of treatment is described.

##### 8.3 Assessment of proportional hazards

- Global Schoenfeld residual test and log(−log) plots.
- The P value is reported for each trial; $P < 0.05$ is regarded as a violation of PH.

##### 8.4 Flexible parametric HR(t)

- Royston-Parmar model (R package `rstpm2`) with a time-varying treatment effect.
- Degrees of freedom are selected by AIC, between 1 and 5 for the baseline hazard and between 1 and 3 for the time-varying effect; the selection rule is prespecified and will not be adjusted based on results.
- HR(t) and its 95% CI are plotted, with the primary cut point marked.
- Sensitivity analysis: baseline degrees of freedom fixed at 3 and time-varying degrees of freedom fixed at 2, to confirm that conclusions are not affected by model selection.

### 8.5 OS

- OS curves are selected by the rules in Section 5.1, and RMST differences ($\tau$ per the rule in Section 6.1) and HR(t) are estimated.
- OS in LITESPARK-022 is from an interim analysis (29% of the final number of events) and is

analysed descriptively only, without inference.

- For LITESPARK-022, no separate RMST at a fixed τ = 30 months is reported; the ΔRMST(τ) curve in Section 7.2 already extends to the maximum feasible τ.

##### 8.6 Comparison of patterns by drug class

1. **Pooling of TKI trials:** the log(late/early HR ratio) of each TKI trial is pooled using a random-effects model (REML estimation of heterogeneity, Hartung-Knapp adjustment of the CI; R package `meta`).
2. **Pooling of ICI trials:** as above, for ICI trials.
3. **Handling of shared control arms:** only one comparison per trial enters the main pooled analysis, prespecified as follows:
    - ASSURE: sunitinib vs placebo (same drug as S-TRAC)
    - SORCE: 1 year vs placebo (treatment duration consistent with the other TKI trials)
    - CheckMate 914 Part B: nivo vs placebo (the primary comparison of the original trial)

    The other comparison is substituted in a sensitivity analysis.
4. **LITESPARK-022:** not pooled with either class; it is presented descriptively alongside the pooled results of the two classes, with a description of the position of its late/early HR ratio CI relative to the pooled values of each class. The control arm of LITESPARK-022 is pembrolizumab (an active control), so its effect represents "belzutifan added on top of ICI", which differs in nature from the placebo-controlled effects of the two classes; this will be stated explicitly in the manuscript. No formal statistical test between LITESPARK-022 and either class is performed, because that would constitute a cross-trial comparison.
5. All pooled analyses combine only within-trial ratio estimates; no arm-level data are pooled.

##### 8.7 Smoothed hazard curves

- Smoothed hazard curves for each arm are plotted with the R package `muhaz` to describe the timing of peak recurrence. This analysis is descriptive only.

---

#### 9. Sensitivity analyses

1. Piecewise analyses using the sensitivity cut points in Section 6.2.
2. KEYNOTE-564 using the curves from each data cutoff in NEJM 2021, Lancet Oncol 2022 and NEJM 2024.
3. Inclusion of PROSPER (RFS, with t = 0 at preoperative randomization), using main-text Fig 2 (corrected version, 24 November 2025; vector figure). The supplementary appendix version in which Day 1 events are censored (raster, 768×480) is reported separately only if it passes the quality checks in Section 5.3; otherwise it is only described in the appendix.
4. ASSURE using the clear cell subset (if the appendix curve is available) or the high-risk ccRCC subset (JAMA Oncol 2017 Fig 2A).
5. PROTECT using the 800 mg population and the ITTAll population.
6. ATLAS using the highest-risk subset and investigator assessment.
7. SORCE using the Leibovich high-risk subset and the clear cell subset.
8. In the class-level pooling, substitution of the other comparison for trials with a shared control arm (Section 8.6).
9. Curves that fail the reconstruction quality checks analysed both with exclusion and with inclusion.

---

#### 9A. Exploratory analyses and reporting rules

1. **SORCE 3 years vs 1 year:** both arms are reconstructed from Fig 2A and 2B, respectively, and the RMST difference and piecewise HR are compared within the trial; this is reported only in the appendix as an exploratory analysis. Because the actual median drug exposure was similar (11.2 months in the 1-year arm and 10.7 months in the 3-year arm), the results will not be used as a main argument.
2. **Table 1:** "discontinuation due to AEs" and "median follow-up" are listed as reported in each original publication, with definitional differences between trials explained in footnotes and without conversion.

---

#### 10. Known limitations (prespecified)

1. Reconstructed IPD are approximations; individual treatment discontinuation times are not available, so periods can be defined only by planned treatment duration.
2. The post-treatment HR is subject to survivor selection bias.
3. Post-treatment follow-up in LITESPARK-022 is short (follow-up range 15.0–40.1 months), so the precision of post-treatment estimates is limited; the results are framed as a testable prediction, and this analysis will be updated when longer follow-up is published.
4. DFS definitions, assessment methods and imaging frequency differ across trials; in some trials imaging frequency decreased after the end of treatment (e.g., SORCE switched to chest X-ray only), which may be confounded with curve convergence.
5. TKI trials were conducted earlier and differed in risk-based eligibility criteria and histology; only patterns are compared across trials.
6. Reconstructed data contain no patient-level covariates, so subgroup or adjusted analyses are not possible.

---

#### 11. Software and reproducibility

- R (version to be recorded at installation): `IPDfromKM`, `survival`, `survRM2`, `rstpm2`, `muhaz`, `meta`.
- WebPlotDigitizer (version to be recorded), for raster figures.
- Python PyMuPDF (1.28.2), for coordinate extraction from vector figures and rendering of PDF pages.
- This plan is registered on OSF to obtain a timestamp; **registration must be completed before digitization of LITESPARK-022 and KEYNOTE-564**.
- Code, coordinate files and reconstructed data will be released at the time of manuscript submission.

---

#### 12. Data availability

Digitized coordinates, reconstructed datasets, and code will be made publicly available upon publication.

---

#### Version history

Version 4.0 (12 October 2026): version registered on OSF. Earlier internal drafts (v1–v3, 11–12 October 2026) were refined using full-text extraction of trial publications; no reconstructed data had been generated at the time of registration.
